# Tixagevimab-cilgavimab (AZD7442) for the treatment of patients hospitalized with COVID-19 (DisCoVeRy): A phase 3, randomized, double-blind, placebo-controlled trial

**DOI:** 10.1101/2024.02.23.24302586

**Authors:** Maya Hites, Clément R. Massonnaud, Simon Jamard, François Goehringer, François Danion, Jean Reignier, Nathalie de Castro, Denis Garot, Eva Larranaga Lapique, Karine Lacombe, Violaine Tolsma, Emmanuel Faure, Denis Malvy, Thérèse Staub, Johan Courjon, France Cazenave-Roblot, Anne Ma Dyrhol Riise, Paul Leturnier, Guillaume Martin-Blondel, Claire Roger, Karolina Akinosoglou, Vincent Le Moing, Lionel Piroth, Pierre Sellier, Xavier Lescure, Marius Trøseid, Philippe Clevenbergh, Olav Dalgard, Sébastien Gallien, Marie Gousseff, Paul Loubet, Fanny Vardon-Bounes, Clotilde Visée, Leila Belkhir, Élisabeth Botelho-Nevers, André Cabié, Anastasia Kotanidou, Fanny Lanternier, Elisabeth Rouveix-Nordon, Susana Silva, Guillaume Thiery, Pascal Poignard, Guislaine Carcelain, Alpha Diallo, Noémie Mercier, Vida Terzic, Maude Bouscambert-Duchamp, Alexandre Gaymard, Marie Anne Trabaud, Grégory Destras, Laurence Josset, Drifa Belhadi, Nicolas Billard, Jérémie Guedj, Thi-Hong-Lien Han, Sandrine Couffin-Cadiergues, Aline Dechanet, Christelle Delmas, Hélène Esperou, Claire Fougerou-Leurent, Soizic Le Mestre, Anabelle Métois, Marion Noret, Isabelle Bally, Sebastián Dergan-Dylon, Sarah Tubiana, Ouifiya Kalif, Nathalie Bergaud, Benjamin Leveau, Joe Eustace, Richard Greil, Edit Hajdu, Monika Halanova, Jose-Artur Paiva, Anna Piekarska, Jesus Rodriguez Baño, Kristian Tonby, Milan Trojánek, Sotirios Tsiodras, Serhat Unal, Charles Burdet, Dominique Costagliola, Yazdan Yazdanpanah, Nathan Peiffer-Smadja, France Mentré, Florence Ader, DisCoVeRy study group

## Abstract

**Background:** Tixagevimab and cilgavimab (AZD7442) are two monoclonal antibodies developed by AstraZeneca for the pre-exposure prophylaxis and treatment of patients infected by SARS-CoV-2. Its effectiveness and safety in patients hospitalized with COVID-19 was not known at the outset of this trial.

**Methods:** DisCoVeRy is a phase 3, adaptive, multicentre, randomized, controlled trial conducted in 63 sites in Europe. Participants were randomly assigned (1:1) to receive placebo or tixagevimab-cilgavimab in addition to standard of care. The primary outcome was the clinical status at day 15 measured by the WHO seven-point ordinal scale. Several clinical, virological, immunological and safety endpoints were also assessed.

**Findings:** Due to slow enrolment, recruitment was stopped on July 1^st^, 2022. The antigen positive modified intention-to-treat population (mITT) was composed of 173 participants randomized to tixagevimab-cilgavimab (N = 91) or placebo (N = 82), 91.9% (159/173) with supplementary oxygen, and 47.4% (82/173) previously vaccinated at inclusion. There was no significant difference in the distribution of the WHO ordinal scale at day 15 between the two groups (odds ratio (OR) 0.93, 95%CI [0.54-1.61]; p = 0.81) nor in any clinical, virological or safety secondary endpoints. In the global mITT (N = 226), neutralization antibody titers were significantly higher in the tixagevimab-cilgavimab group/patients compared to placebo at day 3 (Least-squares mean differences (LSMD) 1.44, 95% Confidence interval (CI) [1.20-1.68]; p < 10^−23^) and day 8 (LSMD 0.91, 95%CI [0.64-1.18]; p < 10^−8^) and it was most important for patients infected with a pre-omicron variant, both at day 3 (LSMD 1.94, 95% CI [1.67-2.20], p < 10^−25^) and day 8 (LSMD 1.17, 95% CI [0.87-1.47], p < 10^−9^), with a significant interaction (p < 10^−7^ and p = 0.01 at days 3 and 8, respectively).

**Interpretation:** There were no significant differences between tixagevimab-cilgavimab and placebo in clinical endpoints, however the trial lacked power compared to prespecified calculations. Tixagevimab-cilgavimab was well tolerated, with low rates of treatment related events.

**Funding:** Trial registration: ClinicalTrials.gov NCT04315948. Registered on 13 March 2020 updated on 22 April 2021.

## Introduction

Even though COVID-19 is no longer a public health emergency,^1^ SARS-CoV-2 continues to circulate worldwide, and therefore individuals unable to mount an adequate immune response (e.g. the elderly and the immunocompromised), or unvaccinated/disease naïve individuals at risk for severe COVID-19, remain vulnerable. Although immunomodulatory agents (corticosteroids, interleukin-1 and 6 inhibitors, JAK inhibitors) and direct-acting antiviral remdesivir have been identified as effective treatments to decrease mortality in sub-groups of patients hospitalized with COVID-19,^2–6^ antibody-mediated SARS-CoV-2 neutralization may be another treatment option in vulnerable patient groups.

Indeed, in the ambulatory setting, the administration of monoclonal antibodies (mAbs) directed against SARS-CoV2 (intravenous sotrovimab or intramuscular tixagevimab-cilgavimab), significantly reduced the risk of hospitalization and death in patients at risk for disease progression, compared to placebo.^7,8^ In the hospital setting, several trials on mAbs for COVID-19 were terminated early for futility (bamlavinimab in 314 patients, and sotrovimab or BRII-196/BRII-198 in 546 patients).^9,10^ Several other trials showed a potential therapeutic benefit of mAbs in this setting. The open-label RECOVERY platform trial was the first to demonstrate a 28-day mortality benefit in seronegative patients at baseline for SARS-CoV-2 infection in patients who received casirivimab-imdevimab plus standard of care (SoC) compared to SoC alone.^11^ The company-sponsored double-blinded, placebo-controlled trial also showed that casirivimab-imdevimab treatment was associated with a significant reduction in 28-day mortality for seronegative patients, but not for seropositive ones.^12^ The mAb cocktail with an extended half-life, tixagevimab-cilgavimab (AZD7442), was also evaluated as an intravenous formulation in association with SoC (which included remdesivir) in the double-blinded, placebo-controlled ACTIV-3-TICO trial in 1417 hospitalized patients with COVID-19.^13^ Although the primary outcome of time to sustained recovery was not met, treatment with tixagevimab-cilgavimab was associated with a decreased 90-day mortality.

Since these trials were completed, the virus itself has evolved, resulting in the emergence of different variants of concern (VOC) and mass vaccination campaigns against SARS-CoV-2 were ramped up worldwide.^14^ In this context, of an evolving pandemic, we report here the results from the DisCoVeRy platform trial that evaluated the efficacy and safety of intravenous tixagevimab-cilgavimab in SARS-CoV-2 antigenic positive patients (i.e those with a high SARS-CoV2 viral load) hospitalized with COVID-19, and followed-up to day 90.

## Materials and Methods

### Study design

DisCoVeRy is a phase 3, adaptive, multicentre, randomized, double-blind, superiority trial for evaluating the efficacy and safety of drugs for the treatment of adults hospitalized for COVID-19. The first four interventions evaluated (lopinavir/ritonavir, lopinavir/ritonavir plus IFN-b-1a, hydroxychloroquine, and remdesivir) have been stopped for futility and the results reported elsewhere.^15–17^ The present analysis is based on the protocol version 15.0 of September 20th, 2021, which evaluates the cocktail of two monoclonal antibodies (mAbs), tixagevimab and cilgavimab developed by AstraZeneca. The study was conducted across 63 sites in 13 European countries (Austria, Belgium, Czech Republic, France, Greece, Hungary, Ireland, Luxembourg, Norway, Poland, Portugal, Slovakia, and Spain). The trial was approved by the Ethics Committees in each country and is sponsored by the Institut national de la santé et de la recherche médicale (Inserm, France); it was conducted in accordance with the Declaration of Helsinki.

### Participants

Eligible participants were adults (> 18 years) hospitalized with SARS-CoV-2 infection confirmed on a RT-PCR performed on a nasopharyngeal (NP) swab within the 5 days preceding randomization, and a time between onset of symptoms and randomization of less than 9 days (extended to 11 days from September 20th, 2021, protocol V15). Participants with prior receipt of investigational or licensed mAb or vaccine indicated for the prevention of SARS-CoV-2 infection or COVID-19 were excluded. From September 20th, 2021 (protocol V15), inclusion of patients with initiated (one dose) or completed (two doses) vaccination was allowed in a proportion of 20% of enrolled participants, provided they would not receive an additional dose in the 30 days following hospital discharge. Full inclusion and exclusion criteria are detailed in the appendix. Written informed consent was obtained from all participants or their legal representative.

### Randomization and masking

Participants were randomly assigned in a 1:1 ratio to receive tixagevimab-cilgavimab or placebo in addition to standard of care (SoC). The randomization was stratified by region, by antigenic status obtained from the result of a rapid antigen test on a NP swab at enrolment, and with protocol V15, by vaccination status. Randomization was performed via an electronic Case Report Form and used computer-generated blocks of various sizes. The allocation of tixagevimab-cilgavimab was double-blinded.

### Procedures

The participants received a single intravenous infusion of tixagevimab-cilgavimab (600 mg at a maximum infusion rate of 50 mg/minute) or placebo (a 0.9% saline solution) within 24 hours following randomization. Participants could not be treated with other monoclonal or polyclonal antibodies directly targeting SARS-CoV-2 during the study. The SoC could include corticosteroids, remdesivir, and other immunomodulatory agents at the discretion of the physician.

Participants were assessed daily while hospitalized. The evaluation of the primary endpoint was at day 15, and participants were contacted by phone to collect the data if they had already been discharged from the hospital. Follow-up assessments were scheduled at days 29, 90, 180, 365 and 456. For all participants discharged from the hospital, assessments at days 29 and 90 were organized as outpatient consultations. Assessments at days 180 and 365 were also organized as outpatient consultations, but only for a subset of 25% of patients enrolled in centers with available resources, selected at day 90, the others were contacted by phone. For the final assessment at day 456, all patients were contacted by phone.

NP swabs or lower respiratory tract samples were obtained at baseline (day 1 pre-treatment) and days 3, 8, 15 (while hospitalized) and 29. Blood samples were obtained at baseline, at days 3, 8, 15 (while hospitalized), and at days 29, 90, 180 and 365 (for the subset of patients evaluated during a medical consultation at these times).

The blood samples included samples for safety laboratory tests, serum samples for serology, drug concentration and anti-drug antibodies measurements, and serum and plasma samples for exploratory objectives.

### Outcomes

The primary endpoint was the clinical status at day 15 measured on the 7-point ordinal scale of the WHO Master Protocol (v3.0, March 3, 2020). The key secondary endpoint was the time from randomization to sustained recovery, defined as being discharged from the index hospitalization, followed by being alive and at home for 14 consecutive days prior to day 90 (details in the appendix). Other secondary efficacy endpoints included: clinical status on the ordinal scale at days 29, 90, 180, and 365; change of the National Early Warning Score 2 (NEWS-2) from baseline to days 3, 8, 15 (if hospitalized), and 29; oxygenation and ventilator-free days from baseline to day 29; time to new oxygen use, non-invasive ventilation or high flow oxygen devices during the first 29 days; time to new invasive mechanical ventilation use during the first 29 days; new need for mechanical ventilation or death by day 15; time to hospital discharge; in-hospital mortality and/or at days 29, 90, 180, 365, or 456; occurrence of new hospitalization between discharge from index hospitalization and days 90, 180 and 365; occurrence of confirmed re-infection with SARS-CoV-2 between discharge and Days 90, 180 and 365.

Safety endpoints included: cumulative incidence of serious adverse events (SAEs), grade 3 and 4 adverse events (AEs), grade 1-2 hypersensitivity-related and infusion-related AEs until day 29, or AE of special interest (AESI); the discontinuation of investigational product for any reason.

Virological endpoints included the measures at baseline, days 3, 8, 15 (while hospitalized) and 29 of: the proportion of participants with detectable SARS-CoV-2 in NP or LRT samples; the normalized quantitative viral load in NP or LRT samples; and their respective change from baseline. They also included the time to first undetectable normalized quantitative viral load in NP swabs.

The pre-specified exploratory analysis of neutralizing activity of patients’ serum samples against their variant of infection, or against delta for pre-omicron variants of infection, was performed. The endpoints related to this analysis included: the neutralizing antibody titers at day 3 and day 8; the change of titers from baseline to days 3 and 8; and the proportion of patients with neutralizing antibody titers below the limit of detection (LOD).

Details on the outcomes are in appendix.

### Statistical analysis

#### Sample size

The study was powered to detect an odds ratio (OR) of 1.5 (an OR greater than 1 indicates superiority of the experimental treatment over the control for each ordinal scale category), with a 90% power and a two-sided type I error of 0.05, for the primary endpoint. We determined that 413 evaluable, antigen positive patients were needed in each arm. Since antigen negative patients can represent as much as 30% of patients, 590 participants (antigen positive or negative) were needed in each arm. Accounting for 5% of non-evaluable patients, we needed to include a total of 620 patients per arm (details in appendix).

#### Populations

The intention-to-treat (ITT) population included all randomized participants analyzed in their assigned randomization group. The modified intention-to-treat (mITT) population included participants from the ITT population for whom the infusion of the allocated treatment was truly initiated. The safety population included all randomized participants, analyzed in their actual treatment group. Each population set was further categorized according to the antigenic status, thereby defining « Antigen positive » ITT and mITT. The set of all patients (i.e. antigenic positive and negative) in ITT, mITT, and safety populations are referred to as the « Global » ITT, mITT, and safety populations.

The efficacy analyses were conducted on the antigen positive mITT population, as it is hypothesized that tixagevimab-cilgavimab may be most effective in antigen positive patients. Efficacy analyses were also conducted on the global mITT population as a sensitivity analysis. Safety analyses were performed on the global safety population. The analyses were stratified by vaccination status, but not by region due to the low number of inclusions in some regions. For global populations, analyses were also stratified by antigenic status. The analyzes of the neutralizing activity of serum samples were performed on patients from the global mITT population with known or imputed variants of infection (details in appendix). Variants of infections were grouped in three categories: (a) pre-omicron (alpha, beta, gamma, delta, other), (b) omicron BA.1, (c) omicron BA.2/5 (omicron BA.2 or BA.5).

The analyses reported here were performed on the pre-specified primary database lock with all endpoints collected until day 90.

#### Statistical tests

The primary endpoint, clinical status at day 15 measured on the 7-point ordinal scale, was analyzed with a proportional odds model. For the key secondary endpoint, time to sustained recovery, cumulative incidence functions were estimated for each group using the Aalen-Johansen estimator, and the recovery rate ratio (RRR) for sustained recovery were estimated using the Fine and Gray method. For virological endpoints, the proportion of subjects with detectable viral loads in NP swabs were compared between groups using a stratified Cochran– Mantel–Haenszel (CMH) test. Time to first undetectable viral load through day 29 was analyzed with the same approach as for the time to sustained recovery. The log_10_ normalized quantitative SARS-CoV-2 viral load in NP swabs were compared between groups using linear mixed models with repeated measures for each patient, with the treatment arm as the main exposure variable, and the time of measurement, time by treatment interaction, and stratification variables as covariates. Patient effect was accounted for as random intercept (log_10_ copies/10 000 cells) and slope (log_10_ copies/10 000 cells per day). The differences in the viral loads, and in the change from baseline of the viral loads between treatment groups were estimated as differences of slopes using the linear mixed effects model.

Analyzes of neutralizing antibody titers were performed on the global mITT population, stratified by vaccination status and by variant of infection (pre-omicron; omicron BA.1; omicron BA.2/5). The log_10_ titers at day 3 and day 8, and the change of titers from baseline to days 3 and 8 were compared between the two arms using an ANCOVA model and results reported as least-square mean differences (LSMD). The proportion of subjects with undetectable neutralizing antibody titers at days 3 and 8 were compared between groups using a stratified CMH test. Details on the statistical tests and handling of missing data are in the appendix. All analyses were done using SAS 9.4.

#### Subgroup analyses

Prespecified subgroup analyses for the primary outcome, the key secondary outcome, and the differences of slopes of normalized viral loads were performed across the following subgroups: vaccination status (yes / no), clinical status at inclusion (ordinal scale 3-4 / ordinal scale 5), duration of symptoms prior to randomization (less than 5 days / 5 days or more), age (less than 65 years / 65 years or more), SARS-CoV-2 serology at inclusion (negative / positive), SARS-CoV-2 variant of infection (pre-omicron / omicron BA.1 / omicron BA.2/5), sex (female / male), and for analyses on the global mITT population, the antigenic status at inclusion (positive / negative). Prespecified subgroup analyses for the serum neutralization outcomes were performed on the following subgroups: vaccination status, SARS-CoV-2 serology at inclusion, and variant of infection. Interactions between the treatment and the subgroup variables were tested and the p-values are reported. Post-hoc subgroup analyses for the cumulative incidence of SAEs, the first safety endpoint, was also performed on the same subgroups in the safety population.

#### Trial safety and monitoring

There was no pre-planned interim analysis for efficacy nor futility, and none were performed. An independent data safety and monitoring board (DSMB) externally reviewed the trial data on two scheduled occasions, as pre-specified in the protocol. The trial was registered on ClinicalTrials.gov (NCT04315948) on 13 March 2020, updated on 22 April 2021. The full protocol and statistical analysis plans are available as supplementary material.

## Results

Due to slow enrolments and *in-vitro* data showing decreased activity of tixagevimab-cilgavimab against the omicron circulating variants,^18,19^ the study steering committee decided to stop the inclusions on July 1^st^ 2022, in all participating sites, before having reached the targeted sample size. Between April 28^th^ 2021, and June 23^rd^ 2022, 237 participants were randomized across 34 sites in 6 European countries (France, n=206 ; Belgium, n=15 ; Norway, n=6 ; Luxembourg, n=5 ; Greece, n=4 ; Portugal, n=1). Five participants were excluded from the ITT population (no valid informed consent form, n=3; withdrawal of consent, n=2) and six excluded from the mITT population (withdrawal of consent, n=2; investigator decision, n=3; transfer to non-participating study site, n=1). Fifty-three participants had a negative antigenic status at inclusion; they were excluded from the antigenic-positive mITT population (figure 1).

**Figure 1.**
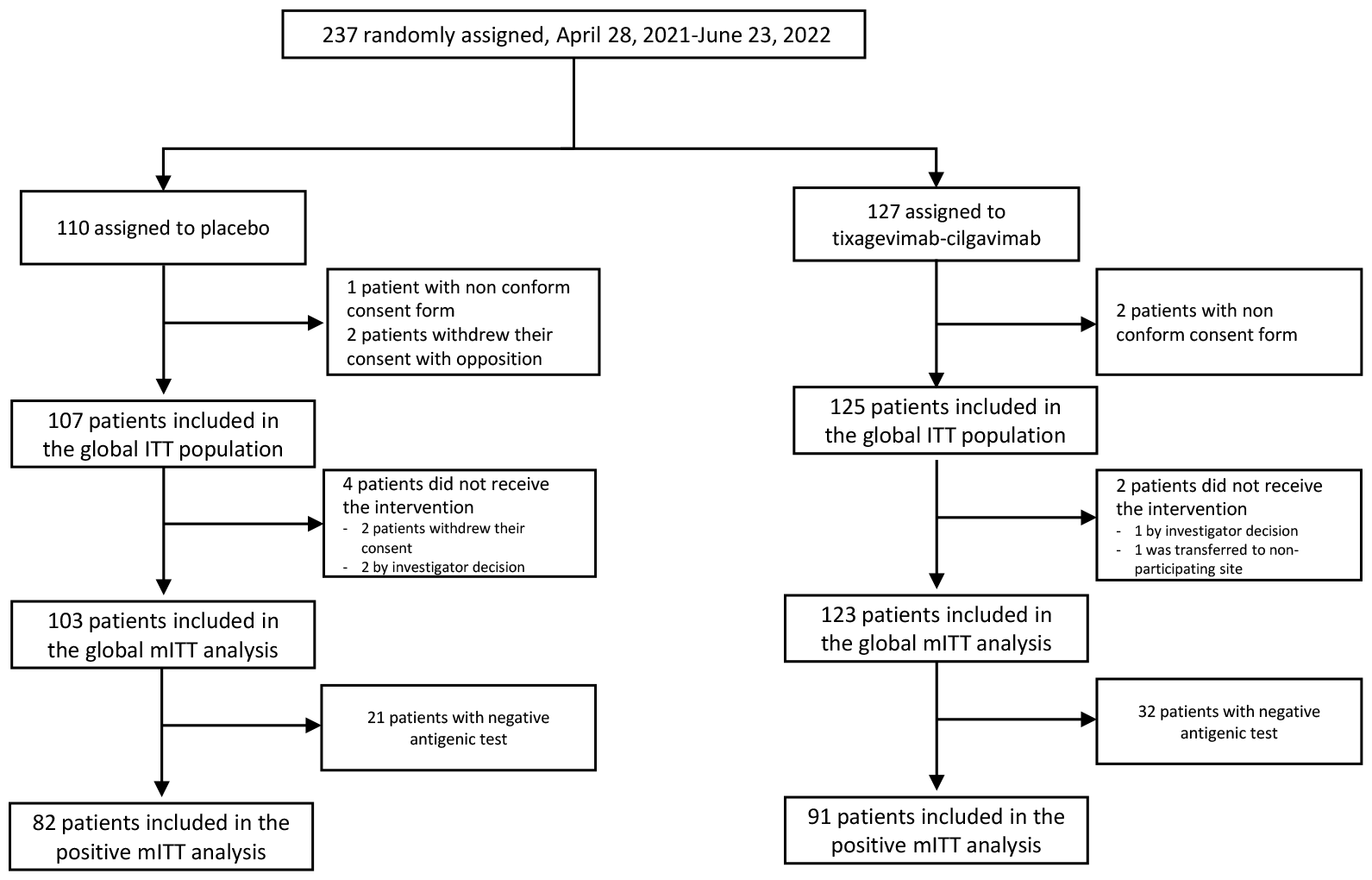
Trial profile.

Participants’ baseline characteristics are presented in table 1, without notable imbalance between arms. Among the 226 participants in the global mITT population set, 173 (76.5%) had a positive antigen test at baseline and vaccination was initiated or completed for 45.1% (102/226) of participants. In the global mITT population, the median age was 66 years (IQR 53 – 76) and 68.6% (155/226) were male. The median time from symptom onset to randomization was 7 days (IQR 6 – 9). Overall, 77% (174/226) of participants had at least one comorbid condition. Most participants were infected with one of the variants circulating before omicron (58.8%, 133/226); others were infected with omicron BA.1 (23.5%, 53/226), or omicron BA.2 or BA.5 (9.7%, 22/226). Data on the variant of infection was missing for 8.0% (18/226) of participants (no sequencing available and not imputed).

**Table 1:**
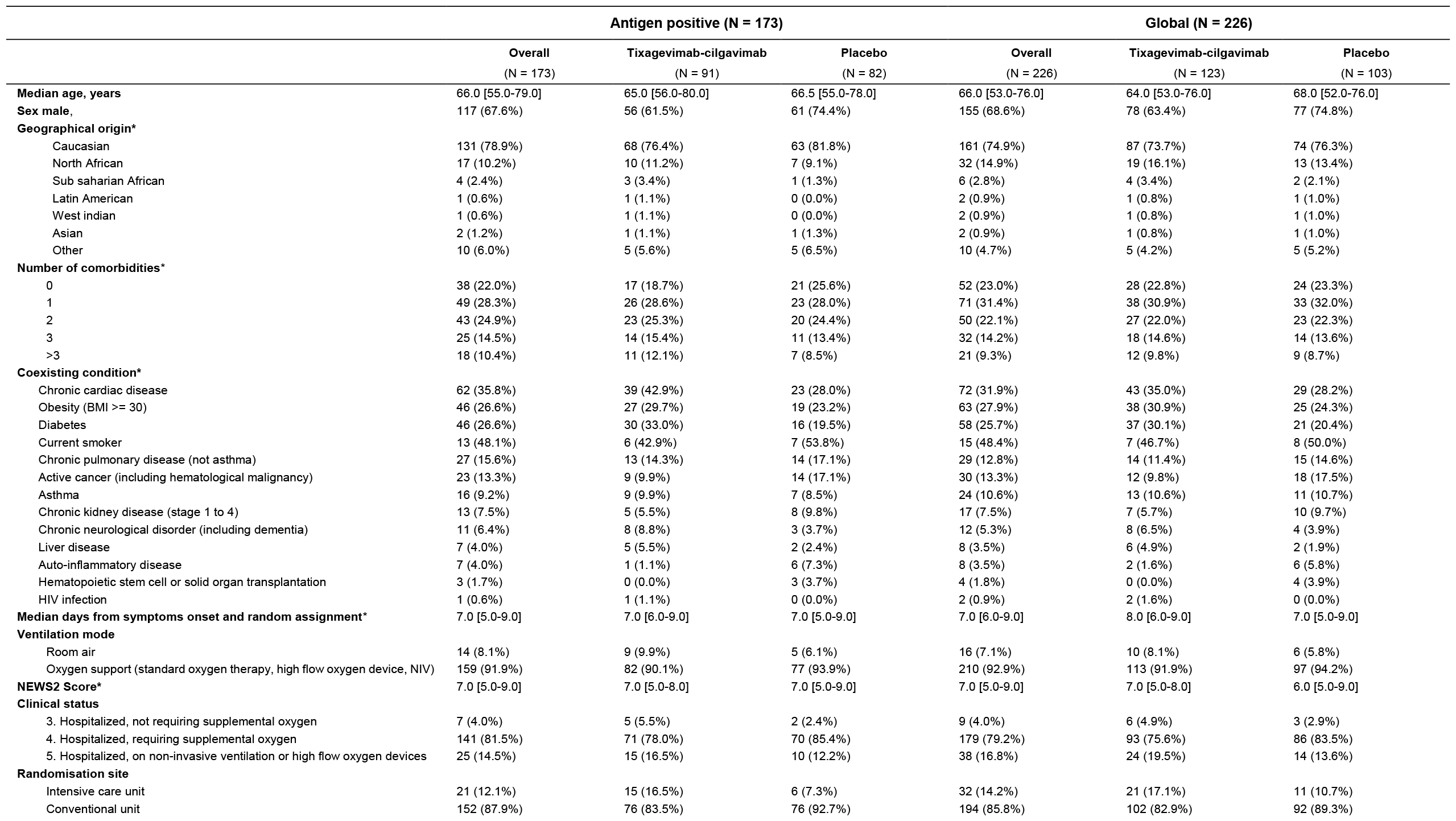
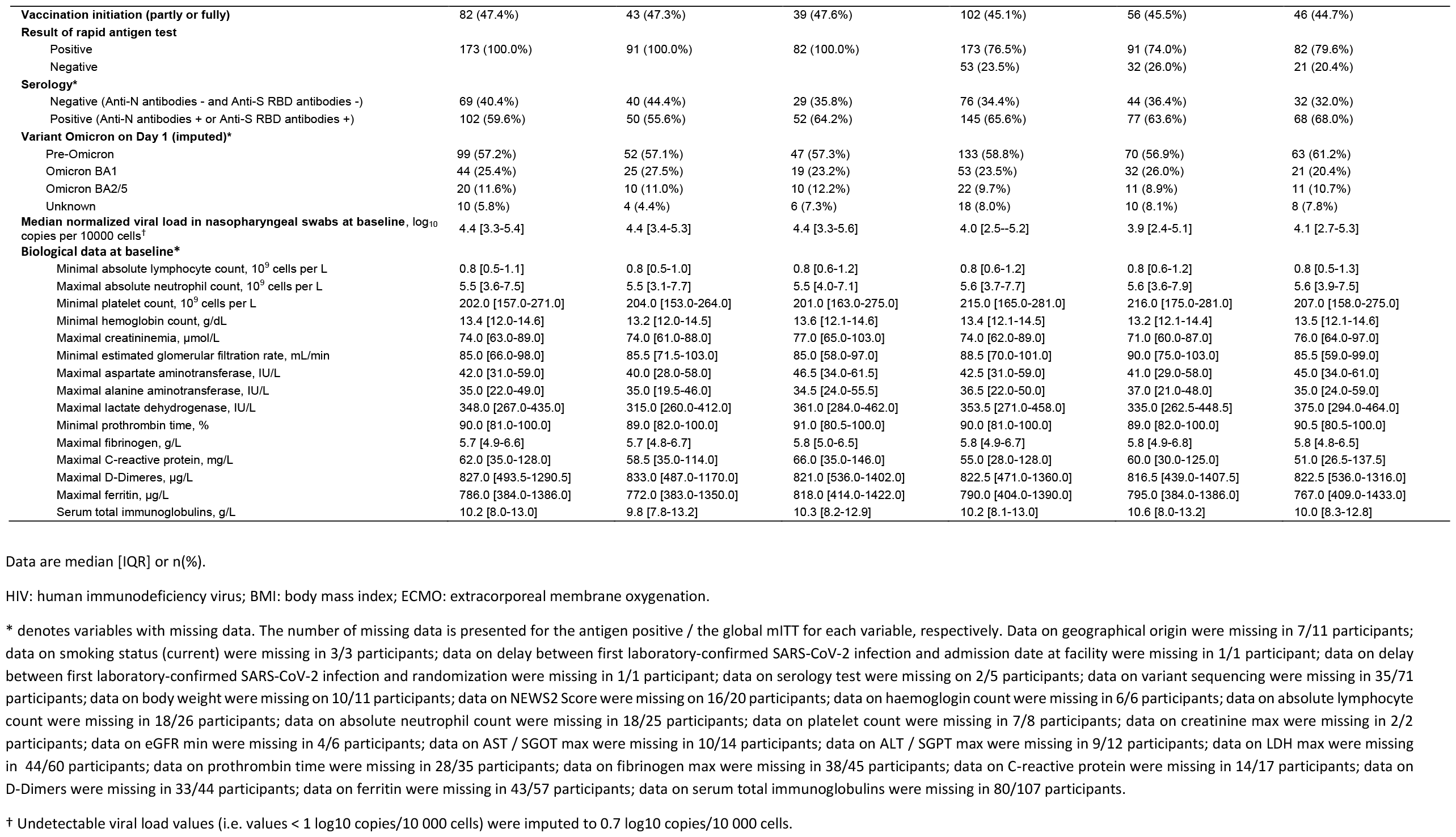
Baseline characteristics of participants in the antigen positive and global modified intention-to-treat populations, overall and according to the treatment group.

Table 2 shows the clinical status of patients in the tixagevimab-cilgavimab and placebo groups according to the WHO ordinal scale, at day 15, day 29, and day 90, for the antigen positive and global mITT populations. There was no significant difference in the distribution of the seven-point ordinal scale at day 15 between the tixagevimab-cilgavimab and control groups (figure 2; table 2). At day 15, 64.7% (112/173) of antigen positive participants were no longer hospitalized (65.9%, 60/91 for tixagevimab-cilgavimab vs. 63.4%, 52/82 for placebo), and 9.8% (17/173) had died (8.8%, 8/91 for tixagevimab-cilgavimab vs. 11.0%, 9/82 for placebo). There was no significant difference in all subgroup analyses according to vaccination status, clinical status at inclusion, variant of infection, duration of symptoms prior to randomization, antigenic status (for the global population), age, sex, or serology (figure S1, appendix). Likewise, there was no significant difference in the distribution of the seven-point ordinal scale at day 29 and at day 90 (figure 2; table 2). The median time to sustained recovery through day 90 was not significantly different between the tixagevimab-cilgavimab and placebo groups, in both the antigen positive and global populations, as well as for any of the subgroup analyses (table 2; figures S2 and S4, appendix). There was no significant interaction between the treatment and the subgroup variables exept for sex, with a greater effect of tixagevimab-cilgavimab for males in both the antigen positive and the global populations (p=0.02 and p=0.04). No significant difference was observed for any other clinical secondary endpoints (table 2; table S2; figure S3, appendix). Analysis of change from baseline in the NEWS-2 score was limited to day 3, due to an important amount of missing data at days 8, 15, and 29.

**Table 2.**
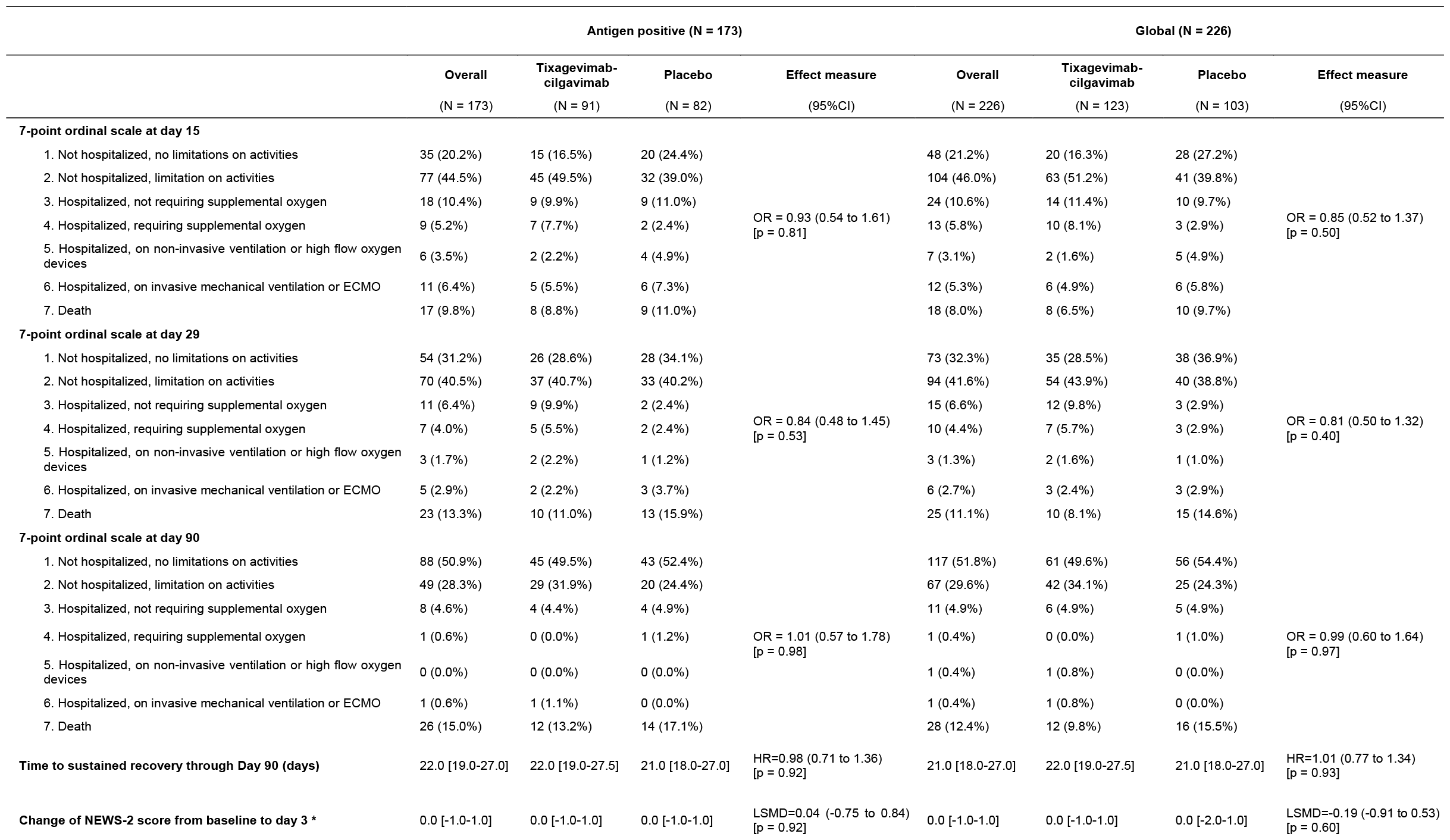
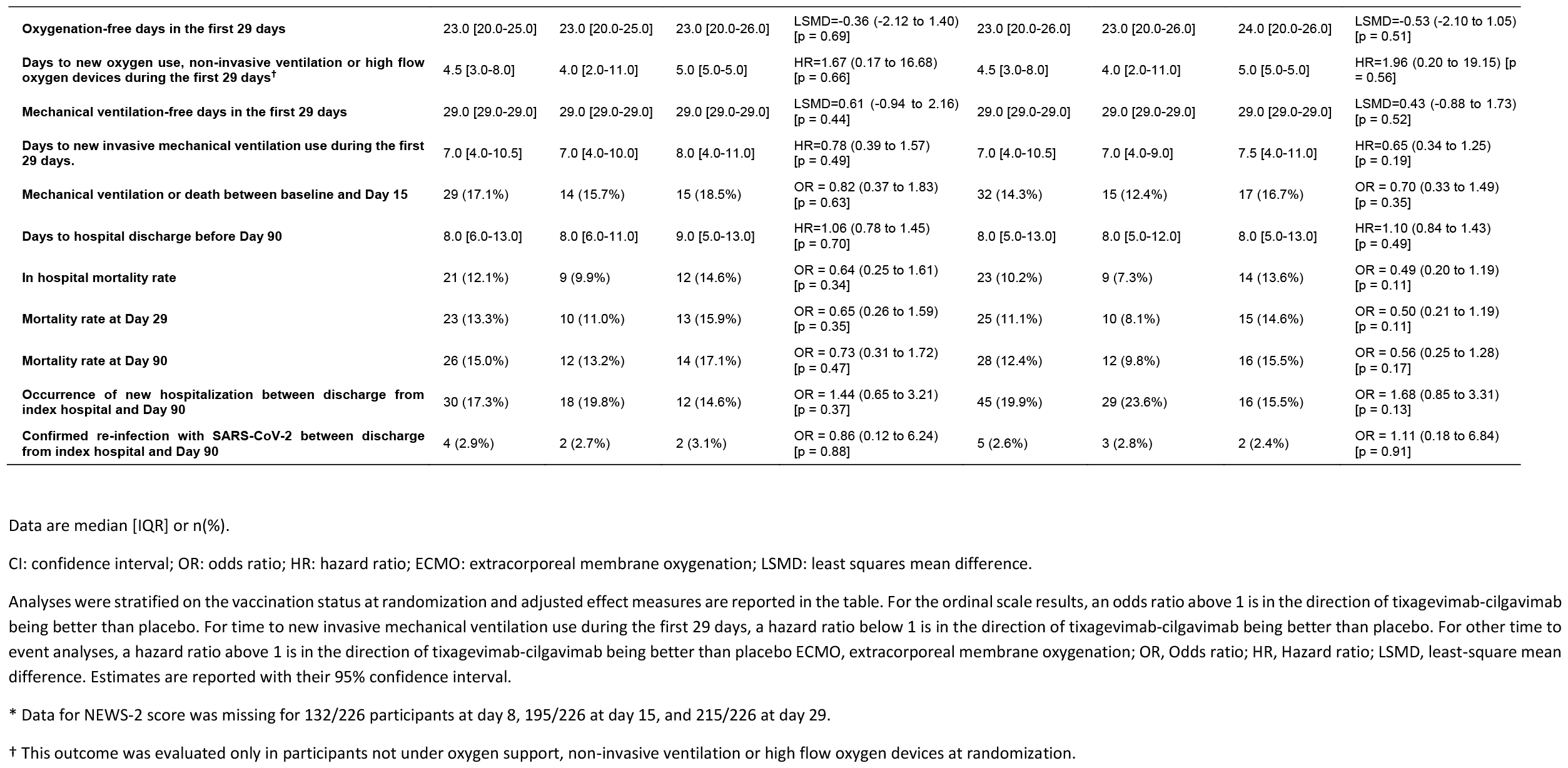
Primary and secondary clinical outcomes in the antigen positive and global modified intention-to-treat populations, overall and according to the treatment group.

|  | Antigen positive (N = 173) |  |  |  | Global (N = 226) |  |  |  |
| --- | --- | --- | --- | --- | --- | --- | --- | --- |
|  | Overall<br>(N = 173) | Tixagevimab-cilgavimab<br>(N = 91) | Placebo<br>(N = 82) | Effect measure<br>(95%CI) | Overall<br>(N = 226) | Tixagevimab-cilgavimab<br>(N = 123) | Placebo<br>(N = 103) | Effect measure<br>(95%CI) |
| **7-point ordinal scale at day 15** |  |  |  |  |  |  |  |  |
| 1. Not hospitalized, no limitations on activities | 35 (20.2%) | 15 (16.5%) | 20 (24.4%) | OR = 0.93 (0.54 to 1.61)<br>[p = 0.81] | 48 (21.2%) | 20 (16.3%) | 28 (27.2%) | OR = 0.85 (0.52 to 1.37)<br>[p = 0.50] |
| 2. Not hospitalized, limitation on activities | 77 (44.5%) | 45 (49.5%) | 32 (39.0%) |  | 104 (46.0%) | 63 (51.2%) | 41 (39.8%) |  |
| 3. Hospitalized, not requiring supplemental oxygen | 18 (10.4%) | 9 (9.9%) | 9 (11.0%) |  | 24 (10.6%) | 14 (11.4%) | 10 (9.7%) |  |
| 4. Hospitalized, requiring supplemental oxygen | 9 (5.2%) | 7 (7.7%) | 2 (2.4%) |  | 13 (5.8%) | 10 (8.1%) | 3 (2.9%) |  |
| 5. Hospitalized, on non-invasive ventilation or high flow oxygen devices | 6 (3.5%) | 2 (2.2%) | 4 (4.9%) |  | 7 (3.1%) | 2 (1.6%) | 5 (4.9%) |  |
| 6. Hospitalized, on invasive mechanical ventilation or ECMO | 11 (6.4%) | 5 (5.5%) | 6 (7.3%) |  | 12 (5.3%) | 6 (4.9%) | 6 (5.8%) |  |
| 7. Death | 17 (9.8%) | 8 (8.8%) | 9 (11.0%) |  | 18 (8.0%) | 8 (6.5%) | 10 (9.7%) |  |
| **7-point ordinal scale at day 29** |  |  |  |  |  |  |  |  |
| 1. Not hospitalized, no limitations on activities | 54 (31.2%) | 26 (28.6%) | 28 (34.1%) | OR = 0.84 (0.48 to 1.45)<br>[p = 0.53] | 73 (32.3%) | 35 (28.5%) | 38 (36.9%) | OR = 0.81 (0.50 to 1.32)<br>[p = 0.40] |
| 2. Not hospitalized, limitation on activities | 70 (40.5%) | 37 (40.7%) | 33 (40.2%) |  | 94 (41.6%) | 54 (43.9%) | 40 (38.8%) |  |
| 3. Hospitalized, not requiring supplemental oxygen | 11 (6.4%) | 9 (9.9%) | 2 (2.4%) |  | 15 (6.6%) | 12 (9.8%) | 3 (2.9%) |  |
| 4. Hospitalized, requiring supplemental oxygen | 7 (4.0%) | 5 (5.5%) | 2 (2.4%) |  | 10 (4.4%) | 7 (5.7%) | 3 (2.9%) |  |
| 5. Hospitalized, on non-invasive ventilation or high flow oxygen devices | 3 (1.7%) | 2 (2.2%) | 1 (1.2%) |  | 3 (1.3%) | 2 (1.6%) | 1 (1.0%) |  |
| 6. Hospitalized, on invasive mechanical ventilation or ECMO | 5 (2.9%) | 2 (2.2%) | 3 (3.7%) |  | 6 (2.7%) | 3 (2.4%) | 3 (2.9%) |  |
| 7. Death | 23 (13.3%) | 10 (11.0%) | 13 (15.9%) |  | 25 (11.1%) | 10 (8.1%) | 15 (14.6%) |  |
| **7-point ordinal scale at day 90** |  |  |  |  |  |  |  |  |
| 1. Not hospitalized, no limitations on activities | 88 (50.9%) | 45 (49.5%) | 43 (52.4%) | OR = 1.01 (0.57 to 1.78)<br>[p = 0.98] | 117 (51.8%) | 61 (49.6%) | 56 (54.4%) | OR = 0.99 (0.60 to 1.64)<br>[p = 0.97] |
| 2. Not hospitalized, limitation on activities | 49 (28.3%) | 29 (31.9%) | 20 (24.4%) |  | 67 (29.6%) | 42 (34.1%) | 25 (24.3%) |  |
| 3. Hospitalized, not requiring supplemental oxygen | 8 (4.6%) | 4 (4.4%) | 4 (4.9%) |  | 11 (4.9%) | 6 (4.9%) | 5 (4.9%) |  |
| 4. Hospitalized, requiring supplemental oxygen | 1 (0.6%) | 0 (0.0%) | 1 (1.2%) |  | 1 (0.4%) | 0 (0.0%) | 1 (1.0%) |  |
| 5. Hospitalized, on non-invasive ventilation or high flow oxygen devices | 0 (0.0%) | 0 (0.0%) | 0 (0.0%) |  | 1 (0.4%) | 1 (0.8%) | 0 (0.0%) |  |
| 6. Hospitalized, on invasive mechanical ventilation or ECMO | 1 (0.6%) | 1 (1.1%) | 0 (0.0%) |  | 1 (0.4%) | 1 (0.8%) | 0 (0.0%) |  |
| 7. Death | 26 (15.0%) | 12 (13.2%) | 14 (17.1%) |  | 28 (12.4%) | 12 (9.8%) | 16 (15.5%) |  |
| **Time to sustained recovery through Day 90 (days)** | 22.0 [19.0-27.0] | 22.0 [19.0-27.5] | 21.0 [18.0-27.0] | HR=0.98 (0.71 to 1.36)<br>[p = 0.92] | 21.0 [18.0-27.0] | 22.0 [19.0-27.5] | 21.0 [18.0-27.0] | HR=1.01 (0.77 to 1.34)<br>[p = 0.93] |
| **Change of NEWS-2 score from baseline to day 3 \*** | 0.0 [-1.0-1.0] | 0.0 [-1.0-1.0] | 0.0 [-1.0-1.0] | LSMD=0.04 (-0.75 to 0.84)<br>[p = 0.92] | 0.0 [-1.0-1.0] | 0.0 [-1.0-1.0] | 0.0 [-2.0-1.0] | LSMD=-0.19 (-0.91 to 0.53)<br>[p = 0.60] |
|  | Overall<br>(N = 173) | Tixagevimab-cilgavimab<br>(N = 91) | Placebo<br>(N = 82) | Effect measure<br>(95%CI) | Overall<br>(N = 226) | Tixagevimab-cilgavimab<br>(N = 123) | Placebo<br>(N = 103) | Effect measure<br>(95%CI) |
| Oxygenation-free days in the first 29 days | 23.0 [20.0-25.0] | 23.0 [20.0-25.0] | 23.0 [20.0-26.0] | LSMD=-0.36 (-2.12 to 1.40)<br>[p = 0.69] | 23.0 [20.0-26.0] | 23.0 [20.0-26.0] | 24.0 [20.0-26.0] | LSMD=-0.53 (-2.10 to 1.05)<br>[p = 0.51] |
| Days to new oxygen use, non-invasive ventilation or high flow oxygen devices during the first 29 days† | 4.5 [3.0-8.0] | 4.0 [2.0-11.0] | 5.0 [5.0-5.0] | HR=1.67 (0.17 to 16.68)<br>[p = 0.66] | 4.5 [3.0-8.0] | 4.0 [2.0-11.0] | 5.0 [5.0-5.0] | HR=1.96 (0.20 to 19.15) [p = 0.56] |
| Mechanical ventilation-free days in the first 29 days | 29.0 [29.0-29.0] | 29.0 [29.0-29.0] | 29.0 [29.0-29.0] | LSMD=0.61 (-0.94 to 2.16)<br>[p = 0.44] | 29.0 [29.0-29.0] | 29.0 [29.0-29.0] | 29.0 [29.0-29.0] | LSMD=0.43 (-0.88 to 1.73)<br>[p = 0.52] |
| Days to new invasive mechanical ventilation use during the first 29 days. | 7.0 [4.0-10.5] | 7.0 [4.0-10.0] | 8.0 [4.0-11.0] | HR=0.78 (0.39 to 1.57)<br>[p = 0.49] | 7.0 [4.0-10.5] | 7.0 [4.0-9.0] | 7.5 [4.0-11.0] | HR=0.65 (0.34 to 1.25)<br>[p = 0.19] |
| Mechanical ventilation or death between baseline and Day 15 | 29 (17.1%) | 14 (15.7%) | 15 (18.5%) | OR = 0.82 (0.37 to 1.83)<br>[p = 0.63] | 32 (14.3%) | 15 (12.4%) | 17 (16.7%) | OR = 0.70 (0.33 to 1.49)<br>[p = 0.35] |
| Days to hospital discharge before Day 90 | 8.0 [6.0-13.0] | 8.0 [6.0-11.0] | 9.0 [5.0-13.0] | HR=1.06 (0.78 to 1.45)<br>[p = 0.70] | 8.0 [5.0-13.0] | 8.0 [5.0-12.0] | 8.0 [5.0-13.0] | HR=1.10 (0.84 to 1.43)<br>[p = 0.49] |
| In hospital mortality rate | 21 (12.1%) | 9 (9.9%) | 12 (14.6%) | OR = 0.64 (0.25 to 1.61)<br>[p = 0.34] | 23 (10.2%) | 9 (7.3%) | 14 (13.6%) | OR = 0.49 (0.20 to 1.19)<br>[p = 0.11] |
| Mortality rate at Day 29 | 23 (13.3%) | 10 (11.0%) | 13 (15.9%) | OR = 0.65 (0.26 to 1.59)<br>[p = 0.35] | 25 (11.1%) | 10 (8.1%) | 15 (14.6%) | OR = 0.50 (0.21 to 1.19)<br>[p = 0.11] |
| Mortality rate at Day 90 | 26 (15.0%) | 12 (13.2%) | 14 (17.1%) | OR = 0.73 (0.31 to 1.72)<br>[p = 0.47] | 28 (12.4%) | 12 (9.8%) | 16 (15.5%) | OR = 0.56 (0.25 to 1.28)<br>[p = 0.17] |
| Occurrence of new hospitalization between discharge from index hospital and Day 90 | 30 (17.3%) | 18 (19.8%) | 12 (14.6%) | OR = 1.44 (0.65 to 3.21)<br>[p = 0.37] | 45 (19.9%) | 29 (23.6%) | 16 (15.5%) | OR = 1.68 (0.85 to 3.31)<br>[p = 0.13] |
| Confirmed re-infection with SARS-CoV-2 between discharge from index hospital and Day 90 | 4 (2.9%) | 2 (2.7%) | 2 (3.1%) | OR = 0.86 (0.12 to 6.24)<br>[p = 0.88] | 5 (2.6%) | 3 (2.8%) | 2 (2.4%) | OR = 1.11 (0.18 to 6.84)<br>[p = 0.91] |

Data are median [IQR] or n(%).
CI: confidence interval; OR: odds ratio; HR: hazard ratio; ECMO: extracorporeal membrane oxygenation; LSMD: least squares mean difference.
Analyses were stratified on the vaccination status at randomization and adjusted effect measures are reported in the table. For the ordinal scale results, an odds ratio above 1 is in the direction of tixagevimab-cilgavimab being better than placebo. For time to new invasive mechanical ventilation use during the first 29 days, a hazard ratio below 1 is in the direction of tixagevimab-cilgavimab being better than placebo. For other time to event analyses, a hazard ratio above 1 is in the direction of tixagevimab-cilgavimab being better than placebo ECMO, extracorporeal membrane oxygenation; OR, Odds ratio; HR, Hazard ratio; LSMD, least-square mean difference. Estimates are reported with their 95% confidence interval.
\* Data for NEWS-2 score was missing for 132/226 participants at day 8, 195/226 at day 15, and 215/226 at day 29.
† This outcome was evaluated only in participants not under oxygen support, non-invasive ventilation or high flow oxygen devices at randomization.

**Figure 2.**
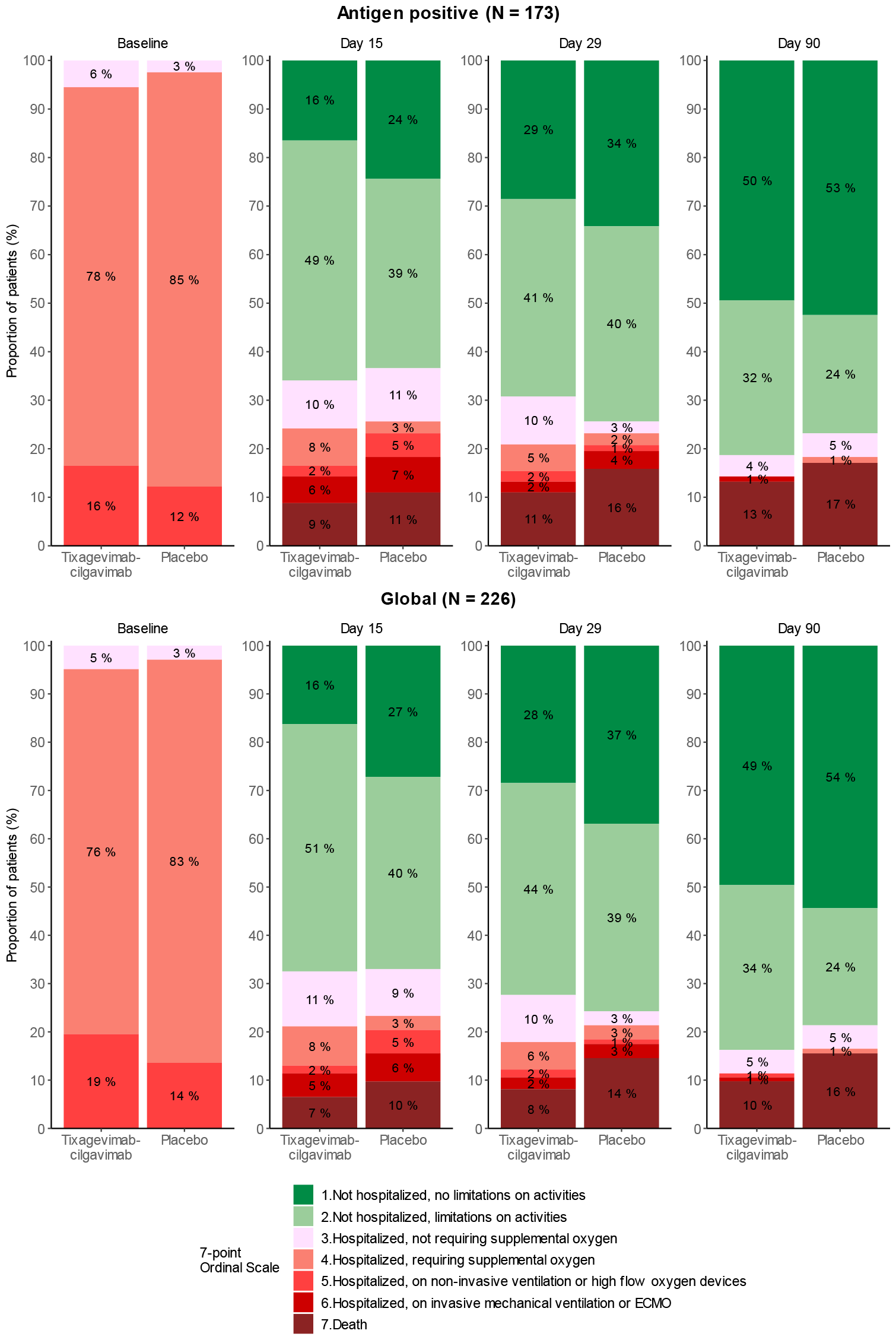
Clinical status on the WHO 7-point ordinal scale at baseline, day 15, day 29, day 90, in the antigenic positive and global modified intention-to-treat populations, according to the treatment group.

Normalized SARS-CoV-2 viral load, measured in NP swabs, were available for 205 (91%) patients at baseline, for 191 (85%) patients at day 3, for 114 (50%) patients at day 8, for 49 (22%) patients at day 15, and for 128 (57%) patients at day 29. There was no significant difference in viral kinetics from baseline to day 29 between the tixagevimab-cilgavimab and control groups, in both the antigen positive and global mITT populations. Accordingly, there was no significant difference between the two groups in the proportion of participants with detectable viral load at each time point, or in the time to first undetectable quantitative viral load through day 29 (figure 3; figure S5; tables S2 and S3, appendix).

**Figure 3.**
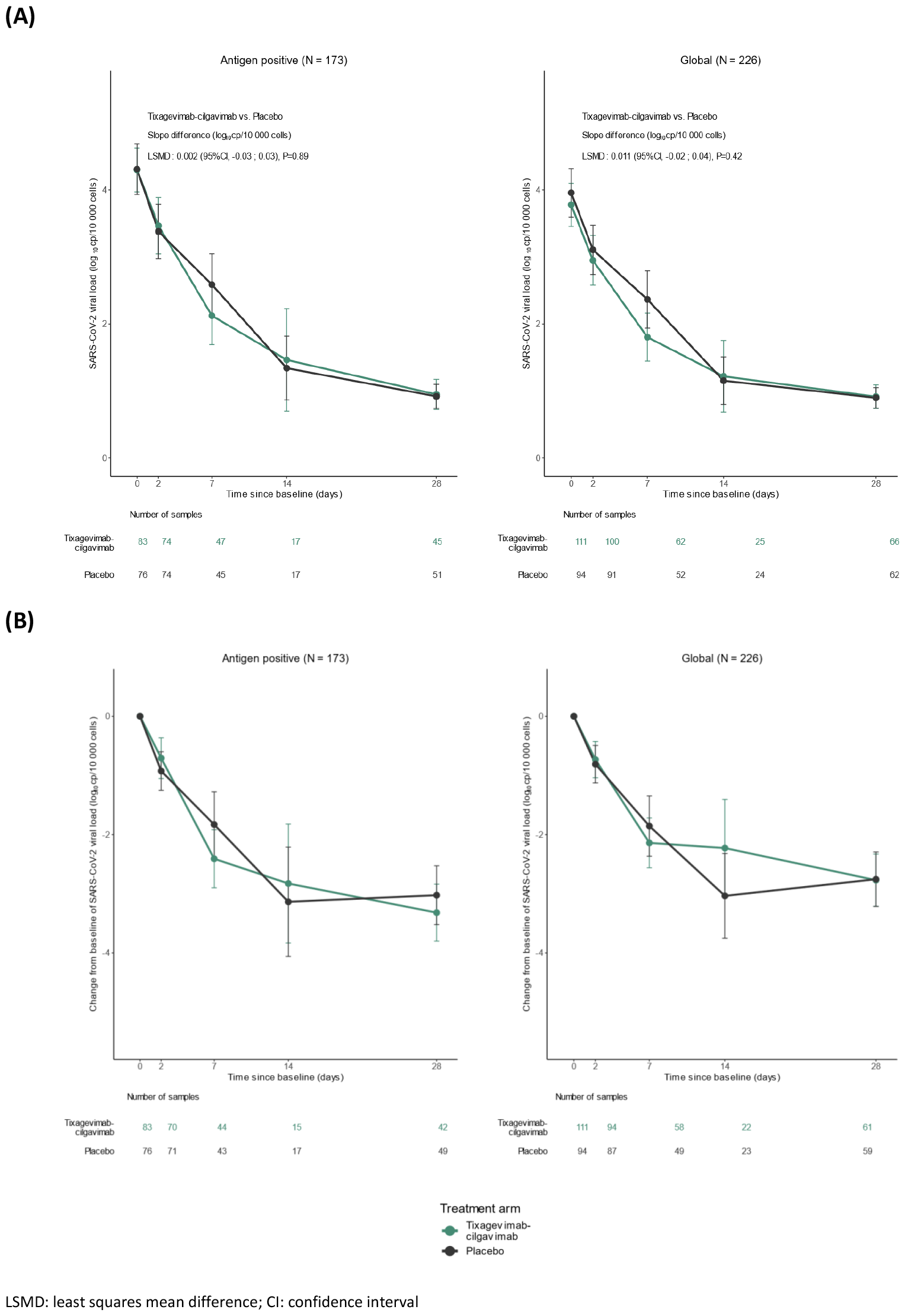
Normalised SARS-CoV-2 viral loads (A) and change from baseline of normalised SARS-CoV-2 viral loads (B) in nasopharyngeal swabs in the antigenic positive and global modified intention-to-treat populations, at each timepoint, according to the treatment group.

Neutralizing antibody titers were available for 198/226 (87.6%) patients at baseline, 179/226 (79.2%) at day 3, and 112/226 (49.5%) at day 8 (table S4, appendix). The proportion of subjects with undetectable neutralizing antibodies in the tixagevimab-cilgavimab arm dropped from 51.9% (56/108) at baseline, to 1% (1/97) at day 3, and 0% (0/60) at day 8, whereas in the placebo arm it dropped from 54.4% (49/90) to 30.5% (25/82) at day 3, and 13.5% (7/52) at day 8 (p < 10^−9^ at day 3 and p = 0.006 at day 8). Figure 4 shows the evolution of the log_10_ neutralizing antibody titers from baseline to day 8. There was a significantly greater increase in neutralizing antibodies in the tixagevimab-cilgavimab group compared to the placebo group both at day 3 (LSMD: 1.44, 95% CI [1.20 - 1.68], p < 10^−23^) and day 8 (LSMD: 0.91, 95% CI [0.64 - 1.18], p < 10^−8^).

**Figure 4:**
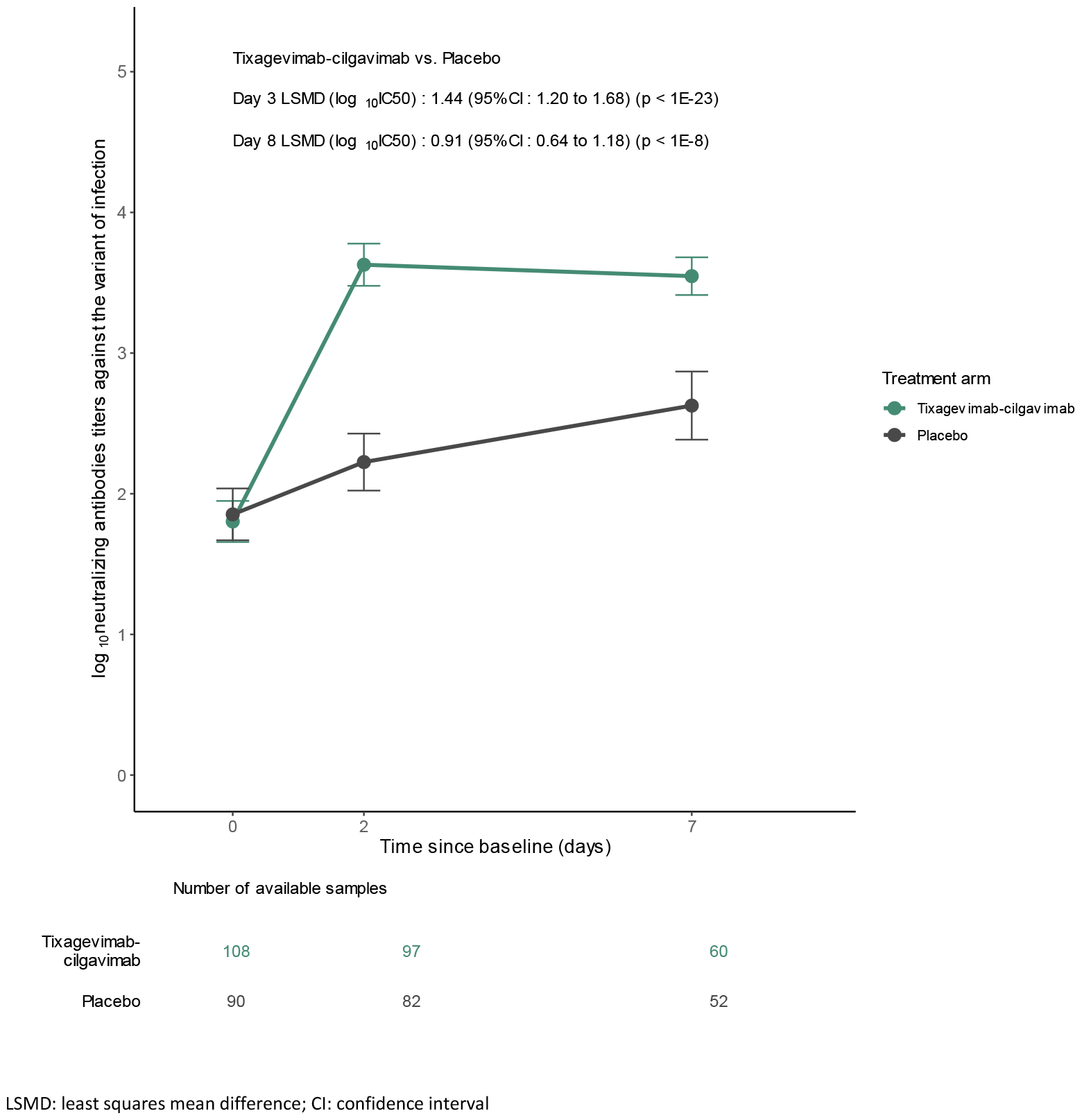
Evolution of log_10_ neutralizing antibody titers against variant of infection, from baseline to day 8 in patients from the global modified intention-to-treat population with known variant of infection at baseline.

Subgroup analyses showed that the difference in the increase of neutralizing antibodies was most important for patients infected with a pre-omicron variant, both at day 3 (LSMD: 1.94, 95% CI [1.67 - 2.20], p < 10^−25^) and day 8 (LSMD: 1.17, 95% CI [0.87 - 1.47], p < 10^−9^), whereas it was not significant for patients infected with omicron BA.1, and reduced for patients infected with BA.2/5. The interactions between the effects of the treatment and the variant at inclusion were significant at days 3 and 8 (p < 10^−7^ and p=0.01, respectively). The interactions with the vaccination status or the serology were also both significant at day 3 (p = 0.006 and p = 0.004), but not at day 8 (figure 5; figure S7, appendix). All prespecified subgroup analyses, and post-hoc analyses of clinical and virological endpoints in the subgroup of patients infected with pre-omicron are presented in the appendix.

**Figure 5.**
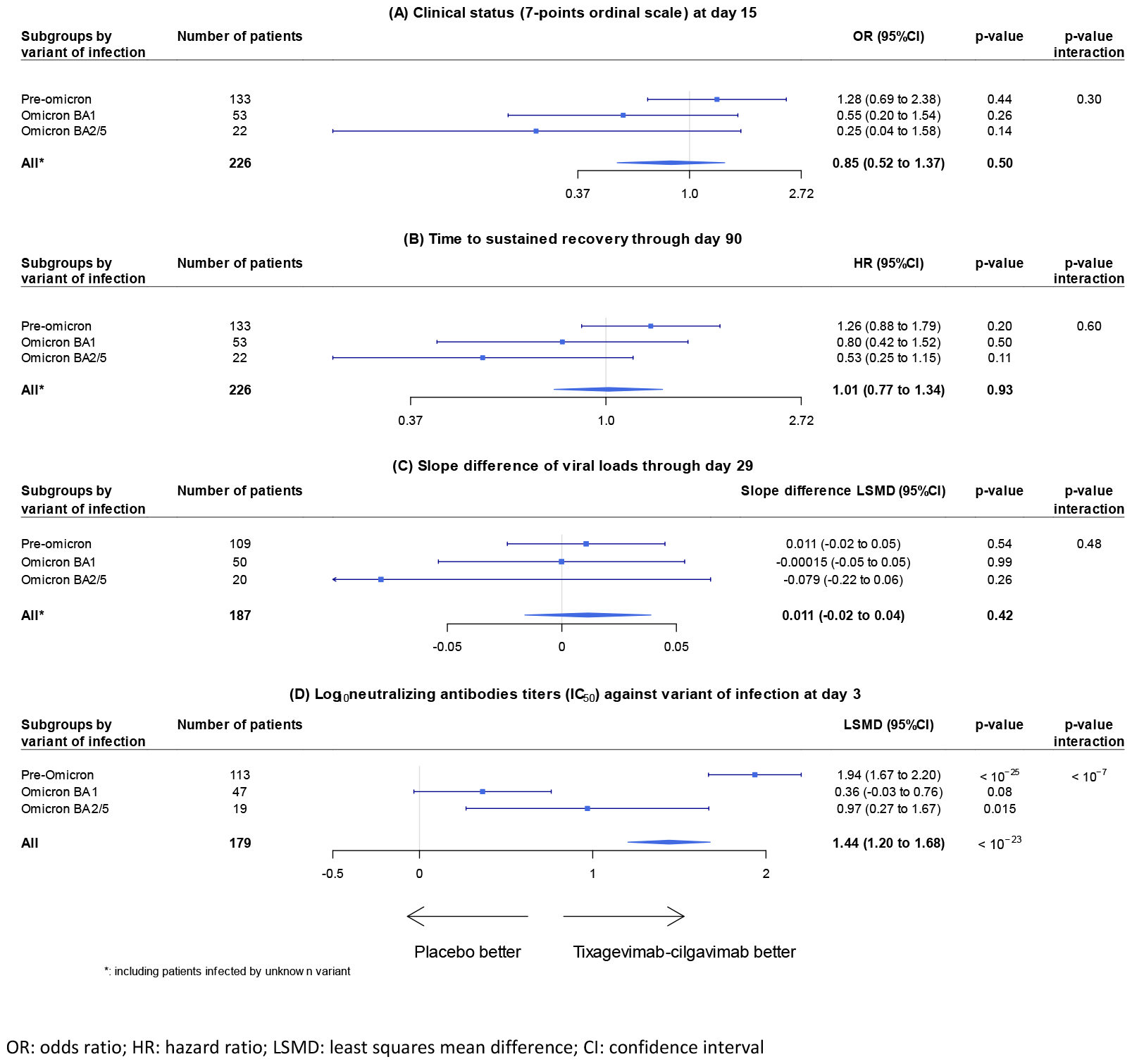
Forest plots of subgroup analyses according to the variant of infection, for the (A) clinical status (7-points ordinal scale) at day 15, (B) time to sustained recovery through day 90, (C) slope difference of viral loads through day 29, (D) log_10_ neutralizing antibody titers against variant of infection at day 3, in patients from the global modified intention-to-treat population.

Safety outcomes are shown in table 3. A total of 178 adverse events (AEs), including 90 serious AEs (SAEs) were reported, of which 28 (31.1%) were considered related to the investigational medicinal product. No grade 1-2 hypersensitivity-related and infusion-related AE until day 29 was reported, and one adverse event of special interest (AESI) was observed in the tixagevimab-cilgavimab group (anaphylactic shock), but considered not linked to the investigational product. In the tixagevimab-cilgavimab group, 51/123 (41.5%) patients had at least one AE, against 45/103 (43.7%) in the control group (p = 0.70); 30 (24.4%) patients in the tixagevimab-cilgavimab group had at least one grade 3 or 4 AE, against 33 (32.0%) in the control group (p = 0.18); 28 (22.8%) patients in the tixagevimab-cilgavimab group had at least one SAE, against 32 (31.1%) in the control group, although the difference was not statistically significant (p = 0.13). Among 19 fatal SAEs overall, none were of cardiac origin. The post-hoc subgroup analyses of SAEs suggested that tixagevimab-cilgavimab was safe regardless of serostatus or vaccination. It also showed, for patients infected with a pre-omicron variant, a statistically significant lower number of patients with at least one SAE in the tixagevimab-cilgavimab group, 10/70 (14.3%) against 22/63 (34.92%) in the control group (p = 0.01; figure S11, appendix).

**Table 3.**
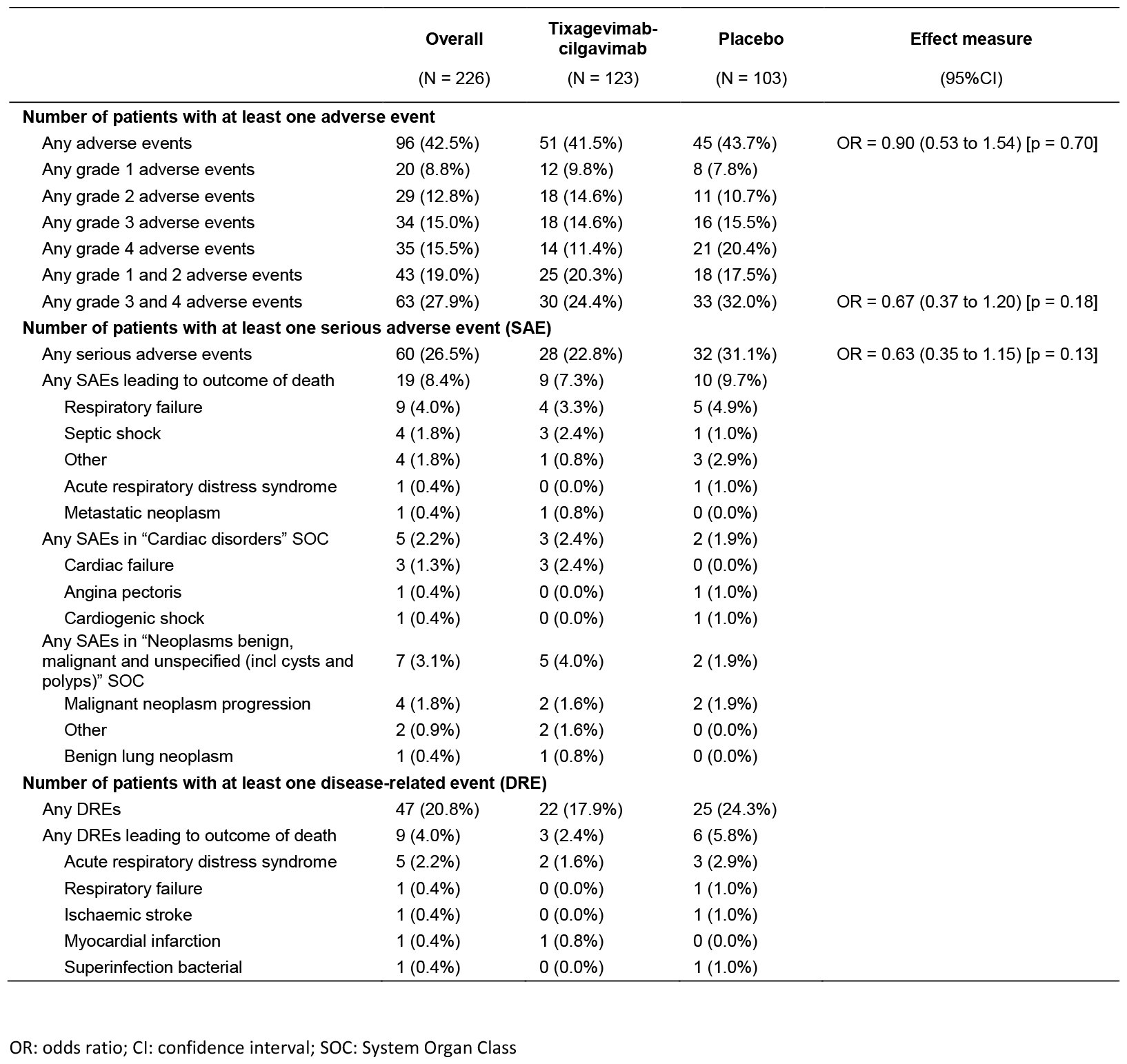
Summary of the number of patients with at least one adverse event, serious adverse event, disease-related event, through day 90 in the global modified intention to treat population, overall and according to the treatment group. Some patients had more than one adverse event (AE) or serious adverse event (SAE). As per protocol, AEs and SAEs do not include disease-related events (DRE).

## Discussion

Here we report the results of the DisCoVeRy multicentric, platform trial comparing intravenous tixagevimab-cilgavimab plus SoC to placebo plus SoC in hospitalized patients with COVID-19, mostly infected with pre-omicron SARS-CoV-2 variants. The trial was terminated early, after only 237 of the 1240 intended inclusions, due to novel *in-vitro* evidence showing loss of significant neutralization activity against omicron circulating variants.^18,19^ Tixagevimab-cilgavimab administration was well tolerated but was neither associated with a better clinical outcome at day 15, nor with improved sustained recovery up to day 90, nor with faster viral clearance, despite a significant increase in neutralizing antibodies against the SARS-CoV-2 variants infecting individuals at days 3 and 8 in patients who received tixagevimab-cilgavimab compared to placebo.

Our results differ from those of the ACTIV-3-TICO trial, the only other randomized controlled trial that has evaluated tixagevimab-cilgavimab in hospitalized patients with COVID-19.^13^ The trial found a 30% relative risk 90-day reduction in mortality (9% vs. 12%; hazard ratio [HR] 0.70 [95%CI 0.50-0.97]; p=0.032). Differences in outcome results may be due to the underpowered nature of the DisCoVeRy trial, but also due to differences in patient populations, their management, and the infecting SARS-CoV-2 variants. Patients, in the DisCoVery trial, were older (median age of 66 vs. 55), and more often male (69% vs. 58%), immunocompromised (20% vs. 9%), with at least one comorbidity (78% vs. 61%), more often vaccinated against COVID-19 (47% vs. 26%), and with a higher seroprevalence for SARS-CoV-2 at baseline (60% vs. 53%) than in the ACTIV-3-TICO trial. Patients in the DisCoVery trial were more severely ill at baseline, with only 4% of patients not requiring supplemental oxygen compared to 23% in the ACTIV-3-TICO trial, with an observed overall greater mortality up until day 90 of 15% compared to 10%, respectively. When considering SoC, 61% of patients in the ACTIV-3-TICO trial received concomitant remdesivir compared to none in the DisCoVeRy trial as remdesivir was not yet considered SoC in Europe at the time of enrolment. Results of the individual patient data meta-analysis of randomized controlled trials on remdesivir later offered more conclusive evidence of remdesivir efficacy in hospitalized patients with COVID-19 not needing mechanical ventilation.^6^ Although the delta variant was the predominant one in both trials, the DisCoVeRy trial enrolled 40% of patients infected with the omicron variant compared to none in the ACTIV-3-TICO trial.^13^ The SARS-CoV-2 omicron variant has been associated with the spread of multiple sub-lineages. They harbor different spike mutations that make them more evasive to vaccine or infection-induced antibodies including therapeutic mAbs. Nevertheless, although infections due to the omicron variant have overall proven to be less virulent than prior variants including delta,^20^ many patients who did require hospitalization were still at risk of dying, as illustrated by the mortality rate of 15% at day 90. A significant neutralizing activity against SARS-CoV-2 was measured in serum taken on days 3 and 8 from patients infected with pre-omicron variants that received tixagevimab-cilgavimab, but was not the case for those infected by omicron variants. This is consistent with *in-vitro* live-virus neutralization assays showing enhanced neutralization escape to tixagevimab-cilgavimab by SARS-CoV-2 serial omicron sub-lineages in comparison to the ancestral strain or delta variant.^18,19^

Results of this trial concerning safety of tixagevimab-cilgavimab should also be considered in-light of other randomized trials and reports of real-life experience. A warning was issued following the post-hoc analyses of the PROVENT pre-exposure prevention trial for SARS-CoV-2; a higher proportion of cardiovascular SAEs (myocardial infarction, cardiac arrhythmia or cardiac failure) were observed in individuals with cardiac risk factors who had received tixagevimab-cilgavimab compared to placebo.^21,22^ Nevertheless, no significant differences in any SAEs, nor in cardiac SAEs between patients who received tixagevimab-cilgavimab (32/123 ; 26% and 5/123 ; 4%, respectively) and those who received placebo (35/103 ; 34% and 4 /123 ; 4%, respectively) were observed in the DisCoVeRy trial. Furthermore, in the post-hoc subgroup analyses of cumulative incidence of SAEs, two times fewer SAEs were observed in the tixagevimab-cilgavimab group compared to the placebo group in patients infected with a pre-omicron variant. These observations are in line with safety results from the ACTIV-3-TICO trial^13^ as well as a population-based propensity-matched cohort study including aggregated healthcare records from 78 million patients in the USA.^23^

The randomized placebo-controlled design, the inclusion of patients vaccinated against SARS-CoV-2, and those infected by other than pre-omicron SARS-CoV-2 variants, the collection of data on normalized viral loads, and on neutralizing antibody titers are strengths of our study. Nevertheless, the final trial size was underpowered to compare safety, clinical efficacy, and virological effects of tixagevimab-cilgavimab to placebo. The early termination of this trial, with only 19% of the planned enrolment, reflects the difficulties to evaluate mAbs in the ever-changing landscape of a pandemic due to a virus with evolving genomics.^24,25^ Already by mid-2022, mAbs previously approved by the US Food and Drug Administration and the European Medicines Agency for clinical use were no longer recommended. Thus, clinical investigators were wary of including patients in our trial when new *in-vitro* evidence showed decreased neutralizing effects against emerging variants.^19^ Further reflections are needed on how best to evaluate mAbs in the clinical setting during a pandemic to rapidly bring new therapeutic options to patients, while preserving patients’ safety. Novel approaches for evaluating mAbs could be considered, such as immuno-bridging trials where correlates of protection such as humoral/ and or cellular immune parameters are evaluated in a controlled trial to establish whether an intervention is effective,^26^ and the adoption of a global surveillance system with criteria for *in-vitro* evaluation of antiviral susceptibility correlated to clinical data.^27^ Modeling antiviral effects while taking into account vaccination status, SARS-CoV2 antibody levels, infecting viral strains, and other parameters, may also provide better insight into viral efficacy of mAbs, as we have previously shown with remdesivir.^28^ It remains to better determine which patients would best benefit clinically from an active antiviral therapy in the hospital setting: is it the NP viral load, the time to symptoms onset, the SARS-CoV2 serological status of the patient, the presence of an immunodeficiency, or other factors that should guide clinicians?^29^

## Data Availability

With publication, deidentified, individual participant data that underlie this Article, along with a data dictionary describing variables in the dataset, will be made available to researchers whose proposed purpose of use is approved by the DisCoVeRy Steering Committee. To request the dataset, please address directly to the corresponding author or to the sponsor's representative to obtain a data access form. All requests will be evaluated by the Trial Management Team and the DisCoVeRy Steering Committee. For accepted requests, data will be shared after signing a data transfer agreement with the study sponsor. Data will be shared directly or through access on the INSERM repository. Related documents, such as the study protocol, statistical analysis plan, and informed consent form, will be made available (with publication) on request to the corresponding author or to the sponsor's representative. The data will be open access for the informed consent form, protocol, and statistical analysis plan.

## Contributors

FA, MH, CD, NPS, MBD, DC, YY, CB, CM, and FM were involved in the design, establishment, and day-to-day management and implementation of the trial. FA, MH, CD, DC, YY and FM obtained funding for the trial. MH, SJ, FG, FD, JR, NdC, DG, ELL, KL, VT, EF, DM, TS, JC, FCR, AMDR, PL, GMB, CR, KA, VLM, LP, PS, XL, MT, PC, OD, SG, MG, PL, FVB, CV, LB, EBN, AC, AK, FL, ERN, SS, GT, NPS, FA included participants in the trial. MBD, AG, MAT, GD and LJ were responsible for the virological analyses. PP and GC were responsible for the immunological analyses ; IB and SDD were involved in the seroneutralization analyses. ADi, NM, VT were responsible of the safety monitoring and analyses. SCD, ADe, CD, HE, CFL, SlM, AM, and MN were in charge of trial promotion and data monitoring. ST, OK, NB, and BL were responsible for the biobanking. MH, JE, RG, EH, MHa, JAP, AP, JRB, TS, KT, MT, ST, SU, GC, ADi, MBD, HE, DC, YY, FM, and FA were voting members of the trial steering committee. THLH, CM, NB, DB, JG, and FM were involved in the statistical analyses. CM, FM, MH, and FA wrote the original draft of the manuscript, which was reviewed and edited by NPS, NM, AG, PP, GC, JG, and DC. MH, CM, DB, NB, THLH, CD, CFL, ADe, and AM had full access to the blinded data. NB had full access to unblinded data. All authors contributed to refinement of and approved this manuscript. The corresponding author had full access to all the data in the study and had final responsibility for the decision to submit for publication

## Acknowledgements

This work received funding from several sources: the European Commission (EU-Response, Grant 101015736), the DIM One Health Île-de-France (R20117HD) and Astra-Zeneca. We thank all participants who consented to enroll in the trial, as well as all study and site staff whose indispensable assistance made the conduct of the DisCoVeRy trial possible (all listed in the appendix, pp 27-36)

## References

1. United Nations. WHO chief declares end to COVID-19 as a global health emergency | UN News [Internet]. 2023 [cited 2023 Nov 3]. Available from: https://news.un.org/en/story/2023/05/1136367

2. The RECOVERY Collaborative Group. Dexamethasone in Hospitalized Patients with Covid-19. N Engl J Med. 2021 Feb 25;384(8):693–704.

3. The WHO Rapid Evidence Appraisal for COVID-19 Therapies (REACT) Working Group, Sterne JAC, Murthy S, Diaz JV, Slutsky AS, Villar J, et al. Association Between Administration of Systemic Corticosteroids and Mortality Among Critically Ill Patients With COVID-19: A Meta-analysis. JAMA. 2020 Oct 6;324(13):1330.

4. Information on COVID-19 Treatment, Prevention and Research [Internet]. COVID-19 Treatment Guidelines. [cited 2023 Nov 3]. Available from: https://www.covid19treatmentguidelines.nih.gov/

5. World Health Organization. Clinical management of COVID-19: living guideline [Internet]. World Health Organization; 2023 Aug [cited 2023 Nov 3]. Report No.: WHO/2019-nCoV/clinical/2023.2. Available from: https://iris.who.int/bitstream/handle/10665/372288/WHO-2019-nCoV-clinical-2023.2-eng.pdf?sequence=1

6. Amstutz A, Speich B, Mentré F, Rueegg CS, Belhadi D, Assoumou L, et al. Effects of remdesivir in patients hospitalised with COVID-19: a systematic review and individual patient data meta-analysis of randomised controlled trials. Lancet Respir Med. 2023 May;11(5):453–64.

7. Montgomery H, Hobbs FDR, Padilla F, Arbetter D, Templeton A, Seegobin S, et al. Efficacy and safety of intramuscular administration of tixagevimab–cilgavimab for early outpatient treatment of COVID-19 (TACKLE): a phase 3, randomised, double-blind, placebo-controlled trial. Lancet Respir Med. 2022 Oct;10(10):985–96.

8. Gupta A, Gonzalez-Rojas Y, Juarez E, Crespo Casal M, Moya J, Falci DR, et al. Early Treatment for Covid-19 with SARS-CoV-2 Neutralizing Antibody Sotrovimab. N Engl J Med. 2021 Nov 18;385(21):1941–50.

9. Self WH, Sandkovsky U, Reilly CS, Vock DM, Gottlieb RL, Mack M, et al. Efficacy and safety of two neutralising monoclonal antibody therapies, sotrovimab and BRII-196 plus BRII-198, for adults hospitalised with COVID-19 (TICO): a randomised controlled trial. Lancet Infect Dis. 2022 May;22(5):622–35.

10. ACTIV-3/TICO LY-CoV555 Study Group. A Neutralizing Monoclonal Antibody for Hospitalized Patients with Covid-19. N Engl J Med. 2021 Mar 11;384(10):905–14.

11. Abani O, Abbas A, Abbas F, Abbas M, Abbasi S, Abbass H, et al. Casirivimab and imdevimab in patients admitted to hospital with COVID-19 (RECOVERY): a randomised, controlled, open-label, platform trial. The Lancet. 2022 Feb;399(10325):665–76.

12. Somersan-Karakaya S, Mylonakis E, Menon VP, Wells JC, Ali S, Sivapalasingam S, et al. Casirivimab and Imdevimab for the Treatment of Hospitalized Patients With COVID-19. J Infect Dis. 2022 Dec 28;227(1):23–34.

13. Holland TL, Ginde AA, Paredes R, Murray TA, Engen N, Grandits G, et al. Tixagevimab– cilgavimab for treatment of patients hospitalised with COVID-19: a randomised, double-blind, phase 3 trial. Lancet Respir Med. 2022 Oct;10(10):972–84.

14. Carabelli AM, Peacock TP, Thorne LG, Harvey WT, Hughes J, COVID-19 Genomics UK Consortium, et al. SARS-CoV-2 variant biology: immune escape, transmission and fitness. Nat Rev Microbiol [Internet]. 2023 Jan 18 [cited 2023 Nov 3]; Available from: https://www.nature.com/articles/s41579-022-00841-7

15. Ader F, Bouscambert-Duchamp M, Hites M, Peiffer-Smadja N, Poissy J, Belhadi D, et al. Remdesivir plus standard of care versus standard of care alone for the treatment of patients admitted to hospital with COVID-19 (DisCoVeRy): a phase 3, randomised, controlled, open-label trial. Lancet Infect Dis. 2022 Feb;22(2):209–21.

16. Ader F, Peiffer-Smadja N, Poissy J, Bouscambert-Duchamp M, Belhadi D, Diallo A, et al. An open-label randomized controlled trial of the effect of lopinavir/ritonavir, lopinavir/ritonavir plus IFN-β-1a and hydroxychloroquine in hospitalized patients with COVID-19. Clin Microbiol Infect. 2021 Dec;27(12):1826–37.

17. Ader F. An open-label randomized, controlled trial of the effect of lopinavir and ritonavir, lopinavir and ritonavir plus interferon-β-1a, and hydroxychloroquine in hospitalized patients with COVID-19: final results. Clin Microbiol Infect. 2022 Sep;28(9):1293–6.

18. Takashita E, Yamayoshi S, Simon V, Van Bakel H, Sordillo EM, Pekosz A, et al. Efficacy of Antibodies and Antiviral Drugs against Omicron BA.2.12.1, BA.4, and BA.5 Subvariants. N Engl J Med. 2022 Aug 4;387(5):468–70.

19. Touret F, Giraud E, Bourret J, Donati F, Tran-Rajau J, Chiaravalli J, et al. Enhanced neutralization escape to therapeutic monoclonal antibodies by SARS-CoV-2 omicron sublineages. iScience. 2023 Apr;26(4):106413.

20. Fan Y, Li X, Zhang L, Wan S, Zhang L, Zhou F. SARS-CoV-2 Omicron variant: recent progress and future perspectives. Signal Transduct Target Ther. 2022 Apr 28;7(1):141.

21. Levin MJ, Ustianowski A, De Wit S, Launay O, Avila M, Templeton A, et al. Intramuscular AZD7442 (Tixagevimab–Cilgavimab) for Prevention of Covid-19. N Engl J Med. 2022 Jun 9;386(23):2188–200.

22. FDA. FACT SHEET FOR HEALTHCARE PROVIDERS: EMERGENCY USE AUTHORIZATION FOR EVUSHELD™ (tixagevimab co-packaged with cilgavimab) [Internet]. 2023 Jan [cited 2023 Nov 3]. Available from: https://www.fda.gov/media/154701/download

23. Birabaharan M, Hill E, Begur M, Kaelber DC, Martin TCS, Mehta SR. Cardiovascular Outcomes After Tixagevimab and Cilgavimab use for Pre-Exposure Prophylaxis Against Coronavirus Disease 2019: A Population-Based Propensity-matched Cohort Study. Clin Infect Dis Off Publ Infect Dis Soc Am. 2023 Apr 17;76(8):1500–3.

24. Karim SSA, Karim QA. Omicron SARS-CoV-2 variant: a new chapter in the COVID-19 pandemic. The Lancet. 2021 Dec;398(10317):2126–8.

25. European Centre for Disease Prevention and Control. SARS-CoV-2 variants of concern as of 20 October 2023 [Internet]. [cited 2023 Nov 3]. Available from: https://www.ecdc.europa.eu/en/covid-19/variants-concern

26. European Medical Agency. Summary report of the Joint EMA-FDA workshop on the efficacy of monoclonal antibodies in the context of rapidly evolving SARS-CoV-2 variants [Internet]. [cited 2023 Nov 3]. Available from: https://www.ema.europa.eu/en/documents/report/summary-report-joint-ema-fda-workshop-efficacy-monoclonal-antibodies-context-rapidly-evolving-sars_en.pdf

27. European Centre for Disease Prevention and Control. SARS-CoV-2 variant mutations conferring reduced susceptibility to antiviral drugs and monoclonal antibodies: a non systematic literature review for surveillance purposes. [Internet]. LU: Publications Office; 2023 [cited 2023 Nov 3]. Available from: https://data.europa.eu/doi/10.2900/192733

28. Lingas G, Néant N, Gaymard A, Belhadi D, Peytavin G, Hites M, et al. Effect of remdesivir on viral dynamics in COVID-19 hospitalized patients: a modelling analysis of the randomized, controlled, open-label DisCoVeRy trial. J Antimicrob Chemother. 2022 Apr 27;77(5):1404–12.

29. Néant N, Lingas G, Gaymard A, Belhadi D, Hites M, Staub T, et al. Association between SARS-CoV-2 viral kinetics and clinical score evolution in hospitalized patients. CPT Pharmacomet Syst Pharmacol. 2023 Oct 11;psp4.13051.

