## Supplementary appendix for "Tixagevimab-cilgavimab (AZD7442) for the treatment of patients hospitalized with COVID-19 (DisCoVeRy): A phase 3, randomized, double-blind, placebo-controlled trial"

by Hites M., Massonnaud C.R., et al.

### Supplementary methods

#### Participants

##### Inclusion criteria:

- Adult ≥18 years of age at time of enrolment
- Hospitalized patients with any of the following criteria:
  - the presence of pulmonary rales/crackles on clinical exam OR
  - SpO2 ≤ 94% on room air OR
  - requirement of supplementary oxygen including high flow oxygen devices or non-invasive ventilation
- A time between onset of symptoms and randomization of less than 9 days (11 days from September 20th, 2021, protocol V15)
- A positive SARS-CoV-2 RT-PCR performed on a NP swab within the 5 days preceding randomization
- The result of a rapid antigen test performed on a NP swab within the 6 hours preceding randomization
- Contraceptive use by men or women
  - Male participants: Contraception for male participants is not required; however, to avoid the transfer of any fluids, all male participants must use a condom from Day 1 and agree to continue for 90 days following administration of IMP
  - Female participants: Women of child-bearing potential must agree to use contraception for 365 days following administration of IMP

##### Exclusion criteria:

- Refusal to participate expressed by patient or legally authorized representative if they are present
- Need for invasive mechanical ventilation and/or ECMO at the time of enrolment
- Spontaneous blood ALT/AST levels > 5 times the upper limit of normal
- Glomerular filtration rate (GFR) < 15 mL/min or requiring maintenance dialysis
- Pregnancy or breast-feeding
- Anticipated transfer to another hospital, which is not a study site within 72 hours following randomization
- Known history of allergy or reaction to any component of the study drug formulation
- Previous hypersensitivity, infusion-related reaction, or severe adverse reaction following administration of monoclonal or polyclonal antibodies.
- *Before September 20th, 2021:* any prior receipt of investigational or licensed vaccine
- Any prior receipt of investigational or licensed mAb indicated for the prevention of SARS-CoV-2 infection or COVID-19, and for those not vaccinated, expected receipt of vaccine in the 30 days following hospital discharge, according to current recommendation in each country.
- Any medical condition which, in the judgment of the investigator, could interfere with the interpretation of the trial results or that preludes to protocol adherence.

**Outcomes**

The primary endpoint was the clinical status at day 15 measured on the 7-point ordinal scale of the WHO Master Protocol (v3.0, March 3, 2020). The key secondary endpoint was the time from randomization to sustained recovery, defined as being discharged from the index hospitalization, followed by being alive and at home for 14 consecutive days prior to day 90. For patients with several discharges before day 90 and re-hospitalisation prior to achieving sustained recovery, the date of the first sustained recovery was considered. Patients discharged but for whom the status 14 days after discharge is not available were censored at the date of last news on vital status post discharge. Patients alive but with no sustained recovery before day 90 were censored at day 90, at the date of loss to follow-up, or at the date of withdrawal. Time to sustained recovery was calculated for patients who died before experiencing the outcome, because death was considered as a competitive risk.

**Virological and Immunological methods**

**Nucleic acid extraction**

Blood and respiratory samples preparation was done using STARlet workstation, (Hamilton©, Villebon-sur-Yvette, France) then RNA extraction was performed on the MGISP-960 using the MGIEasy Nucleic Acid Extraction Kit (BGI, Beijing, China). RNA was extracted from 160 µL of sample eluted in 50 µL of the elution buffer.

**Viral load methods**

After extraction, SARS-CoV-2 and cell detection and quantification were done by RT-PCR on QuantStudio 5 PCR Systems (Thermo Fisher Scientific, Waltham, Massachusetts, USA) using the COVID-19 R-GENE® assay (Argene BioMérieux, Marcy-l'Étoile, France). The SARS-CoV-2 and cell load were measured according to a scale of calibrated plasmid, specially developed by Argene BioMérieux and used according to the manufacturers' instructions. RT-PCR results were interpreted using Fastfinder software v4.6 (Ugentec, Hasselt, Belgium).

SARS-CoV-2 load in plasma samples was expressed as $\log_{10}$ RNA copies/mL. To allow the comparison of respiratory samples of different qualities (cell richness) we followed the same methodology of normalized viral load developed previously in the context of influenza virus infection. In brief, the SARS-CoV-2 viral load was normalized using the number of cells measured (quantification of HPRT-1 housekeeping gene) and expressed in copies per $10^4$ cells.

**Sequencing methods**

After extraction, SARS-CoV-2 sequencing was carried out using COVIDSeq-Test™ (Illumina, San Diego, USA) with Artic V3, V4 or V4.1 primers as they became available. Libraries were sequenced with 100 bp paired-end reads using the NovaSeq 6000 Sequencing system SP flow cell.

To interpret sequencing data, reads were processed using the in-house bioinformatic pipeline seqmet (available at https://github.com/genepii/seqmet). Briefly, trimming of paired reads was performed with cutadapt to remove sequencing adapters and low-quality ends. Alignment to the SARS-CoV-2 reference genome MN908947 was performed by minimap2. Duplicate reads tagged by picard were removed, remaining reads were realigned by abra2 to improve indel detection sensitivity and finally clipped with samtools ampliconclip to remove read ends containing primer sequences. Finally, variant calling was performed by freebayes.

**Serology methods**

The presence of anti-SARS-CoV-2 antibodies was evaluated on serum samples using the Atellica SARS-CoV-2 IgG assay (sCOVG, Siemens Healthineers, Erlangen, Germany), which detects and quantify IgG against RBD domain of the spike (S), and the Architect SARS-COV-2 IgG test (Abbott, Sligo, Ireland), which detects anti-nucleocapsid (N) IgG. Positivity was established according to the threshold value recommended by each manufacturer.

As a quantitative assay, the index value (U/mL) obtain for the sCOVG assay correspond to 21.80 BAU/mL using the World Health Organization (WHO) 1st International 1.0 Standard for anti-SARS-CoV-2 immunoglobulin (NIBSC code: 20/136). This assay has a measuring interval of 0.50–150.00 Index (U/mL) but samples under 1.00 Index (U/mL) were reported as non-reactive reactive according to the manufacturers' instructions. In addition, any sample with a result above 150.00 index (U/mL) was automatically diluted 10 times to achieve a maximum limit of quantification of 1500 index (U/mL), i.e. 32700 BAU/mL.

**Serum neutralization methods**

***Pseudovirus production and titration***

Neutralization assays were performed using lentiviral pseudotypes harboring the SARS-CoV-2 spike and encoding luciferase. Briefly, adherent HEK293T cells were co-transfected with gag/pol and luciferase plasmids, alongside different SARS-CoV-2 spike plasmids (Delta, BA-1 or BA-2) from InvivoGen (plv-SpikeV8, V11 and V12, respectively). Pseudovirus-containing supernatants were harvested after 72 h, centrifuged and filtered through a 0.45 μm filter. Supernatants were then concentrated 50 times on Amicon Ultra (MWCO 100KDa), aliquoted and stored at −80°C. Before use, pseudovirus preparations were tittered using HeLa ACE-2 cells to determine the appropriate dilution necessary to obtain about 150,000 relative light units (RLU) per well in 96-w white plates (Corning #3885).

***Neutralization assay***

Sequential 1/3 dilutions of serum, starting at a 1/10, and Evusheld, starting at varying concentrations depending on the pseudovirus type (Delta: 0.9µg/ml, BA-1: 60µg/ml, BA-2: 2 µg/ml), were incubated with pseudoviruses for 1 h at 37°C in 96-well white plates, before addition of HeLa ACE2 cells (8000 cells per well). Subsequently, plates were incubated for 24 h at 37°C, while being protected from evaporation. After this incubation period, 40µL of DMEM (Gibco 21969035) supplemented with 10% FBS (VWR 97068-086) were added to each well. Following another 24 h of incubation, the medium in each well was aspirated and replaced by 45 µL of 1x cell lysis buffer (OZ Bioscience # LUC1000) for a 60-min incubation under agitation. Thirty µL of luciferin substrate were then added and relative light units (RLU) were measured immediately using a luminometer. The IC50 values were calculated using GraphPad Prism (Version 10).

**Statistical methods**

**Sample size**

The sample size was determined assuming the following distribution of the ordinal scale status at day 15, under SoC: item 1 (not hospitalized, no limitations on activities), 24%; item 2, 35%; item 3, 7%; item 4, 17%; item 5, 6%; item 6, 6%; item 7 (death), 5%. These numbers were obtained using data from the DisCoVery trial between July 1st and December 31, 2020, specifically from 93 patients in the SoC arm, included in the moderate disease strata and with duration of symptoms of less than 9 days. The study was powered to detect an odds ratio of 1.5 (an OR greater than 1 indicates superiority of the experimental treatment over the control for each ordinal scale category), with a 90% power and an two-sided type I error of 0.05, for the primary endpoint, that is for patient with a positive antigen test at inclusion. We determined that 413 evaluable, antigen positive, patients were needed in each arm. Since antigen negative patients can represent as much as 30% of patients, 590 participants (antigen positive or negative) were needed in each arm. Accounting for 5% of non-evaluable patients, we needed to include a total of 620 patients per arm. Inclusion of vaccinated participants (allowed in protocol V15, from September 20th, 2021) could have an impact on the statistical power, depending on the ordinal scale distribution for vaccinated patients under SoC, the expected treatment effect in vaccinated patients, and the proportion of vaccinated patients. The proportion of vaccinated patients was capped at 20% of included participants. Power computations under various scenarios of outcome distribution and treatment effect resulted in a power always greater than 85% (supplementary material), therefore we decided not to change the number of subjects needed with the inclusion of vaccinated participants.

**Statistical tests**

The change of NEWS-2 score from baseline to days 3, 8, 15, and 29, and the number of oxygenation and ventilator-free days from baseline to day 29 were analyzed using ANCOVA models with the treatment group as effect. The time to new oxygen use, non-invasive ventilation or high flow oxygen devices during the first 29 days, and the time to new invasive mechanical ventilation use during the first 29 days were compared between the two groups using a Cox proportional hazards model. The incidence of new mechanical ventilation or death by day 15 in the two groups were compared using a stratified Cochran-Mantel-Haenszel (CMH) test. Time to hospital discharge was analyzed using the same approach as for the key secondary endpoint. Mortality rates were estimated with the Kaplan-Meier method and differences were analyzed using a stratified CMH test. Differences in occurrence of new hospitalization between discharge from index hospitalization and day 90, and occurrence of confirmed re-infection with SARS-CoV-2 between discharge and day 90 were analyzed using a stratified CMH test. For safety endpoints, the number of participants with at least one AE, with at least one grade 3 or 4 AE, and with at least one SAE were compared between groups using a CMH test.

**Handling of missing data**

Missing data for the ordinal scale was handled as follows : if the last scale value available before the missing data for a patient is 7 (death), or if a death date is available, the value for the ordinal scale will be imputed to 7 and carried forward for all subsequent endpoints. For non-hospitalized patients, if the last available value is 1 (not hospitalized, no limitations on activities) or 2 (hospitalized, with limitations on activities), it will be carried forward, otherwise, it will be imputed to 2. For hospitalized patients, a value of 3 (hospitalized, not requiring supplemental oxygen), 4 (hospitalized, requiring supplemental oxygen), 5 (hospitalized on non-invasive ventilation or high flow oxygen devices), or 6 (hospitalized, on invasive mechanical ventilation or extracorporeal membrane oxygenation) will be imputed based on the available clinical data on the day of the missing value, and carried forward until the next non-missing value, or the discharge date, or the date of death.

Missing data for the NEWS-2 score were handled as follows : missing values in variables needed for the NEWS-2 derivation were imputed to the mean if they were missing at baseline, otherwise the last observation was carried forward. Patients that died had their NEWS-2 score imputed to 20 (worst value) after the date of death.

Missing data for oxygen or mechanical ventilation use were imputed using the ordinal scale value, if it was available on the same day. Otherwise, the last observation was carried forward until the next non-missing value, or until the discharge date if discharged before day 29, or until death in case of death before day 29, or until day 29 in the other cases.

If a patient was discharged and no further hospitalization data is available, then the patient will be considered as not being re-hospitalised afterwards. If the ordinal scale value on a given day is "1. Not hospitalized, no limitations on activities" or "2. Not hospitalized, limitation on activities", but the value for the date of discharge is missing, it will be imputed to the day preceding the date on which the ordinal scale value was available.

If the ordinal scale value on a given day is "7. Death", but the date of death is missing, it will be imputed to the first date associated with the ordinal scale value "7. Death".

Missing data for variant of infection was handled as follows: if sequencing data for variant was missing at baseline, it was imputed using available sequencing data for variant at day 3 or day 8. If no sequencing data was available, the variant of infection was imputed to be the variant that was dominant in France at the time of the patient's symptoms onset, according to epidemiological surveys.[1] We identified three periods in which a single variant was dominant (estimated to represent more than 95% of infections): (a) pre-Omicron variant before December 9, 2021; (b) Omicron BA.1 from January 6, 2022 to January 24, 2022; (c) Omicron BA.2/5 after March 28, 2022. If the patient's date of symptom onset fell into a period of transition of circulating variants (from December 9, 2021 to January 6, 2022, or from January 24, 2022 to March 28, 2022), the variant of infection remained "unknown".

Missing viral loads were not imputed. Undetectable viral load values (i.e. values < 1 $log_{10}$ copies/10 000 cells) were imputed 0.7 $log_{10}$ copies/10 000 cells. For the models, in case of several consecutive undetectable values, only the first one was replaced and the subsequent values were discarded (until the next detectable value, if values were available afterwards).

Missing neutralizing antibody titers  were not imputed. For this analysis, the LOD was defined at a value of 30. Values below the LOD (either undetectable or < 30) were imputed to 15 in some analyses.

**Trial safety and monitoring**

There was no pre-planned interim analysis for efficacy nor futility, and none was performed. Data on adverse events and serious adverse events were reviewed monthly by the safety department. An independent data safety and monitoring board (DSMB) externally reviewed the trial data at regular intervals regarding treatment efficacy, safety, and futility. The DSMB gathered as planned after the inclusion of the first 100 patients, on April 20th, 2022, and recommended the continuation of the study.

The protocol specified a list of disease-related events (DREs) which refer to complications that are common in hospitalized patients with COVID-19 and can be serious. DREs were systematically collected in the eCRF but did not require immediate reporting to the sponsor as SAEs, unless assessed as possibly related to the blinded IMP by the investigator.

This list included the following DREs: acute kidney injury/acute renal failure, grade 3 and above transaminase increased, pancreatitis, bacteraemia, bacterial pneumonia, cardiac arrhythmia, cardiac ischemia, cardiac arrest, endocarditis, myocarditis, pericarditis, congestive heart failure, acute respiratory distress syndrome, pneumothorax, pleural effusion, meningitis, encephalitis, stroke, coma, confusion, gastrointestinal hemorrhage, pulmonary thrombosis, arterial or deep vein thromboembolic events.

In the protocol, the adverse events of special interest (AESI) were defined as anaphylaxis and other serious hypersensitivity reactions, including immune complex disease, or grade 3 and 4 injection site reactions or infusion-related events. All AESIs required immediate notification to the sponsor.

**References**

1. Coronavirus : chiffres clés et évolution de la COVID-19 en France et dans le Monde [Internet]. [cited 2023 Nov 3]. Available from: https://www.santepubliquefrance.fr/dossiers/coronavirus-covid-19/coronavirus-chiffres-cles-et-evolution-de-la-covid-19-en-france-et-dans-le-monde

### Supplementary results

**Supplementary table S1: additional baseline characteristics of participants in the antigen positive and global modified intention-to-treat populations, overall and according to the treatment group.**

| | Antigen positive (N = 173) | | | Global (N = 226) | | |
|---|---|---|---|---|---|---|
| | **Overall** (N = 173) | **Tixagevimab-cilgavimab** (N = 91) | **Placebo** (N = 82) | **Overall** (N = 226) | **Tixagevimab-cilgavimab** (N = 123) | **Placebo** (N = 103) |
| **Median age, years** | 66.0 [55.0-79.0] | 65.0 [56.0-80.0] | 66.5 [55.0-78.0] | 66.0 [53.0-76.0] | 64.0 [53.0-76.0] | 68.0 [52.0-76.0] |
| **Sex male**, | 117 (67.6%) | 56 (61.5%) | 61 (74.4%) | 155 (68.6%) | 78 (63.4%) | 77 (74.8%) |
| **Geographical origin*** | | | | | | |
| Caucasian | 131 (78.9%) | 68 (76.4%) | 63 (81.8%) | 161 (74.9%) | 87 (73.7%) | 74 (76.3%) |
| North African | 17 (10.2%) | 10 (11.2%) | 7 (9.1%) | 32 (14.9%) | 19 (16.1%) | 13 (13.4%) |
| Sub saharian African | 4 (2.4%) | 3 (3.4%) | 1 (1.3%) | 6 (2.8%) | 4 (3.4%) | 2 (2.1%) |
| Latin American | 1 (0.6%) | 1 (1.1%) | 0 (0.0%) | 2 (0.9%) | 1 (0.8%) | 1 (1.0%) |
| West indian | 1 (0.6%) | 1 (1.1%) | 0 (0.0%) | 2 (0.9%) | 1 (0.8%) | 1 (1.0%) |
| Asian | 2 (1.2%) | 1 (1.1%) | 1 (1.3%) | 2 (0.9%) | 1 (0.8%) | 1 (1.0%) |
| Other | 10 (6.0%) | 5 (5.6%) | 5 (6.5%) | 10 (4.7%) | 5 (4.2%) | 5 (5.2%) |
| **Number of comorbidities*** | | | | | | |
| 0 | 38 (22.0%) | 17 (18.7%) | 21 (25.6%) | 52 (23.0%) | 28 (22.8%) | 24 (23.3%) |
| 1 | 49 (28.3%) | 26 (28.6%) | 23 (28.0%) | 71 (31.4%) | 38 (30.9%) | 33 (32.0%) |
| 2 | 43 (24.9%) | 23 (25.3%) | 20 (24.4%) | 50 (22.1%) | 27 (22.0%) | 23 (22.3%) |
| 3 | 25 (14.5%) | 14 (15.4%) | 11 (13.4%) | 32 (14.2%) | 18 (14.6%) | 14 (13.6%) |
| >3 | 18 (10.4%) | 11 (12.1%) | 7 (8.5%) | 21 (9.3%) | 12 (9.8%) | 9 (8.7%) |
| **Coexisting condition*** | | | | | | |
| Chronic cardiac disease | 62 (35.8%) | 39 (42.9%) | 23 (28.0%) | 72 (31.9%) | 43 (35.0%) | 29 (28.2%) |
| Obesity (BMI >= 30) | 46 (26.6%) | 27 (29.7%) | 19 (23.2%) | 63 (27.9%) | 38 (30.9%) | 25 (24.3%) |
| Diabetes | 46 (26.6%) | 30 (33.0%) | 16 (19.5%) | 58 (25.7%) | 37 (30.1%) | 21 (20.4%) |
| Current or former smoker | 27 (15.9%) | 14 (15.4%) | 13 (16.5%) | 31 (13.9%) | 15 (12.2%) | 16 (16.0%) |
| Current smoker | 13 (48.1%) | 6 (42.9%) | 7 (53.8%) | 15 (48.4%) | 7 (46.7%) | 8 (50.0%) |
| Chronic pulmonary disease(not asthma) | 27 (15.6%) | 13 (14.3%) | 14 (17.1%) | 29 (12.8%) | 14 (11.4%) | 15 (14.6%) |
| Active cancer(including hematological malignancy) | 23 (13.3%) | 9 (9.9%) | 14 (17.1%) | 30 (13.3%) | 12 (9.8%) | 18 (17.5%) |
| Asthma | 16 (9.2%) | 9 (9.9%) | 7 (8.5%) | 24 (10.6%) | 13 (10.6%) | 11 (10.7%) |
| Chronic kidney disease (stage 1 to 4) | 13 (7.5%) | 5 (5.5%) | 8 (9.8%) | 17 (7.5%) | 7 (5.7%) | 10 (9.7%) |
| Chronic neurological disorder (including dementia) | 11 (6.4%) | 8 (8.8%) | 3 (3.7%) | 12 (5.3%) | 8 (6.5%) | 4 (3.9%) |
| Liver disease | 7 (4.0%) | 5 (5.5%) | 2 (2.4%) | 8 (3.5%) | 6 (4.9%) | 2 (1.9%) |
| Auto-inflammatory disease | 7 (4.0%) | 1 (1.1%) | 6 (7.3%) | 8 (3.5%) | 2 (1.6%) | 6 (5.8%) |
| Hematopoietic stem cell or solid organ transplantation | 3 (1.7%) | 0 (0.0%) | 3 (3.7%) | 4 (1.8%) | 0 (0.0%) | 4 (3.9%) |
| HIV infection | 1 (0.6%) | 1 (1.1%) | 0 (0.0%) | 2 (0.9%) | 2 (1.6%) | 0 (0.0%) |
| **Median days from symptoms onset and random assignment*** | 7.0 [5.0-9.0] | 7.0 [6.0-9.0] | 7.0 [5.0-9.0] | 7.0 [6.0-9.0] | 8.0 [6.0-9.0] | 7.0 [5.0-9.0] |
| **Ventilation mode** | | | | | | |
| Room air | 14 (8.1%) | 9 (9.9%) | 5 (6.1%) | 16 (7.1%) | 10 (8.1%) | 6 (5.8%) |
| Oxygen support (standard oxygen therapy, high flow oxygen device, NIV) | 159 (91.9%) | 82 (90.1%) | 77 (93.9%) | 210 (92.9%) | 113 (91.9%) | 97 (94.2%) |
| **NEWS2 Score*** | 7.0 [5.0-9.0] | 7.0 [5.0-8.0] | 7.0 [5.0-9.0] | 7.0 [5.0-9.0] | 7.0 [5.0-8.0] | 6.0 [5.0-9.0] |
| **Clinical status** | | | | | | |
| 3. Hospitalized, not requiring supplemental oxygen | 7 (4.0%) | 5 (5.5%) | 2 (2.4%) | 9 (4.0%) | 6 (4.9%) | 3 (2.9%) |
| 4. Hospitalized, requiring supplemental oxygen | 141 (81.5%) | 71 (78.0%) | 70 (85.4%) | 179 (79.2%) | 93 (75.6%) | 86 (83.5%) |
| 5. Hospitalized, on non-invasive ventilation or high flow oxygen devices | 25 (14.5%) | 15 (16.5%) | 10 (12.2%) | 38 (16.8%) | 24 (19.5%) | 14 (13.6%) |
| **Randomisation site** | | | | | | |
| Intensive care unit | 21 (12.1%) | 15 (16.5%) | 6 (7.3%) | 32 (14.2%) | 21 (17.1%) | 11 (10.7%) |
| Conventional unit | 152 (87.9%) | 76 (83.5%) | 76 (92.7%) | 194 (85.8%) | 102 (82.9%) | 92 (89.3%) |
| **Vaccination initiation (partly or fully)** | 82 (47.4%) | 43 (47.3%) | 39 (47.6%) | 102 (45.1%) | 56 (45.5%) | 46 (44.7%) |
| **Result of rapid antigen test** | | | | | | |
| Positive | 173 (100.0%) | 91 (100.0%) | 82 (100.0%) | 173 (76.5%) | 91 (74.0%) | 82 (79.6%) |
| Negative | | | | 53 (23.5%) | 32 (26.0%) | 21 (20.4%) |

| | Antigen positive (N = 173) | | | Global (N = 226) | | |
|---|---|---|---|---|---|---|
| | **Overall** (N = 173) | **Tixagevimab-cilgavimab** (N = 91) | **Placebo** (N = 82) | **Overall** (N = 226) | **Tixagevimab-cilgavimab** (N = 123) | **Placebo** (N = 103) |
| **Serology*** | | | | | | |
| Negative (Anti-N antibodies - and Anti-S RBD antibodies -) | 69 (40.4%) | 40 (44.4%) | 29 (35.8%) | 76 (34.4%) | 44 (36.4%) | 32 (32.0%) |
| Positive (Anti-N antibodies + or Anti-S RBD antibodies +) | 102 (59.6%) | 50 (55.6%) | 52 (64.2%) | 145 (65.6%) | 77 (63.6%) | 68 (68.0%) |
| **Variant sequencing at baseline*** | | | | | | |
| Clade 20A | 2 (1.4%) | 0 (0.0%) | 2 (3.1%) | 2 (1.3%) | 0 (0.0%) | 2 (2.7%) |
| Beta, V2 | 2 (1.4%) | 1 (1.4%) | 1 (1.5%) | 2 (1.3%) | 1 (1.2%) | 1 (1.4%) |
| Alpha, V1 | 5 (3.6%) | 3 (4.1%) | 2 (3.1%) | 6 (3.9%) | 3 (3.7%) | 3 (4.1%) |
| Gamma, V3 | 0 (0.0%) | 0 (0.0%) | 0 (0.0%) | 1 (0.6%) | 1 (1.2%) | 0 (0.0%) |
| Delta | 68 (49.3%) | 35 (47.9%) | 33 (50.8%) | 76 (49.0%) | 37 (45.7%) | 39 (52.7%) |
| Omicron 21K (BA.1) | 42 (30.4%) | 24 (32.9%) | 18 (27.7%) | 49 (31.6%) | 29 (35.8%) | 20 (27.0%) |
| Omicron 21L (BA.2) | 18 (13.0%) | 10 (13.7%) | 8 (12.3%) | 18 (11.6%) | 10 (12.3%) | 8 (10.8%) |
| Omicron 22B (BA.5) | 1 (0.7%) | 0 (0.0%) | 1 (1.5%) | 1 (0.6%) | 0 (0.0%) | 1 (1.4%) |
| **Variant Omicron on Day 1 (imputed)** | | | | | | |
| Pre-Omicron | 99 (57.2%) | 52 (57.1%) | 47 (57.3%) | 133 (58.8%) | 70 (56.9%) | 63 (61.2%) |
| Omicron BA1 | 44 (25.4%) | 25 (27.5%) | 19 (23.2%) | 53 (23.5%) | 32 (26.0%) | 21 (20.4%) |
| Omicron BA2/5 | 20 (11.6%) | 10 (11.0%) | 10 (12.2%) | 22 (9.7%) | 11 (8.9%) | 11 (10.7%) |
| Unknown | 10 (5.8%) | 4 (4.4%) | 6 (7.3%) | 18 (8.0%) | 10 (8.1%) | 8 (7.8%) |
| **SAPSII (IGSII) Score***, Median [IQR] | 31.0 [24.0-34.0] | 29.0 [23.0-33.0] | 33.0 [24.0-40.0] | 31.0 [23.5-34.5] | 31.0 [22.0-35.0] | 33.0 [24.0-34.0] |
| **SOFA Score***, Median [IQR] | 3.0 [2.0-3.0] | 3.0 [2.0-3.0] | 3.0 [2.0-4.0] | 3.0 [2.0-3.0] | 2.5 [2.0-3.0] | 3.0 [2.0-4.5] |
| **Median normalized viral load in nasopharyngeal swabs at baseline,** log10 copies per 10000 cells† | 4.4 [3.3-5.4] | 4.4 [3.4-5.3] | 4.4 [3.3-5.6] | 4.0 [2.5--5.2] | 3.9 [2.4-5.1] | 4.1 [2.7-5.3] |
| **Biological data at baseline*** | | | | | | |
| Minimal absolute lymphocyte count, $10^9$ cells per L | 0.8 [0.5-1.1] | 0.8 [0.5-1.0] | 0.8 [0.6-1.2] | 0.8 [0.6-1.2] | 0.8 [0.6-1.2] | 0.8 [0.5-1.3] |
| Maximal absolute neutrophil count, $10^9$ cells per L | 5.5 [3.6-7.5] | 5.5 [3.1-7.7] | 5.5 [4.0-7.1] | 5.6 [3.7-7.7] | 5.6 [3.6-7.9] | 5.6 [3.9-7.5] |
| Minimal platelet count, $10^9$ cells per L | 202.0 [157.0-271.0] | 204.0 [153.0-264.0] | 201.0 [163.0-275.0] | 215.0 [165.0-281.0] | 216.0 [175.0-281.0] | 207.0 [158.0-275.0] |
| Minimal hemoglobin count, g/dL | 13.4 [12.0-14.6] | 13.2 [12.0-14.5] | 13.6 [12.1-14.6] | 13.4 [12.1-14.5] | 13.2 [12.1-14.4] | 13.5 [12.1-14.6] |
| Maximal creatininemia, µmol/L | 74.0 [63.0-89.0] | 74.0 [61.0-88.0] | 77.0 [65.0-103.0] | 74.0 [62.0-89.0] | 71.0 [60.0-87.0] | 76.0 [64.0-97.0] |
| Minimal estimated glomerular filtration rate, mL/min | 85.0 [66.0-98.0] | 85.5 [71.5-103.0] | 85.0 [58.0-97.0] | 88.5 [70.0-101.0] | 90.0 [75.0-103.0] | 85.5 [59.0-99.0] |
| Maximal aspartate aminotransferase, IU/L | 42.0 [31.0-59.0] | 40.0 [28.0-58.0] | 46.5 [34.0-61.5] | 42.5 [31.0-59.0] | 41.0 [29.0-58.0] | 45.0 [34.0-61.0] |
| Maximal alanine aminotransferase, IU/L | 35.0 [22.0-49.0] | 35.0 [19.5-46.0] | 34.5 [24.0-55.5] | 36.5 [22.0-50.0] | 37.0 [21.0-48.0] | 35.0 [24.0-59.0] |
| Maximal lactate dehydrogenase, IU/L | 348.0 [267.0-435.0] | 315.0 [260.0-412.0] | 361.0 [284.0-462.0] | 353.5 [271.0-458.0] | 335.0 [262.5-448.5] | 375.0 [294.0-464.0] |
| Minimal prothrombin time, % | 90.0 [81.0-100.0] | 89.0 [82.0-100.0] | 91.0 [80.5-100.0] | 90.0 [81.0-100.0] | 89.0 [82.0-100.0] | 90.5 [80.5-100.0] |
| Maximal fibrinogen, g/L | 5.7 [4.9-6.6] | 5.7 [4.8-6.7] | 5.8 [5.0-6.5] | 5.8 [4.9-6.7] | 5.8 [4.9-6.8] | 5.8 [4.8-6.5] |
| Maximal C-reactive protein, mg/L | 62.0 [35.0-128.0] | 58.5 [35.0-114.0] | 66.0 [35.0-146.0] | 55.0 [28.0-128.0] | 60.0 [30.0-125.0] | 51.0 [26.5-137.5] |
| Maximal D-Dimeres, µg/L | 827.0 [493.5-1290.5] | 833.0 [487.0-1170.0] | 821.0 [536.0-1402.0] | 822.5 [471.0-1360.0] | 816.5 [439.0-1407.5] | 822.5 [536.0-1316.0] |
| Maximal ferritin, µg/L | 786.0 [384.0-1386.0] | 772.0 [383.0-1350.0] | 818.0 [414.0-1422.0] | 790.0 [404.0-1390.0] | 795.0 [384.0-1386.0] | 767.0 [409.0-1433.0] |
| Serum total immunoglobulins, g/L | 10.2 [8.0-13.0] | 9.8 [7.8-13.2] | 10.3 [8.2-12.9] | 10.2 [8.1-13.0] | 10.6 [8.0-13.2] | 10.0 [8.3-12.8] |
| **Presence of major artifacts precluding analysis of lung parenchyma*,** | 8 (5.4%) | 4 (5.6%) | 4 (5.3%) | 10 (5.3%) | 6 (6.3%) | 4 (4.3%) |
| **Presence of pulmonary embolism**, | 2 (2.2%) | 1 (2.8%) | 1 (1.9%) | 3 (2.7%) | 1 (2.2%) | 2 (3.1%) |
| **Extension of lung parenchyma lesion***, n (%) | | | | | | |
| 0% | 5 (4.0%) | 3 (5.0%) | 2 (3.0%) | 5 (3.0%) | 3 (3.6%) | 2 (2.4%) |
| 1-10% | 18 (14.3%) | 12 (20.0%) | 6 (9.1%) | 22 (13.2%) | 14 (16.9%) | 8 (9.5%) |
| 11-25% | 44 (34.9%) | 20 (33.3%) | 24 (36.4%) | 60 (35.9%) | 30 (36.1%) | 30 (35.7%) |
| 26-50% | 45 (35.7%) | 17 (28.3%) | 28 (42.4%) | 55 (32.9%) | 23 (27.7%) | 32 (38.1%) |
| 51-75% | 13 (10.3%) | 7 (11.7%) | 6 (9.1%) | 22 (13.2%) | 11 (13.3%) | 11 (13.1%) |
| 76-100% | 1 (0.8%) | 1 (1.7%) | 0 (0.0%) | 3 (1.8%) | 2 (2.4%) | 1 (1.2%) |
| **Presence of pleural effusion***, | 12 (8.6%) | 9 (13.4%) | 3 (4.2%) | 14 (7.7%) | 11 (11.8%) | 3 (3.3%) |
| **Presence of mediastinal lymph node***, | 26 (18.7%) | 11 (16.4%) | 15 (20.8%) | 35 (19.1%) | 17 (18.3%) | 18 (20.0%) |

Data are median [IQR] or n(%).

HIV: human immunodeficiency virus; BMI: body mass index; ECMO: extracorporeal membrane oxygenation.

* denotes variables with missing data.

The number of missing data is presented for the antigen positive / the global mITT for each variable, respectively. Data on ethnicity were missing in 0/1 participant; data on geographical origin were missing in 7/11 participants; data on smoking status (current) were missing in 3/3 participants; data on delay between first laboratory-confirmed SARS-CoV-2 infection and admission date at facility were missing in 1/1 participant; data on delay between first laboratory-confirmed SARS-CoV-2 infection and randomization were missing in 1/1 participant; data on serology test were missing on 2/5 participants; data on variant sequencing were missing in 35/71 participants; data on body weight were missing on 10/11 participants; data on NEWS2 Score were missing on 16/20 participants; data on SAPSII (IGSII) Score were missing on 147/190 participants; data on SOFA Score were missing in 151/194 participants; data on Haemoglogin count were missing in 6/6 participants; data on absolute lymphocyte count were missing in 18/26 participants; data on absolute neutrophil count were missing in 18/25 participants; data on Platelet count were missing in 7/8 participants; data on Creatinine max were missing in 2/2 participants; data on eGFR min were missing in 4/6 participants; data on AST / SGOT max were missing in 10/14 participants; data on ALT / SGPT max were missing in 9/12 participants; data on LDH max were missing in 44/60 participants; data on Prothrombin time were missing in 28/35 participants; data on Fibrinogen max were missing in 38/45 participants; data on C-reactive protein were missing in 14/17 participants; data on D-Dimers were missing in 33/44 participants; data on Ferritin were missing in 43/57 participants; data on Serum total immunoglobulins were missing in 80/107 participants; data on plasma pregnancy test were missing in 53/65 participants; data on urine pregnancy test were missing in 50/63 participants; data on presence of major artifacts precluding analysis of lung parenchyma were missing in 0/3 participants; data on extension of lung parenchyma lesions were missing in 21/25 participants; data on presence of pleural effusion were missing in 8/9 participants; data on presence of mediastinal lymph nodes were missing in 8/9 participants.

† Undetectable viral load values (i.e. values < 1 log10 copies/10 000 cells) were imputed to 0.7 log10 copies/10 000 cells.

**Supplementary table S2. Proportion of patients with a detectable viral load, normalized viral load, change from baseline of normalized viral loads in the nasopharyngeal swabs and the plasma, at each sampling time, in the antigenic positive and global modified intention-to-treat populations, overall and according to the treatment group.**

Analyses of detectable viral load were stratified on the vaccination status at randomization and reported odds ratios are adjusted on vaccination status.

| | **Antigen positive (N = 173)** | | | | **Global (N = 226)** | | | |
|---|---|---|---|---|---|---|---|---|
| | **Overall** (N = 173) | **Tixagevimab-cilgavimab** (N = 91) | **Placebo** (N = 82) | **Effect measure** (95%CI) | **Overall** (N = 226) | **Tixagevimab-cilgavimab** (N = 123) | **Placebo** (N = 103) | **Effect measure** (95%CI) |
| **Detectable viral load in NP swabs,** n/N (%) | | | | | | | | |
| - at baseline | 153/159 (96.2%) | 80/83 (96.4%) | 73/76 (96.1%) | | 187/205 (91.2%) | 101/111 (91.0%) | 86/94 (91.5%) | |
| - at day 3 | 130/148 (87.8%) | 64/74 (86.5%) | 66/74 (89.2%) | OR=0.77 (0.29 to 2.09) [p=0.61] | 153/191 (80.1%) | 78/100 (78.0%) | 75/91 (82.4%) | OR=0.72 (0.35 to 1.47) [p=0.36] |
| - at day 8 | 62/92 (67.4%) | 29/47 (61.7%) | 33/45 (73.3%) | OR=0.55 (0.22 to 1.34) [p=0.19] | 66/114 (57.9%) | 31/62 (50.0%) | 35/52 (67.3%) | OR=0.47 (0.22 to 1.01) [p=0.05] |
| - at day 15 | 13/34 (38.2%) | 5/17 (29.4%) | 8/17 (47.1%) | OR=0.48 (0.12 to 1.93) [p=0.30] | 13/49 (26.5%) | 5/25 (20.0%) | 8/24 (33.3%) | OR=0.52 (0.14 to 1.85) [p=0.31] |
| - at day 29 | 18/96 (18.8%) | 10/45 (22.2%) | 8/51 (15.7%) | OR=1.52 (0.54 to 4.28) [p=0.43] | 20/128 (15.6%) | 11/66 (16.7%) | 9/62 (14.5%) | OR=1.15 (0.44 to 3.00) [p=0.77] |
| **Normalized SARS-CoV-2 Viral load in NP swabs,** median (IQR) (in $\log_{10}$ copies per 10000 cells)* | | | | | | | | |
| - at baseline | 4.4 [3.4-5.4] | 4.4 [3.5-5.3] | 4.5 [3.4-5.6] | | 4.0 [2.5--5.2] | 3.9 [2.4-5.1] | 4.1 [2.7-5.3] | |
| - at day 3 | 3.5 [2.0-5.0] | 3.6 [1.9-5.2] | 3.3 [2.1-4.7] | | 2.8 [1.2-4.3] | 2.7 [1.0-4.4] | 2.9 [1.8-4.3] | |
| - at day 8 | 2.2 [0.7-3.7] | 1.8 [0.7-3.2] | 2.6 [0.7-3.8] | | 1.5 [0.7-3.2] | 0.9 [0.7-2.7] | 2.2 [0.7-3.7] | |
| - at day 15 | 0.7 [0.7-1.5] | 0.7 [0.7-1.0] | 0.7 [0.7-1.5] | | 0.7 [0.7-0.7] | 0.7 [0.7-0.7] | 0.7 [0.7-1.3] | |
| - at day 29 | 0.7 [0.7-0.7] | 0.7 [0.7-0.7] | 0.7 [0.7-0.7] | | 0.7 [0.7-0.7] | 0.7 [0.7-0.7] | 0.7 [0.7-0.7] | |
| **Change from baseline in viral load in NP swabs,** median (IQR) (in $\log_{10}$ copies per 10000 cells)* | | | | | | | | |
| - at day 3 | 0.7 [0.0-1.7] | 0.8 [-0.1-1.6] | 0.7 [0.1-1.9] | | 0.7 [0.0-1.7] | 0.7 [-0.2-1.6] | 0.6 [0.0-1.8] | |
| - at day 8 | 2.2 [1.3-3.4] | 2.3 [1.6-3.4] | 1.9 [0.9-3.3] | | 2.1 [1.1-3.3] | 2.2 [1.3-3.1] | 2.1 [0.9-3.3] | |
| - at day 15 | 3.2 [1.9-4.2] | 3.1 [2.1-4.3] | 3.4 [1.6-4.1] | | 3.0 [1.2-3.8] | 2.4 [0.6-3.3] | 3.2 [1.6-3.9] | |
| - at day 29 | 3.2 [2.0-4.6] | 3.3 [2.1-4.6] | 3.2 [1.5-4.4] | | 3.0 [1.4-4.2] | 2.8 [1.6-4.0] | 3.0 [1.2-4.4] | |
| **Undetectable viral load before day 29**, n/N (%) | 93/153 (60.8%) | 46/80 (57.5%) | 47/73 (64.4%) | | 122/187 (65.2%) | 64/101 (63.4%) | 58/86 (67.4%) | |
| **Time to first undetectable quantitative viral load in NP swabs through day 29**, median (IQR) | 17.0 [7.0-28.0] | 14.0 [7.0-29.0] | 26.0 [7.0-28.0] | HR=0.98 (0.67 to 1.42) [p=0.90] | 14.0 [7.0-28.0] | 8.0 [6.5-28.5] | 25.0 [7.0-28.0] | HR=1.04 (0.75 to 1.44) [p=0.81] |
| **Detectable viral load in the plasma**, n/N (%) | | | | | | | | |
| - at baseline | 35/137 (25.5%) | 18/72 (25.0%) | 17/65 (26.2%) | | 42/178 (23.6%) | 22/97 (22.7%) | 20/81 (24.7%) | |
| - at day 3 | 29/129 (22.5%) | 14/67 (20.9%) | 15/62 (24.2%) | | 32/170 (18.8%) | 15/93 (16.1%) | 17/77 (22.1%) | |
| - at day 8 | 6/83 (7.2%) | 2/41 (4.9%) | 4/42 (9.5%) | | 6/103 (5.8%) | 2/53 (3.8%) | 4/50 (8.0%) | |

CI: confidence interval; OR: odds ratio; HR: hazard ratio; LSMD: least squares mean difference ; NP: nasopharyngeal

* Undetectable viral load values (i.e. values < 1 log10 copies/10 000 cells) were imputed to 0.7 log10 copies/10 000 cells.

**Supplementary table S3. Estimated intercepts and slopes of viral load and difference of slopes between randomization arms, in the antigenic positive and global modified intention-to-treat populations, overall, and according to the treatment group and vaccination initiation at randomization.**

Analyses were stratified on vaccination status at randomization and reported effect measure is adjusted on vaccination status at randomization. Intercepts and slopes are reported with their standard errors; difference of slopes is reported with its 95% confidence interval.

| | **Antigen positive (N = 173)** | | | | | **Global (N = 226)** | | | | |
|---|---|---|---|---|---|---|---|---|---|---|
| | **Tixagevimab-cilgavimab** | | **placebo** | | **Effect measure (95%CI)** | **Tixagevimab-cilgavimab** | | **placebo** | | **Effect measure (95%CI)** |
| | Vaccination No | Vaccination Yes | Vaccination No | Vaccination Yes | | Vaccination No | Vaccination Yes | Vaccination No | Vaccination Yes | |
| Intercept, $log_{10}$ cp/10,000 cells (SE) | 3.72 (0.20) | 4.22 (0.20) | 3.73 (0.21) | 4.24 (0.20) | - | 3.41 (0.18) | 3.78 (0.19) | 3.61 (0.19) | 3.97 (0.19) | - |
| Slope, $log_{10}$ cp/10,000 cells/day (SE) | -0.11 (0.01) | -0.12 (0.01) | -0.11 (0.01) | -0.12 (0.01) | 0.002 (-0.03 to 0.03) [p=0.89] | -0.09 (0.01) | -0.10 (0.01) | -0.11 (0.01) | -0.12 (0.01) | 0.011 (-0.02 to 0.04) [p=0.42] |

**Supplementary table S4. Proportion of patients with an undetectable neutralizing antibodies titers, $log_{10}$ neutralizing antibodies titers ($IC_{50}$), and change from baseline of $log_{10}$ neutralizing antibodies titers at each sampling time, on patients from the global modified intention-to-treat population with known variant of infection at baseline, overall and according to the treatment group.**

For all analyses, we consider the $IC_{50}$ titers of neutralizing antibodies targeted at the variant corresponding to the variant of infection at baseline of the patient. Reported OR and LSMD were adjusted by vaccination status and by variant of infection at baseline.

| | **Global (N = 208)** | | | |
|---|---|---|---|---|
| | **Overall** (N = 208) | **Tixagevimab-cilgavimab** (N = 113) | **placebo** (N = 95) | **Effect measure** (95%CI) |
| **Undetectable neutralizing antibodies targeted at the variant of infection**, n/N (%) | | | | |
| Baseline | 105/198 (53.0%) | 56/108 (51.9%) | 49/90 (54.4%) | |
| Day 3 | 26/179 (14.5%) | 1/97 (1.0%) | 25/82 (30.5%) | OR=0.03 (0.00 to 0.27) [$p < 10^{-9}$] |
| Day 8 | 7/112 (6.3%) | 0/60 (0.0%) | 7/52 (13.5%) | NA |
| **$Log_{10}$ neutralizing antibodies titers against the variant of infection,** median (IQR) | | | | |
| Baseline | 1.2 [1.2-2.4] | 1.2 [1.2-2.3] | 1.2 [1.2-2.5] | |
| Day 3 | 3.2 [2.1-3.9] | 3.8 [3.3-4.1] | 2.1 [1.2-3.0] | LSMD=1.44 (1.20 to 1.68) [$p < 10^{-23}$] |
| Day 8 | 3.3 [2.6-3.8] | 3.5 [3.3-4.0] | 2.7 [2.0-3.4] | LSMD=0.91 (0.64 to 1.18) [$p < 10^{-8}$] |
| **Change from baseline of $log_{10}$ neutralizing antibodies titers against the variant of infection**, median (IQR) | | | | |
| to Day 3 | 0.8 [0.3-2.1] | 2.0 [0.9-2.7] | 0.3 [0.0-0.6] | LSMD=1.42 (1.18 to 1.65) [$p < 10^{-23}$] |
| to Day 8 | 1.3 [0.7-2.2] | 1.9 [1.1-2.4] | 0.8 [0.2-1.4] | LSMD=0.87 (0.57 to 1.17) [$p < 10^{-6}$] |

CI: confidence interval; OR: odds ratio; LSMD: least squares mean difference.

**Supplementary figure S1. Forrest plot of subgroup analyses of the primary outcome in the antigen positive (A) and global (B) modified intention-to-treat populations.**

The widths of the confidence intervals have not been adjusted for multiplicity and therefore cannot be used to infer treatment effects.

## (A)

**Antigen positive (N = 173)**

| Subgroups | Number of patients | OR (95%CI) | p-value | p-value interaction |
|---|---|---|---|---|
| **Vaccination status** | | | | |
| Yes | 82 | 0.91 (0.41 to 2.02) | 0.82 | 0.99 |
| No | 91 | 0.94 (0.44 to 1.99) | 0.87 | |
| **Clinical status at inclusion** | | | | |
| 3 or 4 | 148 | 0.99 (0.55 to 1.80) | 0.98 | 0.86 |
| 5 | 25 | 0.79 (0.19 to 3.28) | 0.75 | |
| **Duration of symptoms prior to randomization** | | | | |
| < 5 days | 28 | 0.76 (0.20 to 2.88) | 0.68 | 0.70 |
| >= 5 days | 145 | 0.98 (0.54 to 1.78) | 0.94 | |
| **Age** | | | | |
| < 65 years | 83 | 0.77 (0.35 to 1.73) | 0.53 | 0.61 |
| >= 65 years | 90 | 1.00 (0.47 to 2.14) | 0.99 | |
| **Serology** | | | | |
| Negative | 69 | 0.99 (0.41 to 2.36) | 0.98 | 0.84 |
| Positive | 102 | 0.89 (0.43 to 1.82) | 0.74 | |
| **Variant of infection** | | | | |
| Pre-omicron | 99 | 1.49 (0.73 to 3.03) | 0.28 | 0.21 |
| Omicron BA1 | 44 | 0.43 (0.14 to 1.32) | 0.14 | |
| Omicron BA2/5 | 20 | 0.30 (0.04 to 1.95) | 0.21 | |
| **Sex** | | | | |
| Female | 56 | 0.47 (0.17 to 1.28) | 0.14 | 0.08 |
| Male | 117 | 1.31 (0.67 to 2.57) | 0.42 | |
| **All positive mitt** | **173** | **0.93 (0.54 to 1.61)** | **0.81** | |

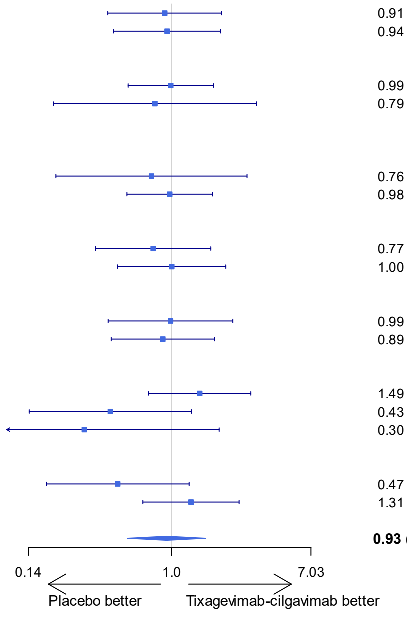

## (B)

**Global (N = 226)**

| Subgroups | Number of patients | OR (95%CI) | p-value | p-value interaction |
|---|---|---|---|---|
| **Antigen test at inclusion** | | | | |
| Positive | 173 | 0.93 (0.54 to 1.61) | 0.81 | 0.33 |
| Negative | 53 | 0.50 (0.17 to 1.44) | 0.20 | |
| **Vaccination status** | | | | |
| Yes | 102 | 0.95 (0.46 to 1.94) | 0.88 | 0.60 |
| No | 124 | 0.76 (0.40 to 1.47) | 0.42 | |
| **Clinical status at inclusion** | | | | |
| 3 or 4 | 188 | 0.85 (0.50 to 1.44) | 0.54 | 0.56 |
| 5 | 38 | 1.22 (0.38 to 3.95) | 0.74 | |
| **Duration of symptoms prior to randomization** | | | | |
| < 5 days | 35 | 0.49 (0.15 to 1.64) | 0.25 | 0.23 |
| >= 5 days | 191 | 0.96 (0.56 to 1.62) | 0.87 | |
| **Age** | | | | |
| < 65 years | 109 | 0.75 (0.36 to 1.53) | 0.42 | 0.90 |
| >= 65 years | 117 | 0.79 (0.41 to 1.53) | 0.48 | |
| **Serology** | | | | |
| Negative | 76 | 0.92 (0.40 to 2.12) | 0.84 | 0.47 |
| Positive | 145 | 0.71 (0.39 to 1.31) | 0.27 | |
| **Variant of infection** | | | | |
| Pre-omicron | 133 | 1.28 (0.69 to 2.38) | 0.44 | 0.30 |
| Omicron BA1 | 53 | 0.55 (0.20 to 1.54) | 0.26 | |
| Omicron BA2/5 | 22 | 0.25 (0.04 to 1.58) | 0.14 | |
| **Sex** | | | | |
| Female | 71 | 0.45 (0.18 to 1.12) | 0.09 | 0.08 |
| Male | 155 | 1.12 (0.62 to 2.02) | 0.70 | |
| **All** | **226** | **0.85 (0.52 to 1.37)** | **0.50** | |

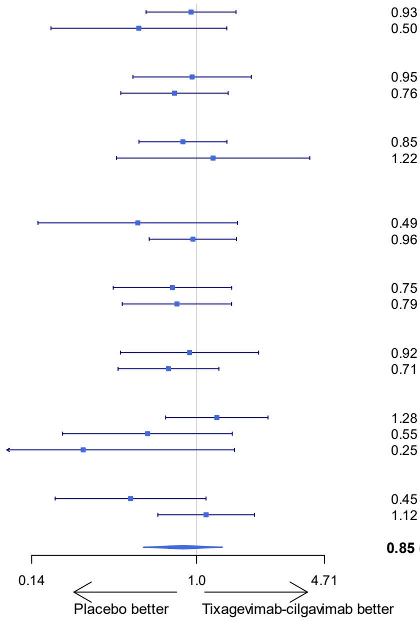

CI: confidence interval; OR: odds ratio

**Supplementary figure S2. Forrest plot of subgroup analyses of the key secondary outcome in the antigen positive (A) and global (B) modified intention-to-treat populations.**

The widths of the confidence intervals have not been adjusted for multiplicity and therefore cannot be used to infer treatment effects.

## (A)

**Antigen positive (N = 173)**

| Subgroups | Number of patients | HR (95%CI) | p-value | p-value interaction |
|---|---|---|---|---|
| **Vaccination status** | | | | |
| Yes | 82 | 0.99 (0.61 to 1.62) | 0.97 | 0.95 |
| No | 91 | 0.99 (0.64 to 1.51) | 0.95 | |
| **Clinical status at inclusion** | | | | |
| 3 or 4 | 148 | 1.04 (0.74 to 1.46) | 0.83 | 0.65 |
| 5 | 25 | 0.94 (0.37 to 2.35) | 0.89 | |
| **Duration of symptoms prior to randomization** | | | | |
| < 5 days | 28 | 1.27 (0.60 to 2.69) | 0.54 | 0.41 |
| >= 5 days | 145 | 0.93 (0.65 to 1.31) | 0.67 | |
| **Age** | | | | |
| < 65 years | 83 | 1.06 (0.67 to 1.67) | 0.80 | 0.81 |
| >= 65 years | 90 | 0.97 (0.59 to 1.58) | 0.90 | |
| **Serology** | | | | |
| Negative | 69 | 1.07 (0.63 to 1.82) | 0.80 | 0.74 |
| Positive | 102 | 0.95 (0.63 to 1.43) | 0.81 | |
| **Variant of infection** | | | | |
| Pre-omicron | 99 | 1.20 (0.79 to 1.83) | 0.38 | 0.56 |
| Omicron BA1 | 44 | 0.73 (0.36 to 1.46) | 0.37 | |
| Omicron BA2/5 | 20 | 0.56 (0.23 to 1.35) | 0.20 | |
| **Sex** | | | | |
| Female | 56 | 0.54 (0.31 to 0.96) | 0.04 | 0.02 |
| Male | 117 | 1.27 (0.86 to 1.88) | 0.22 | |
| **All positive mitt** | **173** | **0.98 (0.71 to 1.36)** | **0.92** | |

Axis: 0.40 – 1.0 – 3.00; ← Placebo better | Tixagevimab-cilgavimab better →

## (B)

**Global (N = 226)**

| Subgroups | Number of patients | HR (95%CI) | p-value | p-value interaction |
|---|---|---|---|---|
| **Antigen test at inclusion** | | | | |
| Positive | 173 | 0.98 (0.71 to 1.36) | 0.92 | 0.85 |
| Negative | 53 | 1.03 (0.60 to 1.75) | 0.93 | |
| **Vaccination status** | | | | |
| Yes | 102 | 1.13 (0.72 to 1.76) | 0.60 | 0.51 |
| No | 124 | 0.94 (0.66 to 1.36) | 0.76 | |
| **Clinical status at inclusion** | | | | |
| 3 or 4 | 188 | 1.06 (0.79 to 1.42) | 0.72 | 0.88 |
| 5 | 38 | 1.08 (0.53 to 2.21) | 0.82 | |
| **Duration of symptoms prior to randomization** | | | | |
| < 5 days | 35 | 1.19 (0.60 to 2.36) | 0.62 | 0.51 |
| >= 5 days | 191 | 0.97 (0.72 to 1.31) | 0.86 | |
| **Age** | | | | |
| < 65 years | 109 | 1.10 (0.75 to 1.63) | 0.63 | 0.60 |
| >= 65 years | 117 | 0.93 (0.61 to 1.41) | 0.73 | |
| **Serology** | | | | |
| Negative | 76 | 1.10 (0.66 to 1.82) | 0.72 | 0.59 |
| Positive | 145 | 0.95 (0.68 to 1.34) | 0.78 | |
| **Variant of infection** | | | | |
| Pre-omicron | 133 | 1.26 (0.88 to 1.79) | 0.20 | 0.60 |
| Omicron BA1 | 53 | 0.80 (0.42 to 1.52) | 0.50 | |
| Omicron BA2/5 | 22 | 0.53 (0.25 to 1.15) | 0.11 | |
| **Sex** | | | | |
| Female | 71 | 0.61 (0.38 to 0.98) | 0.04 | 0.04 |
| Male | 155 | 1.21 (0.87 to 1.68) | 0.27 | |
| **All** | **226** | **1.01 (0.77 to 1.34)** | **0.93** | |

Axis: 0.40 – 1.0 – 3.00; ← Placebo better | Tixagevimab-cilgavimab better →

CI: confidence interval; HR : hazard ratio

**Supplementary figure S3. Forrest plot of subgroup analyses of the difference in slopes of the viral load evolution through day 29 in the antigen positive (A) and global (B) modified intention-to-treat populations.** The widths of the confidence intervals have not been adjusted for multiplicity and therefore cannot be used to infer treatment effects.

**(A)**

**Antigen positive (N = 173)**

| Subgroups | Number of patients | Slope difference LSMD (95%CI) | p-value | p-value interaction |
|---|---|---|---|---|
| **Vaccination status** | | | | |
| Yes | 77 | -0.002 (-0.05 to 0.05) | 0.93 | 0.31 |
| No | 76 | -0.003 (-0.04 to 0.04) | 0.89 | |
| **Clinical status at inclusion** | | | | |
| 3 or 4 | 132 | 0.004 (-0.03 to 0.04) | 0.81 | 0.28 |
| 5 | 21 | -0.011 (-0.10 to 0.08) | 0.81 | |
| **Duration of symptoms prior to randomization** | | | | |
| < 5 days | 25 | -0.042 (-0.14 to 0.06) | 0.41 | 0.86 |
| >= 5 days | 128 | 0.008 (-0.02 to 0.04) | 0.61 | |
| **Age** | | | | |
| < 65 years | 68 | -0.005 (-0.05 to 0.04) | 0.83 | 0.50 |
| >= 65 years | 85 | 0.004 (-0.04 to 0.05) | 0.85 | |
| **Serology** | | | | |
| Negative | 61 | 0.028 (-0.02 to 0.07) | 0.20 | 0.75 |
| Positive | 92 | -0.020 (-0.06 to 0.02) | 0.29 | |
| **Variant at inclusion** | | | | |
| Pre-omicron | 84 | 0.004 (-0.04 to 0.04) | 0.85 | 0.17 |
| Omicron BA1 | 42 | 0.005 (-0.05 to 0.06) | 0.87 | |
| Omicron BA2/5 | 20 | -0.079 (-0.22 to 0.06) | 0.26 | |
| **Sex** | | | | |
| Female | 51 | -0.027 (-0.08 to 0.02) | 0.27 | 0.15 |
| Male | 102 | 0.020 (-0.02 to 0.06) | 0.32 | |
| **All antigenic positive** | **153** | **0.002 (-0.03 to 0.03)** | **0.89** | |

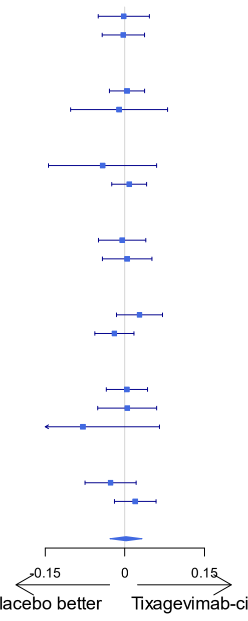

**(B)**

**Global (N = 226)**

| Subgroups | Number of patients | Slope difference LSMD (95%CI) | p-value | p-value interaction |
|---|---|---|---|---|
| **Antigen test at inclusion** | | | | |
| Positive | 153 | 0.002 (-0.03 to 0.03) | 0.89 | 0.50 |
| Negative | 34 | 0.029 (-0.03 to 0.09) | 0.34 | |
| **Vaccination status** | | | | |
| Yes | 93 | 0.008 (-0.04 to 0.05) | 0.71 | 0.42 |
| No | 94 | 0.006 (-0.03 to 0.04) | 0.73 | |
| **Clinical status at inclusion** | | | | |
| 3 or 4 | 158 | 0.014 (-0.02 to 0.04) | 0.38 | 0.47 |
| 5 | 29 | -0.013 (-0.09 to 0.06) | 0.73 | |
| **Duration of symptoms prior to randomization** | | | | |
| < 5 days | 30 | -0.029 (-0.09 to 0.03) | 0.35 | 0.51 |
| >= 5 days | 157 | 0.018 (-0.01 to 0.05) | 0.24 | |
| **Age** | | | | |
| < 65 years | 83 | 0.010 (-0.03 to 0.05) | 0.64 | 0.33 |
| >= 65 years | 104 | 0.008 (-0.03 to 0.05) | 0.72 | |
| **Serology** | | | | |
| Negative | 67 | 0.035 (-0.01 to 0.08) | 0.11 | 0.25 |
| Positive | 120 | -0.016 (-0.05 to 0.02) | 0.34 | |
| **Variant of infection** | | | | |
| Pre-omicron | 109 | 0.011 (-0.02 to 0.05) | 0.54 | 0.48 |
| Omicron BA1 | 50 | -0.00015 (-0.05 to 0.05) | 1.00 | |
| Omicron BA2/5 | 20 | -0.079 (-0.22 to 0.06) | 0.26 | |
| **Sex** | | | | |
| Female | 61 | -0.021 (-0.07 to 0.03) | 0.38 | 0.33 |
| Male | 126 | 0.025 (-0.01 to 0.06) | 0.15 | |
| **All** | **187** | **0.011 (-0.02 to 0.04)** | **0.42** | |

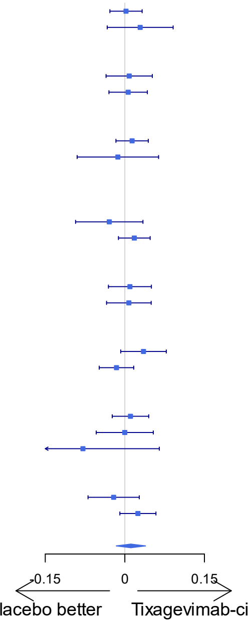

CI: confidence interval; LSMD: least squares mean difference.

**Supplementary figure S4: Cumulative incidence curve of time to sustained recovery through day 90, with death as competing event, in the antigen positive and global modified intention-to-treat populations, according to the treatment group.**

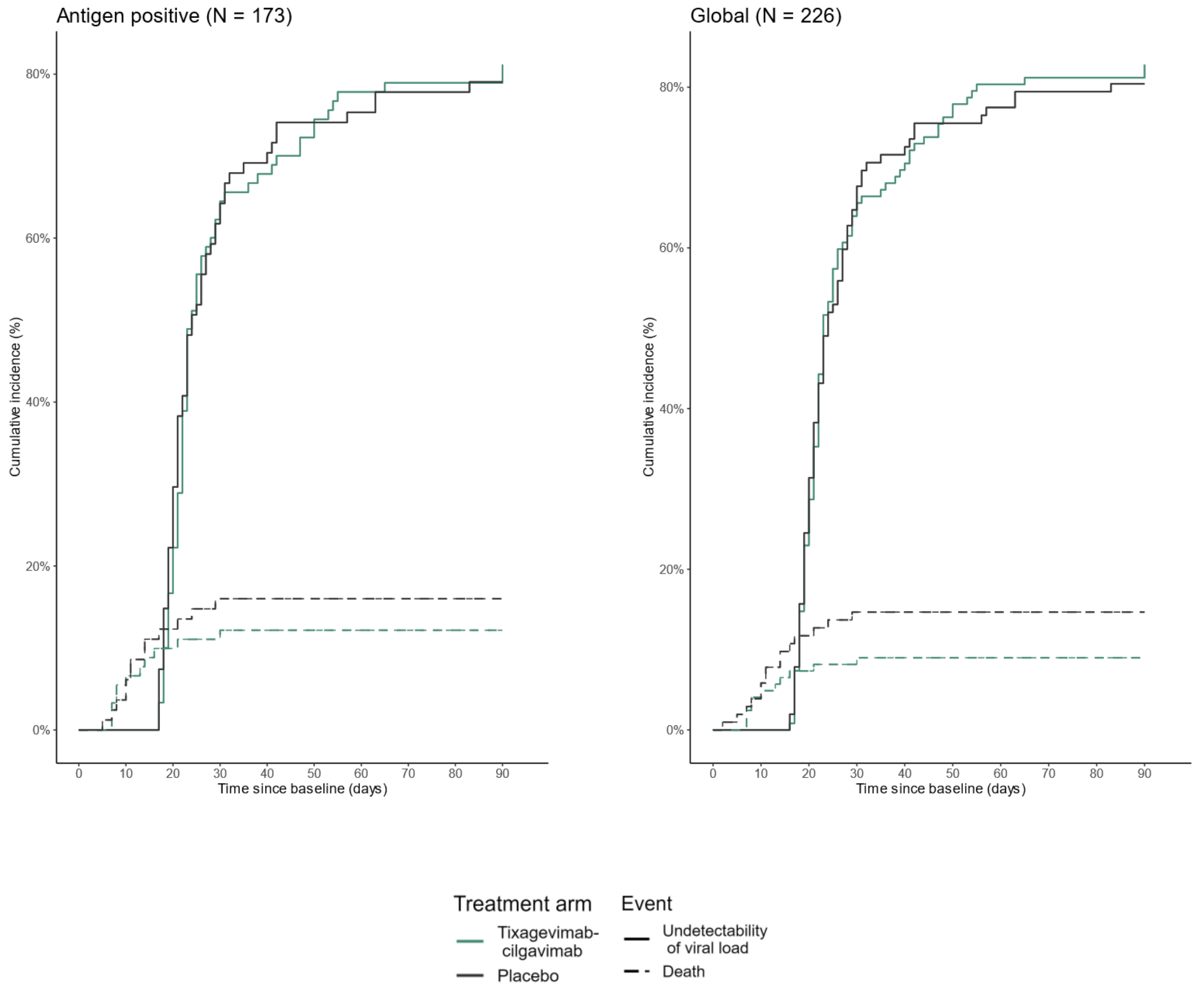

**Supplementary figure S5: Cumulative incidence curve of time to undetectability of viral load in nasopharyngeal swabs through day 29, with death as competing event, in the antigen positive and global modified intention-to-treat populations, according to the treatment group.**

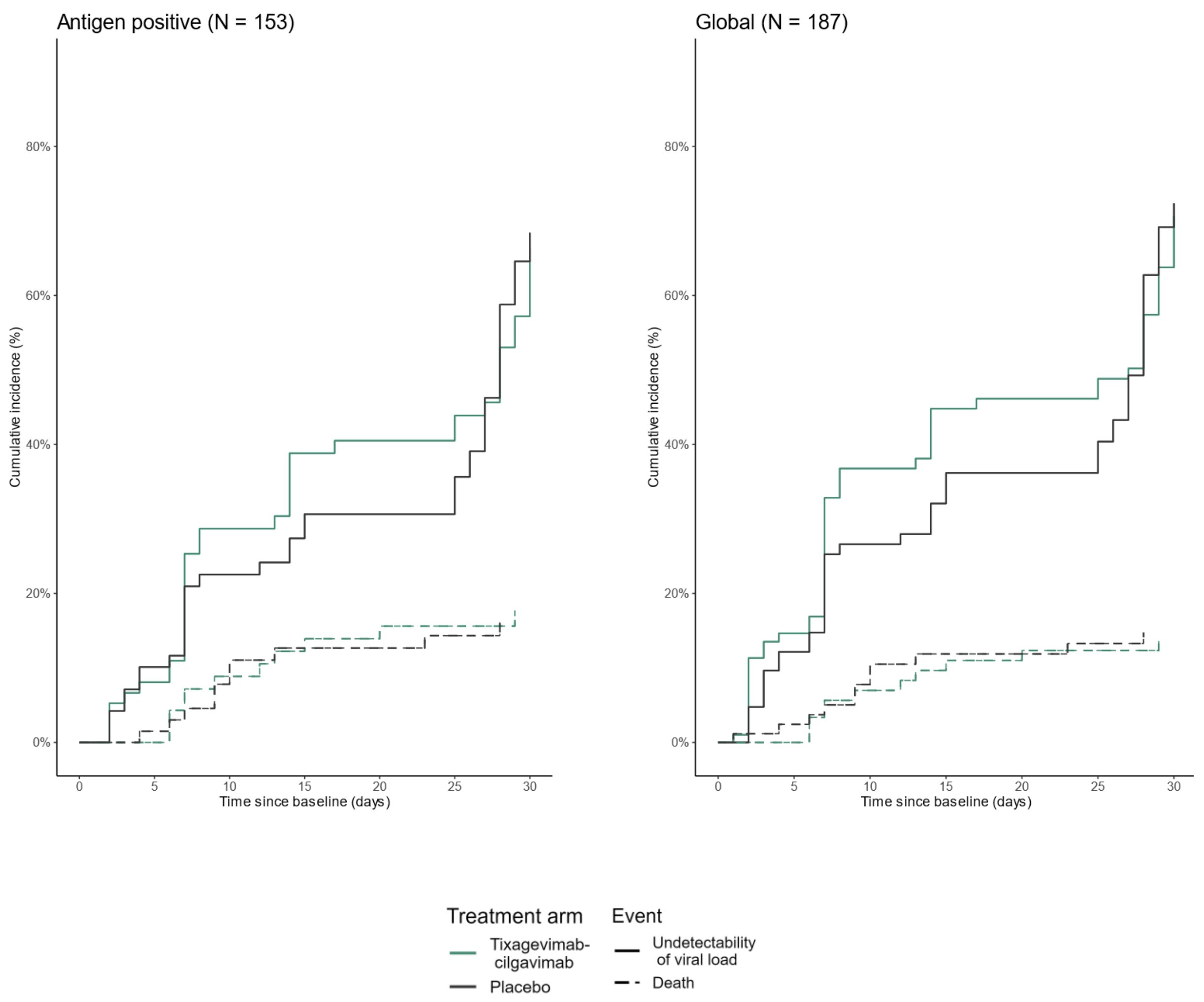

**Supplementary figure S6: Forrest plot of subgroup analyses on the evolution of log10 neutralizing antibodies titers (IC50) against variant of infection, at day 3 (A) and day 8 (B), in patients from the global modified intention-to-treat population, with known variant of infection at baseline.**

## (A)

**Log10 IC50 seroneutralisation against the variant of infection at Day3**

| Subgroups | Number of patients | LSMD (95%CI) | p-value | p-value Interaction |
|---|---|---|---|---|
| **Vaccination status** | | | | |
| Yes | 82 | 1.10 (0.81 to 1.39) | <1E-9 | 0.006 |
| No | 97 | 1.73 (1.38 to 2.08) | <1E-15 | |
| **Serology** | | | | |
| Negative | 65 | 1.97 (1.56 to 2.38) | <1E-12 | 0.004 |
| Positive | 114 | 1.20 (0.94 to 1.46) | <1E-14 | |
| **Variant of infection** | | | | |
| Pre-Omicron | 113 | 1.94 (1.67 to 2.20) | <1E-25 | <1E-7 |
| Omicron BA1 | 47 | 0.36 (-0.03 to 0.76) | 0.080 | |
| Omicron BA2/5 | 19 | 0.97 (0.27 to 1.67) | 0.015 | |
| **All** | **179** | **1.44 (1.20 to 1.68)** | **<1E-23** | |

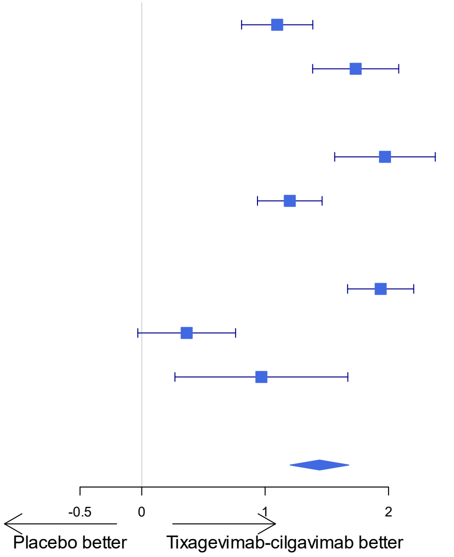

## (B)

**Log10 IC50 seroneutralisation against the variant of infection at Day8**

| Subgroups | Number of patients | LSMD (95%CI) | p-value | p-value interaction |
|---|---|---|---|---|
| **Vaccination status** | | | | |
| Yes | 47 | 0.69 (0.33 to 1.05) | <1E-3 | 0.20 |
| No | 65 | 1.04 (0.66 to 1.42) | <1E-5 | |
| **Serology** | | | | |
| Negative | 43 | 1.03 (0.53 to 1.54) | <1E-3 | 0.38 |
| Positive | 69 | 0.79 (0.49 to 1.09) | <1E-5 | |
| **Variant of infection** | | | | |
| Pre-Omicron | 74 | 1.17 (0.87 to 1.47) | <1E-9 | 0.01 |
| Omicron BA1 | 30 | 0.22 (-0.35 to 0.79) | 0.46 | |
| Omicron BA2/5 | 8 | 0.78 (0.46 to 1.10) | 0.005 | |
| **All** | **112** | **0.91 (0.64 to 1.18)** | **<1E-08** | |

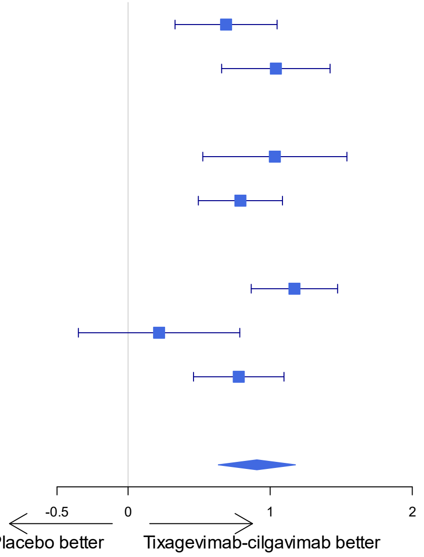

CI: confidence interval; LSMD: least squares mean difference.

**Supplementary figure S7. Clinical status on the WHO 7-point ordinal scale at baseline, day 15, day 29, day 90, in patients infected with a pre-omicron variant, from the global modified intention-to-treat population, according to the treatment group.**

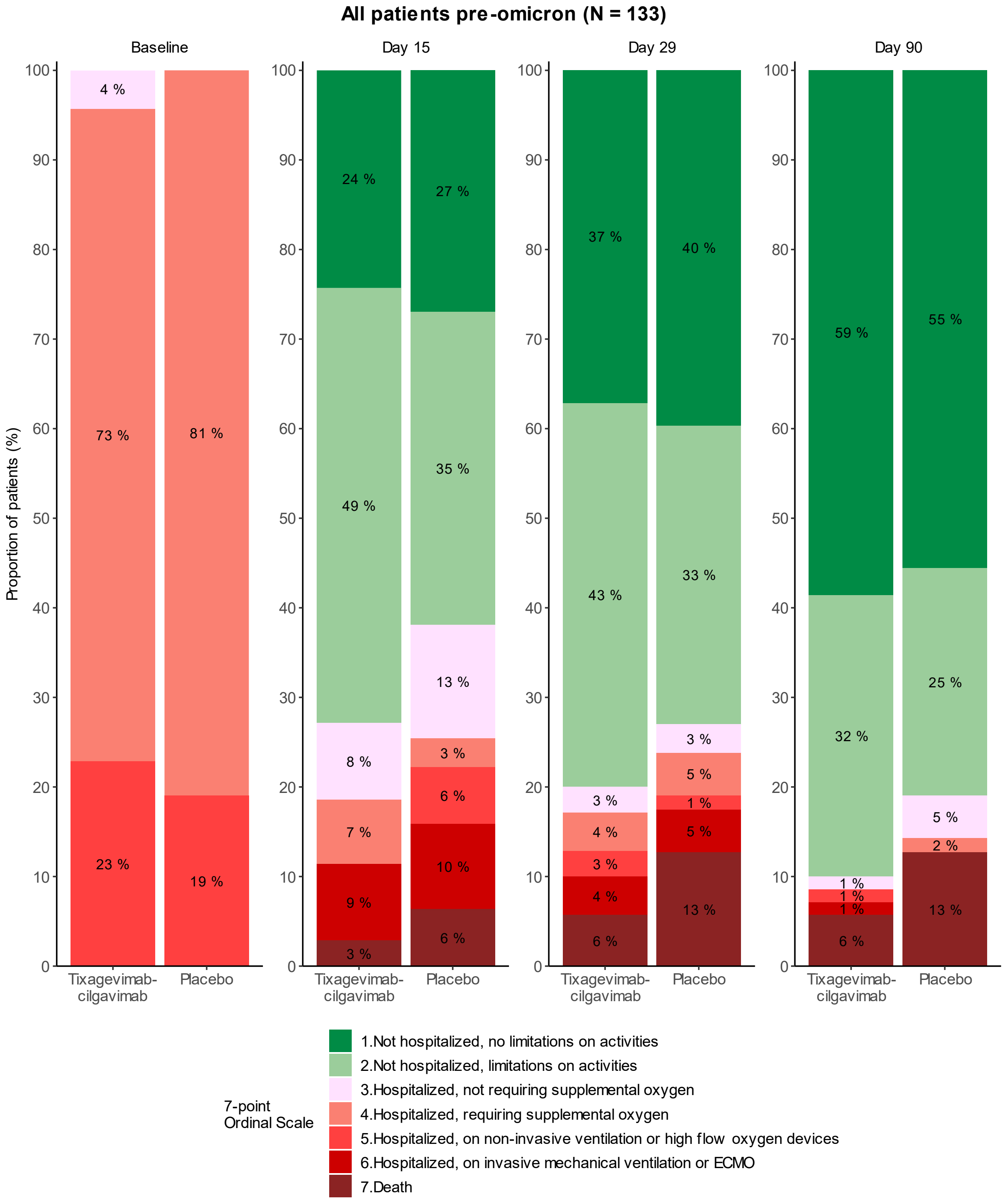

**Supplementary figure S8: Normalised SARS-CoV-2 viral loads in nasopharyngeal swabs in patients infected with a pre-omicron variant from the global modified intention-to-treat population, at each timepoint, and as change from baseline, according to the treatment group.**

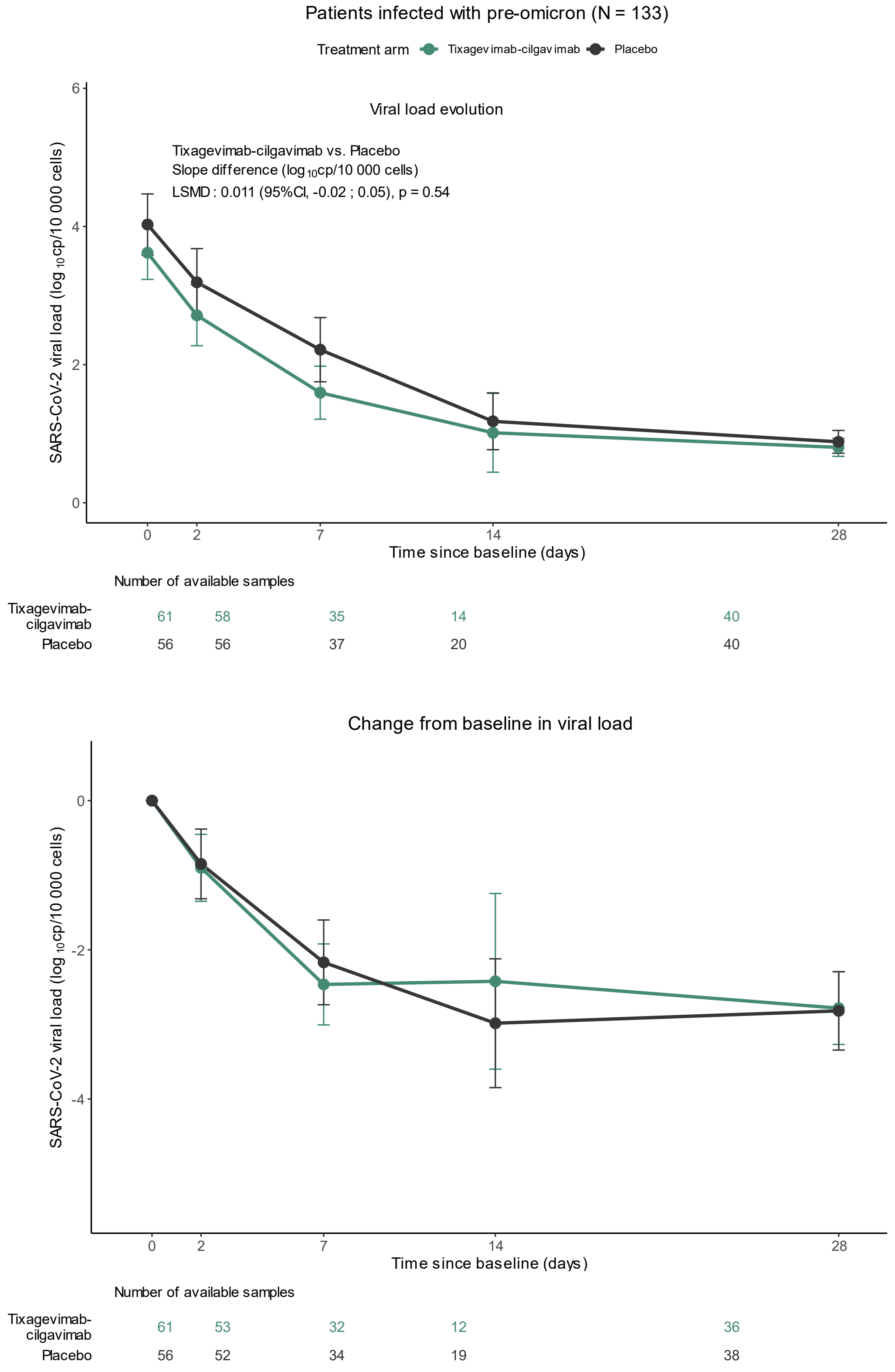

CI: confidence interval; LSMD: least squares mean difference.

**Supplementary Figure S9: Evolution of $log_{10}$ neutralizing antibody titers against variant of infection, from baseline to day 8 in patients infected with a pre-omicron variant, from the global modified intention-to-treat population.**

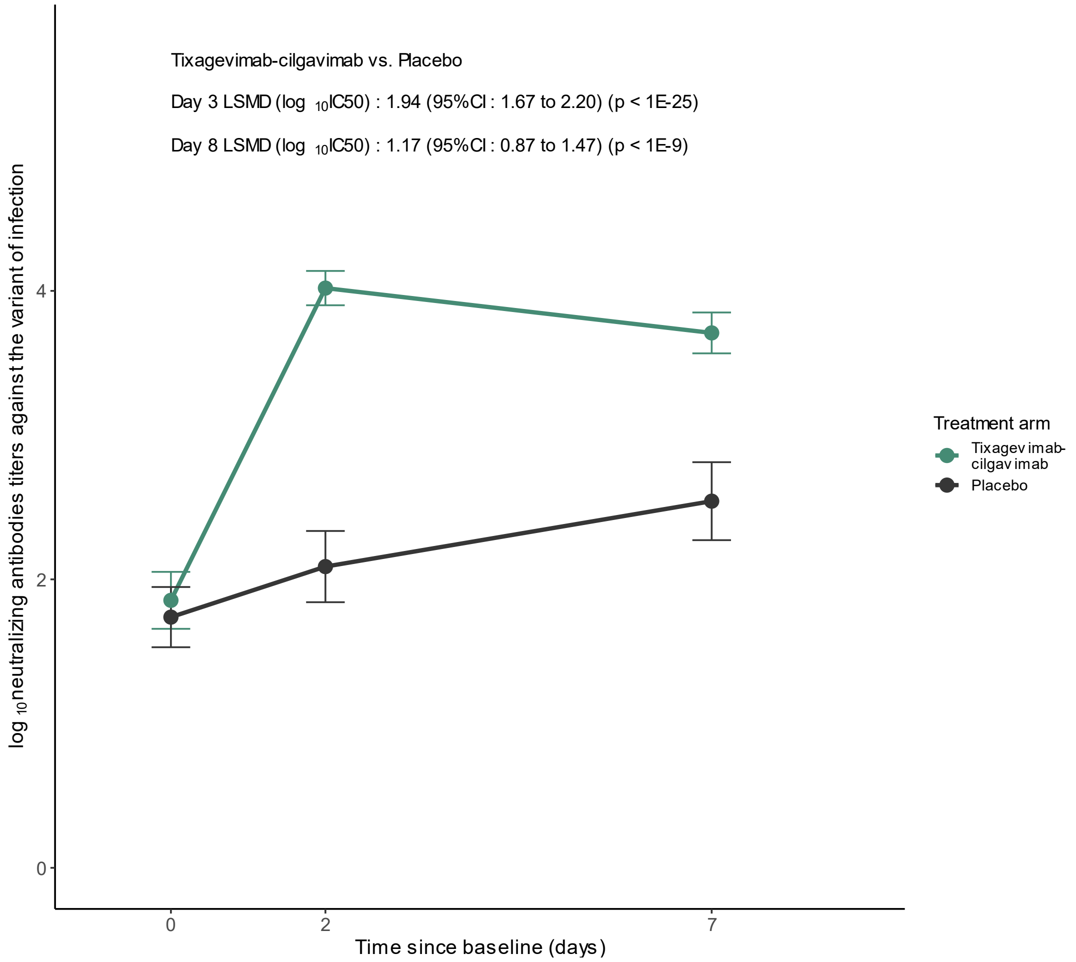

Number of available samples

| | | | |
|---|---|---|---|
| Tixagevimab-cilgavimab | 66 | 59 | 36 |
| Placebo | 59 | 54 | 38 |

CI: confidence interval; LSMD: least squares mean difference.

**Supplementary Figure S10: Cumulative incidence curve of time to sustained recovery through day 90 with death as competing event, in patients infected with a pre-omicron variant, from the global modified intention-to-treat population.**

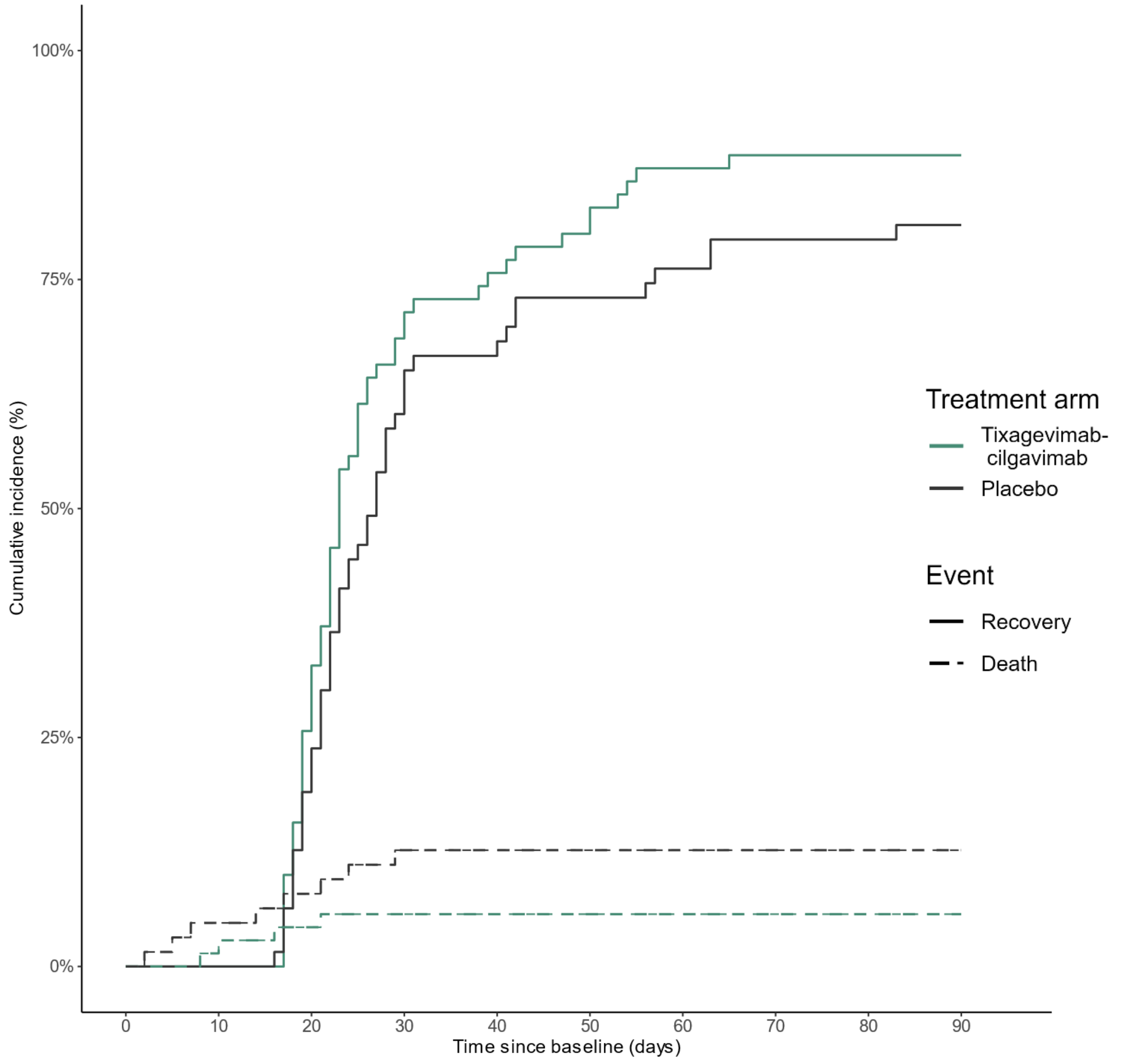

**Supplementary figure S11: Forest plot of the subgroup analyses of the number of patients with at least one serious adverse event of any type, in the safety population set, through day 90.**

Number of patients with at least one serious adverse event through day 90

| Subgroups | Number of patients | OR (95% CI) | p-value |
|---|---|---|---|
| **Antigen test at inclusion** | | | |
| Positive | 173 | 0.77 (0.39 to 1.52) | 0.45 |
| Negative | 53 | 0.28 (0.07 to 1.12) | 0.07 |
| **Vaccination status** | | | |
| Yes | 102 | 0.84 (0.37 to 1.89) | 0.67 |
| No | 124 | 0.44 (0.18 to 1.10) | 0.08 |
| **Clinical status at inclusion** | | | |
| 3 or 4 | 188 | 0.66 (0.34 to 1.29) | 0.23 |
| 5 | 38 | 0.47 (0.11 to 2.01) | 0.30 |
| **Duration of symptoms prior to randomization** | | | |
| < 5 days | 35 | 0.75 (0.18 to 3.15) | 0.70 |
| >= 5 days | 191 | 0.65 (0.33 to 1.28) | 0.21 |
| **Age** | | | |
| < 65 years | 109 | 0.44 (0.16 to 1.23) | 0.11 |
| >= 65 years | 117 | 0.93 (0.42 to 2.04) | 0.86 |
| **Serology** | | | |
| Negative | 76 | 0.55 (0.21 to 1.45) | 0.20 |
| Positive | 145 | 0.69 (0.30 to 1.59) | 0.38 |
| **Variant of infection** | | | |
| Pre-omicron | 133 | 0.31 (0.13 to 0.73) | 0.01 |
| Omicron BA1 | 53 | 1.38 (0.42 to 4.50) | 0.61 |
| Omicron BA2/5 | 22 | 2.20 (0.35 to 13.91) | 0.38 |
| **Sex** | | | |
| Female | 71 | 0.98 (0.31 to 3.08) | 0.97 |
| Male | 155 | 0.57 (0.27 to 1.21) | 0.14 |
| **All patients** | **226** | **0.63 (0.35 to 1.15)** | **0.13** |

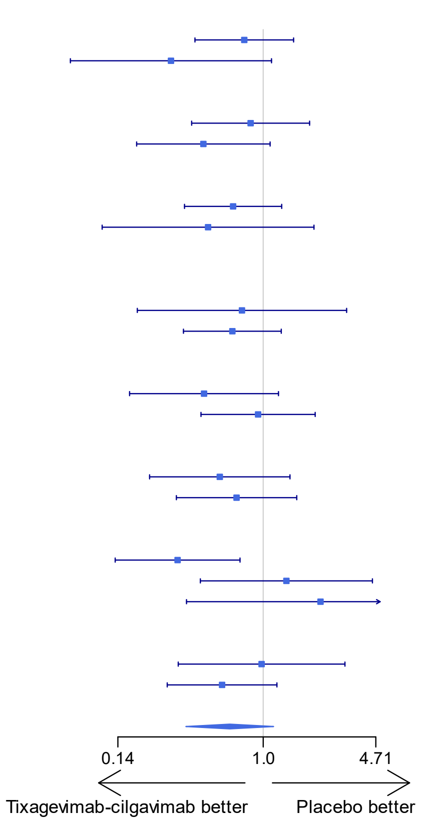

CI: confidence interval; OR: odds ratio

#### DisCoVeRy Study Group

**Sponsor:** Sandrine Couffin-Cadiergues, Hélène Espérou (Pôle de Recherche Clinique, Inserm, Paris, France)

**Investigators** (countries in alphabetical order):

***Austria:*** Alexander Egle, Alexander Egle, Richard Greil**,** Michael Grundbichler, Volker Hauck, Lukas Ratzinger, **(**Paracelsus Medical University Salzburg, SCRI-CCCI and AGMT)

***Belgium:*** Felix De Bièvre, Maya Hites, Frederique Jacobs, Eva Larranaga, Englert Pierre, Gil Verschelden (Hôpital Erasme, Cliniques Universitaires de Bruxelles) ; Philippe Clevenbergh, Sophie Leemans, Evelyne Maillart (Hôpital Brugmann, Bruxelles) ; Leila Belkhir, Julien De Greef, Jean Cyr Yombi (Cliniques Universitaires de Saint Luc, Bruxelles) ; Ides Colin, Clotilde Visée (Pôle Hospitalier Jolimont, Mons and Warquignies) ; Mathias Leys, Pieter Samaey (Centre hospitalier AZ Groeninge, Kortrijk) ; Janneke Cox, Peter Messiaen, Jeroen Van der Hilst, Karlijn Van Halem (Jessa Hospital, Hasselt).

***Czech Republic:*** Zula Badaev, Vyacheslav Grebenyuk, Milan Trojanek (Bulovka University Hospital, Praha) ; Veronika Bednarova, Anna Chaloupka, Katarina Dolezalova, Andrej Nagy, Michal Rezek, Filip Soucek, Alzbeta Trckova (St. Anne's University Hospital, Brno).

***France***: Adeline Bauvois, Cyril Charron, Laetitia Coutte, Guillaume Geri, Mathieu Godement, Segolene Greffe, Romain Jouffroy, Edouard Jullien, Jean-Emmanuel Kahn, Marion Pepin, Matthieu Petit, Elisabeth Rouveix-Nordon (Hôpital Ambroise Paré, Assistance Publique – Hôpitaux de Paris) ;
Claire Andrejak, Damien Basille, Cedric Joseph, Jean Philippe Lanoix, Sylvie Lion, Jean-Luc Schmit, Sandrine Soriot-Thomas (Centre Hospitalier Universitaire de Amiens) ; Carine Belzunce, David Bougon, Pierre-Alain Charretier, Renaud Chouquer, Didier Dorez, Etienne Escudier, Samuel Gay, Serge Hautefeuille, Cecile Janssen, Albrice Levrat, Gabriel Macheda, Mylene Maillet, Aldric Manuel, Audrey Merlet, Michel Muller, Sylvain Panh, Emilie Piet, Marion Rougon, Michel Sirodot, Violaine Tolsma, Caroline Varache, Vitrat Vitrat (Centre Hospitalier Annecy-Genevois) ; Kevin Bouiller, Catherine Chirouze, Dejan Ilic, Jean-Christophe Navellou, Noemie Tissot (Centre Hospitalier Universitaire de Besançon) ; Simon Bessi, Christelle Chantalat, Stephane Jaureguiberry (Hôpital Bicêtre, Assistance Publique – Hôpitaux de Paris) ; Antoine Bachelard, Chloe Bertin, Lio Collias, Odile Fleurot, Nathan Peiffer-Smadja, Valentina Isernia, Xavier Lescure, Bastien Mallo, Caroline Proux, Yazdan Yazdanpanah (Hôpital Bichat - Claude Bernard, Assistance Publique – Hôpitaux de Paris) ; Charles Cazanave, Philippe Boyer, Alexandre Boyer, Hoang-Nam Bui, Cedric Carrier, Benjamin Clouzeau, Arnaud Desclaux, Maylis Ducours, Cedric Gil-Jardine, Alexandre Gros, Didier Gruson, Julie Leitao, Maelig Lescure, Denis Malvy, Didier Neau, Maxime Poteau, Charline Sazio (Centre Hospitalier Universitaire de Bordeaux) ; Clara Bourrachot, Violaine Corbin, Kevin Grapin, Christine Jacomet, Henri Laurichesse, Olivier Lesens, Natacha Mrozek, Leo Sauvat, Aurelien Simon, Clement Theis,

Magali Vidal (Centre Hospitalier Universitaire de Clermont-Ferrand) ; Simon Gravier, Ciprian Ion, Damien Kayser, Martin Martinot, Mahsa Mohseni-Zadeh (Hospices Civils de Colmar) ; Brice Guerpillon, Lydie Khatchatourian, Nadia Saidani, Jean-Philippe Talarmin (Centre Hospitalier Cornouaille, Quimper) ; Marion Dollat,Nicolas Gambier, Marie-Aude Khuong, Yacine Tandjaouilambiotte (Hôpital Delafontaine, Saint-Denis) ; Marielle Buisson, Francois Xavier Catherine, Pascal Chavanet, Michel Duong, Clementine Esteve, Sophie Mahy, Lionel Piroth, Pascal Andreu, Francois Aptel, Pierre-Emmanuel Charles, Marie Labruyere, Sebastien Prin, Jean-Pierre Quenot, Jean-Baptiste Roudaut, Nicolas Aswad, Guillaume Beltramo, Amaury Berrier, Philippe Bonniaud, Charlotte Dignoire, Nicolas Favrolt, Marjolaine Georges, Claudio Rabec, Maximilien Spanjaard, (Centre Hospitalier Universitaire de Dijon) ; Constance Guillaud, Georgeta Lascu Dubos, Jean-Daniel Lelievre, Matthieu Mahevas, Sebastien Gallien, Hanane Mehawej, Giovanna Melica, Marc Michel, Florence Runyo, Noemie Saada, William Vindrios (Hôpital Henri-Mondor, Assistance Publique – Hôpitaux de Paris) ; Veronique Delcey, Sandra Devatine, Aude Kopp, Victoria Manda, Anne-Lise Munier, Pierre Sellier (Hôpital Lariboisière, Assistance Publique – Hôpitaux de Paris) ; Arnaud Salmon-Rousseau, Emmanuel Faure (Centre Hospitalier Régional Universitaire de Lille) ; Florence Ader, Dulce Alfaiate, Bernard Allaouchiche, Agathe Becker, Nicolas Benech, Arthur Bert, Elodie Berton, Laurent Bitker, Julien Bohe, Evelyne Braun, Olivier Cannesson, Paul Chabert, Pierre Chauvelot, Christian Chidiac, René Chumbi Flores, Anne Conrad, Gilles Devouassoux, Tristan Ferry, Frederic Gormand, Agathe Granger, Pierre Grumet, Lucille Jay, Emilie Joffredo, Thomas Jullien, Lize Kiakouama, Melanie Levrad, Ibrantina Lucena e Silva, Mehdi Mezidi, Patrick Miailhes, Oriane Pelton, Thomas Perpoint, Thierry Petitjean, Francois Philit, Cecile Pouderoux, Jean-Christophe Richard, Gaelle Richard-Colmant, Sandrine Roux, Pascal Seve, Fabrice Thiolliere, Claire Triffault Fillit, Florent Valour, Olivia Vassal, Florent Wallet, Hodanne Yonis (Hospices Civils de Lyon) ; Sylvie Abel, Bastien Bigeard, André Cabié, Ornella Cabras, Karine Guitteaud, Melanie Lehoux, Athena Marquise, Sandrine Pierre-François, Jean-Marie Turmel (Centre Hospitalier Universitaire de Martinique, Fort-de-France) ; Haffiane Abdelmagid, Alexis Hanisch, Flore Lacassin-Beller, Marion Mirabel, Isabel Roca Cerezo, Arnaud Sement (Centre Hospitalier Mont de Marsan) ; Charlotte Boulle, Jordan Lejeune, Vincent Le Moing, David Morquin, Nathalie Pansu, Lucas Perez (Centre Hospitalier Universitaire de Montpellier) ; Mathieu Baldacini, Samuel Degoul, Anne-Florence Dureau, Carmen Ionescu, Khaldoun Kuteifan, Guylaine Labro, Joy Mootien, Pierre Oudeville, Luis Pinto, Antoine Poidevin, Yannick Rabouel, Laure Stiel (Centre Hospitalier de Mulhouse Sud-Alsace) ; Guillaume Baronnet, Elisabeth Baux, Sybille Bevilacqua, Alice Corbel, François Goehringer, Eliette Jeanmaire, Antoine Kimmoun, Benjamin Lefevre (Centre Hospitalier Régional Universitaire de Nancy) ; David Boutoille, Emmanuel Canet, Colin Deschamvres, Benjamin Gaborit, Charlotte Garret, Pauline Lamouche Wilquin, Jean-Baptiste Lascarrou, Paul Le Turnier, Anne Sophie Lecompte, Raphael Lecomte, Jeremie Lemarie, Maelle Martin, Arnaud Felix Miailhe, Gregoire Ottavy, Gwennaelle Querne, François Raffi, Jean Reignier, Amelie Seguin, Stijn Sruvay, Olivier Zambon (Centre Hospitalier Universitaire de Nantes) ; Fanny Lanternier, Cléa

Melenotte, Claire Rouzaud, Alexandra Serris (Hôpital Necker, Assistance Publique – Hôpitaux de Paris) ; Julia Beatini, Matthieu Buscot, David Chirio, Johan Courjon, Eric cua, Aurelien Degoutte, Elisa Demonchy, Jacques Durant, Sylvie Leroy, Charles-Hugo Marquette, Celine Michelangeli, Veronique Mondain, Bernard Prouvost-Keller, Pascal Pugliese, Karine Risso (Centre Hospitalier Universitaire de Nice) ; Stephanie Bulyez, Aurele Buzancais, Aurelien Daurat, Romaric Larcher, Paul Loubet, Guillaume Louart, Laurent Muller, Nicolas Perin, Claire Roger (Centre Hospitalier Universitaire de Nîmes) ; Guillaume Beraud, Melanie Catroux, France Cazenave-Roblot, Valentin Giraud, Gwenael Le Moal, Jean-Philippe Martellosio (Centre Hospitalier Universitaire de Poitiers) ; Firouze Bani-Sadr, Messaline Bermejo, Joel Cousson, Antoine Goury, Maxime Hentzien, Gautier Julien, Bruno Mourvillier, Yohan Nguyen, Thibault Tromeur (Centre Hospitalier Universitaire de Reims) ; Jean-Sebastien Allain, Marion Baldeyrou, François Benezit, Marie Dewitte, Fabrice Laine, Bruno Laviolle, Solene Patrat-Delon, Matthieu Revest, Pierre Tattevin (Centre Hospitalier Universitaire de Rennes) ;
Hafid Ait-Oufella, Diane Bollens, Thibault Chiarabini, Patrick Ingiliz, Karine Lacombe, Benedicte Lefebvre, Jean-Luc Meynard, Nadia Valin (Hôpital Saint-Antoine, Assistance Publique – Hôpitaux de Paris) ; Elisabeth Botelho-Nevers, Sandrine Accassat, Celine Cazorla, Elodie De Magalhaes, Anne Fresard, Amandine Gagneux-Brunon, Marie-France Lutz, Jerome Morel, Flora Schein, Guillaume Thiery (Centre Hospitalier Universitaire de Saint-Étienne) ; Nathalie De Castro, Blandine Denis, Geoffroy Liegeon, Jean-Michel Molina, Emma Rubenstein, Mariagrazia Tateo (Hôpital Saint-Louis, Assistance Publique – Hôpitaux de Paris) ; Jessie Cattelan, Raphael Clere-Jehl, Francois Danion, Yves Hansmann, Julie Helms, Christine Kummerlen, Nicolas Lefebvre, Sebastien Loewert, Hamid Merdji, Ferhat Meziani, Alexandra Monnier, Walid Oulehri, Hassene Rahmani, Yvon Ruch, Antoine Studer, Charles Ambroise Tacquard, Axel Ursenbach (Centre Hospitalier Universitaire de Strasbourg) ; Marwa Bachir, Ruxandra Calin, Julie Chas, Ludovic Lassel, Marine Nadal, Christia Palacios, Gilles Pialoux, Martin Siguier (Hôpital Tenon, Assistance Publique – Hôpitaux de Paris) ; Muriel Alvarez, Fabienne Calvas, Helene Catala, Alexa Debard, Pierre Delobel, Monique Galitzky, Camille Garnier, Gaspard Grouteau, Nicolas Guibert, Pauline Lansalot-Matras, Simon Lauret, Lucie Lelievre, Guillaume Martin Blondel, Julien Mazieres, Elise Noelsavina, Sarah Pellerin, Sandrine Pontier-Marchandise, Lydie Porte, Gregoire Prevot, Stella Rousset, Kamila Sedkaoui, Claire Thalamas (Centre Hospitalier Universitaire de Toulouse) ; Frederic Bastides, Laetitia Bodet Contentin, Claudia Carvalho-Schneider, Francois Coustilleres, Pierre-Francois Dequin, Stephan Ehrmann, Thomas Flament, Denis Garot, Maeva Gourraud, Antoine Guillon, Sophie Jacquier, Charlotte Larrat, Annick Legras, Adrien Lemaignen, Marion Lacasse, Julie Mankikian, Emmanuelle Mercier, Lucile Rouault, Charlotte Salmon Gandonniere (Centre Hospitalier Universitaire de Tours) ; Tiphaine Guy, Marie Gousseff (Centre Hospitalier Bretagne-Atlantique, Vannes) ; Emma D'Anglejan (Centre Hospitalier de Versailles).
***Greece***: Paraskevi Fragkou, Ioannis Grigoropoulos, Charalampos Moschopoulos, Sotirios Tsiodras, Georgios Tsioulos, Nikolaos Tziolos (University General Hospital, Athens) ; Anastasia Kotanidou, Maria Theodorakopoulou, Edison Yajai (Evaggelismos General Hospital, Athens) ; Carolina

Akinosoglou, Vasiliki Dimakopoulou, Marcos Maragkos, Elena Polyzou, Argyrios Tzouvelekis (University General Hospital of Patras, Patras) ;

***Hungary:*** Imre Bakos, Eva Sandor, Istvan Varkonyi (Debreceni Egyetem Klinikai Központ Infektológiai Klinika, Debrecen) ; Richard Armos, Szili Balazs, Takacs Istvan (Semmelweis University-Department of Internal Medicine and Oncology, Budapest)

***Ireland:*** Colm Bergin, Brian Kent, Colm Kerr, Liam Townsend (St.James Hospital, Dublin)

***Luxembourg***: Myriam Alexandre, Nassera Aouali, Vic Arendt, Tania Bintener, Pierre Braquet, Esther Calvo Lasso de la Vega, Joelle Colling, Gaelle Damilot, Thomas Henin, Christian Michaux, Jean Reuter, Therese Staub, Lamia Skhiri, Christophe Werer, Ali Znati (Centre Hospitalier de Luxembourg) ;

***Norway:*** Jan Erik Berdal, Olav Dalgard, Ole Kristian Fossum (Akershus University Hospital, Lorenskog) ; Hedda Hoel Simen Schive (Lovisenberg Diakonal Hospital, Oslo) ; Ane-Kristine Fin, Aleksander-Rygh Holten, Synne Jenum, Maghild Eide Macpherson, Anne Ma Dyrhol Riise, Kristian Tonby (Oslo University Hospital, Oslo) ;

***Portugal***: Sandra Braz, Joao Miguel Ferreira Ribeiro (Centro Hospital Universitário de Lisboa Norte, Hospital de Santa Maria); José-Artur Paiva, Roberto Roncon-Albuquerque (Centro Hospitalar Universitário São João de Porto).

***Spain***: Sandra de la Rosa Riestra, Zaira Raquel Palacios Baena, Patricia Perez Palacios, Jesus Rodriguez Bano (Hospital Universitario Virgen Macarena, Sevilla)

#### Acknowledgements

We acknowledge all the patients enrolled in the study and all the health-care workers involved in patients' management.

We acknowledge the members of the Data Safety Monitoring Board:
Stefano Vella (chair), Karen Barnes, Guanhua Du, Mike Jacobs, Catia Marzolini, Donata Medaglini, Mical Paul, Stuart Pocock, Sylvie Van Der Werf, Patrick Yéni, Phaik Yeong Cheah and the independent statistician Tim Collier.

We acknowledge the DisCoVeRy Steering Committee (by alphabetical order):
Voting members: Florence Ader, Maude Bouscambert, Guislaine Carcelain, Dominique Costagliola, Alpha Diallo, Hélène Espérou, Joe Eustace, Richard Greil, Edit Hajdu, Monika Halanova, Maya Hites, France Mentre, José-Artur Paiva, Anna Piekarska, Jesus Rodriguez Bano, Marie-Thérèse Staub, Kristian Tonby, Milan Trojanek, Sotirios Tsiodras, Serhat Unal, Yazdan Yazdanpanah.
Non-voting members: José-Ramón Arribas Lopez, Pascale Augé, Nathalie Bergaud, Charles Burdet, Paraskevi C. Fragkou, Ahmet Cagkan Inkaya, Mireille Caralp, Beata Cecetkova, Florence Chung, Sandrine Couffin-Cadiergues, Aline Dechanet, Christelle Delmas, Jacques Demotes, Marina Dumousseaux, Alexander Egle, Fionnuala Keane, Soizic Lemestre, Julia Lumbroso, Clément Massonnaud, Noémie Mercier, Nicolas Minvielle, Marion Noret, Nathan Peiffer-Smadja, Carina Pereira, Julien Poissy, Jean Reuter, Juliette Saillard, Vida Terzic, Mary-Anne Trabaud, Marius Trøseid, Sarah Tubiana.

We acknowledge the French DisCoVeRy Trial Management Team (by alphabetical order):
Florence Ader, Nathalie Bergaud, Nicolas Billard, Maude Bouscambert, Charles Burdet, Dominique Costagliola, Sandrine Couffin-Cadiergues, Aline Dechanet, Christelle Delmas, Alpha Diallo, Marina Dumousseaux, Hélène Esperou, Assia Ferrane, Claire Fougerou-Leurent, Alexandre Gaymard, Benjamin Hamze, Lien Han, Maya Hites, Vinca Icard, Soizic Le Mestre, Benjamin Leveau, Clément Massonnaud, France Mentré, Noémie Mercier, Joe Miantezila-Basilua, Nicolas Minvielle, Marion Noret, Nathan Peiffer-Smadja, Juliette Saillard, Sarah Tubiana.

We acknowledge the Working Groups of the DisCoVeRy Trial (by alphabetical order):
Clinical aspects: Florence Ader, Charles Burdet, Christelle Delmas, Marina Dumousseaux, Maya Hites, Nathan Peiffer-Smadja, Juliette Saillard.
Study products: Sandrine Couffin-Cadiergues, Aline Dechanet, Christelle Delmas, Marina Dumousseaux, Assia Ferrane, Benjamin Hamze, Juliette Saillard.

Methodology and Statistics: Charles Burdet, Dominique Costagliola, Lien Han, Clément Massonnaud, France Mentré.

Data: Toni Alfaiate, Nicolas Billard, Charles Burdet, Aline Dechanet, Christelle Delmas, Claire Fougerou-Leurent, Annabelle Metois, Noémie Mercier, Joe Miantezila-Basilua, Nicolas Minvielle, Vida Terzic.

Virology: Maude Bouscambert, Charles Burdet, Alexandre Gaymard, Maya Hites, Benjamin Leveau, Bruno Lina, France Mentré, Florence Morfin-Sherpa, Bruno Simon, Hadrien Regue.

Immunology: Laurent Abel, Florence Ader, Nathalie Bergaud, Maude Bouscambert, Charles Burdet, Guislaine Carcelain, Alexandre Gaymard, Maya Hites, Clément Massonnaud, France Mentré, Nathan Peiffer-Smadja, Pascal Poignard, Mary-Anne Trabaud, Vincent Vieillard.

Genetics (host): Laurent Abel, France Mentré.

Genetics (SARS-CoV-2): Nathalie Bergaud, Grégory Destras, Laurence Josset, Hadrien Regue, Bruno Simon.

Collection of biological human samples: Aline Dechanet, Nathalie Bergaud, Christelle Delmas, Ouifiya Kalif, Benjamin Leveau, Valentine Piquard, Céline Sakonda, Sarah Tubiana.

Safety matters: Aline Dechanet, Christelle Delmas, Alpha Diallo, Assia Ferrane, Noemie Mercier, Joe Miantezila-Basilua, Nicolas Minvielle, Laurence Moachon, Nathan Peiffer-Smadja, Vida Terzic.

Regulatory affairs, Monitoring and Financial management: Sandrine Couffin-Cadiergues, Aline Dechanet, Christelle Delmas, Marina Dumousseaux, Hélène Esperou, Assia Ferrane, Claire Fougerou-Leurent, Soizic Le Mestre, Noemie Mercier, Nicolas Minvielle, Marion Noret, Juliette Saillard, Vida Terzic.

Communication: Florence Ader, Sandrine Couffin-Cadiergues, Aline Dechanet, Christelle Delmas, Marina Dumousseaux, Hélène Esperou, Claire Fougerou-Leurent, Maya Hites, France Mentré, Marion Noret, Nathan Peiffer-Smadja, Priscille Rivière, Juliette Saillard, Léa Surugue, Yazdan Yazdanpanah.

Drug supply*:* Christelle Delmas, Marina Dumousseaux, Juliette Saillard.

Inserm-Transfert*:* Pascale Augé, Mireille Caralp, Nathalie Dugas, Florence Chung, Julia Lumbroso.

Clinical Research Unit CHU Bichat-Claude Bernard*:* Camille Couffignal.

Data hosting: the COVID-19 Partners Platform and Hervé Le Nagard.

We acknowledge the executive management staff of Inserm: Gilles Bloch (CEO), Didier Samuel (CEO), François Chambelin, Carine Delrieu, Claire Giry, Jean-Christophe Hébert, Frédérique Le Saulnier, Claire de Marguerye, Dominique Pella, Damien Rousset, Teodora Yovkova.

We acknowledge the RENARCI network: Marion Noret, Albert Sotto and Pierre Tattevin.

We acknowledge the pharmacists, the virologists and all others research staffs:

*Austria:* Sabine Brennsteiner-Fahrner, Johanna Feldinger, Natascha Gager, Helmut Gringinger, Astrid Haab, Josef Klappacher, Margit Langanger, Ingrid Liedtke, Petra Luft, Eva Mertin, Doris Probst, Michaela Schachner, Verena Schrattenecker, Sabrina Sgonc, Karin Spitzlinger, Monika Wetscher.

*Belgium:* Tessa Acke, Buse Al, Sophiane Arbaj, Claire Barthelemy, Tatiana Besse, Anne Bouvier, Celine Daoust, Anuschka De Clercq, Fem De Plus, Hilde De Vlieger, Ingrid Dekeyser, Sofie Desmet, Inge Dreesen, Jarode Ducrot, Soukaina El Baroudi, Fatima El Bouazaoui, Adeline Gaillet, Vanessa Galand, Anne-Catherine Gerard, Jasmien Gevers, Severine Halleux, Estelle Hawia, Nabil Hayef, Angeline Henry, Youssef Jaadar, Pieter Jan De Jonghe, Nina Kamun Mwinkeu, Zineb Khalil, Valerie Kin, Dimphny Kindt, Idalie Lawson, Francoise Leunen, Justine Malaise, Sylvie Martens, Odette Mukangenzi, Viviane Ngungu-Yandja, Emelien Putzeys, Jente Reumers, Robert Rugwiro, Axelle Scarniere, Cinderella Smissaert, Eveline Van steenkiste, Ellen Vandekerkhof, Florence Vandenberghe Astrid Vandorpe, Kimberly VanHees, Lorenz Vanneste.

*Czech Republic:* Eva Bendova, Martina Bizosova, Alzbeta Dorota Dostalova, Petra Feberova, Daniela Harapatova, Zuzana Hrdlickova, Vera Kajerova, Barbora Krauzova, Jana Kriklanova, Hnat Kurhan, Jiri Litzman, Pavlina Martinkova, Elka Nycova, Silvia Rajecka, Filip Ruzicka, Miluse Smejkalova, Nadezda Sojkova, Vladimir Soska, Pavla Stepankova.

*France:* Jerome Aboab, Anne Adda, Genevieve Afantchao, Salima Allaf, Marie-Alexandra Alyanakian, Camille Alzina, Audrey Ambert, Corinne Amiel, Gaelle Amiotte, Maiwenn Andree-Bilien, Quam Aquereburu, Daphne Archinard, Clemence Arrive, Romain Atger, Mathilde Audry, Marine Aussedat, Sylvain Auvity, Christelle Auvray, Veronique Avettand, Loredana Baboi, Prissile Bakouboula, Caroline Barau, Marc Bardou, Aurelie Barrail Tran, Amelie Barreau, Laurie Barthelemi, Aurelie Baudin, Delphine Bauwens, Sophie Bayer, Christine Bechet, Guillaume Becker, Said Belhadj, Ghezzoul Bellabes, Gaetan Beltzung, Ines Ben Ghezala, Noussaier Ben Salem, Philippe Benoit, Raida Benrayana, Juliette Berger, Willy Berlier, Noemie Bernier, Virginie Bernigal, Marjory Berns, Marion Berthe, David Berthelot, Frederique Bertholon, Anais Beulaygue, Iris Bichard, Stephane Blanc, Stephanie Blin, Sophie Boddaert, Helene Boivin, Benedicte Bonnard, Carine Bonnin, Stephanie Bonte, Jeremy Bontemps, Karim Bouadam, Celia Boucheraine, Nihed Boughdiri, Elodie Boulay, Johan Bourbon, Philippine Bourrassaud, Maude Bouscambert-Duchamp, Nadege Bouskila, Caroline Boutin, Magali Bouvier, Meriem Bouzair, Nathalie Braghini, Maureen Brasseur, Sophie Breaud, Amenie Brebion, Sylvie Brice, Corinne Brochier, Pascal Brouard, Guillaume Brouillet, Anne-Sophie Bruyer, Isabelle Calmont, Valentine Campana, Anthony Carmona, Lauren Carraz, Muriel Carvalho Verlinde, Nadine Casimir, Nicolas Cassou, Sandrine Castelain, Audrey Castet-Nicolas, Cedric Castex, Guylaine Castor, Laetitia Caturla, Astrid Causel, Magali Centelles, Margaux Chabert, Marie-Laure Chaix, Lynda Chalal, Helene Chambrin-Lauvray, Malkhone Chansombat, Bruno Chanzy, Carole Charles, Julie Charligny, Delphine

Chartier, Caroline Chaudier, Ounissa Cherifi, Dominique Chiappini, Anne-Laure Clairet, Sabrina Cochennec, Isabelle Combes, Stephanie Cossec, Auriane Couderc, Helene Coyard, Amelie Cransac, Isabelle Credoz, Magali Cremel, Benjamin Cuinet, Olivia Da Conceicao, Anne Daguenel-Nguyen, Edith Dauchy, Marina Davier, Dominique De Debriel, Anne De Monte, Alexis De Rougemont, Lucile Decanter, Constance Delaugerre, Mickael Delaunay, Delphine Denis, Marine Deppenweiler, Lydia Derridj Rekkeb, Adrien Dersigny, Diane Descamps, Mireille Desille-Dugast, Maelle Detoc, Emma Devienne, Sandrine Dhenin, Aisse Diallo, Anne Claire Diata, Jerome Dimet, Nikita Dobremel, Regine Doncesco, Marie Dorard-Chabrier, Ouarda Douache, Bernard Drenou, Jaqueline Dubois, Clara Duran, Julie Durif, Charlene Duschene, Emilie Dussosoy, Loubna Elotmani, Jean-Francois Emile, Anne Esling, Laure Esteve, Samira Fafi-Kremer, Gaelle Fajole, Marie-Sarah Fangous, Magali Farines Raffoul, Raynald Feliho, Marie-Paule Ferdinand, Virginie Ferre, Julie Fillon, Arianna Fiorentino, Laurent Flet, Djeneba Fofana, Jean-Eudes Fontan, Catherine Francois, Alais Frelat, Valerie Furlan, Audrey Gabassi, Anas Gahbiche, Julie Gall, Jean Gallot-Lavalee, Valerie Galvan, Nicolas Gambier, Ludivine Garrier, Karen George, Taghi bijan Ghaleh Marzban, Carine Ghionda, Coralie Gibouleau, Morgane Gilg, Marion Girard, Lauriane Giroudot, Fanny Giroux, Claudine Gniadek, Cindy Godard, Sandrine Gohier, Geraldine Gonfrier, Alain Gravet, Jennifer Griffonnet, Isabelle Gros, Michele Grosdenier, Thomas Gualdi, Franck Guerin, Elodie Guerineau, Catherine Guichard, Audrey Guillier, Marie Gyselinck, Sofia Haddou, Smina Hadj Mahfoud, Estelle Haeberle, Catherine Hamon-Bouer, Cedric Hartard, Delphine Haye, Cecile Henquell, Anais Henric, Yves-Edouard Herpe, Berthe Marie Himbert, Catherine Hoskovec, Anne Hutt-Clauss, Vinca Icard, Meriem Imarazene, Cylia Imekhlaf, Lina Iratni, Elodie Issorat, Jacques Izopet, Aude Jacob, Elodie Jacqueroux, Julie Jambon, Valentine Jaspart-Le Du, Janick Jean-Marie, Julien Jeanpetit, Anne-Lise Jegu, Adeline Jeudy, Helene Jeulin, Camille Joachim, Stephanie Jolas, Estelle Josse, Caroline Jouy, Jean-Yves Jouzeau, Zelie Julia, Camille Jurado, Anna Kabala, Ouifiya Kafif, Jean- Daniel Kaiser, Marie Aude Kamp, Isabelle Komla Soukha, Gaella Kotokp Youkou, Laura Kramer, Marie Kroemer, Carine Labruyere, Blandine Lafitte, Sandra Lagarrigue, Caroline Laheurte, Laure Lalande, Jean-Louis Lamaignere, Julie Lamarque, Sylvie Lanier, Philippe Lanotte, Dihia Larkeche, Elodie Laugel, Severine Lauraire-Ho, Morgane Le Bras, Melaine Le Brazic, Chloe Le Brun, Manuela Le Cam, Philippe Le Corvoisier, Marie Le Donge, Delphine Le Febvre De Nailly, Sylvie Le Gac, Sophie Le Guen-Granville, Clotilde Le Tiec, Marie Lebouc, Jennifer Legrand, Quentin Lepiller, Marie-Laurence Lepilliez, Karine Leplus, Celine Leroy, Marianne Leruez-Ville, Hubert Lesur, Cyrielle Letaillandier, Benjamin Leveau, Nicolas Leveque, Sophie Lloret, Celine Loiseau, Sandra Lomazzi, Francoise Louni, Marie Louyot, Thomas Lucas, Maxime Luu, Isabelle Madelaine, Catherine Malaplate, Philippe Manivet, Jean-Michel Mansuy, Julie Marechal, Cecile Mariet, Eric Marquis, Guillaume Marrane, Nora Martel, Delphine Martineau, Solene Marty-Quinternet, Laurene Masson, Diane Maugars, Agnes Maurer, Clementine Mayala Kanda, Marianne Maynard, Julien Mazet, Mouniya Mebarki, Catherine Metzger, Marine Minier, Domitille Molinari, Gwenael Monnier, Maelle Monphile, Samantha Montagne, Clara Mora, Laurence Morand-Joubert, Florence Morfin Sherpa, Sophie Morice, Alexia

Moulin, Sophie Mugnier, Aline Murhula, Catherine Mutter, Marina Nabeih-Fakhori, Fatiha Najioullah, Pierre-André Natella, Krystel Nerini, Remi Neviere, Julianne Oddonne, Caroline Oger, Stanislas Ogoudjobi, Jeremie Orain, Laure Ottomani, Fariza Ouali, Lynda Oualit, Carole Ouisse, Sonia Ould Younes, Nabil Oumarou, Christophe Padoin, Maider Pagadoy, Gaelle Pagand, Jean-Baptiste Pain, Rachel Parat, Stephanie Parat, Maguy Parrinello, Elodie Parry, Anne Pattyn, Mathilde Pelletier, Veronique Pelonde-Erimee, Claire Pernin, Corinne Pernot, Pauline Perreau, Agnes Pezzi, Ketsia Pierre, Michael Pierre, Carine Pifaut, Sylvie Pillet, Sylvie Pineau, Valentine Piquard, Isabelle Pironneau, Amandine Pisoni, Lydie Poinsignon, Isabelle Popov, Chloe Porche, Isabelle Portier, Benedicte Poumaroux, Severine Poupblanc, Brigitte Pouzet, Joanne Praneuf, Anaele Pregny, Miresta Previlon, Michel Prevot, Florent Prion, Nina Pronina, Mathieu Putigny, Julie Quantin, Julie Queva, Charlene Raimbert, Pascaline Rameau, Carole Ramousset, Marie Ramstein, Florence Raynal, Benedicte Razat, Anais Razurel, Jennifer Reutenauer, Lucie Richard, Clemence Rieu, Aurelie Risal, Noemie Roberjot, Camila Rocaboy, Sophie Rochas, Audrey Rodallec, Veronique Ronat, Anne-Marie Roque-Afonso, Pauline Rosique, Patrick Rossignol, Guillaume Rousseau, Catherine Roussel, Jessica Rousson, Perrine Roux, Celine Rouzeau, Renee Flore Roy-Ema, Mathilde Rudelin, Sylvie Sacher Huvelin, Thiziri Sadaoui, Zaina Safir, Virginie Salas, Meriem Salhi, Nicolas Sans, Amandine Saubion, Celine Schaeffer, Anne Schieber Pachart, Evelyne Schvoerer, Verane Schwiertz, Justine Sevin, Hasni Si Albdelkader, Veronique Simeon, Florian Smagghe, Justine Sourice, Sylvie Spinar, Thomas Sternjacob, Valerie Suez-Panama, Muhtadi Suliman Haj Mukhtar, Marie Sylvain, Shabnam Taheri, Martine Tching-Sin, Fatima Tendjaoui, Elise Thebault, Marine Thibault, Vincent Thibault, Pelagie Thibaut, Alaki Thiemele, Elise Tissot, Marion Tissot, Irit Touitou, Thanh Huy Christian Tran, Thomas Tritz, Marie Capucine Trousset, Edouard Tuaillon, Sarah Tubiana, Bruno Turlin, Julie Vardanega, Charline Vauchy, Christelle Vauloup Felous, Marion Vazel, Veronique Venard, Celine Verstuyft, Celine Viaud, Magalie Vieille, Laurent Villeneuve, Solenne Villot, Sylvia Wehrlen-Pugliese, Celine Wilpotte, Rosa Wilson, Scarlett Wise.

***Greece :*** Eirini Daikou, Efthymios Dimitrakis, Helen Fytrou, Eleni Kraniotaki, Eleni Lamprou, Tsakona Maria, Georgios Schinas, Nikolaos Siafakas, Fani Veini.

***Hungary***: Vasarhelyi Barna, Ibolya Borsi, Viktoria Jurecsko, Szoke Katalin, Erzsebet Komaromi, Vecsei Margit, Chini Mohammadali.

***Ireland*** : Binny Benny, Niamh, Galvin, Aisling, Gavin, Edel, ODea, Nalini Panti,

***Luxembourg****:* Joel Aerts, Emmanuelle Andlauer-Vialette, Valerie Arnould, Gaelle Auquier, Carlo Braunert, Charlotte Delcave, Maria Dima, Laurence Floener, Jean-Hugues Francois, Sandrine Garnier, Gregory Gaudillot, Georges Gilson, Jerome Graas, Elise Jourdan, Isabelle Larosche, Sara Macchi,

Guillerme Marques, Aurore Martinez, Alessandra Moussel, Caroline Peelman, Laury Phelix, Daniela Valoura.

***Norway***:

Zeineb Al-Karbawi, Jessica Andreassen, Jannicke Dokken, Wenche Ekornhol, Simon Engebraten, Ingeborg Fiane, Dag Fossum, Gro Mellem Gulbrandsen, Ingeborg Holand, Marina Ilyas, Elisabeth Toverud Landaas, Gunn Helen Malmstrom, Mia Martinsson, Ingunn Melkeraaen, Katerina Nezvalova-Henriksen, Merie Nguen Dang, Sara Nur, Kjerstin Rostad, Linda Gail Skeie, Kristin Stoen Gunnarsen, Merete Tollefsen, Elin Westerheim.

Poland: Maciej Borowiec, Joanna Kazmierczak, Miroslawa Krajewska, Dalinda Szpruch.

***Portugal:*** Jesus Ana Lucia, Joao Andrade Ribeiro, Maria Arminda Leonor, Tatiana Bento, Diana Cacador, Raquel Carneiro, Ana Carrondo, Patricia Catarino Carvalho, Susana Cecilio, Vanessa Codea, Murielle Dardenne, Rui Farinha, Jose Friaes, Vanessa Gaspar Rodrigues, Tiago Guimaraes, Ana Sofia Homemde Mello, Ruben Jorge, Ana Lima, Ana Lopes, Tatiana Lopes Rodrigues, Antonio Madureira, Carla Melo, Joana MonteirodaCosta, Cristiana Mota, Olinda Ourique, Margarida Pacheco Carvalho, Domingas Palmas, Fernanda Pedro, Siliva Pereira, Daniela Ramos, Fernanda Reis, Raquel Ribeiro, Ana Ribeiro, Joao Ribeiro, Catia Rodrigues, Luis Santos, Fatima Soares Dias, Fernanda Valerio.

***Slovakia:*** Martina Antosova, Lucia Jedlickova, Michal Kolorz, Lucia Rahelova, Robert Rosolanka, Edita Trtilkova.

***Spain :***

Jose Manuel Carretero, Ivan Corriente, Marta Fernandez Regana, Ana Belen Gomez Sanchez, Antonio Gonzalez, David Antonio Gutierrez Campos, Virginia Palomo Jimenez, Elena Ruiz, Ana Silva Campos.

We acknowledge service providers : Cleanweb (eCRF), Excelya (monitoring aspects), Theradis PHARMA (Drug flow, IWRS and Collection of biological human samples).

We acknowledge the support of ASTRA-ZENECA and Biomerieux.
