## Supplementary documents for "Tixagevimab-cilgavimab (AZD7442) for the treatment of patients hospitalized with COVID-19 (DisCoVeRy): A phase 3, randomized, double-blind, placebo-controlled trial"

by Hites M., Massonnaud C.R., et al.

CONSORT 2010 checklist……………………….………………………………….. Page 2

Protocol V15.0………………………………………………………………………. Page 4

Statistical analysis plan for primary and secondary endpoints for version 15 and beyond of the protocol (evaluation of AZD7442)…………………………………………………………….… Page 95

Statistical analysis plan for exploratory endpoints –Virologic efficacy in respiratory and blood samples………………………………………………………………………………… Page 131

Statistical analysis plan for exploratory endpoints – Serum neutralization of SARS-CoV-2 in blood samples………………………………………………………………………………… Page 141

### CONSORT 2010 checklist of information to include when reporting a randomised trial*

| Section/Topic | Item No | Checklist item | Reported on page No |
|---|---|---|---|
| **Title and abstract** | | | |
| | 1a | Identification as a randomised trial in the title | 1 |
| | 1b | Structured summary of trial design, methods, results, and conclusions (for specific guidance see CONSORT for abstracts) | 6 |
| **Introduction** | | | |
| Background and objectives | 2a | Scientific background and explanation of rationale | 7 |
| | 2b | Specific objectives or hypotheses | 7 |
| **Methods** | | | |
| Trial design | 3a | Description of trial design (such as parallel, factorial) including allocation ratio | 8 |
| | 3b | Important changes to methods after trial commencement (such as eligibility criteria), with reasons | 8 |
| Participants | 4a | Eligibility criteria for participants | 8 |
| | 4b | Settings and locations where the data were collected | 8 |
| Interventions | 5 | The interventions for each group with sufficient details to allow replication, including how and when they were actually administered | 9 |
| Outcomes | 6a | Completely defined pre-specified primary and secondary outcome measures, including how and when they were assessed | 10 |
| | 6b | Any changes to trial outcomes after the trial commenced, with reasons | 10 |
| Sample size | 7a | How sample size was determined | 11 |
| | 7b | When applicable, explanation of any interim analyses and stopping guidelines | 13 |
| Randomisation: | | | |
| Sequence generation | 8a | Method used to generate the random allocation sequence | 9 |
| | 8b | Type of randomisation; details of any restriction (such as blocking and block size) | 9 |
| Allocation concealment mechanism | 9 | Mechanism used to implement the random allocation sequence (such as sequentially numbered containers), describing any steps taken to conceal the sequence until interventions were assigned | 9 |
| Implementation | 10 | Who generated the random allocation sequence, who enrolled participants, and who assigned participants to interventions | 9 |
| Blinding | 11a | If done, who was blinded after assignment to interventions (for example, participants, care providers, those | 9 |

| | | | |
|---|---|---|---|
| | | assessing outcomes) and how | |
| | 11b | If relevant, description of the similarity of interventions | 12 |
| Statistical methods | 12a | Statistical methods used to compare groups for primary and secondary outcomes | 12 |
| | 12b | Methods for additional analyses, such as subgroup analyses and adjusted analyses | 12 |
| **Results** | | | |
| Participant flow (a diagram is strongly recommended) | 13a | For each group, the numbers of participants who were randomly assigned, received intended treatment, and were analysed for the primary outcome | 13 |
| | 13b | For each group, losses and exclusions after randomisation, together with reasons | 13 |
| Recruitment | 14a | Dates defining the periods of recruitment and follow-up | 13 |
| | 14b | Why the trial ended or was stopped | 13 |
| Baseline data | 15 | A table showing baseline demographic and clinical characteristics for each group | Table 1 |
| Numbers analysed | 16 | For each group, number of participants (denominator) included in each analysis and whether the analysis was by original assigned groups | 14 |
| Outcomes and estimation | 17a | For each primary and secondary outcome, results for each group, and the estimated effect size and its precision (such as 95% confidence interval) | 14 |
| | 17b | For binary outcomes, presentation of both absolute and relative effect sizes is recommended | 14 |
| Ancillary analyses | 18 | Results of any other analyses performed, including subgroup analyses and adjusted analyses, distinguishing pre-specified from exploratory | 15 |
| Harms | 19 | All important harms or unintended effects in each group (for specific guidance see CONSORT for harms) | 16 |
| **Discussion** | | | |
| Limitations | 20 | Trial limitations, addressing sources of potential bias, imprecision, and, if relevant, multiplicity of analyses | 18 |
| Generalisability | 21 | Generalisability (external validity, applicability) of the trial findings | 17 |
| Interpretation | 22 | Interpretation consistent with results, balancing benefits and harms, and considering other relevant evidence | 17 |
| **Other information** | | | |
| Registration | 23 | Registration number and name of trial registry | 13 |
| Protocol | 24 | Where the full trial protocol can be accessed, if available | 13 |
| Funding | 25 | Sources of funding and other support (such as supply of drugs), role of funders | 13 |

Citation: Schulz KF, Altman DG, Moher D, for the CONSORT Group. CONSORT 2010 Statement: updated guidelines for reporting parallel group randomised trials. BMC Medicine. 2010;8:18.
© 2010 Schulz et al. This is an Open Access article distributed under the terms of the Creative Commons Attribution License (http://creativecommons.org/licenses/by/2.0), which permits unrestricted use, distribution, and reproduction in any medium, provided the original work is properly cited.

*We strongly recommend reading this statement in conjunction with the CONSORT 2010 Explanation and Elaboration for important clarifications on all the items. If relevant, we also recommend reading CONSORT extensions for cluster randomised trials, non-inferiority and equivalence trials, non-pharmacological treatments, herbal interventions, and pragmatic trials. Additional extensions are forthcoming: for those and for up-to-date references relevant to this checklist, see www.consort-statement.org.

La science pour la santé
From science to health

| Réservé au promoteur | N° Inserm | EU-DRACT | CPP | CNIL | N° Clinical Trial |
|---|---|---|---|---|---|
| | C20-15 | 2020-000936-23 | Réf. CPP: 3777<br>Réf. CNRIPH : 20.03.06.51744 | DR-2020-115 | NCT04315948 |
| | Qualification Réglementaire | ☒ RIPH du 1° médicament<br>☐ RIPH du 1° médicament thérapie innovante<br>☐ RIPH du 1° dispositif médical<br>☐ RIPH du 1° hors produit de santé | | ☐ RIPH du 2° à risques et contraintes minimes<br>☐ RIPH du 3° non interventionnelle | |

### Etude multicentrique, randomisée, adaptative, de l'efficacité et de la sécurité des traitements des patients adultes hospitalisés pour une infection COVID-19

### Multi-centre, adaptive, randomized trial of the safety and efficacy of treatments of COVID-19 in hospitalized adults

#### DisCoVeRy

**VERSION 15.0 of September 20th, 2021**

#### CONFIDENTIEL

| | |
|---|---|
| **Investigateur coordonnateur :**<br>**Florence Ader**<br>**Fonction** : **PU-PH (M.D., Ph.D)**<br>**Unité Inserm d'affiliation : Centre International de Recherche en Infectiologie (CIRI) - Inserm 1111**<br>**DR de rattachement : DR Auvergne Rhône Alpes**<br>**Adresse :**<br>**Service des Maladies infectieuses**<br>**Hopital de la Croix-Rousse, Hospices Civils de Lyon**<br>104, Grande-Rue de la Croix-Rousse<br>69317 LYON cedex 04, FRANCE<br>Tel : +33 (0)472 071 560<br>Mail: | **Méthodologie et statistique :**<br>**France Mentré / Charles Burdet**<br><br>**Unité Inserm d'affiliation : Infection. Antimicrobials. Modelling. Evolution (IAME), UMR 1137**<br>**Adresse:**<br>46 rue Henri Huchard, 75018 Paris, France<br>Tel : +33 (0)140 257 941<br>Email:<br><br> |

☐ **Monocentrique** ☐ **Nationale**
☒ **Multicentrique** ☒ **Européenne/Internationale**

| | | |
|---|---|---|
| **Promoteur :**<br>Inserm - Pôle Recherche Clinique (PRC)<br>Biopark, Bâtiment A, 8 rue de la Croix Jarry, 75013 Paris | **Contact France :**<br>Christelle Delmas,<br>Cheffe de projet<br><br><br>Tél: 01 82 53 33 68<br>Fax: 01 44 23 67 10 | **Contact Europe**<br>Juliette Saillard,<br>Cheffe de projet<br><br><br>Tél: 01 44 23 67 20<br>Fax: 01 44 23 67 10 |

CONFIDENTIAL

#### Summary of DisCoVeRy

**Terminology:** The novel coronavirus-induced disease described in December 2019 in Wuhan is designated COVID-19, and the causal RNA coronavirus is designated SARS-CoV-2.

**Background:** In early 2020, as this protocol was being initially developed, there were no approved treatments for COVID-19. As of January 2020, WHO identified remdesivir and lopinavir/ritonavir to be candidates of interest to be tested in clinical trials. Additional considerations based on emerging literature data have led to consider other drugs such as hydroxychloroquine and interferon ß-1a to be tested. Since then, additional drugs that also require evaluation have emerged, including monoclonal antibodies. The purpose of this randomized trial is to provide substantial evidence on the efficacy, or lack of efficacy and safety of these treatments.

**Randomisation:** This protocol describes a randomized trial among adults (≥18-year-old) hospitalized for COVID-19 that randomly allocates them between 2 arms: Standard of Care (SoC) + placebo *versus* SoC + AZD7442 (previous treatment arms were SoC + hydroxychloroquine suspended on May 24, 2020 and stopped on June 17, 2020, SoC + lopinavir/ritonavir and SoC + lopinavir/ritonavir plus interferon ß-1a, both stopped on June 29, 2020, and SoC + remdesivir which was stopped on 19/01/2021). Corticosteroids and anticoagulants have been added to the SoC on October 2, 2020. Other arms can be added as evidence emerges from other candidate therapeutics.

**Adaptive design:** The interim trial data will be monitored by a Data Safety Monitoring Board. If good evidence emerges while the trial is continuing that some other treatment(s) should also be evaluated, then it will be centrally decided that one or more extra arms may be added while the trial is in progress. However, in this version of the protocol, there will be no pre-planned interim analysis for efficacy nor futility, and the possibility to add arms will be very limited given the blinded and placebo-controlled nature of this phase of the study.

**Outcomes to be recorded:** At randomization, information is collected on the identity of the randomising physician and of the patient, and on age, sex, and an ordinal 7-point scale of severity. The main outcome is the clinical status on the ordinal 7-point scale at day 15.

**Numbers to be randomised:** A total of 620 patients per arm will be included.

CONFIDENTIAL

**Simplicity of procedures:** To facilitate collaboration even in hospitals that suddenly become overloaded, patient entry and all other trial procedures have been greatly streamlined.

**Need for DisCoVeRy protocol:** if many hospitals in several different countries collaborate through a compatible master protocol then reliable results will emerge more rapidly than if different hospitals or countries were to establish separate trials, and the findings are, therefore, more likely to be helpful in controlling the present pandemic. The interim results will be monitored confidentially, allowing the trial to continue across sites and countries without release of results in settings where an outbreak would wane before the trial study had reliably answered the principal questions it was designed to address.

CONFIDENTIAL

#### Link with Solidarity Trial

The DisCoVeRy protocol is based on the WHO Master protocol version 3.0 of March 3, 2020.

That WHO master protocol was largely based on a series of deliberations of the WHO R&D Blueprint Clinical Trials Expert Group, including clinical trialists, coronavirus experts, regulatory and ethics experts, and clinicians, some treating COVID-19 patients.[1] Based on these deliberations, the National Institute of Allergy and Infectious Diseases (NIAID) at the US National Institutes of Health drafted a Master Protocol, which was then adapted by the WHO to facilitate international implementation of the global Solidarity protocol.

From April 2nd, 2020 to January 19th, 2021 (stop of inclusion in remdesivir arm), DisCoVeRy was an add-on trial of the Solidarity Trial (protocol v15.0, August 6, 2020). At the end of follow-up of each patient included in a treatment arm shared between the DisCoVeRy and the Solidarity trials, the three outcomes of the Solidarity trial will be uploaded by the investigator on the specific website.

CONFIDENTIAL

---

[1] https://www.who.int/blueprint/priority-diseases/key-action/COVID-19_Treatment_Trial_Design_Master_Protocol_synopsis_Final_18022020.pdf
https://apps.who.int/iris/bitstream/handle/10665/330694/WHO-HEO-RDBlueprintnCoV-2020.4-eng.pdf
https://apps.who.int/iris/bitstream/handle/10665/330692/WHO-HEO-RDBlueprintnCoV-2020.2-eng.pdf

#### Statement of Compliance

The study will be carried out in accordance with the following, as applicable:

- The protocol version <to be completed> - date <to be completed>
- All National and Local Regulations and Guidance applicable at each site
- The International Council for Harmonisation of Technical Requirements for Registration of Pharmaceuticals for Human Use (ICH) E6(R2) Good Clinical Practice, and the Belmont Report: Ethical Principles and Guidelines for the Protection of Human Subjects of Research,
- National and ethical regulations

The signature below provides the necessary assurance that this study will be conducted according to all stipulations of the protocol including statements regarding confidentiality, and according to local legal and regulatory requirements, and ICH E6(R2) GCP guidelines.

Site Investigator Signature:

Signed: ______________________________ Date: ______________

Name:

Title:

CONFIDENTIAL

CONFIDENTIAL

CONFIDENTIAL

### 1. Protocol Summary

#### 1.1. Rationale for Proposed Clinical Study

In December 2019, the Wuhan Municipal Health Committee identified an outbreak of viral pneumonia cases of unknown cause. Coronavirus RNA was quickly identified in some of these patients. This novel coronavirus has been designated SARS-CoV-2, and the related coronavirus disease is termed COVID-19. Currently, there are no approved therapeutic agents available for coronaviruses. The purpose of this randomised trial is to provide substantial evidence on the efficacy, or lack of efficacy, and safety of these treatments given the large number of cases.

This protocol is based on the protocol produced by the National Institute of Health for the World Health Organization (WHO), version of March 3, 2020, which further led to the Solidarity protocol of WHO. This study supports the integration of the Solidarity WHO trial worldwide.

The supplementary investigations in the DisCoVeRy trial correspond to the add-on studies planned by the WHO Solidarity protocol: "Particular countries, or particular groups of hospitals, may want to collaborate in making further measurements or observations, such as serial virology, serial blood gases or chemistry, serial lung imaging, or serial documentation of other aspects of disease status (eg, through linkage to electronic healthcare records and routine medical databases)."

The current protocol lays out the general principles of how the multi-intervention trial would be implemented. As new interventions are added, the protocol will be amended and reviewed by IRBs/ECs and applicable regulatory agencies before implementation.

As of January 2020, WHO identified remdesivir and lopinavir/ritonavir to be candidates of interest to be tested in clinical trials Additional considerations based on emerging literature data have led to consider other drugs such as hydroxychloroquine, interferon ß-1a and monoclonal antibody AZD7442 to be tested.

In previous versions of the DisCoVeRy protocol, remdesivir, lopinavir/ritonavir with or without interferon ß-1a and hydroxychloroquine were evaluated as potential treatments for COVID-19. These treatments have been discontinued based on analyses review by both DSMC/DSMB, the Solidarity Executive Group and the DisCoVeRy steering committee.

CONFIDENTIAL

Nevertheless, there are still unmet medical needs for treating hospitalized patients with COVID-19; a highly effective anti-viral drug against SARS-CoV2 infection in this setting has not yet been identified. Therefore, in the present version of the protocol, the monoclonal antibody (mAb) AZD7442 developed by AstraZeneca is being evaluated. AZD7442 is a long-acting double mAb combination derived from convalescent patients that developed SARS-CoV-2 infection. Two antibodies (Abs) bind simultaneously to non-overlapping sites of the spike glycoprotein, thus blocking the receptor-binding domain of the spike from interacting with human angiotensin-converting enzyme 2 (ACE2) to neutralize wild-type SARS-CoV-2 virus infection. The cocktail has been engineered with half-life extension technology to extend neutralization potential durability following a single administration. The combination of two long acting mAbs, such as AZD7442, is also designed to reduce the risk of developing escape mutant strains.

Currently, several countries are enrolling in DisCoVeRy and new European countries will soon be launched. The recent launch of the 5-year multi-national 'European Research and Preparedness Network for Pandemics and Emerging Infectious Diseases' — EU-RESPONSE project coordinated by Inserm and funded by the European Commision will allow for rapid European expansion and further evolution of the DisCoVeRy trial, which is the work package 1 of EU-RESPONSE. The WP1 is the first deliverable of the project that will lead to a new multinational European multi-modal (phase II and III) adaptive platform trial for emerging infectious diseases in general (work package 2) [2].
https://ec.europa.eu/info/news/commission-supports-international-clinical-research-network-treat-covid-19-2020-sep-18_en).

#### 1.2. Study Design

DisCoVeRy is a randomized controlled trial among adults (≥18-year-old) hospitalized for COVID-19. This study is an adaptive, randomized, open or blinded, depending on the drug to be evaluated, clinical trial to evaluate the safety and efficacy of possible therapeutic agents in hospitalized adult patients diagnosed with COVID-19. The study is a multi-centre/country trial that will be conducted in various sites in Europe with Inserm as sponsor.

The study will compare different investigational therapeutic agents to a control group managed with the SoC including corticosteroids and anticoagulants. There will be interim monitoring to allow early stopping for safety and to introduce new therapies as they become available. If one therapy proves to be superior to others

[2] https://ec.europa.eu/info/news/commission-supports-international-clinical-research-network-treat-covid-19-2020-sep-18_en

CONFIDENTIAL

in the trial, this treatment may become part of the SoC for comparison(s) with new experimental treatment(s).

This version of the protocol, therefore, describes a randomized blinded placebo-controlled trial among adults (≥18-year-old) hospitalized for COVID-19 that randomly allocates them (1:1 ratio) between 2 arms: SoC + placebo *versus* SoC + AZD7442. The most recent changes to the protocol are **to allow inclusions of vaccinated** (fully or partly) **patients** against SARS-CoV2 in a proportion of 20% enrolled subjects, and to extend the period between the onset of symptoms and randomization from **less than 9 to less than 11 days.** The rationale for these changes is that this protocol is being implemented during this evolving pandemic, and therefore it must be adapted to it.

When the protocol was first written, vaccination against SARS-CoV2 had only just begun. Fortunately, vaccination coverage against SARS-CoV2 has significantly increased all over Europe, and around the world. However, it has become apparent that some vaccinated individuals do not develop adequate protection against circulating SARS-CoV2; these patients may develp COVID-19, they may require hospitalization, and they may have poor outcomes.[1–3] As numbers of vaccinated individuals increase around the world, it is important to determine whether the AZ-7442 monoclonal antibody cocktail is effective in non-vaccinated, but also in vaccinated individuals hospitalized with a symptomatic COVID-19.
The time period between onset of symptoms and randomization has also been extended because few patients are being hospitalized with less than 9 days of symptoms. Although it is believed that mABs should be administered as early as possible in the course of COVID-19 disease to have greatest efficacy, preliminary results from the Recovery trial have shown that the casirivimab + imdevimab cocktail is effective in improving outcomes in patients with negative serology for SARS-CoV2.[4] There is a good correlation between high viral loads for SARS-CoV2 and negative serology, and the primary analysis in this version of the protocol will indeed be performed in patients with a positive antigen test on nasopharyngeal swab at enrolment

Randomization will now be stratified by:

- Region (according to the administrative definition in each country)
- Antigenic status (positive or negative), obtained from the result of a rapid antigen test on nasopharyngeal swab performed at enrolment
- Vaccination initiation (yes or no): vaccination is initiated if at least one injection of any vaccine againt SARS-CoV- 2 was received prior to enrolment, whatever the delay.

CONFIDENTIAL

The primary analyses will be conducted on patients with antigen-positive results. A positive antigenic test is evidence of high viral shedding consistent with a recently started or uncontrolled infection. Overall, the number of antigen-negative patients will be at most 30% of all included subjects. The number of patients with vaccination (partly or fully) will be 20% of antigene positive patients and 20% of antigene negative patients. Sensitivity analyses will be performed in all patients, stratified by antigenic status and vaccination initiation.

Other arms can be added as evidence emerges regarding other therapeutic candidates. If additional arms are added or dropped from the trial, randomization will proceed with an equal probability of assignment to each of the remaining arms.

Because of the possibility that standards of supportive care may vary between trial centres and may also be optimized over time as more is learned about successful management of COVID-19, comparisons of safety and efficacy will be based on data from concurrently randomized participants.

A global independent data and safety monitoring board (DSMB) monitors interim data to make recommendations about early study closure or changes to conduct, including adding or removing treatment arms. However, the current version of the protocol does not allow for efficacy or futility analysis, and the ability to add trial arms will be limited by the study being blinded and placebo-controlled during the investigation of AZD7442.

All subjects will undergo a series of efficacy and safety assessments, including laboratory assays.

Subjects will be assessed at baseline, and at Days 3, 8 and 15 while hospitalized. Patients will be contacted by phone at Day 15 for evaluation of the Primary Endpoint if they have been discharged prior to Day 15-, and 14-days following hospital discharge for efficacy assessment.

Further follow-up assessments will be organized at Days 29, 90, 180, 365 and 456. If discharged from the hospital, days 29 and 90 assessments will be organized as outpatients' consultations for all. For Days 180 and 365 assessments, a subset of 25% of patients enrolled in centers with available resources and selected at Day 90 will be evaluated during a medical consultation, while the other will be contacted by phone. For Day 456, all patients will be contacted by phone.

CONFIDENTIAL

Nasopharyngeal swabs (NP) or lower respiratory tract samples will be obtained at baseline (Day 1 pre-treatment) and at Days 3, 8, 15 (while hospitalized) and 29 (while hospitalized or, if discharged from the hospital, in the outpatient setting).

Blood samples will be obtained at baseline (Day 1 pre-treatment) and at Days 3, 8, 15 (while hospitalized), at Days 29 and 90, and at Days 180 and 365 (for the subset of patients evaluated during a medical consultation at these times).
Thoracic computed tomography (CT)-scan will be obtained at baseline, depending on the centre's imagery capacities.

#### 1.3. Objectives and Endpoints

The overall objective of the study is to evaluate the clinical efficacy and safety of investigational therapeutics relative to the placebo arm among hospitalized adult patients who have COVID-19.

| **OBJECTIVES** | **ENDPOINTS (OUTCOME MEASURES)** |
|---|---|
| **Primary** | |
| Evaluate the clinical efficacy of different investigational therapeutics relative to the placebo arm in patients hospitalized with COVID-19 | Subject clinical status (on a 7-point ordinal scale) at Day 15:<br>1. Not hospitalized, no limitations on activities<br>2. Not hospitalized, limitation on activities<br>3. Hospitalized, not requiring supplemental oxygen<br>4. Hospitalized, requiring supplemental oxygen<br>5. Hospitalized, on non-invasive ventilation or high flow oxygen devices<br>6. Hospitalized, on invasive mechanical ventilation or ECMO<br>7. Death |
| **Key Secondary** | |
| Evaluate the efficacy of the investigational product on sustained recovery from index COVID-19 infection | Time from randomization to sustained recovery, defined as being discharged from the index hospitalization, followed by being alive and at home for 14 consecutive days prior to Day 90 |
| **Secondary** | |
| 1. Evaluate the **clinical efficacy** of different investigational therapeutics as compared to the placebo arm as assessed by: | |
| Ordinal scale:<br>• Subject clinical status on an ordinal scale on Days 29, 90, 180 and 365 | Status on an ordinal scale assessed at Day 29, 90, 180 and 365 |

CONFIDENTIAL

| OBJECTIVES | ENDPOINTS (OUTCOME MEASURES) |
| --- | --- |
| National Early Warning Score 2 (NEWS-2):<br>• Change from baseline to Days 3, 8, 15 (while hospitalized) and 29 in NEWS-2 | NEWS-2 at baseline (Day 1 pre-treatment), at Days 3 and 8, 15 (if patient is still hospitalized), and at Days 29 |
| Oxygenation:<br>• Oxygenation-free days during the first 28 days (to Day 29)<br>• Incidence and duration of new oxygen use, non-invasive ventilation or high flow oxygen devices during the first 28 days (to Day 29) | Duration of supplemental oxygen (if applicable) |
| Mechanical Ventilation:<br>• Ventilator-free days during the first 28 days (to Day 29).<br>• Incidence and duration of invasive mechanical ventilation use during the first 28 days (to Day 29)<br>• Need for mechanical ventilation or death by Day 15 | Duration of mechanical ventilation (if applicable)<br><br>Mechanical ventilation or death between baseline and Day 15 |
| Hospitalization<br>• Time to hospital discharge from randomization | Date of discharge from hospital |
| Mortality<br>• In-hospital mortality<br>• 29-day mortality<br>• 90-day mortality<br>• 180-day mortality<br>• 365-day mortality<br>• 456-day mortality | Date and cause of death (if applicable) |
| Long term health status<br>• Any hospitalization<br><br>• Any confirmed re-infection with SARS-CoV-2 | Occurrence of new hospitalization between discharge from index hospitalization and Days 90, 180 and 365<br><br>Occurrence of confirmed re-infection with SARS-CoV-2 between discharge from index hospitalization and Days 90, 180 and 365 |
| 2. Evaluate **<u>the safety</u>** of different investigational therapeutics through 456 days of follow-up as compared to the control arm as assessed by: | |
| Cumulative incidence of serious adverse events (SAEs) | SAEs |
| Cumulative incidence of Grade 3 and 4 events (AEs) | Grade 3 and 4 adverse events |
| Cumulative incidence of Grade 1- 2 hypersensitivity-related and infusion related AEs until D29 visit | Grade 1-2 hypersensitivity-related and infusion related AEs until D29 visit |
| Cumulative incidence of AEs of Special Interest | AEs of Special Interest |
| Discontinuation of investigational therapeutics (for any reason) | Discontinuation of investigational therapeutics (for any reason) |
| **Exploratory** | |

CONFIDENTIAL

| OBJECTIVES | ENDPOINTS (OUTCOME MEASURES) |
|---|---|
| Evaluate the virologic efficacy of different investigational therapeutics as compared to the control arm as assessed by:<br>• Percent of subjects with detectable SARS-CoV-2 in NP or lower respiratory tract sample at baseline (Day 1 pre-treatment) and at Days 3, 8, 15 (while hospitalized) and 29<br>• Normalized quantitative SARS-CoV-2 viral load in NP or lower respiratory tract sample at baseline (Day 1 pre-treatment) and at Days 3, 8, 15 (while hospitalized) and 29<br>• Normalized quantitative SARS-CoV-2 viral load in blood at baseline (Day 1 pre-treatment) and at Days 3 and 8<br>• Detection of variants of SARS-CoV-2 in NP or lower respiratory tract sample at baseline (Day 1 pre-treatment) to determine which variant is responsible for the infection) and at Days 3, 8, 15 (while hospitalized) and 29, and in case of transfer into ICU, to monitor the emergence of escape or virulence mutations) | Qualitative (Ct value) and quantitative PCR (normalized viral load) for SARS-CoV-2 in NP swab or lower respiratory tract sample at baseline (Day 1 pre-treatment), at Days 3 and 8, 15 (if patient is still hospitalized), and at Days 29<br><br>Qualitative (Ct value) and quantitative (viral load) PCR for SARS-CoV-2 in blood at baseline (Day 1 pre-treatment) and at Days 3, and 8 (if patient is still hospitalized)<br><br>Whole Genome Sequencing in NP or lower respiratory tract sample at baseline (Day 1 pre-treatment) and at Days 3, 8, 15 (while hospitalized) and 29, and in case of transfer into ICU |
| Evaluate the pharmacokinetics of AZD7442 | On Day 1, plasma concentration before and at the end of infusion (peak)<br>Plasma concentration at Days 3 and 8, 15 (if patient is still hospitalized), and at Days 29, 90, and in a subset of 25% of enrolled patients, at Days 180 and 365 |
| Evaluate the development of anti-drug antibodies (ADA) against the monoclonal antibodies | Anti-AZD7442 antibodies on Day 1 (pre-treatment), D15 (while hospitalized), Days 29, 90, and in a subset of 25% of enrolled patients, at Days 180 and 365 |
| Identify host genetic variants having an impact:<br>(1) in the development of severe clinical disease in individuals infected by SARS-CoV-2<br>(2) in the response in terms of safety and efficacy to investigational antiviral drugs | Whole genome sequencing (WGS) of the subjects |
| Evaluate the immune-inflammatory response during treatment of COVID-19 | Blood levels of immunothrombosis markers, IFNs (and anti-IFNs antibodies), cytokines and complement activation products at baseline (Day 1 pre-treatment), at Days 3 and 8, 15 (if patient is still hospitalized), and |

| OBJECTIVES | ENDPOINTS (OUTCOME MEASURES) |
|---|---|
| | at Days 29, 90, and in a subset of 25% of enrolled patients, at Days 180 and 365 |
| Evaluate the kinetics and functional capacities of patients anti-SARS-CoV-2 antibodies to evaluate the added value of the therapeutic antibody compared to the host's immunity | Blood levels and functional capacities of patients anti-SARS-CoV-2 immunoglobulins at baseline (Day 1 pre-treatment), at Days 3 and 8, 15 (if patient is still hospitalized), and at Days 29, 90, and in a subset of 25% of enrolled patients, at Days 180 and 365. SARS-CoV-2 and ADCC-NK functional neutralization ability of the blood at baseline (Day 1 pre-treatment) and at Days 3, 8, (while hospitalized), 29, and in a subset of 25% of enrolled patients, at Day 180 |

#### 1.4. Study Population

The study population is hospitalized adult (≥18 years old) patients with COVID-19. Selection criteria are presented below.

##### 1.4.1. Inclusion criteria

1. Adult ≥18 years of age at the time of enrolment
2. Hospitalized patients with any of the following criteria:
   a. the presence of pulmonary rales/crackles on clinical exam OR
   b. $SpO_2 \leq 94\%$ on room air OR
   c. requirement of supplementary oxygen including high flow oxygen devices or non-invasive ventilation
3. A time between onset of symptoms and randomization of **less than 11 days**
4. A positive SARS-CoV-2 PCR performed on a NP swab **within the 5 days** preceding randomization
5. The result of a rapid antigen test performed on a NP swab **within the 6 hours** preceding randomization
6. Contraceptive use by men or women.
   a. Male participants: Contraception for male participants is required; to avoid the transfer of any fluids, all male participants must use a condom from Day 1 and agree to continue for 90 days following administration of IMP.
   b. Female participants: Women of child-bearing potential must agree to use contraception for 365 days following administration of IMP. Acceptable birth control methods are listed in section 8.5.

CONFIDENTIAL

##### 1.4.2. Exclusion criteria

1. Refusal to participate expressed by patient or legally authorized representative
2. Need for invasive mechanical ventilation and/or ECMO at the time of enrolment
3. Spontaneous blood ALT/AST levels > 5 times the upper limit of normal
4. Glomerular filtration rate (GFR) < 15 mL/min or requiring maintenance dialysis
5. Pregnancy or breast-feeding
6. Anticipated transfer to another hospital, which is not a study site within 72 hours following randomization
7. Known history of allergy or reaction to any component of the study drug formulation.
8. Previous hypersensitivity, infusion-related reaction, or severe adverse reaction following administration of monoclonal or polyclonal antibodies.
9. Any prior receipt of investigational or licensed mAb/biologic indicated for the prevention of SARS-CoV-2 infection or COVID-19, and for those not vaccinated, expected receipt of vaccine in the 30 days following hospital discharge, according to current recommendation in each country.
10. Any medical condition which, in the judgment of the investigator, could interfere with the interpretation of the trial results or that preludes to protocol adherence.

#### 1.5. Study Phase

Phase 3

#### 1.6. Sites

Site selection will be determined as information becomes available about the epidemiology of COVID-19, and sites will be activated based on the number of local/regional cases and the willingness of local investigators to participate in the study. Multiple sites will need an appropriate Institutional Review Board (IRB) or Ethics Committee (EC) approval, but activation will be dependent on the incidence of COVID-19 at the site.

#### 1.7. Study intervention

All participants will receive the SoC of the participating center. Since October 2, 2020, corticosteroids and anticoagulants are part of the SoC, following the results from recent literature and the agreement of members of the DisCoVeRy trial Steering Committee.

CONFIDENTIAL

Participants randomized to the AZD7442 group will receive a total dose of 600 mg AZD7442 *via* a co-administered (both mAbs mixed in a single infusion) single IV infusion at a maximal infusion rate of 50 mg/minute. The patients randomized to the placebo arm will receive a saline solution. Preparation of an opaque vial will be made by an unblinded pharmacist (or his/her designee), and administration will be performed by a blinded nurse. Patients and investigators will be blinded.
All participants will be monitored closely for 2 hours after the infusion, as there is a risk of infusion reaction and hypersensitivity (including anaphylaxis) with any biological agent.

#### 1.8. Study Duration

The study will last for up to 3 years.
An individual subject will complete the study in 456 days, from screening at day -1 to follow-up on day 456 ± 14 days. The end of the study is defined as completion of the last 456-day data collection in the trial.

#### 1.9. Schedule of Assessments

CONFIDENTIAL

#### Table 1: Schedule of Assessments

| Day +/- Window | Screening[1] | Baseline (Day 1)[2] | 2-14 | 15[3] ± 2 | 29[4] ± 3 | 90[4] ± 7 | 180[5] ± 14 | 365[5] ± 14 | 456[5] ± 14 |
|---|---|---|---|---|---|---|---|---|---|
| **ELIGIBILITY** | | | | | | | | | |
| Eligibility criteria | X | | | | | | | | |
| Informed consent | X | | | | | | | | |
| Review SARS-CoV-2 results | X | | | | | | | | |
| Rapid Antigen test | X | | | | | | | | |
| **STUDY INTERVENTION** | | | | | | | | | |
| Randomization[12] | | X | | | | | | | |
| Standard of Care (SoC) + placebo | | Administration of SoC according to guidelines[13] Single administration of placebo at day 1 | Administration of SoC according to guidelines | | | | | | |
| Or SoC + AZD7442 | | Administration of SoC according to guidelines[13] Single administration of AZD7442 on day 1 | Administration of SoC according to guidelines | | | | | | |
| **STUDY PROCEDURES** | | | | | | | | | |
| Demographics & Medical History | X | | | | | X[6] | X[6] | X[6] | X[6] |
| Vital status | | | Daily while hospitalized | X | X | X | X | X | X |
| Medication review | X | | Daily while hospitalized[11] | X | X | X[14] | X[14] | X[14] | X[14] |
| NEWS-2 | | X | Days 3, 8 (all ± 1 day) if hospitalized | X | X | | | | |
| Ventilation mode | | X | Daily while hospitalized[11] | X | X | | | | |
| Oxygenation | | X | Daily while hospitalized[11] | X | X | | | | |
| Ordinal scale | | X | | X | X | X | X | X | |
| Thoracic CT scan | X | | | | | | | | |
| Adverse event evaluation | X | X | Daily while hospitalized | X | X | X | X | X | X |
| **SAFETY LABORATORY** | | | | | | | | | |
| Safety biological and inflammatory tests[7,8] | X[9] | | Day 3, 8 (all ± 1 day) if hospitalized | X | X | X | X[10] | X[10] | |
| Urinary pregnancy test for females of childbearing potential | X[9] | | | X | X | X | X[10] | X[10] | |
| Qualitative (Ct value) PCR for SARS-CoV-2 in NP swab or lower respiratory tract sample | | X[15] | Day 8 (± 1 day) if hospitalized[16] | | | | | | |
| **RESEARCH LABORATORY** | | | | | | | | | |
| Serum concentration of AZD7442 | | X (before and at end of infusion) | Days 1 (end of infusion) Days 3, 8 (all ± 1 day) if hospitalized | X | X | X | X[10] | X[10] | |
| Serum sample for AZD7442 ADA assessment | | X (before infusion) | | X | X | X | X[10] | X[10] | |
| Serum for exploratory objectives | | X | Days 3, 8 (all ± 1 day) if hospitalized | X | X | X | X[10] | X[10] | |
| Plasma for exploratory objectives | | X | Days 3, 8 (all ± 1 day) if hospitalized | X | X | X | X[10] | X[10] | |
| Whole blood for genetic analysis[18] | | | X | | | | | | |
| Nasopharyngeal swab and/or lower respiratory tract samples (for patients in the Intensive | | X | Days 3, 8 (all ± 1 day) if hospitalized On the day of transfer into ICU, if applicable | X | X | | | | |

CONFIDENTIAL

| Day +/- Window | Screening[1] | Baseline (Day 1)[2] | 2-14 | 15[3] ± 2 | 29[4] ± 3 | 90[4] ± 7 | 180[5] ± 14 | 365[5] ± 14 | 456[5] ± 14 |
|---|---|---|---|---|---|---|---|---|---|
| Care Unit) for exploratory objectives | | | | | | | | | |

*Notes:*

1. *Refer to Section 10.1 of the protocol for details of data to be collected at screening*
2. *Baseline assessments will be performed prior to study drug administration (Day 1 pre-treatment)*
3. *Data will be collected only during hospitalization, except for the ordinal scale which will be collected by phone for discharged patients*
4. *If discharged from the hospital, visits will be conducted in the outpatient setting*
5. *By phone call or during a medical consultation*
6. *Information on re-hospitalization and SARS-CoV-2 reinfection*
7. *Creatinine; Liver enzymes and hepatic function (LDH, ALT, AST); cell counts (neutrophils and total lymphocyte counts, haemoglobin, platelets); coagulation and inflammation status (fibrinogen, D-Dimer, prothrombin time, C-reactive protein, ferritinemia, serum total immunoglobulins G. Serum total immunoglobulins G assessed only at screening)*
8. *Any laboratory tests performed as part of routine clinical care within the specified visit window can be used for safety laboratory testing*
9. *Laboratory tests performed in the 36 hours prior to enrolment will be accepted*
10. *During a medical consultation, in a subset of 25% of enrolled participants*
11. *In hospitalized patients, medication review, oxygenation and ventilation mode will be collected up to day 29*
12. *Randomization will be done in the 6 hours following rapid antigen testing*
13. *SoC should be administered on day 1 in the 24 hours following randomization*
14. *Concomitant medicatians will be captured based on potential relatedness to AEs post D29 visit*
15. *Ct value and lineage of the virus performed during hospitalization and before infusion will be accepted*
16. *Qualitative (Ct value) for SARS-CoV-2 in NP swab or lower respiratory tract sample if patient hospitalized at D8*
17. *If the participant agrees to the host genome analysis*

CONFIDENTIAL

### 2. Introduction

#### 2.1. Study Rationale

COVID-19 is a respiratory disease caused by a novel coronavirus (SARS-CoV-2) and causes substantial morbidity and mortality. A limited number of vaccines and COVID-19 antibody therapies have received authorization for emergency use in selected countries, such as the US and UK, and other areas are expected to follow. Given the great number of individuals touched by the disease and the heterogeneous clinical presentation of the COVID-19, there is still a vast unmet need for therapeutic options. This clinical study is designed to evaluate potential therapeutics for the treatment of adult patients hospitalized with COVID-19.

At present, there is no specific antiviral therapy for coronavirus infections.
Few treatment studies have been conducted because most human coronavirus strains cause self-limited disease and care is supportive.

After the SARS coronavirus was identified in 2002 during a large outbreak, there was an increased interest in the development of specific therapeutic agents. SARS-CoV case-patients were treated with corticosteroids, type 1 IFN agents, convalescent plasma, ribavirin, or lopinavir/ritonavir based on pre-clinical data supporting potential efficacy except for ribavirin. Since the SARS-CoV outbreak, new therapeutic agents targeting viral entry proteins, proteases, polymerases, and methyltransferases have been tested, however, none of them has been shown to be effective in clinical trials.

#### 2.2. Background

##### 2.2.1. Purpose of Study

Coronavirus (CoVs) are positive-sense single stranded enveloped RNA viruses, many of which are commonly found in humans and cause mild symptoms. Over the past two decades, emerging pathogenic CoVs capable of causing life-threatening disease in humans and animals have been identified, namely severe acute respiratory syndrome coronavirus (SARS-CoV) and Middle Eastern respiratory syndrome coronavirus (MERS-CoV).

In December 2019, the Wuhan Municipal Health Committee (Wuhan, China) identified an outbreak of viral pneumonia cases of unknown cause. Coronavirus RNA was quickly identified in some of these patients. This novel coronavirus has been abbreviated as SARS-COV-2 and has 89% nucleotide identity with bat SARS-like-CoVZXC21 and 82% with that of human SARS-CoV [5].

CONFIDENTIAL

This novel coronavirus has been designated SARS-CoV-2, and the disease caused by this virus has been designated COVID-19. Outbreak forecasting and mathematical modelling suggest that these numbers will continue to rise [6]. Global efforts to evaluate novel antivirals and therapeutic strategies to treat COVID-19 have intensified. There is an urgent public health need for rapid development of novel interventions.

##### 2.2.2. Potential therapeutics

**Potential antiviral drugs**

As of January 2020, WHO identified remdesivir and lopinavir/ritonavir to be interesting candidates to be tested in clinical trials. Additional considerations based on emerging data available on March 29th, 2020 led to consider other drugs such as hydroxychloroquine and interferon ß-1a to be tested.

Consequently, remdesivir, hydroxychloroquine, lopinavir/ritonavir either alone or in combination with IFN-ß1a were identified as drug candidates to be evaluated based on their broad antiviral activity, the *in-vitro* and *in-vivo* data showing activity against coronaviruses, preliminary clinical data in patients with COVID-19 and extensive clinical safety database (see below). They were implemented in the DisCoVeRy trial.

On May 23, 2020, in light of new available data and aware of the investigators' responsibility to ensure patient safety when conducting randomised trials with the hydroxychloroquine arm, the Solidarity Executive Committee, which includes a member representing DisCoVeRy, had an extraordinary meeting.

They decided by mutual agreement during this meeting to issue the following recommendations:
- to establish communication between the various international randomized trials in progress to exchange on the results of possible interim analyses of the hydroxychloroquine/SoC arm *versus* SoC;
    - to conduct simultaneously a comprehensive review of all existing published data on hydroxychloroquine for the treatment of COVID-19 from randomised and non-randomised studies;
    - to temporarily suspend, with immediate effect, the allocation of patients in the hydroxychloroquine arm until further notice, with no pre-supposition as to what further action should be taken.

Consequently, the DisCoVeRy trial, sponsored by Inserm, continued in France and other European countries with suspension of the hydroxychloroquine arm after May 24, 2020.

CONFIDENTIAL

At this stage, close communication was established between the independent data monitoring committees of three international trials (DisCoVeRy, Solidarity, Recovery) to share the conclusions of the interim analyses. This allowed them to consider the combined evidence from three major randomized clinical trials, each evaluating hydroxychloroquine vs. SoC for treatment of hospitalized COVID-19 patients in different countries around the world. The results confirmed with a high degree of confidence that hydroxychloroquine was not effective at reducing mortality in hospitalized COVID-19 patients. These results are consistent with a growing number of national guidelines for the treatment of COVID-19 that do not support the use of hydroxychloroquine for hospitalized patients such as the French guidelines [7] or the NIH guidelines in the USA [8].

On June 17, 2020, based on these analyses and a review of the available published evidence [9–13], the Solidarity Executive Group and the DisCoVeRy steering committee made the common decision to cease the hydroxychloroquine arm of the Solidarity/DisCoVeRy trial.

On June 27, 2020, based on analyses review by both DSMC/DSMB, the Solidarity Executive Group and the DisCoVeRy steering committee made the common decision to cease the lopinavir/ritonavir and lopinavir/ritonavir plus interferon ß-1a arms of the Solidarity/DisCoVeRy trial for futility.

It remained important to continue the evaluation of remdesivir compared to the SoC for COVID-19 patients hospitalized to establish evidence regarding the effect of remdesivir on mortality. This evaluation, therefore, has continued in the WHO Solidarity trial in which centers on all continents of the world participate (> 30 participating countries) and in the DisCoVeRy clinical trial across Europe (centers open in France, Luxembourg, Austria, and Belgium).

Although there have been some indications that COVID-19 patients could benefit from remdesivir therapy, the overall conclusion by the non-profit Magic Evidence Ecosystem Foundation (MAGIC) based on data from over 7000 patients across the 4 trials (including the WHO Solidarity Trial) is that "the evidence suggested no important effect on mortality, need for mechanical ventilation, time to clinical improvement, and other patient-important outcomes". Based on this analysis, the WHO current guidelines recommend against the use of remdesivir in COVID-19 patients (https://www.who.int/news-room/feature-stories/detail/who-recommends-against-the-use-of-remdesivir-in-covid-19-patients). Thus, remdesivir was not further investigated in the Solidarity trial; however, investigators were allowed to use remdesivir at their own discretion and inclusions continued in the DisCoVeRy trial.

CONFIDENTIAL

On January 13th, 2021, the Discovery DSMB evaluated an interim report based on 776 patients of whom 389 received remdesivir and 387 received standard of care. The DSMB recommended that patient recruitment be suspended because of lack of evidence of efficacy of remdesivir after 15 days and a very low probability to conclude with the inclusion of additional participants. There was also no evidence for treatment efficacy at day 29 (on the same scale or on mortality), nor in the analysis restricted to moderate-risk participants at day 15. This recommendation has been endorsed by the Discovery Europe Steering Committee on January 19th 2021.

Nevertheless, there are still unmet medical needs for treating hospitalized patients with COVID-19; a highly effective anti-viral drug against SARS-CoV2 infection in this setting has not yet been identified. In this light, recent *in-vitro*, *in-vivo* and some clinical data in humans suggest that mabs directed against the spike protein of SARS-CoV2 may be a promising therapeutic agent against coronaviruses.

Monoclonal antibodies are used to bind to one specific substance in the body to mimic, block, or cause changes to provide a very specific and effective therapeutic intervention for a disease. Over the last year, we have learned that SARS-CoV2 has a spike protein made up of two sub-units (S1 and S2); this protein is very important for cell entry and infection as it binds to human epithelial cells via the ACE2 receptor. Neutralizing monoclonal antibodies aim to block the interaction of the virus with the ACE2 host cell receptor, thus inhibiting viral entry.

Very recently, two mAbs have received Emergency Use Authorization by the FDA and other regulatory agencies (including the European Medicines Agency) for the treatment of mild to moderate infection.
The first Emergency Use Authorization (EUA) to use bamlanivimab against SARS-CoV-2 was issued in November 2020 by the FDA for the early treatment (within 10 days of symptoms onset) of high-risk patients. The EUA excluded patients who are hospitalized or who require oxygen therapy due to COVID-19. In March 2021, the European Medicines Agency (EMA) issued that bamlanivimab, alone or in combination with Etesevimab could be used for patients with COVID-19 who do not require oxygen supplementation [3]. The efficacy and the safety of the combination is based on two clinical studies i) a phase 2, randomized, double-blind study in 577 individuals with mild to moderate infection, with a virological primary endpoint (significant change in viral load from baseline on day 11) ii) preliminary results from a randomized phase 3 study, double blind, placebo controlled, in 1035 high-risk individuals with a mild to moderate infection, with a

[3] https://www.ema.europa.eu/en/documents/referral/eli-lilly-company-limited-antibody-combination-bamlanivimab/etesevimab-covid19-article-53-procedure-conditions-use-conditions-distribution-patients-targeted_en.pdf

CONFIDENTIAL

clinical primary endpoint (hospitalization/death). In the latter study (BLAZE-1), the combination bamlanivimab/etesevimab was associated with a reduction in the risk of hospitalization and death that varied between 70 to 85%, depending on the dosage used.

The second combination is REGN-COV-2 (Regeneron), a double mAb combination of casirivimab and imdevimab which showed better viral clearance and fewer COVID-19 related medical visits compared to placebo in patients with mild COVID-19 (NCT04425629). In the interim analysis in which data from 275 patients were reported, the REGN-COV2 antibody cocktail reduced viral load, with a greater effect in patients whose immune response against SARS-CoV2 had not yet been initiated or who had a high viral load at baseline [14]. Safety outcomes were similar in the treatment and the placebo groups. Early March 2021, preliminary data of a phase 3 study in 4567 infected non-hospitalized patients were released [4], suggesting that the study met its primary endpoint, with casirivimab/imdevimab significantly reducing the risk of hospitalisation or death by 70% compared to placebo.

It is important to note that that the studies mentioned above focused on non-hospitalized patients, treated within 5 days of symptom onset. At the present time, there is no demonstration that monoclonal antibodies can be beneficial to hospitalized patients. Accordingly, in France, a Temporary Use Authorization (TUA) was issued on 02/27/2021 for the use of bamlanivimab in the treatment of adults with a positive PCR for SARS-CoV-2 and a mild to moderate COVID-19 and at high risk of progressing to a severe form of COVID-19 within 5 days of symptoms onset. Then TUAs were issued on 03/08/2021 for the combination bamlanivimab/etesevimab and for the combination casirivimab/imdevimab for patients at high risk of progressing to a severe form of COVID-19 and within 5 days of symptom onset as well. All these TUA concern patients with a mild or moderate form of COVID-19 and exclude patients who require oxygen therapy. DisCoVeRy, on the contrary, include patients hospitalized for COVID-19 who have evidence of lower respiratory disease or who require supplemental oxygen.

The use of monoclonal antibodies is currently being tested in hospitalized patients with moderate to severe disease, in particular in the framework of the Activ-3 platform, which recruits on patients within 12 days of symptom onset. In the framework of this platform, bamlanivimab study arm has been stoped for futility by DSMB. In a firm-sponsored study, REGN-COV-2 study arm in hospitalized

[4] https://www.roche.com/media/releases/med-cor-2021-03-23.htm

CONFIDENTIAL

patients with high oxygen requirements was stopped for futility [5]; preliminary results suggest encouraging results in the arm including patients with low oxygen requirements [6]. REGN-COV-2 is also currently being tested in patients with moderate to severe COVID-19 in the UK Recovery trial (NCT04426695).

In the present version of the protocol, an arm was added to the DisCoVeRy trial to investigate the benefit of AZD7442 (AstraZeneca) in COVID-19 patients. AZD7442 is a long acting double mAb combination derived from convalescent patients that developed SARS-CoV-2 infection. Two Abs bind to non-overlapping sites of the spike glycoprotein resulting in a synergistic blocking of the receptor-binding domain of the spike from interacting with human angiotensin-converting enzyme 2 (ACE2) neutralizing wild-type SARS-CoV-2 virus infection. The cocktail has been engineered with half-life extension technology to extend neutralization potential durability following a single administration. The combination of two long acting mABs such as AZD7442 is also designed to reduce the risk of developing escape mutant strains. Importantly this cocktail maintains an efficient neutralizing activity against the variants of concern circulating in France and Europe and this, not only on the B.1.117 (UK) but also aginst the B.1.351 (south-african). therefore they differ from the two cockails of MAbs with ATU in France from Eli-Lilly (no activity against B.1.351) or RGNR (partial activity against B.1.351) [15].

In pre-clinical experiments, AZD7442 has been shown to block the binding of the SARS-CoV-2 virus to host cells and protect against infection in cell and animal disease models [16].

AZD7442 is advancing into two Phase III clinical trials: the PROVENT trial has evaluated the safety and efficacy of AZD7442 to prevent infection for up to 12 months. PROVENT is a phase III, randomized, double-blind, placebo-controlled, multi-centre trial assessing the safety and efficacy of a single 300mg dose of AZD7442 compared to placebo for the prevention of COVID-19. The trial was conducted in 87 sites in the US, UK, Spain, France and Belgium. 5,197 participants were randomised in a 2:1 ratio to receive a single intramuscular (IM) dose of either 300mg of AZD7442 (n = 3460) or saline placebo (n = 1,737), administered in two separate, sequential IM injections. The primary efficacy endpoint was the first case of any SARS-CoV-2 RT-PCR positive symptomatic illness occurring post dose prior to day 183. Participants were adults 18 years-old and over who would benefit from prevention with the long-acting antibody AZD7442,

[5] https://investor.regeneron.com/news-releases/news-release-details/regn-cov2-independent-data-monitoring-committee-recommends/
[6] https://investor.regeneron.com/news-releases/news-release-details/regeneron-announces-encouraging-initial-data-covid-19-antibody

defined as having increased risk for inadequate response to active immunisation (predicted poor responders to vaccines or intolerant of vaccine) or having increased risk for SARS-CoV-2 infection, including those whose locations or circumstances put them at appreciable risk of exposure to the SARS-CoV-2 virus. Participants at the time of screening were unvaccinated and had a negative point-of-care SARS-CoV-2 serology test. Preliminary results have been released by the AZ firm on the 20th of August 2021, stating that AZD7442 achieved a statistically significant reduction in the incidence of symptomatic COVID-19, the trial's primary endpoint. AZD7442 reduced the risk of developing symptomatic COVID-19 by 77% (95% confidence interval (CI): 46, 90), compared to placebo. The trial accrued 25 cases of symptomatic COVID-19 at the primary analysis. There were no cases of severe COVID-19 or COVID-19-related deaths in those treated with AZD7442. In the placebo arm, there were three cases of severe COVID-19, which included two deaths. We are currently waiting for the peer-reviewed publication.

The second trial STORM CHASER has evaluated post-exposure prophylaxis and pre-emptive treatment in approximately 1,100 participants but did not meet primary endpoint. Indeed, the primary endpoint was the first case of any SARS-CoV-2 RT-PCR positive symptomatic illness occurring post dose prior to day 183, with a follow-up of 15 months. Nevertheless, there was a 73% reduction of symptomatic cases in the AZ-7442 treatment arm compared to placebo in those PCR negative at baseline.

In addition, other drugs might soon emerge that also require evaluation. This review is conducted regularly as more data become available. The intention is to assess the evidence available for these candidates regarding safety and efficacy.

#### Corticosteroids

Corticosteroids were added to the SoC on October 2, 2020 based on emerging literature.

The Recovery clinical trial randomised 2104 patients to receive dexamethasone and 4321 to receive usual care [17]. Dexamethasone was administered orally or intravenously at a dose of 6 mg once daily for up to 10 days (or until hospital discharge if sooner). In the dexamethasone group, the incidence of death was lower than that in the usual care group among patients receiving invasive mechanical ventilation (29.3% vs. 41.4%; rate ratio, 0.64; 95% CI, 0.51 to 0.81) and among those receiving oxygen without invasive mechanical ventilation (23.3% vs. 26.2%; rate ratio, 0.82; 95% CI, 0.72 to 0.94), but not among those who were receiving no respiratory support at randomization (17.8% vs. 14.0%; rate ratio, 1.19; 95% CI, 0.91 to 1.55). The use of dexamethasone resulted in lower 28-day mortality

CONFIDENTIAL

among those who were receiving either invasive mechanical ventilation or oxygen alone at randomization but not among those receiving no respiratory support.

Corticosteroids have previously been evaluated in patients with Acute Respiratory Distress Syndrome (ARDS), in a non-COVID context. In a multicentre, randomised controlled trial, 139 moderate-to-severe ARDS patients (Berlin criteria) were randomly assigned 139 to a dexamethasone group and 138 to a control group. Patients in the dexamethasone group received an intravenous dose of 20 mg once daily from day 1 to day 5, which was reduced to 10 mg once daily from day 6 to day 10. At 60 days, 29 (21%) patients in the dexamethasone group and 50 (36%) patients in the control group had died (between-group difference −15·3% [–25·9 to −4·9]; p=0·0047) [18]. In COVID-19 patients with moderate to severe ARDS, the Codex trial, using the same schedule of corticotherapy, found an increase in ventilator free days, without any difference in mortaliy [19].

The main difference between these trials is the degree of severity of the patients. In the Recovery trial, only 15% of patients were receiving invasive ventilation, while all the patients in the ARDS trial were receiving invasive ventilation, with 85% of moderate ARDS and 15% of severe ARDS. We can hypothetize that the Recovery study paves the way for the corticotherapy schedule in COVID-19 for non-critical patients, within the first stage of respiratory failure. The question of the best corticotherapy schedule for the most severe patients with ARDS in COVID-19 is not totally answered.

A recent meta-analysis evaluated the administration of systemic corticosteroids to critically ill patients with COVID-19 using the data from 7 randomized trials (including Recovery and Codex). It included 678 patients treated with systemic corticosteroids (dexamethasone, hydrocortisone, or methylprednisolone) and 1025 patients receiving placebo or SoC. There were 222 deaths among the 678 patients randomized to corticosteroids and 425 deaths among the 1025 patients randomized to usual care or placebo (summary OR, 0.66 [95% CI, 0.53-0.82]; P < .001 based on a fixed-effect meta-analysis) [20]. The sub-groups analyses are in favor of the use of dexamethasone (Recovery schedule) or dexamethasone/methylprednisolone ("ARDS schedule"), but not in favor of hydrocortisone at "sepsis dose".

In light of this literature, and because the majority of patients included in the DisCoVeRy clinical trial are COVID-19 patients needing oxygen support, we decided to include corticosteroids in the standard of care (SoC) for those patients needing oxygen.

CONFIDENTIAL

The investigators are requested to use the dosing assessed in the Recovery trial: dexamethasone 6 mg daily for 10 days or until discharge, whichever is shorter. In case of a patient with ARDS in an ICU ward, investigators are free to change the schedule for an ARDS one (for instance dexamethasone 20 mg per day for 5 days, and then 10 mg per day for 5 days, as in the last ARDS trial, with early administration).

**Anticoagulants**

Anticoagulants were added to the SoC on October 2, 2020 based on emerging literature. The predominant coagulation abnormalities in patients with COVID-19 suggest a hypercoagulable state and an increased risk of venous thromboembolism and pulmonary embolism [21–23]. In a retrospective study of 449 patients with severe COVID-19, venous thromboembolism prophylaxis was associated with improved survival when compared with no prophylaxis, especially in patients with high D-dimer values [24]. In a retrospective series of 2773 individuals hospitalized with COVID-19, in whom 786 (28 percent) received systemic anticoagulation, anticoagulation was associated with improved in-hospital survival in intubated patients (71 percent, versus 37 percent for those who were not anticoagulated) [25].

According to numerous guidance by organizations and scientific societies, venous thromboembolism (VTE) prophylaxis should be prescribed in all hospitalized medical, surgical or ICU patients [26,27]. Full-dose anticoagulation should be undertaken for patients with proven venous thromboembolism including pulmonary embolism. The SoC will thus include venous thromboembolism prophylaxis and/or anticoagulant treatment according to the protocol applied in each center for dose and duration.

#### 2.3. Risk/Benefit Assessment

Despite great efforts to search for a highly effective anti-viral treatment against SARS-CoV2 infection in hospitalized COVID-19 patients, none have yet been identified. Given the unmet medical need for safe and effective therapeutics, the global clinical research task force must continue to carry out therapeutics' evaluations worldwide. This is the commitment of the DisCoVeRy clinical trial. The approval of a new drug product requires substantial evidence of effectiveness and a demonstration of safety for the drug's intended use. A benefit-risk assessment is indispensable, based on rigorous scientific standards to ensure that the product's benefits outweigh its risks for the intended population.

##### 2.3.1. Known Potential Risks

For each new therapeutic agent under investigation, findings from the preclinical and clinical studies will be briefly described in this section and the IB will be in the eTMF. The final decision to include or exclude breastfeeding or pregnant women and children depends on a risks and benefits assessment.

The potential risks of participating in this study are those associated with having blood drawn, the intravenous (IV) catherization, possible reactions to AZD7442, and breach of confidentiality. Drawing blood may cause transient discomfort and fainting. Fainting is usually transient and managed by having the subject lie down and elevate his/her legs. Bruising at the blood collection sites may occur but can be prevented or lessened by applying pressure to the blood draw site for a few minutes after the blood is taken. Intravenous catheterization may cause insertion site pain, phlebitis, haematoma formation, and infusate extravasation; less frequent but significant complications include bloodstream and local infections. Working in an aseptic (sterile) manner will decrease the risk of infection at the site where blood is to be drawn or at catheter site.

##### 2.3.2. Risks to Privacy

Subjects will be asked to provide personal health information (PHI). All attempts will be made to keep this PHI confidential within the limits of the law. However, there is a chance that unauthorized persons will see the subject's PHI. All study records will be kept in a locked file cabinet or maintained in a locked room at the participating clinical site. Electronic files will be password protected.

Only people who are involved in the conduct, oversight, monitoring, or auditing of this study will be allowed access to the PHI that is collected.

Any publications from this study will not use information that will identify subjects by name. Organizations that may inspect and/or copy research records maintained at the participating site for quality assurance and data analysis include groups such as the IRB/EC, Sponsor and the pertinent regulatory authorities.

##### 2.3.3. Known Potential Benefits

The candidate therapeutic(s) being evaluated may or may not improve clinical outcome of an individual adult subject with COVID-19 who participates in this study. However, there is potential benefit to society from their participation in this study resulting from insights gained about the therapeutic agents under study as well as the natural history of the disease. While there may not be benefits for an individual subject, there may be benefits to society if a safe, efficacious therapeutic agent can be identified during this global COVID-19 outbreak.

CONFIDENTIAL

##### 2.3.4. Assessment of Potential Risks and Mitigation Strategies

**Potential and identified risks in Humans**

AZD7442 is a combination of 2 human mAbs, with non-overlapping epitopes directed against the Receptor Binding Domain (RBD) of the SARS-CoV-2 spike protein for neutralization of the virus. Neither mAb have any human target. Neutralizing mAbs may or may not improve clinical outcome of an individual adult subject with COVID-19. However, there is a potential benefit to society from their participation in this study resulting from insights gained about the therapeutic agent under study as well as the natural history of the disease. While there may not be benefits for an individual subject, there may be benefits to society if a safe, effective therapeutic agent can be identified during this global COVID-19 outbreak. The AZD7442 has already been studied in phase I and II trials (see Investigator Brochure); it appears to be well-tolerated.

However, there are also some possible adverse events or risks related to treatment with AZD7442 mAbs:

1. Potential risks are associated with the administration of any immunoglobulin, including polyclonal immunoglobulin preparations and mAbs. The potential risks include, but are not limited to, injection site reactions, infusion-related reactions, anaphylaxis and other serious hypersensitivity reactions including immune complex disease, and antibody-dependent enhancement of disease (ADE). Injection site reactions may manifest as local inflammation, redness, itching, pain, bruising, infection, or excessive bleeding at the site of injection. These reactions should be managed according to standard clinical practice. Other reactions include infusion-related reaction (excluding hypersensitivity and anaphylaxis) occurring during infusion of AZD7442/placebo or within 30 minutes to 2 hours after the initiation of first AZD7442/placebo infusion (in rare cases up to 24h). These reactions most often are mild in intensity but they can be severe and there have been reports of fatal events, although no severe or fatal reactions have been reported for AZD7442 to date. In a phase 1 trial no infusion related reactions were reported for AZD7442/Placebo. . Anaphylaxis, although rare usually occurs after subsequent exposure to an antigen, and is most commonly accompanied by severe systemic skin and/or mucosal reactions and difficulty breathing
   ADE is a theoretical risk. ADE is an increased binding efficiency of virus-antibody complexes to Fc receptor bearing cells, which triggers virus entry. The mAbs in AZD7442 have been designed to prevent binding to cellular Fc receptors, so the risk of ADE occurring via this mechanism should range from very low to none.
2. There is also a theoretical risk for causing an antibody-dependent enhancement of SARS-CoV2 infection (antigen-antibody complex), or in other

CONFIDENTIAL

words for amplifying the infection or triggering harmful immunopathology. However, the Fc fragment of the monoclonal antibodies have been manipulated to ablate the effector function, thus reducing this theoretical risk.

3. Furthermore, different variants of SARS-CoV2 are currently emerging in different world spots leading to a possibility that AZD7442 may prove to be ineffective against one or several of these mutant strains. Mutations in SARS-CoV-2 may reduce mAbs efficacy by decreasing the binding affinity of the antigen-antibody complex. Nevertheless, the bispecific mAb combination such as AZD7442 based on a mAb cocktail including 2 antibodies derived from COVID-19 convalescent donors binding two different sites has a better chance to suppress viral escape than a single mAb. Finally, AZD7442 has a very long half-life, thus possibly offering a long protection against SARS-CoV2 infection, but the long half-life could also theoretically play a role in the emergence of resistant mutant strains through sustained exposure to sub-optimal concentrations.

While multiple SARS-CoV-2 variants are circulating globally, rapidly spreading variants have been reported, initially emerging in the United Kingdom and South Africa. The B.1.1.7 lineage (aka 201/501Y.V1, VOC 202012/01) of SARS-CoV-2 was first identified in cases from Kent, United Kingdom in September 2020. The B.1.351 lineage (aka 20H.501Y.V2) of SARS-CoV-2 was first identified in cases from Nelson Mandela Bay, South Africa in October 2020. This variant emerged independently of B.1.1.7, yet while it shares some of the same substitutions, has been defined by additional spike protein substitutions of concern in the receptor binding domain. Preliminary in vitro studies, not yet formally validated to study efficacy of mABs against variants, have shown that AZD7442 and the individual mAbs that comprise AZD7442 (AZD8895 and AZD1061) neutralize B.1.1.7 and B.1.351 emergent strains (https://doi.org/10.1101/2021.01.25.428137)
Please see IB (in eTMF) for additional information.

**Mitigation strategies**

- Administration of the mABs only in a setting where respiratory and circulatory support is available should an adverse reaction to dosing occur.
- Education of site staff specifically related to administration of mAbs.
- Regular laboratory assessments.
- Exclusion criteria have been adapted to take into account these risks.

Taking into account the measures taken to minimize risk of participants in this study, the potential risks identified in association with AZD7442 are justified by the anticipated benefits that may be afforded to participants at risk of COVID-19.

The constantly evolving landscape of mutations of key proteins of SARS-CoV-2 must strengthen our vigilance regarding the full maintenance of activity of the AZD7442 on SARS-CoV-2 over the course of the trial and the potential therapeutic selective pressure resulting from its use. Consequently, the monitoring of strain variants, which mutation hotspot patterns have been and will be identified in different geographic areas is of critical importance during the course of the DisCoVeRy trial. Very close virological follow-up of patients will be carried out through a regular genomic screening of the strains as stated in the 1.3 section and the table 1 in the section 1.9 (schedule of assessments).

A close and individual virological follow-up is planned in case of suspected virological escape to treatment between D0 and D8. In case of absence of viral decay between D0 and D8 or a PCR Ct value less than 31 at D8, the nasopharyngeal sample assessed at D8 will be sequenced in the country of inclusion  or sent to the National Reference Center for Respiratory Viruses in Lyon (France). All sequences obtained will be centralized at the National Reference Center for Respiratory Viruses in Lyon to allow a global analysis and thus monitor the emergence of an escape variant.

### 3. Study Objectives

#### 3.1. Primary Objective

The primary objective of the study is to evaluate the clinical efficacy of the investigational product compared to the placebo arm in patients hospitalized with COVID-19 using a 7-point ordinal scale assessing clinical status at Day 15.

#### 3.2. Secondary Objectives

##### 3.2.1. Key secondary objectives

Evaluate the efficacy of the investigational product on sustained recovery from index COVID-19 infection.

##### 3.2.2. Additional secondary objectives

**Clinical efficacy**

Evaluate clinical efficacy of different investigational therapeutics as compared to the control arm as assessed by:

Ordinal scale

- Subject clinical status on an ordinal scale on Day 29, 90, 180 and 365

National Early Warning Score 2 (NEWS-2)

- Change from baseline to Days 3, 8, 15 (while hospitalized) and 29 in NEWS-2

CONFIDENTI

Oxygenation

- Oxygenation-free days during the first 28 days (to Day 29)
- Incidence and duration of new oxygen use, non-invasive ventilation or high flow oxygen devices during the first 28 days (to Day 29)

Mechanical Ventilation

- Ventilator-free days during the first 28 days (to Day 29).
- Incidence and duration of invasive mechanical ventilation use during the first 28 days (to Day 29)
- Need for mechanical ventilation or death by Day 15

Hospitalization

- Time to hospital discharge from randomization

Mortality

- In-hospital mortality
- 29-day mortality
- 90-day mortality
- 180-day mortality
- 365-day mortality
- 456-day mortality

Long term health status

- Any hospitalization
- Any confirmed re-infection with SARS-CoV-2

**Safety**

Evaluate the safety of different investigational therapeutics through 456 days of follow-up as compared to the placebo arm as assessed by:

- Cumulative incidence of serious adverse events (SAEs)
- Cumulative incidence of Grade 3 and 4 adverse events (AEs)
- Cumulative incidence of Grade 1-2 hypersensitivity-related and infusion related adverse events (AEs) until D29 visit
- Cumulative incidence of AEs of Special Interest
- Discontinuation of investigational therapeutics (for any reason)

#### 3.3. Exploratory Objective

Evaluate the virologic efficacy of different investigational therapeutics as compared to the control arm as assessed by:

CONFIDENTIAL

- Percent of subjects with detectable SARS-CoV-2 in NP or lower respiratory tract sample at baseline (Day 1 pre-treatment) and at Days 3, 8, 15 (while hospitalized) and 29
- Normalized quantitative SARS-CoV-2 viral load in NP or lower respiratory tract sample at baseline (Day 1 pre-treatment) and at Days 3, 8, 15 (while hospitalized) and 29
- Normalized quantitative SARS-CoV-2 viral load in blood at baseline (Day 1 pre-treatment) and at Days 3 and 8
- Detection of variants of SARS-CoV-2 in NP or lower respiratory tract sample at baseline (Day 1 pre-treatment) and at Days 3, 8, 15 (while hospitalized) and 29, and in case of transfer into ICU

Evaluate the pharmacokinetics of AZD7442

Evaluate the development of anti-drug antibodies (ADA) against the monoclonal antibodies

Identify host genetic variants having an impact:
- in the development of severe clinical disease in individuals infected by SARS-CoV-2
- in the response in terms of safety and efficacy to investigational antiviral drugs

Evaluate the immune-inflammatory response during treatment of COVID-19

Evaluate the kinetics and functional capacities of patients anti-SARS-CoV-2 antibodies to evaluate the added value of the therapeutic antibody compared to the host's immunity

### 4. Study Endpoints

#### 4.1. Primary Endpoint

Clinical status of subject on Day 15 (on a 7-point ordinal scale):

1. Not hospitalized, no limitations on activities.
2. Not hospitalized, limitation on activities.
3. Hospitalized, not requiring supplemental oxygen.
4. Hospitalized, requiring supplemental oxygen.
5. Hospitalized, on non-invasive ventilation or high flow oxygen devices.
6. Hospitalized, on invasive mechanical ventilation or ECMO.
7. Death.

#### 4.2. Secondary Endpoints

CONFIDENTIAL

##### 4.2.1. Key secondary endpoint

Time from randomization to sustained recovery, defined as being discharged from the index hospitalization, followed by being alive and home for 14 consecutive days prior to Day 90.

##### 4.2.2. Additional secondary objectives

**For efficacy assessment**

- Status on an ordinal scale assessed at Day 29, 90, 180 and 365
- NEWS-2 at baseline (Day 1 pre-treatment), at Days 3 and 8, 15 (if patient is still hospitalized), and at Days 29at baseline and at Days 3, 8, 15 (while hospitalized) and at Day 29
- Duration of supplemental oxygen (if applicable)
- Duration of mechanical ventilation (if applicable)
- Date of discharge from hospital
- Date and cause of death (if applicable)
- Mechanical ventilation or death between baseline and Day 15
- Occurrence of new hospitalization between discharge from index hospitalization and Days 90, 180 and 365
- Occurrence of confirmed re-infection with SARS-CoV-2 between discharge and Days 90, 180 and 365

**For safety assessment**

- SAEs
- Grade 3 and 4 adverse events
- Grade 1-2 hypersensitivity-related and infusion related adverse events (AEs) until D29 visit
- AEs of Special Interest
- Discontinuation of investigational therapeutics (for any reason)

##### 4.3. Exploratory Endpoint

- Qualitative (Ct Value) and quantitative PCR (normalized viral load) for SARS-CoV-2 in NP swab or lower respiratory tract sample at baseline (Day 1 pre-treatment), at Days 3 and 8, 15 (if patient is still hospitalized), and at Days 29
- Qualitative (Ct value) and quantitative PCR (viral load) for SARS-CoV-2 in blood at baseline (Day 1 pre-treatment) and at Days 3, and 8 (if patient is still hospitalized)
- Whole Genome Sequencing in NP or lower respiratory tract sample at baseline (Day 1 pre-treatment) and at Days 3, 8, 15 (while hospitalized) and 29, and in case of transfer into ICU

CONFIDENTIAL

- Serum AZD7442 concentration on Day 1 before and after end of infusion (peak), on Days 3 and 8, 15 (if patient is still hospitalized), and at Days 29, 90, and in a subset of 25% of enrolled patients, at Days 180 and 365
- Anti-AZD7442 antibodies Day 1 (pre-treatment), 15 (if patient is still hospitalized), 29, 90, and in a subset of 25% of enrolled patients, at Days 180 and 365
- Whole genome sequencing (WGS) of the subjects
- Blood levels of immunothrombosis markers, IFNs (and anti-IFNs antibodies), cytokines and complement activation products at baseline (Day 1 pre-treatment) and at Days 3, 8, 15 (while hospitalized), 29, 90, and in a subset of 25% of enrolled patients, at Days 180 and 365
- Blood levels and functional capacities of patients anti-SARS-CoV-2 immunoglobulins at baseline (Day 1 pre-treatment), and at Days 3 and, 8, 15 (while if patient is still hospitalized), and at Days 29, 90, and in a subset of 25% of enrolled patients, at Days 180 and 365

### 5. Study Design

#### 5.1. Overall Design

This study is an adaptive, randomized, blinded multicentre/country trial to evaluate the safety and efficacy of therapeutic agents in hospitalized adult patients diagnosed with COVID-19.

Several therapeutic agents have already been evaluated, and discontinued, as presented in the Introduction.
In the present version of the protocol, the study is randomizing adult hospitalized, COVID-19 patients not requiring invasive mechanical ventilation, to one of 2 arms (1:1 ratio): SoC + placebo or SoC + AZD7442.
Other arms can be added as evidence emerges from other candidate therapeutics.
In addition, since October 2, 2020, corticosteroids for patients needing oxygen therapy and anticoagulants have been part of the SoC.

An independent data and safety monitoring board (DSMB) will actively monitor trial data to make recommendations about early study cessation for safety or changes in study arms. If one therapy shows unequivocal efficacy compared with placebo/SoC, then it may be added to the control arm for comparison(s) with new experimental treatment(s). In the current version of the protocol, there will be no pre-planned interim efficacy or futility analysis conducted, thus the efficacy results will not be available until the primary analysis.

Randomization will be stratified by:

CONFIDENTIAL

- Region (according to the administrative definition in each country)
- Antigenic status (positive or negative), obtained from the result of a rapid antigen test on nasopharyngeal swab performed at enrolment
- Vaccination initiation (yes or no): vaccination is initiated if at least one injection of any vaccine againt SARS-CoV- 2 was received prior to enrolment, whatever the delay.

The primary analyses will be conducted on patients with antigen-positive results. A positive antigenic test is evidence of high viral shedding consistent with a recently started or uncontrolled infection. Overall, the number of antigen-negative patients will be at most 30% of all included subjects. The number of patients with vaccination (partly or fully) will be limited to 20% of all participants, split evenly between antigen positive and antigen negative patients (i.e. vaccinated patients can make up at most 20% of antigene positive patients and 20% of antigene negative patients). Sensitivity analyses will be performed in all patients, stratified by antigenic status and vaccination initiation.

Notwithstanding that the number of strata should be kept to the minimum and the ones selected of major importance, the time of onset of symptoms should be considered for stratification or at least, collection of these data is essential for proper subgroup analyses, and in consideration of the pilot data it might be one key factor to consider as to whether there should be stratification on this variable.

All subjects will undergo a series of efficacy and safety assessments, including laboratory assays. Subjects will be assessed at baseline, and at Days 3, 8 and 15 while hospitalized. Patients will be contacted by phone at Day 15 for evaluation of the Primary Endpoint if they have been discharged prior to Day 15, and 14 days following hospital discharge for efficacy assessment. Further follow-up assessments will be organized at Days 29, 90, 180, 365 and 456. If discharged from the hospital, days 29 and 90 assessments will be organized as outpatients' consultations for all. For Days 180 and 365 assessments, a subset of 25% of patients enrolled in centers with available resources and selected at Day 90 will be evaluated during a medical consultation, while the other will be contacted by phone. For Day 456, all patients will be contacted by phone.

Nasopharyngeal swabs (NP) or lower respiratory tract samples will be obtained at baseline (Day 1 pre-treatment) and at Days 3, 8, 15 (while hospitalized) and 29 (while hospitalized or, if discharged from the hospital, in the outpatient setting).
Blood samples will be obtained at baseline (Day 1 pre-treatment) and at Days 3, 8, 15 (while hospitalized), at Days 29 and 90, and at Days 180 and 365 (for the subset of patients evaluated during a medical consultation at these times).

CONFIDENTIAL

Thoracic computed tomography (CT)-scan will be obtained at baseline, depending on the centre's imagery capacities.

##### 5.2. Scientific Rationale for Study Design

This study utilizes an adaptive design that maximizes efficiency in identifying a safe and efficacious therapeutic agent for COVID-19 during the current outbreak. Some investigational products may be in limited supply and this study design enables continuation of the study even if a product becomes unavailable. In addition, the adaptive design allows for the evaluation of new therapeutic agents as they are identified. As the study is a multicentre, multinational randomized controlled study, it will be possible to acquire rigorous data about the safety and efficacy of investigational therapeutic agents for COVID-19 that will lead to generalizable evidence.

Randomization is essential for establishing efficacy of these new therapeutic agents. Also, collecting clinical and virologic data on enrolled patients using a standardized timeline and collection instruments should provide valuable information about the clinical course of and morbidities associated with severe COVID-19 in a diverse group of hospitalized adult patients.

It was initially decided to conduct an open trial as preparing placebo for all concomitant treatment groups was not feasible to start rapidly. In addition, the placebo design was complex to handle in a study with several active treatment groups with different routes of administration and potential additional arms to be added.

The current protocol will implement a blinding and placebo to match the single treatment arm with active AZD7442. Please see section 7.5 for details on preparation of active drug and placebo.

#### 6. Study Population

Male and non-pregnant female adults ≥18 years of age hospitalized with COVID-19 who meet all eligibility criteria will be enrolled.

Children and pregnant women will be excluded from the study.

The estimated time from screening to end of study for an individual subject is approximately 456 days.

##### 6.1. Inclusion Criteria

CONFIDENTIAL

In order to be eligible to participate in this study, a patient must meet all of the following inclusion criteria:

1. Adult ≥18 years of age at the time of enrolment
2. Hospitalized patients with any of the following criteria:
    a. the presence of pulmonary rales/crackles on clinical exam OR
    b. SpO2 ≤ 94% on room air OR
    c. requirement of supplementary oxygen including high flow oxygen devices or non-invasive ventilation
3. A time between onset of symptoms and randomization of **less than 11 days**
4. A positive SARS-CoV-2 PCR performed on a NP swab **within the 5 days** preceding randomization
5. The result of a rapid antigen test performed on a NP swab **within the 6 hours** preceding randomization
6. Contraceptive use by men or women.
    a. Male participants: Contraception for male participants is required; to avoid the transfer of any fluids, all male participants must use a condom from Day 1 and agree to continue for 90 days following administration of IMP.
    b. Female participants: Women of child-bearing potential must agree to use contraception for 365 days following administration of IMP. Acceptable birth control methods are listed in section 8.5.

#### 6.2. Exclusion Criteria

An individual who meets any of the following criteria will be excluded from participation in this study:

1. Refusal to participate expressed by patient or legally authorized representative
2. Need for invasive mechanical ventilation and/or ECMO at the time of enrolment
3. Spontaneous blood ALT/AST levels > 5 times the upper limit of normal
4. Glomerular filtration rate (GFR) < 15 mL/min or requiring maintenance dialysis
5. Pregnancy or breast-feeding
6. Anticipated transfer to another hospital, which is not a study site within 72 hours following randomization
7. Known history of allergy or reaction to any component of the study drug formulation.
8. Previous hypersensitivity, infusion-related reaction, or severe adverse reaction following administration of monoclonal or polyclonal antibodies.
9. Any prior receipt of investigational or licensed mAb/biologic indicated for the prevention of SARS-CoV-2 infection or COVID-19, and for those not

CONFIDENTIAL

vaccinated, expected receipt of vaccine in the 30 days following hospital discharge, according to current recommendation in each country.

10. Any medical condition which, in the judgment of the investigator, could interfere with the interpretation of the trial results or that preludes to protocol adherence.

#### 6.3. Exclusion of Specific Populations

Children will be excluded from the study under this version of the protocol.

As the effects on the foetus are not fully known from nonclinical reproductive toxicity studies, pregnant women will not be eligible for the study.

#### 6.4. Inclusion of Vulnerable Participants

In the DisCoVeRy trial, vulnerable participants are defined as patients who may be under legal protection.

A specific procedure has been written in order to include adult patients under legal protection: the legal representative must sign the informed consent form.

#### 6.5. Lifestyle Considerations

During this study, women of childbearing potential are asked to avoid getting pregnant during the study from Day 1 up to Day 365.
Men are asked to agree using contraception for 90 days following randomization.

#### 6.6. Screen Failures

After the screening evaluations have been completed, the investigator or his/her designee is to review the inclusion/exclusion criteria and determine the subject's eligibility for the study.

Only the reason for ineligibility will be collected on screen failures. Subjects who are found to be ineligible will be told the reason for ineligibility.

Individuals who do not meet the criteria for participation in this study (screen failures) because of an abnormal laboratory finding may be rescreened once the abnormality is resolved.

#### 6.7. Strategies for Recruitment and Retention

##### 6.7.1. Recruitment

CONFIDENTIAL

It is anticipated that patients with COVID-19 will present to participating hospitals, and that no other efforts to recruit potential subjects are needed. Recruitment efforts may also include dissemination of information about this study to other medical professionals / hospitals.

Several European countries are enrolling in DisCoVeRy and other European countries will soon join. The recent launch of the 5-year multi-national 'European Research and Preparedness Network for Pandemics and Emerging Infectious Diseases' — EU-RESPONSE project coordinated by Inserm and funded by the European Commision will allow the rapid European expansion and further evolution of the DisCoVeRy study. The project will also help build a new multinational European Adaptive Platform Trial for emerging infectious diseases in general [7].
In France, the RENARCI network is involved in the recruitment of participating sites. In Europe, the recrtuiment of sites will be stimulated using the ECRIN network.

Information regarding this trial may be provided to potential subjects who have previously participated in other trials conducted at the sites and to medical care providers who have cases of COVID-19 admitted to their hospital or in the referral area. Other forms and/or mechanisms of recruitment may also be used. The IRB/EC will approve the recruitment process and all materials prior to use.

In participating centers, all consecutive hospitalized patients that are confirmed to have SARS-CoV-2 will be assessed for eligibility. Screening will begin with a brief discussion with study staff. Some will be excluded based on demographic data and medical history i.e. pregnancy, < 18 years of age, severe renal failure, etc. Subject Inclusion and Exclusion Criteria must be confirmed by a study clinician named on the delegation log.

Information about the study will be presented to potential subjects (or legal representative) and questions will be asked to determine potential eligibility. Screening procedures can begin only after informed consent is obtained.

##### 6.7.2. Retention

Participating subjects will be reminded of subsequent visits.

##### 6.7.3. Compensation Plan for Subjects

An indemnity of 70 euros in compensation for the constraints suffered (including transport costs) is provided for each of the following visits:

[7] https://ec.europa.eu/info/news/commission-supports-international-clinical-research-network-treat-covid-19-2020-sep-18_en

CONFIDENTIAL

- D90 (all patient) performed in outpatient setting,
- D180 and D365 (subset of 25% of enroll patients) performed in outpatient setting.

##### 6.7.4. Costs

There is no cost to subjects for the research tests, procedures/evaluations and study product while taking part in this study.

### 7. Study Product

#### 7.1. Investigational Therapeutic

##### 7.1.1. Study Product Description

AZD7442, a cocktail of AZD8895 and AZD1061, human IgG1κ mAbs directed against the RBD of the SARS-CoV-2 spike protein. The antibodies were derived from mAbs COV2-2196 and COV2-2130, respectively, that were cloned from B cells isolated from COVID-19 convalescent patients. AZD8895 has an overall molecular weight of approximately 149 kDa, and AZD1061 has an overall molecular weight of approximately 151 kDa. Both mAbs are expressed in Chinese hamster ovary cells, and the manufacture of clinical supplies is conducted in accordance with current Good Manufacturing Practices. AZD7442 will be supplied as separate vials of AZD8895 and AZD1061 containing 150 mg colorless to slightly yellow, clear to opalescent solutions for injection. The solutions contain 100 mg/mL of active ingredient (AZD8895 or AZD1061) in 20 mM Lhistidine/L-histidine hydrochloride, 240 mM sucrose, and 0.04% (w/v) polysorbate 80, at pH 6.0. The label-claim volume is 1.5 mL.

The placebo will be a 0.9% (w/v) NaCl solution for infusion also called saline. The placebo will be supplied as a single 10-mL, clear and colorless vial.

AZD7442/placebo drug product will be packaged as a kit per patient which will contain the full treatment course for the patient.

**Table 2: Study medication overview.**

| **Intervention Name** | **AZD7442** | **Placebo** |
|---|---|---|
| **Dose Formulation** | AZD8895: single-use vial (100 mg/mL)<br>AZD1061: single-use vial (100 mg/mL) | 0.9% (w/v) saline |

CONFIDENTIAL

| | | |
|---|---|---|
| **Dosage Level(s) (mg)** | 600 mg dose (300 mg AZD8895 and 300 mg AZD1061) | 0.9% (w/v) saline |
| **Dose Volume (mL)** | Refer to pharmacy procedures | Refer to pharmacy procedures |
| **Route of administration** | IV infusion | IV infusion |
| **Use** | Experimental | Placebo-comparator |
| **IMP and NIMP** | IMP | IMP |
| **Sourcing** | AstraZeneca | Sponsor |
| **Packaging and Labeling** | Study Intervention will be provided as four single-use 10R vials packaged in one carton per patient, and labeled appropriately | Study Intervention will be provided as one single-use 10 mL vial, packaged in one carton per patient, and labeled appropriately |

##### 7.1.2. Dosing and Administration

The AZD7442/placebo will be administered within 24 hours following randomization. Participants will receive via IV infusion from a single IV bag a total dose of 600 mg AZD7442 *via* co-administration (both mAbs mixed in a single infusion) at a maximum infusion rate of 50 mg/minute or placebo. Time of study drug administration to the subject will be recorded on the appropriate data collection form (eCRF).

A **blinded** trained professional will administer the entire contents of the IV bag using IV administration sets containing low protein binding 0.2 µm or 0.22 µm polyethersulfone (PES) filters. The target infusion time is 30 minutes, and the target infusion rate is 20 mg/minute. The catheter will be flushed with 5 mL normal saline at the end of the infusion. The IV line will be flushed according to local practices to ensure the full dose is administered.
All participants must be under nurse or doctor surveillance during the infusion and should be monitored closely for 2 hours after the infusion, as there is a risk of infusion reaction and hypersensitivity (including anaphylaxis) with any biological agent. Though less frequent, infusion-related reactions can also occur later on within the first 24 hours from the start of infusion.

**Precautionary medications**

The clinical site should have necessary equipment and medications for the management of any infusion reaction.

Premedication for infusions is not planned.

CONFIDENTIAL

The investigators and sponsor may decide to recommend premedication, if the frequency of infusion reactions among participants warrants it. If minor infusion reactions are observed, administration of acetaminophen, 500 mg to 1000 mg, antihistamines and/or other appropriately indicated medications may be given prior to the start of infusions for subsequent participants. The decision to implement premedication for infusions in subsequent participants will be made by the investigator and sponsor and recorded in the study documentation. Any premedication given will be documented as a concomitant therapy.

##### Symptoms and Signs

Symptoms and signs that may occur as part of an infusion reaction include, but are not limited to, fever, chills, nausea, headache, bronchospasm, hypotension, angioedema, throat irritation, rash including urticaria, pruritus, myalgia, and dizziness.

Infusion-related reactions' severity will be assessed and reported using the Division of AIDS (DAIDS) Table for Grading the Severity of Adult and Pediatric Adverse Events, Corrected version 2.1 (July 2017) [NIAID Division of AIDS. *Division of AIDS (DAIDS) Table for Grading the Severity of Adult and Pediatric Adverse Events Corrected Version 2.1, July 2017*. 2017] [8].

##### Site Needs

The clinical site should have necessary equipment, medications, adequately qualified and experienced staff with appropriate medical cover for the management of any infusion reaction, which may include, but is not limited to, oxygen, IV fluid, epinephrine (adrenaline), acetaminophen (paracetamol) and antihistamine.

The pharmacy will be provided with labels to be placed on the IV bag before dispensing (refer to the pharmacy procedure).

The sponsor provides 0.9% (w/v) saline and IV bags, similarly shrouded.

##### Management of Infusion Reactions including Discontinuation

Investigators will use their clinical judgement and standard of care to evaluate and manage all infusion reactions. If an infusion reaction occurs, then supportive care should be used in accordance with the signs and symptoms. If a severe and potentially life-threatening infusion reaction occurs with AZD7442 or placebo, its use should be permanently or temporarily discontinued. For dose discontinuation,

[8] https://rsc.niaid.nih.gov/clinical-research-sites/grading-severity-adult-pediatric-adverse-events-corrected-version-two-one

CONFIDENTIAL

refer to sections 7.2.2 Dose Discontinuation and 9.1.4. Discontinuation of Study Drug.

If a participant is not infused with AZD7442/placebo or the complete infusion is not given, all follow-up procedures and reporting's outlined in the protocol, should be adhered to as indicated.

#### 7.2. Justification for Dose

Human efficacious doses for AZD7442 were evaluated using in vitro potency data (virus neutralizing activity of AZD7442 against SARS-CoV-2) and PK data. In addition, a viral dynamic model was developed to understand the pharmacodynamic effects of AZD7442 inhibiting a SARS-CoV-2 infection and the resulting immune response. In the viral dynamic model the AZD7442 partition ratio between lung epithelial lining fluid-to-serum is assumed as 1% and the potency (IC80; inhibiting SARS-CoV-2 by 80%) as 40 ng/mL. With the dose of 600 mg IV predicted to result in exposures in the epithelial lining fluid that exceed the IC80, this dose when administered before the time of peak viral load (~ 7 days after day of infection) is expected to result in reduction of the peak viral load and earlier eradication of the viral load. When administered after the peak viral load has been reached, AZD7442 is still expected to result in earlier viral load eradication compared to when drug is not present.

IV was selected over IM as the administration route for this treatment study because peak concentrations are achieved immediately after IV administration while it takes > 7 days after IM injection to achieve peak concentrations and early treatment is critical for this patient population. Also, the peak concentrations following IV infusion are about three times higher than following IM injection at the same dose level and higher Cmax may be correlated with better efficacy.

##### 7.2.1. Dose Escalation

Not Applicable

##### 7.2.2. Dose Discontinuation

For patients treated with AZD7442/placebo, in case of any suspected allergic reaction including vasogenic shock, drug infusion should be stopped immediately. The infusion can be resumed only after signs and symptoms recovery within 20mn. The infusion speed will be then decreased by half compared to initial speed rate at the time of the occurrence of infusions-related reactions.
Refer also to section 9.1.4. Discontinuation of Study Drug

CONFIDENTIAL

#### 7.3. Preparation/Handling/Storage/Accountability

##### 7.3.1. Acquisition

AZD7442/placebo will be shipped to the site from the Sponsor drug repository. The site will provide all other supplies.

Sponsor Procedures for ordering and accepting drug of AZD7442 (AZD8895 and AZD1061) and placebo, will be described in the pharmacy procedure.

##### 7.3.2. Accountability

The site PI is responsible for study product distribution and disposition and has ultimate responsibility for study product accountability. The site PI may delegate to the participating site's research pharmacist responsibility for study product accountability.
The participating site's research pharmacist will be responsible for maintaining complete records and documentation of study product receipt, accountability, reconciliation, inventory, dispensation, storage conditions, and final disposition of the study product(s); the pharmacy procedure details the requirements when performing accountability-related tasks at site.
All study product(s), whether administered or not, must be documented in the sponsor interactive web response system (IWRS). The sponsor's monitoring staff will verify IWRS records, temperatures logs, and AZD7442/placebo-preparation documents per the site monitoring plan.

Only participants enrolled in the study may receive AZD7442/placebo and only authorized site staff may supply or administer AZD7442/placebo.

##### 7.3.3. Destruction

After the study treatment period has ended or as appropriate over the course of the study after study product accountability has been performed, disposition of unused and used active vials should occur as noted:

Unused and Used active vials:

- Should be destroyed following applicable pharmacy procedures or by the site's selected destruction vendor. Following the pharmacy procedure and the site's procedure for the destruction of hazardous material or study product destruction policy/standard operating procedure (SOP) when destroying used and unused items.
- A certificate of destruction should be provided to the sponsor and retained in the Pharmacy Binder once completed as per pharmacy procedure.

CONFIDENTIAL

#### 7.4. Product Storage and Stability

The AZD7442/placebo must be stored in a secure facility, environmentally controlled, and monitored (manual or automated) facility in accordance with the labeled storage conditions, with access limited to the investigator and authorized site staff. All products must be stored at 2-8°C and used within the assigned expiry date on the label. The investigator or designee must confirm appropriate temperature conditions have been maintained during transit and on-site storage for all AZD7442/placebo received and any discrepancies are reported and resolved before use of the AZD7442/placebo.
Total time from needle puncture of the vial to the start of administration must not exceed:

- 24 hours at 2 °C to 8 °C
- 4 hours at room temperature.

If the final product is stored at both refrigerated and ambient temperatures, the total time must not exceed 24 hours.
For IV infusion, if the infusion solution was stored refrigerated, the infusion solutions must be allowed to equilibrate to room temperature prior to commencement of administration.
Each AZD8895, AZD1061 and placebo vial must be used only once to prepare a single dose.

#### 7.5. Preparation

A total of two vials of AZD8895 and two vials of AZD1061 are required for the dosing of the solution at 600 mg (see Table 2). Six mL from a single placebo vial is required for the dosing of the placebo to patient.
Sites should follow standard and local aseptic procedures, the clinical study protocol and pharmacy procedure for all activities. All dispensing activities should be documented according to local procedures and pharmacy procedure.

Use a single 0.9% (w/v) saline IV bag for each AZD8895/AZD1061 infusion or placebo infusion. The IV bag can be made of polyolefin (PO) or polyvinylchloride (PVC).

The target volume of IV bag is 100 mL. However, an IV bag volume of 250 mL is acceptable if a 100 mL IV bag is not available.

Preparation of the IV bag are performed by an **unblinded** qualified professional. This unblinded qualified professional must only prepare the drug and must not perform any other study procedures such as drug administration or entering data into study documents apart from the drug preparation source records, see

CONFIDENTIAL

pharmacy procedure. The administration will be performed by a **blinded** trained professional, see section 7.1.2 of the study protocol.

The AZD7442/placebo does not contain preservatives and any unused portion must be kept in a secured place until monitoring can be done (at the sponsor's request).
If there are any defects noted with the AZD7442/placebo before the infusion, the sponsor should be notified immediately.
If there are any defects noted with the AZD7442/placebo during or after infusion, the investigator and the sponsor should be notified immediately.

### 8. Measures to Minimize Bias: Randomization and Blinding

Since June 29, 2020, the study will randomize participants 1:1 to standard of care alone or with investigational product added. If additional arms are added to or dropped from the trial, randomization will proceed with an equal probability of assignment to each of the remaining arms.

The randomization procedure and link with DisCoVeRy e-CRF will be described in an SOP.

Randomization will be stratified by region (according to the administrative definition in each country), by the antigenic status (positive or negative), obtained from the result of a rapid antigen test on nasopharyngeal swab performed at enrolment, and by the vaccination initiation (yes or no). Vaccination is initiated if at least one injection of any vaccine against SARS-CoV-2 was received prior to enrolment, whatever the delay.

The randomization list will be computer-generated, by various size blocks. It will be directly implemented in the electronic Case Report Form.

#### 8.1. Study Intervention Compliance

Each dose of study product will be administered by a member of the clinical research team that is qualified and licensed to administer the study product. Administration and date, time, and location of injection will be entered into the case report form (CRF).

#### 8.2. Blinding

CONFIDENTIAL

Neither the participant nor any of the investigators or Sponsor staff who are involved in the treatment or clinical evaluation and monitoring of the participants will be aware of the study intervention received. Since AZD7442 and placebo are visually distinct prior to dose preparation (due to differences in container closure), AZD7442/placebo will be handled by an unblinded pharmacist (or representative) at the study site.

The patient, the site Investigator and study center staff will be blinded to the AZD7442/placebo allocation. The study center pharmacist and representative will be unblinded to the AZD7442/placebo allocation and will prepare AZD7442 or placebo for a patient as specified by the randomization scheme (and only the pharmacist and representative will know the randomization/treatment allocation detail). Lot numbers of AZD7442 dispensed will be recorded by the pharmacists and monitored by an unblinded monitor. Other study center staff and monitors will be not be given access to lot number information.

The randomization code should not be broken except in medical emergencies when the appropriate management of the participant requires knowledge of the treatment randomization. The investigator documents and reports the action to the Sponsor, without revealing the treatment given to the participant to the Sponsor staff.

The Sponsor retains the right to break the code for SAEs that are unexpected and are suspected to be causally related to the AZD7442/placebo and that potentially require expedited reporting to regulatory authorities. Randomization codes will not be broken for the planned analyses of data until all decisions on the evaluability of the data from each individual participant have been made and documented.

CONFIDENTIAL

#### 8.3. Procedures for Unblinding

The IWRS will be programmed with blind-breaking instructions. In case of an emergency, in which the knowledge of the specific blinded AZD7442/placebo will affect the immediate management of the participant's condition (eg. Antidote available), the investigator has the sole responsibility for determining if unblinding of a participants' AZD7442/placebo assignment is warranted. Participant safety must always be the first consideration in making such a determination. If a participant's AZD7442/placebo assignment is unblinded, the Sponsor must be notified within 24 hours after breaking the blind

#### 8.4. Concomitant Therapy

During the course of the DisCoVeRy trial (456 days of follow-up), included participants cannot be treated with other monoclonal or polyclonal antibodies which directly target SARS COV-2.

Whatever the randomization arm, patients can be vaccinated with any available SARS-CoV-2 vaccine after a delay post-infection that respects the recommandation within his/her country. In most European countries this delay is of 90 days post infection.

The standard of care will include corticosteroids for those patients requiring oxygen therapy as this drug was shown to reduce mortality of COVID-19 patients who needed oxygen support and venous thromboembolism prophylaxis or treatment according to guidelines for the management of COVID-19.

Participants cannot be included in another clinical trial investigating the efficacy of any treatment for COVID-19. However, the physician in charge of the patient is allowed to administer other drugs that are not antivirals including - but not limited - to tocilizumab, sarulimab, etc. These drugs can be used as part of the standard of care.

The list of medications received prior to enrolment will be assessed only from 7 days prior to enrolment to day 29 and will be detailed in the MOP. Post day 29, concomitant medications will be review based on potential relatedness to AEs.

#### 8.5. Acceptable birth method control

##### 8.5.1. Male participants

To avoid the transfer of any fluids, all male participants must use a condom from Day 1 and agree to continue through 90 days following administration of AZD7442/placebo.

##### 8.5.2. Female participants

Women not of childbearing potential are defined as women who are either permanently sterilized (hysterectomy, bilateral oophorectomy, or bilateral salpingectomy), or who are postmenopausal. Women will be considered postmenopausal if they have been amenorrheic for 12 months prior to the planned date of randomization without an alternative medical cause. The following age-specific requirements apply:

- Women < 50 years old would be considered postmenopausal if they have been amenorrheic for 12 months or more following cessation of exogenous hormonal treatment.

CONFIDENTIAL

- Women ≥ 50 years old would be considered postmenopausal if they have been amenorrheic for 12 months or more following cessation of all exogenous hormonal treatment.

Women of childbearing potential must use one highly effective form of birth control. A highly effective method of contraception is defined as one that can achieve a failure rate of less than 1% per year when used consistently and correctly. Women of childbearing potential who are sexually active with a non-sterilized male partner must agree to use one highly effective method of birth control, as defined in the list below this paragraph, from Day 1 and agree to continue through 365 days following administration of AZD7442/placebo. Periodic abstinence (calendar, symptothermal, post-ovulation methods), withdrawal (coitus interruptus), spermicides only, and lactational amenorrhea method are not acceptable methods of contraception. Female condom and male condom should not be used together. All women of childbearing potential must have a negative pregnancy test result at the screening visit and during the study as in the laboratory manual (**Table 1: Schedule of Assessments**).

List of highly effective birth control methods:

- Intrauterine device
- Intrauterine hormone-releasing system (IUS)
- Bilateral tubal occlusion
- Vasectomized partner (provided that partner is the sole sexual partner of the woman of childbearing potential study participant and that the vasectomized partner has received medical assessment of the surgical success).
- Combined (estrogen- and progestogen-containing hormonal contraception) with inhibition of ovulation
    - Oral (combined pill)
    - Intravaginal
    - Injectable
    - Transdermal (patch)
- Progestogen-only hormonal contraception with inhibition of ovulation
    - Oral
    - Injectable
    - Implantable

#### 8.6. Rescue Medicine

Not Applicable

#### 8.7. Non-Research Standard of Care

CONFIDENTIAL

Not Applicable

### 9. Study Intervention Discontinuation and Subject Discontinuation / Withdrawal

#### 9.1. Halting Criteria and Discontinuation of Study Intervention

##### 9.1.1. Individual Drugs Halting

For an individual subject, the infusion must be immediately stopped if they have a suspected drug-related event of hypersensitivity (Grade 2 or higher) during the infusion.

##### 9.1.2. Study Halting for Safety

Given severity of illness in COVID-19, there are no pre-specified stopping rules. Instead, there will be close oversight by the protocol team and frequent DSMB reviews for safety.

##### 9.1.3. Withdrawal from Randomized Treatment or from the Study

Patients are free to withdraw from participation in the study at any time upon request, without any consequence. Patients should be listed as having withdrawn consent only when they no longer wish to participate in the study and no longer authorize the Investigators to make efforts to continue to obtain their outcome data. Every effort should be made to encourage patients to remain in the study for the duration of their planned outcome assessments. Patients should be educated on the continued scientific importance of their data, even if they discontinue study drug. In the case of a patients becoming lost to follow-up, attempts to contact the patient should be made and documented in the patient's medical records. Please see 9.1.6 (Lost to Follow-Up) for further details.

##### 9.1.4. Discontinuation of Study Drug

Participants will receive a single dose of 600 mg AZD7442. It may be necessary for a participant to temporarily interrupt their administration of AZD7442/placebo.
If the administration of AZD7442/placebo is not resumed, the participant should remain in the study to be evaluated.

Unless the patient withdraws consent, those who discontinue study drug early should remain in the study for further acquisition of endpoint measurements. The reason for patient discontinuation or interruption of IV administration of study drug should be documented in the case report form.

##### 9.1.5. Withdrawal of Patients from the Study

CONFIDENTIAL

A patient may be removed from the study for the following reasons post initial dosing; however, whenever possible the patient should be followed for safety evaluations per protocol:

- Patient withdraws consent
- Discontinuation from the study is requested by the patient for any reason
- Discontinuation from the study is requested by or one of the investigators upon medical judgment
- Particularly in the arm standard of care alone for patients with severe disease
- Death of the patient
- Termination of the study
- Lost to follow-up

Patients who withdraw from this study or are lost to follow-up after signing the informed consent form (ICF) and administration of the study product, will not be replaced. The reason for patient discontinuation from the study will be recorded on the appropriate case report form.

###### 9.1.6. Lost to Follow-Up

A subject will be considered lost to follow-up if he or she fails to appear for a follow-up assessment and cannot be contacted with good effort. These efforts will be documented in the subject's record.

Site personnel, or an independent third party, may attempt to collect the survival status of the participant within legal and ethical boundaries for all participants randomized, including those who did not get the AZD7442/placebo. Public sources may be searched for survival status information. If survival status is determined as deceased, this will be documented, and the participant will not be considered lost to follow-up. Sponsor personnel will not be involved in any attempts to collect survival status information.

#### 10. Study Assessments and Procedures

##### 10.1. Screening Assessment

After obtaining the informed consent, some or all of the following assessments are performed to determine eligibility requirements as specified in the inclusion and exclusion criteria:

- Perform a SARS-CoV-2 rapid antigen test and obtain Ct value, lineage
- Focused medical history, including the following information:
  - Day of onset of COVID-19 symptoms
  - Day of hospitalization

CONFIDENTIAL

- COVID-19 vaccination initiation (yes/no) and if yes history of COVID-19 vaccination
    - History of chronic medical conditions related to inclusion and exclusion criteria
    - Medication allergies or contraindication
    - Review medications and therapies for this current illness and record on the appropriate CRF
- Counsel subjects to use adequate birth control methods required during the study to avoid pregnancy
- Obtain age and sex
- Obtain weight and height
- Obtain ethnicity
    - In the context of a new disease, the ethnicity might be a prognostic factor for COVID-19. Moreover, the ethnicity is known to be an important factor for drug concentrations and the pharmacodynamics / pharmacokinetic of study drugs may be different according to ethnicity
- Obtain blood for screening laboratory evaluations if not done in the preceding 36 hours:
    - Liver enzymes and hepatic function (LDH, ALT, AST);
    - Cell counts (white cells including absolute neutrophils and lymphocyte counts, red cells through haemoglobin, platelet count);
    - Coagulation and inflammatory status (fibrinogen, D-Dimer, prothrombin time]); C-reactive protein, ferritinemia, serum total immunoglobulins G);
    - Renal function: creatininemia.

The following data collected before randomization will be reported centrally to the WHO Solidarity trial (only for treatment arms shared with Solidarity):
• Country, hospital (from a list of approved hospitals) and randomising doctor
• Confirmation that informed consent has been obtained
• Patient identifiers, admission date, age and sex
• Patient characteristics (yes/no): current smoking, diabetes, heart disease, chronic lung disease, chronic liver disease, asthma, HIV

Clinical screening laboratory evaluations will be performed locally by the site laboratory.

The overall eligibility of the subject to participate in the study will be assessed once all screening values are available. The screening process can be suspended prior to complete assessment at any time if one or more exclusion criteria are identified by the study team. Study subjects who qualify should be randomized as soon as possible after enrolment.

CONFIDENTIAL

After inclusion, a SARS-CoV-2 RT-PCR test will be conducted in all patients.

Analysis of the immune-inflammatory response and of the kinetics and functional capacities of patients anti-SARS-CoV-2 antibodies will be analyzed centrally.

The volume of venous blood to be collected is presented in
Table 4: Venepuncture **Volumes**.

#### 10.2. Efficacy Assessment

For all baseline assessments and follow-up visits, refer to SoA for procedure to be done, and details below for each assessment.

##### 10.2.1. Measures of clinical support

At each study day while hospitalized, the following measure of clinical support should be assessed according to SoA (see Table 1: Schedule of Assessments):

- Hospitalization
- Oxygen requirement
- High flow oxygen devices
- Non-invasive mechanical ventilation (via mask)
- Mechanical ventilator requirement (via endotracheal tube or tracheostomy tube)
- ECMO requirement

##### 10.2.2. Ordinal Scale

The ordinal scale is an assessment of the clinical status at the first assessment of a given study day. The scale is as follows:

1. Not hospitalized, no limitations on activities
2. Not hospitalized, limitation on activities;
3. Hospitalized, not requiring supplemental oxygen;
4. Hospitalized, requiring supplemental oxygen;
5. Hospitalized, on non-invasive ventilation or high flow oxygen devices;
6. Hospitalized, on invasive mechanical ventilation or ECMO;
7. Death.

Participants who answer yes to the question "are you able to carry out all the activities, including physical activities, that you carried out before the first symptoms of COVID-19?" will have a score of 1 (Not hospitalized, no limitations on activities). Participants who answer no to the question "are you able to carry out all the activities, including physical activities, that you carried out before the first

CONFIDENTIAL

symptoms of COVID-19?" will have a score of 2 (Not hospitalized, limitations on activities).
The scale will be obtained at baseline, and Day 15 (Primary Endpoint) and at Day 29.
At Days 15, if the patient is discharged from hospital, it will be collected by phone.
At Days 29, 90, 180 and 365, if the patient did not attend the scheduled visit, it will be collected by phone.

##### 10.2.3. Sustained recovery

Time from randomization to sustained recovery, defined as being discharged from the index hospitalization, followed by being alive and home for 14 consecutive days prior to Day 90.

The participants will be contacted by phone 14 days after hospital discharge to assess vital status and re-hospitalisation since index discharge. Several attempts will be made to contact the participant or his/her relatives.

Residence or facility groupings to define home are:
1) Independent/community dwelling with or without help, including house, apartment, undomiciled/homeless, shelter, or hotel;
2) Residential care facility (e.g., assisted living facility, group home, other non-medical institutional setting);
3) Other healthcare facility (e.g., skilled nursing facility, acute rehab facility);
4) Long-term acute care hospital (hospital aimed at providing intensive, longer term acute care services, often for more than 28 days).

Home is defined as the level of residence or facility where the participant was residing prior to hospital admission leading to enrollment in this protocol. Lower (less intensive) level of residence or facility will also be considered as home. By definition, "home" cannot be a "short-term acute care" facility. Participants previously affiliated with a "long-term acute care" hospital recover when they return to the same or lower level of care.

Readmission from "home" may occur and if this occurs within 14 days of the first discharge to "home", then the primary endpoint will not be reached until such time as the participant has been at home for 14 consecutive days.

Participants residing in a facility solely for public health or quarantine purposes will be considered as residing in the lowest level of required residence had these public health measures not been instated.

CONFIDENTIAL

Some recovering patients are discharged from the hospital while still requiring continuous low flow supplementary oxygen to maintain satisfactory blood oxygenation for some period of time.

##### 10.2.4. NEWS-2 Score

The NEWS-2 score has demonstrated an ability to discriminate patients at risk of poor outcomes [28]. This score is based on 7 clinical parameters. The NEWS Score is being used as an efficacy measure.

This should be evaluated at the first assessment of a given study day. These parameters can be obtained from the hospital chart using the last measurement prior to the time of assessment. This is recorded for the day obtained. i.e. on Day 3, Day 3 score is obtained and recorded as Day 3.

**Tc**

| **Physiological parameter** | **3** | **2** | **1** | **Score**<br>**0** | **1** | **2** | **3** |
|---|---|---|---|---|---|---|---|
| Respiration rate (per minute) | ≤8 | | 9–11 | 12–20 | | 21–24 | ≥25 |
| $SpO_2$ Scale 1 (%) | ≤91 | 92–93 | 94–95 | ≥96 | | | |
| $SpO_2$ Scale 2 (%) | ≤83 | 84–85 | 86–87 | 88–92<br>≥93 on air | 93–94 on oxygen | 95–96 on oxygen | ≥97 on oxygen |
| Air or oxygen? | | Oxygen | | Air | | | |
| Systolic blood pressure (mmHg) | ≤90 | 91–100 | 101–110 | 111–219 | | | ≥220 |
| Pulse (per minute) | ≤40 | | 41–50 | 51–90 | 91–110 | 111–130 | ≥131 |
| Consciousness | | | | Alert | | | CVPU |
| Temperature (°C) | ≤35.0 | | 35.1–36.0 | 36.1–38.0 | 38.1–39.0 | ≥39.1 | |

*SpO2 scale: For patients confirmed to have hypercapnic respiratory failure on blood gas analysis on either a prior or their current hospital admission, and requiring supplemental oxygen, we recommend (i) a prescribed oxygen saturation target range of 88–92%, and (ii) that the dedicated SpO2 scoring scale (Scale 2) on the chart should be used to record and score the oxygen saturation for the NEWS. The decision to use SpO2 scale 2 should be made by a competent clinical decision maker and should be recorded in the patient's clinical notes. In all other circumstances, the regular NEWS SpO2 scale 1 should be used.*

*CVPU: Confusion or arousable only to voice (V) or pain (P), or unresponsive (U).*

In addition, following clinical information should be collected:

- Symptoms
- Complications
- Assessment of measures of clinical support
    - Oxygen requirement and flow
    - High flow oxygen devices requirement
    - Non-invasive mechanical ventilation and parameters of ventilation
    - Mechanical ventilator requirement (via endotracheal tube or tracheostomy tube) and parameters of ventilation
    - Hemodynamic support, drug and dose
    - ECMO requirement and parameters

The following data collected during follow-up will be reported centrally to the WHO Solidarity trial, only for treatment arms shared with Solidarity (which is not the case of the AZD7442 IMP):

- When patients die or are discharged, follow-up ceases and it is reported
- Which study drugs were given (and for how many days)
- Whether ventilation or intensive care was received (and, if so, when)
- Date of discharge, or date and cause of death.

##### 10.2.5. Long term health status

###### Hospitalization

Hospitalization following hospital discharge is defined as the admission from "home" (as defined in 10.2.3) to a short-term acute care facility.
The event of hospitalization following hospital discharge will be assessed by phone at days 90, 180 and 365 or during a medical consultation, for a subset of 25% of enrolled subjects.

###### Reinfection with SARS-CoV-2

Reinfection with SARS-CoV-2 will be assessed by phone at days 180 and 365 or during a medical consultation, for a subset of enrolled subjects. A positive RT-PCR or Antigen test will be required in order to confirm reinfection.

##### 10.2.6. Exploratory Assessment

###### Viral Shedding

NP swabs or lower respiratory tract samples will be collected at different time points (see Table 1: Schedule of Assessments) and stored as outlined in the

CONFIDENTIAL

laboratory manual. The decay of viral shedding of SARS-CoV-2 in NP or respiratory tract samples will be analyzed using normalized viral load. Lack of control of the infection or virological escape from the treatment can be monitored on these viral kinetics data

If virology assays can be set up with enough numbers of specimens tested, this data will be submitted as part of the Clinical Study Report. This may be submitted separately, as a supplemental Clinical Study Report. In France, these samples can be centralized and analyzed in the National Reference Center (CNR) of Respiratory Virusesin Lyon.

##### Concentrations of study medications and Anti-Drug Antibodies

Concentration of AZD7442 and the presence of Anti-Drug Antibodies (ADA) will be measured at different time points (see Table 1: Schedule of Assessments). The PK and ADA results will not be available to site. The actual date and time for each sample will be collected in the eCRF.

Samples will be collected, labelled stored and shipped as detailed in the Laboratory Manual. A central laboratory will be used for the logistic arrangements.

Samples for determination of serum drug concentration and serum anti-drug antibodies will be assayed using validated bioanalytical methods. Full details of the analytical drug concentrations and ADA incidence results will be described in separate Bioanalytical Reports.
Placebo samples will be analyzed for ADA but will not be analyzed for PK, unless there is a need to confirm that correct treatment has been given to study participants.
Drug concentration information that may unblind the study will not be reported to investigative sites or blinded personnel until the study has been unblinded.

##### Thoracic CT scan at baseline

Thoracic CT-scan will be performed at enrolment (baseline), whenever possible in centers managing COVID-19 patients. A standardized framework will be used to assess the CT scans in each centre.

##### Genetic variants associated with severity of disease and/or drug response

Whole blood for whole exome sequencing will be collected at baseline in subjects who agree to participle in the genetic analysis.

##### Immune-inflammatory response during treatment of COVID-19

Blood levels of immunothrombosis markers, IFNs (and anti-IFNs antibodies), and complement activation products will be measured at different time points.

CONFIDENTIAL

#### Kinetics and functions of anti-SARS-CoV-2 antibodies

Blood levels of anti-SARS-CoV-2 immunoglobulins will be measured at different time points.
SARS-CoV-2 functional neutralization and ADCC ability of the blood will be determined at different time points.

#### 10.3. Safety and Other Assessments

##### 10.3.1. Safety procedures

Study procedures are specified in the SoA. A study physician licensed to make medical diagnoses and listed will be responsible for all trial-related medical decisions. If follow-up assessments are conducted in the outpatient setting, each center has to manage the collection of the data and the completion of the CRF.

- Physical examination: A symptom-directed (targeted) physical examination (including vital signs, including pulse rate, pulse oximetry, blood pressure, and body temperature) will be performed to evaluate for any possible adverse event.
- Clinical laboratory evaluations (performed in local laboratories):
    - Fasting is not required before collection of laboratory samples.
    - Blood will be collected at the time points indicated in the laboratory manual. Clinical laboratory parameters include
        - renal function: creatininemia;
        - liver enzymes and hepatic function: total bilirubin, LDH, ALT, AST;
        - cell counts: white cells including absolute neutrophils and lymphocyte counts, red cells through haemoglobin, platelet count,
        - coagulation and inflammatory status: fibrinogen, D-Dimer, prothrombin time, C-reactive protein, ferritinemia.
    - This testing will be performed at each clinical trial site in real time.

CONFIDENTIAL

**Table 4: Venepuncture Volumes**

| Day +/- Window | Screening / Baseline | 3 | 8[1] ± 1 | 15[1] ± 2 | 29 ± 3 | 90 ± 7 | 180[3] ± 14 | 365[3] ± 14 |
|---|---|---|---|---|---|---|---|---|
| Safety Laboratory tests[2] | X 15mL | X 15mL | X 15mL | X 15mL | X 15mL | X 15mL | X 15mL | X 15mL |
| Serum for exploratory objectives | X 6mL | X 6mL | X 6mL | X 6mL | X 6mL | X 6mL | X 6mL | X 6mL |
| Plasma for exploratory objectives | X 6mL | X 6mL | X 6mL | X 6mL | X 6mL | X 6mL | X 6mL | X 6mL |
| Serum for drug concentration | X 5mL | X 2.5mL | X 2.5mL | X 2.5mL | X 2.5mL | X 2.5mL | X 2.5mL | X 2.5mL |

| | | | | | | | | |
|---|---|---|---|---|---|---|---|---|
| Serum for anti-drug antibodies | X<br>3,5mL | | | X<br>3,5mL | X<br>3,5mL | X<br>3,5mL | X<br>3,5mL | X<br>3,5mL |
| Blood for genetic analysis (EDTA) | X<br>6mL | | | | | | | |
| Total volume | 41,5mL | 29.5mL | 29.5mL | 33mL | 33mL | 33mL | 33mL | 33mL |
| Maximum total for all study days | | | | | | 199,5mL | | 265,5mL |

[1]*For hospitalized patients only*

[2]*Creatinin; Liver enzymes and hepatic function (LDH, ALT, AST); cell counts (neutrophils and total lymphocyte counts, haemoglobin, platelets); coagulation and inflammation status (fibrinogen, D-Dimer, prothrombin time, C-reactive protein, ferritinemia, serum total immunoglobulins G).*

[3] In a subset of 25% of enrolled participants

If a physiologic parameter, e.g., vital signs, or laboratory value is outside of the protocol-specified range, then the measurement may be repeated once if, in the judgment of the investigator, the abnormality is the result of an acute, short-term, rapidly reversible condition or was an error.
A physiologic parameter may also be repeated if there is a technical problem with the measurement caused by malfunctioning, or an inappropriate measuring device (i.e., inappropriate-sized BP cuff).

##### 10.3.2. Virological surveillance

NP swab or lower respiratory tract sample will be collected at the time points indicated in the laboratory manual.
After screening assessment (refer to section Screening Assessment), virological laboratory parameters include:

- At day 8 when patient is hospitalized: Qualitative (Ct value) PCR for SARS-CoV-2 at each clinical trial site in real time ;
- If Ct value <31 at D8 or a lack of viral load decline between D1 and D8 indicating an incomplete antiviral response:
    - Quantitative PCR for SARS-CoV-2 D1/D8
    - Whole Genome sequencing for escape variants D1/D8

  In France, these samples will be centralized and analyzed in the CNR of Respiratory Viruses of virology in Lyon as soon as possible.

#### 10.4. Adverse Events and Serious Adverse Events

##### 10.4.1. Definition of Adverse Event (AE)

AE means any untoward medical occurrence associated with the use of an intervention in humans, whether or not considered intervention related. An AE can, therefore, be any unfavourable and unintended sign (including an abnormal laboratory finding), symptom, or disease temporally associated with the use of medicinal (investigational) product.

All AEs will be assessed for severity and relationship to study intervention.

CONFIDENTIAL

Any medical condition that is present at the time that the subject is screened will be considered as baseline and not reported as an AE. However, if the severity of any pre-existing medical condition increases, it should be recorded as an AE.

Given the nature of severity of the underlying illness, subjects will have many symptoms and abnormalities in vitals and laboratory. All Grade 3 and 4 AEs will be captured as AEs in this trial and will be considered as notable events. In addition, Grade 1-2 hypersensitivity-related and infusion related AEs will be captured up until and including the visit at D29. Examples of hypersensitivity-related events include, but are not limited to: Skin rash, hives, itching, swelling, wheezing, itchy/watery eyes.

##### 10.4.2. Definition of Serious Adverse Event (SAE)

An SAE is defined as "An AE or suspected adverse reaction is considered serious if, in the view of either the investigator or the sponsor, it results in any of the following outcomes:

- Death,
- a life-threatening AE,
- prolongation of existing hospitalization,
- a persistent or significant incapacity or substantial disruption of the ability to conduct normal life functions,
- or a congenital anomaly/birth defect.
- important medical events

Important medical events that may not result in death, be life-threatening, or require hospitalization may be considered serious when, based upon appropriate medical judgment, they may jeopardize the patient or subject and may require medical or surgical intervention to prevent one of the outcomes listed in this definition. Examples of such medical events include allergic bronchospasm requiring intensive treatment in an emergency room or at home, blood dyscrasias or convulsions that do not result in inpatient hospitalization, or the development of drug dependency or drug abuse."

"Life-threatening" refers to an AE that at occurrence represents an immediate risk of death to a subject. An event that may cause death if it occurs in a more severe form is not considered life-threatening. Similarly, a hospital admission for an elective procedure is not considered a SAE.

All SAEs, as with any AE, will be assessed for severity and relationship to study intervention.

CONFIDENTIAL

All SAEs will be recorded on the appropriate SAE CRF.

All SAEs will be followed through resolution or stabilization by a licensed study physician.

All SAEs will be reviewed and evaluated and those considered as pertaining to participants safety will be immediately sent to the DSMB (all SAE are periodically reviewed), and the IRB/EC according the applicable local requirements.

##### 10.4.3. Definition of Notable Events

Selected Grade 1 and 2 hypersensitivity and infusion related events and all grade 3 and 4 AEs are considered as notable events and must be collected into eCRF (AE section) within 24 hours after the investigator becomes aware of the confirmed event. All notable events associated to seriousness criteria must be reported as SAE.

##### 10.4.4. Definition of Adverse Event of Special Interest (AESI)

An AESI is an AE that required an immediate notification to sponsor. AESIs for AZD7442 are listed below, they include:

- Anaphylaxis and other serious hypersensitivity reactions, including immune complex disease
- Grade 3 and 4 injection site reactions or infusion-related events

All AESI have to be sent by the investigator to the sponsor immediately and no later than 24h after being aware of it using SAE form.

##### 10.4.5. Protocol-Specified Exempted Serious Events

These events may refer to disease- or complication-related events that are common in Covid-19 patients and can be serious.
They are reported systematically on the eCRF in the pre-defined daily data sections. However, they will NOT be reported as SAE in the eCRF, unless the investigator considered that there is a reasonable possibility that the study intervention (blinded investigational agent/ placebo or study-supplied SoC treatment) enhanced or caused the event. DSMB will review periodically these complications.
These events may occur during the initial hospitalization, lead to a re-admission, or occur in a later hospitalization during follow-up.
Protocol-specified exempted serious events are:

- Acute kidney injury / Acute renal failure
- Grade 3 and above transaminase increased
- Pancreatitis

CONFIDENTIAL

- Bacteremia
- Bacterial pneumonia
- Cardiac arrhythmia / Cardiac ischemia / Cardiac arrest
- Endocarditis
- Myocarditis
- Pericarditis
- Congestive heart failure
- Acute Respiratory Distress Syndrome
- Pneumothorax / Pleural effusion
- Meningitis / Encephalitis
- Stroke / Cerebrovascular accident
- Coma / Confusion
- Gastrointestinal hemorrhage
- Pulmonary thrombosis
- Arterial or deep vein thromboembolic events

##### 10.4.6. Suspected Unexpected Serious Adverse Reactions (SUSAR)

A SUSAR is any SAE where a causal relationship with the study product is at least reasonably possible but is not listed in the Investigator Brochure (IB), Package Insert, and/or Summary of Product Characteristics. All SUSAR and SAR will be sent to DSMB in real time for a safety review.

##### 10.4.7. Classification of an Adverse Event

The determination of seriousness, severity, and causality will be made by an on-site investigator who is qualified (licensed) to diagnose AE information, provide a medical evaluation of AEs, and classify AEs based upon medical judgment. This includes but is not limited to physicians, physician assistants, and nurse practitioners.

**Severity of Adverse Events**

All AEs and SAEs will be assessed for severity, according to the Division of AIDS (DAIDS) Table for Grading the Severity of Adult and Paediatric Adverse Events, version 2.1 (July 2017).

For AEs not included in the protocol defined grading system, the following guidelines will be used to describe severity.

- Mild (Grade 1): Events that are usually transient and may require only minimal or no treatment or therapeutic intervention and generally do not interfere with the subject's usual activities of daily living.

CONFIDENTIAL

- Moderate (Grade 2): Events that are usually alleviated with additional specific therapeutic intervention. The event interferes with usual activities of daily living, causing discomfort but poses no significant or permanent risk of harm to the research subject.

- Severe (Grade 3): Events interrupt usual activities of daily living, or significantly affects clinical status, or may require intensive therapeutic intervention. Severe events are usually incapacitating.

- Very severe (Grade 4): Events that are potentially life threatening.

AEs characterized as intermittent require documentation of onset and duration of each episode. The start and stop duration of each reported AE will be recorded on the appropriate CRF. Changes in the severity of an AE will be documented to allow an assessment of the duration of the event at each level of intensity.

**Relationship to Study Intervention**

For each reported adverse reaction, the PI or designee must assess the relationship of the event to the study product using the following guideline:

- Related – The AE is known to occur with the study intervention, there is a reasonable possibility that the study intervention caused the AE, or there is a temporal relationship between the study intervention and event. Reasonable possibility means that there is evidence to suggest a causal relationship between the study intervention and the AE.

- Not Related – There is not a reasonable possibility that the administration of the study intervention caused the event, there is no temporal relationship between the study intervention and event onset, or an alternate aetiology has been established.

##### 10.4.8. Time Period and Frequency for Event Assessment and Follow-Up

For this study, all Grade 3 and 4 AEs, AESI and all SAEs occurring from the time the informed consent is signed through the Day 456 (end of study) visit will be documented, recorded, and reported. Selected Grade 1 and 2 hypersensitivity and infusion related events will be captured up until and including the visit at D29.

**Investigators Reporting of AEs**

Information on selected Grade 1 and 2 hypersensitivity and infusion related events and all Grade 3 and Grade 4 AEs should be recorded on the appropriate CRF. All clearly related signs, symptoms, and results of diagnostic procedures

CONFIDENTIAL

performed because of an AE should be grouped together and recorded as a single diagnosis. If the AE is a laboratory abnormality that is part of a clinical condition or syndrome, it should be recorded as the syndrome or diagnosis rather than the individual laboratory abnormality. Each protocol-defined reported AE will also be described in terms of duration (start and stop date), severity, association with the study product, action(s) taken, and outcome.

##### 10.4.9. Serious Adverse Event Reporting

**Investigators Reporting of SAEs**

- **e-CRF Notification:**

Any AE that meets a protocol-defined criterion as a SAE must be notified immediately (within 24 hours of site awareness) on an SAE form.

When the eCRF SAE notification form is completed by the investigator, an automatic email is sent immediately to the Inserm safety department.

- **Back up circuit when eCRF is unavalaible:**

The investigator must report all SAE/SAR using the CRF SAE printed form, dated and signed to:

Institut Thématique Santé Publique – Recherche Clinique et Thérapeutique (ITSP – RCT)
Mission Réglementation et Qualité et Recherche Clinique (RQRC)


If an SAE is declared by paper circuit, it is the responsibility of the site to re-enter the form in the eCRF

- **Relevant documentation related to SAE** (e.g. hospitalization report, laboratories results…):

The investigator sends all relevant coded documentation related to the SAE to the Inserm safety department, by fax: 01 53 94 60 02 or by.
The Inserm safety department will review and assess the SAE for regulatory reporting and potential impact on study subject safety and protocol conduct.

At any time after completion of the study, if the site PI or appropriate sub-investigator becomes aware of an SAE, the site PI or appropriate sub-investigator will report the event to the Inserm safety department.

**Regulatory Reporting of safety data**

CONFIDENTIAL

All Suspected Unexpected Adverse Reactions (SUSAR) have to be reported, within the legal timeframe, by the sponsor to the National Competent Authority of the Member State concerned and the Ethic Committees according to the local requirements.

The timelines for expedited initial reporting (day 0) starts as soon as the information containing the minimum reporting criteria has been received by the sponsor.

For fatal and life-threatening SUSAR, the sponsor should report at least the minimum information without delay after being made aware of the case.
SUSAR which are not fatal and not life-threatening are to be reported within 15 calendar days.
For fatal and life-threatening SAR, the sponsor should report at least the minimum information without delay after being made aware of the case to NCA according to the local requirements.

If significant new information on an already reported case is received by the sponsor, this information should be reported as a follow-up report within 8 days after being made aware of the relevant complementary information.

Once a year throughout the clinical trial, the sponsor should submit to the national competent authority and the Ethics Committee (if applicable) of the Member States, an annual safety report including the list of all Serious Adverse Reaction.

The complet safety data circuit is available in the safety management plan of the Discovery trial.

###### 10.4.10. Reporting of Pregnancy

Pregnancy is not an AE. However, any pregnancy that occurs during study participation should be reported to the sponsor on the appropriate CRF form. Pregnancy should be followed to outcome. The investigator has to notify to the sponsor, immediately and no later than 48 hours after being made aware of it, any pregnancy and its outcome.
The investigator has to follow the subject until the end of the pregnancy or its interruption and to notify the outcome to the sponsor using the “Final pregnancy notification form”.

#### 10.5. Safety concerns including unanticipated Problems

##### 10.5.1. Definition of Unanticipated Problems (UP)

CONFIDENTIAL

An Unanticipated Problem is any event, incident, experience, or outcome that meets the following criteria:

- Unexpected in terms of nature, severity, or frequency given (a) the research procedures that are described in the protocol-related documents, such as the EC-approved research protocol and informed consent document; and (b) the characteristics of the subject population being studied;
- Related or possibly related to participation in the research ("possibly related" means there is a reasonable possibility that the incident, experience, or outcome may have been caused by the procedures involved in the research); and
- Suggests that the research places subjects or others at a greater risk of harm (including physical, psychological, economic, or social harm) than was previously known or recognized.

##### 10.5.2. Unanticipated Problem Reporting

To satisfy the requirement for prompt reporting, unanticipated problems (UP) will be reported using the following timeline:

- UPs that are SAEs will be reported to the IRB/EC and to the Statistical and Data Coordinating Centreand study sponsor within 24 hours of the investigator becoming aware of the event per the above describe SAE reporting process.
- Any other UP will be reported to the IRB/EC and to the SDCC/study sponsor within 3 days of the investigator becoming aware of the problem.

##### 10.5.3. Reporting Unanticipated Problems to Subjects

Subjects will be informed of any UPs that occur as part of their participation in this study.

#### 11. Statistical Considerations

This study is intended to allow for adaptations with the ability to add a new experimental arm if one becomes available. A brief summary is provided here. Details will be described in the statistical analysis plan, to be finalized prior to the first DSMB meeting.

If additional data become available to add an experimental therapy, analyses of experimental arms will be performed comparing concurrently enrolled control subjects.

##### 11.1. Statistical Hypotheses

The primary outcome uses an ordinal severity scale with 7 categories, analysed using the proportional odds model. This model assumes that the treatment to

CONFIDENTIAL

control odds ratio of being classified in a given severity category "i" or better is the same for each category. The null hypothesis being tested is whether the odds of improvement on the ordinal scale is the same for the control and experimental treatment arms (i.e., whether the common odds ratio differs is 1).

#### 11.2. Sample Size Determination

There will be no pre-planned interim analysis for efficacy nor futility in this version of the protocol. If good evidence emerges while the study is continuing that some other treatment(s) should also be evaluated, then it will be decided that one or more extra arms may be added while the trial is in progress. For treatment arms shared with Solidarity, those decisions will be made globally by the Executive Group of the International Steering Committee of Solidarity from information of its DSMC.

<u>Sample size estimates provided as a reference but not to indicate final number of patients to be randomised.</u>

The odds ratio represents the odds of improvement in the ordinal scale for treatment relative to control control [29] shows that the sample size to detect a given odds ratio for 1:1 randomization using a 2-tailed test at level αis given by

$$\frac{12\left(z_{\alpha/2}+z_{\beta}\right)^2}{\theta^2\left(1-\sum_{i=1}^{6} p_i^3\right)},$$

where $\theta$ is the log odds ratio, $p_i$ is the overall probability (combined over both arms) of being in the ith category of the ordinal outcome, and $z_{\alpha/2}$ and $z_{\beta}$ are the $1-\alpha/2$ and βth quantiles of the standard normal distribution.

**Table 5** displays the outcomes under standard of care used for sample size determination. They were obtained from patients included in DisCoVery trial from July 1st to December 31 2020, with results available in the database locked on January 6, 2021 for the DSMB. More precisely, D15 outcomes of 93 patients from the SOC arm, included in the moderate disease strata and with duration of symptoms of less of 9 days are reported and used.

**Table 5: Outcomes on Day 15 used for sample size determination**

| | outcome (%) |
|---|---|
| Death | 5 |

CONFIDENTIAL

| | |
|---|---|
| Hospitalized, on mechanical ventilation or ECMO | 6 |
| Hospitalized, on non-invasive ventilation or high flow oxygen devices | 6 |
| Hospitalized, requiring supplemental oxygen | 17 |
| Hospitalized, not requiring supplemental oxygen | 7 |
| Not hospitalized, limitation on activities | 35 |
| Not hospitalized, no limitations on activities | 24 |

The targeted strength of evidence for a stand-alone registration trial in essence is two-sided p=0.05, which corresponds to a (one-sided) false positive error rate of 0.025.

Since a large proportion of patients are moderately ill patients, we power the study for an odds ratio 1.5 (an odds ratio higher than 1 indicates superiority of the experimental treatment over the control for each ordinal scale category), with 90% power and using an overall two-sided type I error rate of 0.05.

The number of subject is computed for a power of 90% in patients included in the randomization strata with positive antigene (Ag+) results at baseline.
We would need 413 evaluable patients per arm with Ag+ results.

Despite the changes in the inclusion/exclusion criteria, we decided not to change the number of subjects needed.
For the delay of symptoms before inclusion from 9 to 11 days, as the number of subjects was computed for the primary analysis on only positive antigen patients it will have no impact.

The inclusion of vaccinated (fully or partly) patients could have an impact on the statistical power for an unchanged number of subjects, depending:

- on the distribution of the outcome at day 15 in vaccinated patients receiving SoC, (which migh be different that the expected distribution presented in the **Table 5**)
- on the expected treatment effect in vaccinated patients,

CONFIDENTIAL

– on the proportion of patients vaccinated.

For the last point, we decided to cap the proportion of vaccinated patients to 20% of antigene positive patients.

No data are yet available on the change brought by vaccination for the first two two assumptions. **Table 6** presents power calculation, for a sample size of 413 per arm with Ag + results, for various scenario varying

- the outcome at day 15 in vaccinated vs non-vaccinated patients
- the treatement effect in vaccinated vs non-vaccinated patients

**Table 6. Power computation according for 413 subjects per arm and 20% vaccinated patients, for various assumptions on treatment effect and outcome at day 15 in vaccinated vs non-vaccinated patients**

| **Outcome distribution in SoC arms in Vaccinated vs Non-vaccinated patients** | **OR of treatment effect in vaccinated patients** | | |
|---|---|---|---|
| | **OR = 1.3** (**lower** treatment effect than in non-vaccinated) | **OR = 1.5** (**same** treatment effect than in non-vaccinated) | **OR = 1.7** (**better** treatment effect than in non-vaccinated) |
| OR = 0.8 (**worse** outcome at day 15 in vaccinated than in non-vaccinated) | 86% | 90% | 93% |
| OR = 1 (**same** outcome at day 15 in vaccinated than in non-vaccinated) | 86% | 90% | 93% |
| OR = 1.2 (**better** outcome at day 15 in vaccinated than in non-vaccinated) | 86% | 90% | 93% |

As can be seen in **Table 6**, the power is always greater than 85%, hence the decision not to change the number of subjects to be included.

The number of evaluable Ag- patients will be limited to at most 30% of the total number of patients, hence at most 177 Ag- patients per arm. At most 20% of vaccinated Ag- patients will be included.

The number of evaluable patients per arm is 590. Therefore, we need to include 620 patients per arm (434 Ag+ and 186 Ag-) to account for 5% not evaluable patients (informed consent retracted, not valid, …).

#### 11.3. Populations for Analyses

CONFIDENTIAL

The following analysis sets are defined:

- Positive intention-to-treat (Positive ITT): all randomized patients with positive SARS-CoV-2 antigenic status at randomization who will be analysed in their randomization group.
- Global intention-to-treat (Global ITT): all randomized patients with positive or negative SARS-CoV-2 antigenic status at randomization who will be analysed in their randomization group.
- Positive modified intention-to-treat (Positive mITT): patients of the Positive ITT in whom the allocated treatment was initiated.
- Global modified intention-to-treat (Global mITT): patients of the Global ITT in whom the allocated treatment was initiated.
- Positive Safety analysis set: all randomized patients with positive SARS-CoV-2 antigenic status at randomization and analysed in their actual treatment group.
- Global Safety analysis set: all randomized patients with positive or negative SARS-CoV-2 antigenic status at randomization and analysed in their actual treatment group.

The primary analysis for efficacy will be performed in the Positive intention-to-treat set. The primary analysis for safety will be performed in the Global Safety analysis set.

#### 11.4. Statistical Analyses

##### 11.4.1. General Approach

A statistical analysis plan (SAP) will be developed and filed with the study sponsor prior to database lock.

The primary analyses will be conducted on Ag+ patients. All tests will be stratified by geographic region (if sample size allows) and vaccination initiation (yes or no). For the sensitivity analysis, all tests on the full population will be additionaly stratified by antigenic status at enrollment (Ag+ and Ag-).

At the time all participants complete their Day 90 visit or have experienced a sustained recovery, a database lock will be initiated, and the study participants will be unblinded to the clinical study team members who are not involved with site operations or the classification of safety data. This version of the database will serve as the primary analysis dataset in this version of the protocol (the AZD7442 phase of the study), which will include the Clinical Study Report (CSR) output production, including formal testing of the primary and key secondary efficacy endpoints. When all participants complete their last safety follow-up

CONFIDENTIAL

visit, a final clinical database lock will occur, and the CSR outputs will be updated using the final data.

##### 11.4.2. Analysis of the Primary Efficacy Endpoint

The ordinal scale will be used to estimate a proportional odds model. The primary hypothesis test will be based on a test of whether the common odds ratio for treatment is equal to one. As noted earlier, the hypothesis test is, for large sample sizes, nearly the same as the Wilcoxon rank sum test.

Therefore, the procedure produces a valid p-value regardless of whether the proportional odds model is correct. Nonetheless, estimation and confidence intervals do require the model to be correct. Accordingly, we will evaluate model fit using a goodness-of-fit likelihood ratio test.

The distribution of severity results will be summarized by treatment arm as percentages. The validity of the proportionality assumption will be evaluated and tested. Intercurrent events for the primary estimand include the treatment of participants with any COVID-19 disease treatment. Intercurrent events will be handled by a treatment policy strategy. Efforts to minimize loss-to-follow-up will be considerable. However, small amounts of missing data may occur. In such cases, participants without final outcome data will be excluded from the analysis. Sensitivity analyses will evaluate the impact of making different assumptions about missing observations. These sensitivity analyses will be fully defined in the SAP.

##### 11.4.3. Analysis of the Key Secondary Efficacy Endpoint

The evaluation for the key secondary endpoint, time to sustained recovery through Day 90, will be performed only if the primary end point is significant (hierarchical testing). It will be performed using competitive risk to compare the cumulative incidence functions for sustained recovery between the treatment groups, taking into account the "competing risk" of death in analysing sustained recovery. Intercurrent events include the treatment of participants with any COVID-19 disease treatment. Intercurrent events will be handled by a treatment policy strategy. Missing data will be considered missing for the primary analysis of the key secondary endpoint. Sensitivity analyses will evaluate the impact of making different assumptions about missing observations. Sensitivity analyses will be carried out using outcomes, such as requiring continuous low flow oxygen upon discharge, which are variations of the definition of sustained recovery. These sensitivity analyses will be fully defined in the SAP.
In addition, sensitivity analyses will be carried out using outcomes which are variations of the definition of sustained recovery. These outcomes will use

CONFIDENTIAL

definitions of sustained recovery that consider continuous use of supplemental oxygen reported during the 14-day period at home following discharge. These outcomes are cited as secondary endpoints.

##### 11.4.4. Analysis of the Secondary Endpoint(s)

1) Differences in time-to-event endpoints (e.g., time to discharge) by treatment will be summarized with Kaplan-Meier curves and 95% confidence bounds.
2) Change in ordinal scale at specific time points will be summarized by proportions (e.g., proportion who have a 1-, 2-, 3-, or 4-point improvement or 1-, 2-, 3-, 4-point worsening).
3) Duration of event (e.g., duration of mechanical ventilation) will be summarized according to median days with quartiles.
4) Incidence data (e.g., incidence of new oxygen use) will be summarized as a percent with 95% confidence intervals.
5) Categorical data (e.g., 29-day mortality or ordinal scale by day) will be summarized according to proportions with confidence intervals on the difference or odds ratios for a binary or multiple category scale, respectively.

Missing data procedures will be described in the SAP.

##### 11.4.5. Safety Analyses

Safety endpoints include death through Day 29, SAEs, discontinuation of study infusions, and severe AEs. These events will be analysed univariately and as a composite endpoint. Time-to-event methods will be used for death and the composite endpoint. Each AE will be counted once for a given participant and graded by severity and relationship to COVID-19 or study intervention. AEs will be coded using the current version of the Medical Dictionary for Regulatory Activities (MedDRA). AEs will be presented by system organ class, duration (in days), start- and stop-date. Adverse events leading to premature discontinuation from the study intervention and serious treatment-emergent AEs should be presented either in a table or a listing.

##### 11.4.6. Baseline Descriptive Statistics

Baseline characteristics will be summarized by treatment arm. For continuous measures the mean and standard deviation will be summarized. Categorical variables will be described by the proportion in each category (with the corresponding sample size numbers).

##### 11.4.7. Planned Interim and Early Analyses

**Early analyses**

CONFIDENTIAL

Additional early analyses include monitoring enrolment, baseline characteristics, and follow-up rates throughout the course of the study by the study team. Analyses will be conducted blinded to treatment assignment.

**Interim analyses**

A data and safety monitoring board (DSMB) will monitor ongoing results to ensure patient well-being and safety as well as study integrity. It will also evaluate the ermergence of new variants in patients without viral load decrease at D8. The DSMB will be asked to communicate the results with the DSMC of Solidarity trial (only for treatment arms shared with Solidarity) when there is clear and substantial evidence of a treatment difference. More details about the interim analyses are described below as well as a separate guidance document for the DSMB. The composition and the charter of the DSMB will be communicated to the ANSM, as well as the minutes of each DSMB meeting.

**Interim Safety Analyses**

The AE / SAE data are reviewed every 2 weeks by the trial safety team. If there are a concerning number of unexpected AEs, the DSMB will be asked to review safety data in an *ad hoc* meeting.

In addition, the DSMB will review safety data after 100, 200 and 300 subjects are entered into the study, and then every 200 new patients are included, with a maximum of 1 DSMB meeting per week. Other *ad hoc* reviews will be undertaken if there are other specific safety concerns. The study will not stop enrolment awaiting these DSMB reviews, though the DSMB may recommend temporary or permanent cessation of enrolment based on their safety reviews.

No pre-specified stopping guideline will be defined because there are various aspects of potential harm that could be studied.

All the above is intended for all randomised patients.

An independent statistician will prepare these closed reports for DSMB review and recommendations. Analyses will be presented with blinded codes for treatment arms to protect against the possibility that the DSMB report may fall into the wrong hands. A DSMB charter will further describe procedures and membership. An additional document on statistical issues related to monitoring will be provided to the DSMB prior to interim analyses.

##### 11.4.8. Sub-Group Analyses

Analyses will be performed for both the primary efficacy and the key secondary efficacy endpoints, first for the Ag+ population second for the Ag+ population of

CONFIDENTIAL

not vaccinated patients, and third for the full (Ag+/Ag-) population, to evaluate the treatment effect across various subgroups. The following surbgroups will be studied by rapid antigen test result at inclusion (Ag+ vs Ag-, for the full population analysis), vaccination initiation (yes vs no, except for the analysis in non vaccinated patients), geographic region, clinical status at inclusion (7-point ordinal scale of 3 OR 4 vs 5), duration of symptoms prior to enrolment (<3 vs ≥ 3 days AND < 5 vs ≥ 5 days), age (< 50 vs [50;70[ vs ≥ 70), SARS-CoV-2 serology at inclusion (seronegative vs seropositive post infection vs seropositive post vaccination), SARS-CoV-2 lineage at inclusion (categories to be defined from blind analysis),and sex. For each population of analysis, a forest plot will display confidence intervals across subgroups. Interaction tests will be conducted to determine whether the effect of treatment varies by subgroup.

### 12. Supporting Documentation and Operational Considerations

#### 12.1. Regulatory, Ethical, and Study Oversight Considerations

This study will be conducted in conformity with the principles set forth in The Belmont Report: Ethical Principles and Guidelines for the Protection of Human Subjects of Research and the ICH E6(R2).

IRBs/ECs will review and approve this protocol, associated informed consent documents, recruitment material, and handouts or surveys intended for the subjects, prior to the recruitment, screening, and enrolment of subjects. Site IRBs may have additional national and local regulations.

Any amendments to the protocol or consent materials will be approved by the IRB/EC before they are implemented. IRB/EC review and approval will occur at least annually throughout the duration of the study. The investigator will notify the IRB/EC of deviations from the protocol and SAEs, as applicable to the IRB/EC policy.

The Sponsor must receive the documentation that verifies IRB/EC-approval for this protocol, informed consent documents, and associated documents prior to the recruitment, screening, and enrolment of subjects, and any IRB/EC-approvals for continuing review or amendments as required by the Sponsor.

##### 12.1.1. Informed Consent Process

Informed consent is a process that is initiated prior to an individual agreeing to participate in a trial and continuing throughout the individual’s trial participation. Investigators or designated research staff will obtain a subject’s informed consent

CONFIDENTIAL

in accordance with the national ethics and regulatory requirements and local regulations and policy, and ICH E6 GCP before any study procedures or data collection are performed.

Subjects will receive a concise and focused presentation of key information about the clinical trial, verbally and with a written consent form. The key information about the study will be organized and presented in lay terminology and language that facilitates understanding why one might or might not want to participate.

ICFs will be IRB/EC-approved, and subjects will be asked to read and review the consent form. Subjects (or legally authorize representatives) must sign the ICF prior to starting any study procedures being done specifically for this trial. Once signed, a copy of the ICF will be given to the subject for their records.

New information will be communicated by the site PI to subjects who consent to participate in this trial in accordance with IRB/EC requirements. The informed consent document will be updated, and subjects will be re-consented per IRB/EC requirements, if necessary.

**Requirements for Permission by Parents/Guardians and Assent by Children (in case of a minor)**

Not available

**Other Informed Consent Procedures**

Subjects may withdraw permission to use samples for secondary use at any time. They will need to contact the study site and the samples will be removed from the study repository after this study is completed and documentation will be completed that outlines the reason for withdrawal of permission for secondary use of samples.

##### 12.1.2. Study Termination and Closure

In Section 8, Study Intervention Discontinuation and Subject Discontinuation/Withdrawal, describes the temporary halting of the study.

This study may be prematurely terminated if there is sufficient reasonable cause, including but not limited to:

- Determination of unexpected, significant, or unacceptable risk to subjects
- Results of interim analysis
- Insufficient compliance to protocol requirements
- Data that are not sufficiently complete and/or not evaluable
- Regulatory authorities

CONFIDENTIAL

If the study is prematurely terminated, the site PI will promptly inform study subjects and the IRB/EC as applicable. The site PI will assure appropriate follow-up for the subjects, as necessary.

The sponsor will notify regulatory authorities as applicable.

##### 12.1.3. Confidentiality and Privacy

Subject confidentiality is strictly held in trust by the participating investigators, their staff, and the sponsor(s) and their agents. This confidentiality is extended to cover clinical information relating to subjects, test results of biological samples, and all other information generated during participation in the study. No identifiable information concerning subjects in the study will be released to any unauthorized third party. Subject confidentiality will be maintained when study results are published or discussed in conferences.

The study monitor, other authorized representatives of the sponsor, representatives of the IRB/EC, and/or regulatory agencies may inspect all documents and records required to be maintained by the investigator, including but not limited to, medical records (office, clinic, or hospital) and pharmacy records for the subjects in this study. The clinical study site will permit access to such records.

All source records including electronic data will be stored in secured systems in accordance with institutional policies and federal regulations.

All study data and research specimens that leave the site (including any electronic transmission of data) will be identified only by a coded number that is linked to a subject through a code key maintained at the clinical site. Names or readily identifying information will not be released unless the sponsor approves and it aligns with the consent form, or according to laws for required reporting.

##### 12.1.4. Secondary Use of Stored Specimens and Data

Secondary Human Subject Research is the re-use of identifiable data or identifiable biospecimens that were collected from some other ''primary'' or ''initial'' activity, such as the data and samples collected in this protocol. Any use of the sample or data for secondary research purposes, however, will be presented in a separate protocol and require separate IRB/EC approval.

Each sample will be labelled only with a barcode and a unique tracking number to protect subject confidentiality. Secondary research with coded samples and data may occur, however, subject confidentiality will be maintained as described

CONFIDENTIAL

for this protocol. An IRB/EC review of the secondary research using coded specimens is required.

The subject's decision can be changed at any time by notifying the study doctors or nurses in writing. If the subject subsequently changes his/her decision, the samples will be destroyed if the samples have not been used for research or released for a specific research project.

###### 12.1.5. Data Sharing for Secondary Research

Data from this study may be used for secondary research. All of the individual subject data collected during the trial will be made available after de-identification. The SAP and Analytic Code will also be made available. This data will be available immediately following publication, with no end date.

The investigator may request removal of data on individual study subjects from data repositories in the event that a research subject withdraws or changes his or her consent. However, some data that have been distributed for approved research use cannot be retrieved.

##### 12.2. Key Roles and Study Governance

Decisions related to the study will be made by a protocol team that includes representatives from all countries, and separate networks within a country.

###### 12.2.1. Safety Oversight

**Protocol team oversight**

The protocol team will review pools of AE data every 2 weeks to ensure there no significant number of unexpected AEs (AEs that do not fit with the known course of COVID-19). If there are a significant number of unexpected AEs, the DSMB will be asked to review unblinded safety data in an ad hoc meeting.

**Data Safety Monitoring Committee**

Safety oversight will be conducted (i) by a specific international independent DisCoVeRy DSMB that monitors subject safety treatment efficacyoutcomes specific to DisCoVeRy; (ii) by a Global international DSBM for the three outcomes shared with the WHO Solidarity protocol, for treatment arms shared with Solidarity. The DisCoVeRy DSMB members will be separate and independent of study personnel participating in this trial and should not have scientific, financial or other conflict of interest related to this study.

The DisCoVeRy DSMB will consist of members with appropriate expertise to contribute to the interpretation of the data from this trial. The DisCoVeRy I DSMB

CONFIDENTIAL

should be as broadly informed as possible regarding emerging evidence from related studies as well as from the conduct of this DisCoVeRy Protocol. The DisCoVeRy DSMB will operate under the guidelines of a charter that will be written at the organizational meeting of the DSMB. The DSMB will review SAEs on a regular basis and ad hoc during this trial. The Medical Monitor will be responsible for reviewing SAEs in real time. All SAR and SUSAR will be sent to the DSMB in real time for a safety review.

The interim trial results will be monitored by a Data Monitoring Committee. If good evidence emerges while the trial is continuing that some other treatment(s) should also be evaluated, then it will be decided that one or more extra arms will be added while the trial is in progress. All decisions will be made globally by the Executive Group of the International Steering Committee of Solidarity from information of its DSMC (for treatment arms shared with Solidarity).

The study will not stop enrolment awaiting these DSMB reviews, though the DSMB may recommend temporary or permanent cessation of enrolment based on their safety reviews.

Additional data may be requested by the DSMB, and interim statistical reports may be generated as deemed necessary. The DSMB may receive data in aggregate and presented by treatment arm. The DSMB will review grouped data in the closed session only. As an outcome of each review/meeting, the DSMB will make a recommendation as to the advisability of proceeding with study interventions (as applicable), and to continue, modify, or terminate this study.

##### 12.2.2. Clinical Monitoring

Clinical site monitoring is conducted to ensure that the rights and well-being of trial subjects are protected, that the reported trial data are accurate, complete, and verifiable. Clinical Monitoring also ensures conduct of the trial isin compliance with the currently approved protocol/ amendment(s), ICH, GCP, and with applicable regulatory requirement(s) and sponsor requirements. Clinical monitoring will also verify that any critical study procedures are completed following specific instructions in the protocol-specific MOP.

Monitoring for this study will be performed by Clinical Trial Unit or Contrat Research Organization. Details of clinical site monitoring are documented in a clinical monitoring plan (CMP). The CMP describes in detail who will conduct the monitoring, at what frequency monitoring will be done, at what level of detail monitoring will be performed, and the distribution of monitoring reports. Monitoring visits will include, but are not limited to, review of regulatory files, accountability records, CRFs, ICFs, medical and laboratory reports, site study

CONFIDENTIAL

intervention storage records, training records, and protocol and GCP compliance. Site monitors will have access to each participating site, study personnel, and all study documentation according to the sponsor-approved site monitoring plan. Study monitors will meet with site Pis to discuss any problems and outstanding issues and will document site visit findings and discussions.
Because DisCoVeRy is a trial involving COVID-19 treatment, remote Source Data Verification (SDV) will only be considered for site where temporary national emergency measures limit on-site monitoring and if in line with national law.
It will focus on the quality control of critical data (primary efficacy data and important safety data).

##### 12.2.3. Data Handling and Record Keeping

**Data Collection and Management Responsibilities**

Data collection is the responsibility of the study personnel at the participating clinical study site under the supervision of the site PI. The site PI must maintain complete and accurate source documentation.

Clinical research data from source documentation (including, but not limited to, AE/SAEs, concomitant medications, medical history, physical assessments, clinical laboratory data, some worksheet when dated and signed by investigator) will be entered by the clinical study site into CRFs via a 21 CFR Part 11-compliant internet data entry system. The data system includes password protection and internal quality checks, such as automatic range checks, to identify data that appear inconsistent, incomplete, or inaccurate. Aes and concomitant medications will be coded according to the most current versions of MedDRA and WhoDrug, respectively.

The clinical trial unit (APHP-DEBRC) for this study will be responsible for data management, quality review, analysis, and reporting of the study data.

The IND sponsor is responsible for review of data collection tools and processes, and review of data and reports.

AEs will be coded according to the MedDRA dictionary version 23.0 or higher.

A separate study specific Study Data Standardization Plan (SDSP) appendix will be developed which describes the technical recommendations for the submission of human study data and related information in a standardized electronic format throughout product development.

At the end of the study, a copy of all datasets including annotated CRFs and data dictionary will be provided to the sponsor.

CONFIDENTIAL

##### Study Record Retention

Study related records, including the regulatory file, study product accountability records, consent forms, subject source documents and electronic records should be maintained for a period of 2 years following the date a marketing application is approved for the investigational product for the indication for which it is being investigated; or, if no application is to be filed or if the application is not approved for such indication, until 2 years after the investigation These documents should be retained for a longer period, however, if required by local policies or regulations. No records will be destroyed without the written consent of the sponsor. Consent forms with specimen retention linked to identifiable specimens will be maintained for as long as the specimens remain in identifiable format, and a minimum of three years after use of the identifiable specimens in non-exempt human subject research.

##### Source Records

Source data are all information in original records (and certified copies of original records) of clinical findings, observations, or other activities in a clinical trial necessary for the reconstruction and evaluation of the trial. Source data should be attributable, legible, contemporaneous, original, accurate, and complete. Each participating site will maintain appropriate medical and research records for this trial, in compliance with ICH GCP, regulatory, and institutional requirements. Data recorded in the CRF derived from source documents should be consistent with the data recorded on the source documents.

Interview of subjects is sufficient for obtaining medical history. Solicitation of medical records from the subject's primary care provider is not required.

##### 12.2.4. Protocol Deviations

A protocol deviation is any noncompliance with the clinical trial protocol, any process that is noted in the protocol and refers to details in the protocol-specific SOP, or GCP requirements or any critical study procedures with specific instructions in ancillary documents referenced in the protocol such as a protocol-specific MOP.

The noncompliance may be either on the part of the subject, the investigator, or the study site staff. Following a deviation(s), corrective actions should be developed by the site and implemented promptly. All individual protocol deviations will be addressed in subject study records.

It is the responsibility of the site PI and personnel to use continuous vigilance to identify and report deviations within five working days of identification of the protocol deviation, or within five working days of the scheduled protocol-required

CONFIDENTIAL

activity. All deviations must be promptly reported per the protocol deviation reporting procedures. Protocol deviations must be sent to the IRB/EC per their guidelines. The site PI and personnel are responsible for knowing and adhering to their IRB/EC requirements. A completed copy of the Protocol Deviation Form must be maintained in the Regulatory File, as well as in the subject's chart if the deviation is subject specific.

##### 12.2.5. Data Sharing Policy

To avoid premature release of data, this protocol specifies that efficacy data from a trial that has not yet been completed due to insufficient enrolment should not be released. After an outbreak has ended at a given site, the study would be paused.

An independent monitoring committee would review results to make recommendations regarding whether the study should continue or stop for safety guided by the pre-specified *Data Safety Monitoring Board Charter*.

The study is registered on the clinicaltrials.gov site (NCT04315948).

Importantly, under this protocol, the investigators would remain blinded to any results of analyses; the study data would only be released if the trial were either stopped on the basis of a recommendation from the monitoring committee or had reached its targeted number of endpoints or amount of participant follow-up.

##### 12.2.6. Human Data Sharing Plan

See above.

##### 12.2.7. Publication

Following completion of the study, the lead PI is expected to publish the results of this research in a scientific journal, with consideration to the clarifications under section 10.1.10 above and all contracts signed for the trial.

##### 12.2.8. Conflict of Interest Policy

The independence of this study from any actual or perceived influence, such as by the pharmaceutical industry, is critical. Therefore, any actual conflict of interest of persons who have a role in the design, conduct, analysis, publication, or any aspect of this trial will be disclosed and managed. Furthermore, persons who have a perceived conflict of interest will be required to have such conflicts managed in a way that is appropriate to their participation in the design and conduct of this trial.

CONFIDENTIAL

### 13. Additional Considerations

#### 13.1. Research Related Injuries

For any potential research related injury, the site PI or designee will assess the subject.
Study personnel will try to reduce, control, and treat any complications from this study. Immediate medical treatment may be provided by the participating study site.

As needed, referrals to appropriate health care facilities will be provided to the subject. The site PI should then determine if an injury occurred as a direct result of the tests or treatments that are done for this study.

If it is determined by the participating site PI that an injury occurred to a subject as a direct result of the tests or treatments that are done for this trial, then referrals to appropriate health care facilities will be provided to the subject.

Study personnel will try to reduce, control and treat any complications from this trial. Immediate medical treatment may be provided by the participating site, such as giving emergency medications to stop immediate allergic reactions.

INSERM, as the sponsor, has taken out an insurance contract per country of civil responsibility, in accordance with local legal and regulatory guidelines.

#### 13.2. Abbreviations

| | |
|---|---|
| ADA | Anti-Drug_Antibodies |
| AE | Adverse Event |
| ALT | Alanine Transaminase |
| APHP-DEBRC | Assistance Publique – Hopitaux de Paris, Department of Epidemiology, Biostatistic and Clinical Research, Bichat Hospital DEBRC - France |
| AST | Aspartate Transaminase |
| BP | Blood Pressure |
| CFR | Code of Federal Regulations |
| CI | Confidence Interval |
| CLIA | Clinical Laboratory Improvement Amendments |
| CMP | Clinical Monitoring Plan |
| CMS | Clinical Material Services |
| Cr | Creatinine |
| CRF | Case Report Form |

CONFIDENTIAL

| | |
|---|---|
| CROMS | Clinical Research Operations and Management Support |
| CSR | Clinical Study Report |
| CQMP | Clinical Quality Management Plan |
| DSMB | Data Safety Monitoring Board |
| DSMC | Data Safety Monitoring Committee |
| EC | Ethics Committee |
| eCRF | Electronic Case Report Form |
| eTMF | Electronic Trial Master File |
| FDA | Food and Drug Administration |
| FWA | Federal Wide Assurance |
| GCP | Good Clinical Practice |
| GLP | Good Laboratory Practices |
| Hgb | Haemoglobin |
| HR | Heart Rate |
| IB | Investigator's Brochure |
| ICD | International Classification of Diseases |
| ICF | Informed Consent Form |
| ICH | International Council for Harmonisation |
| IFN-ß-1a | Interferon beta-1a |
| IMP | Investigational Medicinal Product |
| IND | Investigational New Drug Application |
| IRB | Institutional Review Board |
| IV | Intravenous |
| IWRS | Interactive Web Response System |
| MCG | Microgram |
| MedDRA | Medical Dictionary for Regulatory Activities |
| MERS | Middle East Respiratory Syndrome |
| MOP | Manual of Procedures |
| N | Number (typically refers to subjects) |
| NDA | New Drug Application |
| NP | Nasopharyngeal |
| PHI | Protected Health Information |
| PI | Principal Investigator |
| PK | Pharmacokinetics |
| PLT | Platelet |
| PP | Per Protocol |
| PT | Prothrombin Time |
| RBD | Receptor Binding Domain |
| SAE | Serious Adverse Event |
| SAP | Statistical Analysis Plan |
| SARS | Severe Acute Respiratory Syndrome |

CONFIDENTIAL

| | |
|---|---|
| SDCC | Statistical and Data Coordinating Centre |
| SDSP | Study Data Standardization Plan |
| SNP | Single Nucleotide Polymorphisms |
| SOA | Schedule of Assessments |
| SOC | System Organ Class |
| SOP | Standard Operating Procedure |
| SUSAR | Suspected Unexpected Serious Adverse Reaction |
| T. Bili | Total Bilirubin |
| UP | Unanticipated Problem |
| US | United States |
| WBC | White Blood Cell |
| WHO | World Health Organization |

CONFIDENTIAL

#### 13.4. Protocol Amendment History

| Version | Date | Description of Change | Brief Rationale |
|---|---|---|---|
| 5.0 | 22.03.2020 | Hydroxychloroquine arm implementation | |
| 7.0 | 05.04.2020 | DisCoVeRy is an add-on trial of the Solidarity Trial | |
| 8.0 | 23.06.2020 | Hydroxychloroquine arm stopped | |
| 9.0 | 29.06.2020 | Lopinavir/Ritonavir and Lopinavir/Ritonavir + IFN arms stopped | |
| 10.0 | 01.10.2020 | Corticosteroids and anticoagulants added to SoC.<br>5-year multi-national 'European Research and Preparedness Network for Pandemics and Emerging Infectious Diseases' — EU-RESPONSE project will allow the rapid European expansion and further evolution of the DisCoVeRy study and build a new multinational European Adaptive Platform Trial for emerging infectious diseases in general. | |
| 11.0 | 08.12.2020 | Update of the sample size determination. | |
| 12.0 | 09.03.2021 | Remdesivir arm stopped, AZD7442 arm implementation | |
| 13.0 | 29.03.2021 | Update of background and exclusion criteria as requested by the French Ethics Committee | |
| 14.0 | 19.04.2021 | Updates requested by French competent authority | |
| 15.0 | 20.09.2021 | Inclusion of vaccinated patients. Extended period since symptom onset | |

CONFIDENTIAL

CONFIDENTIAL

### 15. References

1. Loconsole D, Stea ED, Sallustio A, et al. Severe COVID-19 by SARS-CoV-2 Lineage B.1.1.7 in Vaccinated Solid-Organ Transplant Recipients: New Preventive Strategies Needed to Protect Immunocompromised Patients. *Vaccines*. 2021;9(8):806. doi:10.3390/vaccines9080806
2. Rabinowich L, Grupper A, Baruch R, et al. Low immunogenicity to SARS-CoV-2 vaccination among liver transplant recipients. *Journal of Hepatology*. 2021;75(2):435-438. doi:10.1016/j.jhep.2021.04.020
3. Soiza RL, Scicluna C, Thomson EC. Efficacy and safety of COVID-19 vaccines in older people. *Age and Ageing*. 2021;50(2):279-283. doi:10.1093/ageing/afaa274
4. RECOVERY Collaborative Group, Horby PW, Mafham M, et al. *Casirivimab and Imdevimab in Patients Admitted to Hospital with COVID-19 (RECOVERY): A Randomised, Controlled, Open-Label, Platform Trial*. Infectious Diseases (except HIV/AIDS); 2021. doi:10.1101/2021.06.15.21258542
5. Chan JF-W, Yuan S, Kok K-H, et al. A familial cluster of pneumonia associated with the 2019 novel coronavirus indicating person-to-person transmission: a study of a family cluster. *The Lancet*. 2020;395(10223):514-523. doi:10.1016/S0140-6736(20)30154-9
6. Wu JT, Leung K, Leung GM. Nowcasting and forecasting the potential domestic and international spread of the 2019-nCoV outbreak originating in Wuhan, China: a modelling study. *Lancet*. 2020;395(10225):689-697. doi:10.1016/S0140-6736(20)30260-9
7. HCSP. Covid-19 : utilisation de l'hydroxychloroquine. Accessed January 4, 2021. https://www.hcsp.fr/Explore.cgi/Telecharger?NomFichier=hcspa20200524_covidutilidelhydro.pdf
8. NIH. Management of COVID-19 | Coronavirus Disease COVID-19. COVID-19 Treatment Guidelines. https://www.covid19treatmentguidelines.nih.gov/overview/management-of-covid-19/
9. Al-Kofahi M, Jacobson P, Boulware DR, et al. Finding the Dose for Hydroxychloroquine Prophylaxis for COVID-19: The Desperate Search for Effectiveness. *Clin Pharmacol Ther*. 2020;108(4):766-769. doi:10.1002/cpt.1874
10. Rosenberg ES, Dufort EM, Udo T, et al. Association of Treatment With Hydroxychloroquine or Azithromycin With In-Hospital Mortality in Patients With COVID-19 in New York State. *JAMA*. 2020;323(24):2493-2502. doi:10.1001/jama.2020.8630
11. Hernandez AV, Roman YM, Pasupuleti V, Barboza JJ, White CM. Hydroxychloroquine or Chloroquine for Treatment or Prophylaxis of COVID-19: A Living Systematic Review. *Annals of Internal Medicine*. 2020;173(4):287-296. doi:10.7326/M20-2496
12. Paccoud O, Tubach F, Baptiste A, et al. Compassionate Use of Hydroxychloroquine in Clinical Practice for Patients With Mild to Severe COVID-19 in a French University Hospital. *Clinical Infectious Diseases*. Published online June 18, 2020:ciaa791. doi:10.1093/cid/ciaa791
13. Torjesen I. Covid-19: Hydroxychloroquine does not benefit hospitalised patients, UK trial finds. *BMJ*. Published online June 8, 2020:m2263. doi:10.1136/bmj.m2263
14. Weinreich DM, Sivapalasingam S, Norton T, et al. REGN-COV2, a Neutralizing Antibody Cocktail, in Outpatients with Covid-19. *N Engl J Med*. 2021;384(3):238-251. doi:10.1056/NEJMoa2035002
15. Wang P, Nair MS, Liu L, et al. *Antibody Resistance of SARS-CoV-2 Variants B.1.351 and B.1.1.7*. Immunology; 2021. doi:10.1101/2021.01.25.428137

CONFIDENTIAL

16. Zost SJ, Gilchuk P, Case JB, et al. Potently neutralizing and protective human antibodies against SARS-CoV-2. *Nature*. 2020;584(7821):443-449. doi:10.1038/s41586-020-2548-6
17. Dexamethasone in Hospitalized Patients with Covid-19 — Preliminary Report. *New England Journal of Medicine*. 2020;0(0):null. doi:10.1056/NEJMoa2021436
18. Villar J, Ferrando C, Martínez D, et al. Dexamethasone treatment for the acute respiratory distress syndrome: a multicentre, randomised controlled trial. *Lancet Respir Med*. 2020;8(3):267-276. doi:10.1016/S2213-2600(19)30417-5
19. Tomazini BM, Maia IS, Cavalcanti AB, et al. Effect of Dexamethasone on Days Alive and Ventilator-Free in Patients With Moderate or Severe Acute Respiratory Distress Syndrome and COVID-19: The CoDEX Randomized Clinical Trial. *JAMA*. 2020;324(13):1307. doi:10.1001/jama.2020.17021
20. The WHO Rapid Evidence Appraisal for COVID-19 Therapies (REACT) Working Group, Sterne JAC, Murthy S, et al. Association Between Administration of Systemic Corticosteroids and Mortality Among Critically Ill Patients With COVID-19: A Meta-analysis. *JAMA*. 2020;324(13):1330. doi:10.1001/jama.2020.17023
21. CRICS TRIGGERSEP Group (Clinical Research in Intensive Care and Sepsis Trial Group for Global Evaluation and Research in Sepsis), Helms J, Tacquard C, et al. High risk of thrombosis in patients with severe SARS-CoV-2 infection: a multicenter prospective cohort study. *Intensive Care Med*. 2020;46(6):1089-1098. doi:10.1007/s00134-020-06062-x
22. Ranucci M, Ballotta A, Di Dedda U, et al. The procoagulant pattern of patients with COVID-19 acute respiratory distress syndrome. *J Thromb Haemost*. 2020;18(7):1747-1751. doi:10.1111/jth.14854
23. Connors JM, Levy JH. Thromboinflammation and the hypercoagulability of COVID-19. *J Thromb Haemost*. 2020;18(7):1559-1561. doi:10.1111/jth.14849
24. Tang N, Bai H, Chen X, Gong J, Li D, Sun Z. Anticoagulant treatment is associated with decreased mortality in severe coronavirus disease 2019 patients with coagulopathy. *J Thromb Haemost*. 2020;18(5):1094-1099. doi:10.1111/jth.14817
25. Paranjpe I, Fuster V, Lala A, et al. Association of Treatment Dose Anticoagulation With In-Hospital Survival Among Hospitalized Patients With COVID-19. *J Am Coll Cardiol*. 2020;76(1):122-124. doi:10.1016/j.jacc.2020.05.001
26. Spyropoulos AC, Levy JH, Ageno W, et al. Scientific and Standardization Committee communication: Clinical guidance on the diagnosis, prevention, and treatment of venous thromboembolism in hospitalized patients with COVID-19. *J Thromb Haemost*. 2020;18(8):1859-1865. doi:10.1111/jth.14929
27. Bai C, Chotirmall SH, Rello J, et al. Updated guidance on the management of COVID-19: from an American Thoracic Society/European Respiratory Society coordinated International Task Force (29 July 2020). *Eur Respir Rev*. 2020;29(157). doi:10.1183/16000617.0287-2020
28. Smith GB, Prytherch DR, Meredith P, Schmidt PE, Featherstone PI. The ability of the National Early Warning Score (NEWS) to discriminate patients at risk of early cardiac arrest, unanticipated intensive care unit admission, and death. *Resuscitation*. 2013;84(4):465-470. doi:10.1016/j.resuscitation.2012.12.016
29. Whitehead J. Sample size calculations for ordered categorical data. *Statist Med*. 1993;12(24):2257-2271. doi:10.1002/sim.4780122404

CONFIDENTIAL

**Multi-centre, adaptive, randomized trial of the safety and efficacy of treatments of COVID-19 in hospitalized adults**

### Statistical analysis plan for primary and secondary endpoints for version 15 and beyond of the protocol (evaluation of AZD7442)

**CONFIDENTIAL**

#### Signatures

| RESPONSIBILITY | NAME | SIGNATURE | DATE |
|---|---|---|---|
| **Statistician** | Lien Han | | 2023-01-09 |
| **Statistician** | Toni Alfaiate | | 2023-01-09 |
| **Methodologist** | Clément Massonnaud | | 2023-01-09 |
| **Chief methodologist** | France Mentré | | 2023-01-09 |
| **DisCoVeRy Trial steering committee** | Dominique Costagliola | | 2023-01-12 |

#### Revision history

| VERSION NO. | EFFECTIVE DATE | DESCRIPTION |
|---|---|---|
| 1.0 | 29-08-2021 | First version (protocol V14) |
| 2.0 | 25-11-2021 | Second version (protocol V15) |
| 3.0 | 09-01-2023 | Amendments after early stop of inclusions |

#### List of abbreviations

| ABBREVIATION | DESCRIPTION |
|---|---|
| AE | Adverse event |
| ALT | Alanine aminotransferase |
| AST | Aspartate aminotransferase |
| BMI | Body mass index |
| CRP | C-reactive protein |
| DAIDS | Division of AIDS |
| DSMB | Data and Safety Monitoring Board |
| DSMC | Data and Safety Monitoring Committee |
| ECMO | Extra-corporeal membrane oxygenation |
| INR | International normalized ratio |
| ITT | Intention-to-treat |
| MedDRA | Medical Dictionary for Regulatory Activities |
| mITT | Modified intention-to-treat |
| NEWS | National Early Warning Score |
| OR | Odds ratio |
| PCT | Procalcitonin |
| SAE | Serious adverse event |
| SAP | Statistical analysis plan |
| SGOT | Serum glutamic-oxalacetic transaminase |
| SGPT | Serum glutamic-pyruvic transaminase |
| SoC | Standard of Care |

5.1. Patient disposition ... 21
5.2. Protocol violations ... 23
6. Demographics and other baseline characteristics ... 23
6.1. Descriptive statistics ... 23
6.2. Demographic data ... 23
6.3. Medical and surgical history ... 24
6.4. Clinical evaluation ... 24
6.5. Biological data ... 24
6.6. Chest CT data ... 25
6.7. Previous and concomitant treatment ... 25
6.8. Study treatment ... 27
7. Analysis ... 27
7.1. Analysis of the primary endpoint ... 27
7.2. Analysis of key secondary endpoint ... 28
7.3. Analysis of the secondary endpoints ... 28
7.3.1. Ordinal scale ... 28
7.3.2. National Early Warning Score 2 (NEWS-2) ... 29
7.3.3. Oxygenation ... 29
7.3.4. Mechanical ventilation ... 30
7.3.5. Hospitalization ... 30
7.3.6. Mortality ... 31
7.3.7. Long term health status ... 31
7.3.8. Safety ... 32
8. Sensitivity analysis ... 33
9. Subgroup analyses ... 33
10. Appendix: ... 34
10.1. Power computation ... 34
10.2. NEWS-2 scoring system ... 34
References ... 36

### 1. Overview of the study

#### 1.1. Introduction

This document summarizes the endpoints and the statistical methods that will be used to analyse the DisCoVeRy data for primary and secondary endpoints. Specific documents will be prepared for exploratory endpoints.

The statistical analyses will be performed after two global database locks if the trial is not modified: the first after all patients have reached D90, for the primary, key secondary, and other efficacy and safety analyses available at day 90; and the second after all patients have completed the last study visit, for all planned analyses.

This Statistical Analysis Plan (SAP) details the different populations of analysis and the statistical methodology that will be used to analyze the data. It describes the efficacy and safety variables and anticipated data transformations and manipulations, and other details of the analyses not provided in the study protocol.

The analyses described are based upon the clinical study protocol V15.0 dated September 20, 2021. This new version of the Statistical Analysis Plan has been edited following the decision by the Discovery Steering Committee on June 23, 2022 to stop the inclusions on July 1st, 2022 due to a low rate of inclusion. It is therefore adapted to account for a smaller sample size than initially planned.

#### 1.2. Study objectives

##### 1.2.1. Primary objective

The primary objective of the study is to evaluate the clinical efficacy of the investigational product compared to the placebo arm in patients hospitalized with COVID-19 using a 7-point ordinal scale assessing clinical status at Day 15.

##### 1.2.2. Key Secondary objective

The key secondary objective is to evaluate the efficacy of the investigational product on sustained recovery from index COVID-19 infection.

##### 1.2.3. Secondary objectives

**Clinical efficacy**

To evaluate the clinical efficacy of the investigational product as compared to the control arm as assessed by:

- Ordinal scale
    - Subject clinical status on an ordinal scale on Days 29, 90, 180, and 365.
- National Early Warning Score (NEWS)

Change from baseline to Days 3, 8, 15 (if patient is still hospitalized during these 3 visits), and 29 in NEWS-2.

- Oxygenation
    - Oxygenation free days in the first 28 days (to Day 29).

  - Incidence and duration of new oxygen use, non-invasive ventilation or high flow oxygen devices during the first 28 days (to Day 29).
- Mechanical ventilation
  - Ventilator free days during the first 28 days (to Day 29).
  - Incidence and duration of invasive mechanical ventilation use during the first 28 days (to Day 29).
  - Need for mechanical ventilation or death by Day 15.
- Hospitalization
  - Time to hospital discharge from randomization.
- Mortality
  - In-hospital mortality
  - 29-day mortality.
  - 90-day mortality
  - 180-day mortality
  - 365-day mortality
  - 456-day mortality
- Long term health status
  - Any hospitalization between discharge from index hospitalization and Days 90, 180 and 365.
  - Any confirmed re-infection with SARS-CoV2 between discharge from index hospitalization and Days 90, 180 and 365.

**Safety**

To evaluate the safety of different investigational therapeutics through 456 days of follow-up as compared to the control arm as assessed by:

- Cumulative incidence of serious adverse events (SAEs)
- Cumulative incidence of Grade 3 and 4 adverse events (AEs).
- Cumulative incidence of Grade 1-2 hypersensitivity related and infusion related AEs until D29 visit
- Cumulative incidence of AEs of Special Interest
- Discontinuation or temporary suspension of investigational product (for any reason)

#### 1.3. Study endpoints

##### 1.3.1. Primary endpoint

The primary endpoint is the clinical status at Day 15 on a 7-point ordinal scale:

1. Not hospitalized, no limitation on activities;
2. Not hospitalized, limitation on activities;
3. Hospitalized, not requiring supplemental oxygen;
4. Hospitalized, requiring supplemental oxygen;

5. Hospitalized, on non-invasive ventilation or high flow oxygen devices;
6. Hospitalized, on invasive mechanical ventilation or ECMO;
7. Death.

###### 1.3.2. Key secondary endpoint

The key secondary endpoint is the time from randomization to sustained recovery, defined as being discharged from the index hospitalization, followed by being alive and at home for 14 consecutive days prior to Day 90. A detailed description of sustained recovery including the definition of home is presented in section 10.2.3 of the trial protocol (V15.0 dated September 20, 2021).

###### 1.3.3. Secondary endpoints

**Clinical efficacy**

- Status on an ordinal scale assessed at Day 29, 90, 180 and 365
- NEWS-2 at baseline (Day 1 pre-treatment), at Days 3, 8 and 15 (if patient is still hospitalized during these 3 visits), and at Days 29.
- Duration of supplemental oxygen (if applicable).
- Duration of mechanical ventilation (if applicable).
- Date of discharge from hospital.
- Date and cause of death (if applicable).
- Mechanical ventilation or death between baseline and Day 15.
- Occurrence of new hospitalization between discharge from index hospitalization and Days 90, 180 and 365.
- Occurrence of confirmed re-infection with SARS-CoV-2 between discharge and Days 90, 180 and 365.

**Safety**

- SAEs.
- Grade 3 and 4 of adverse events.
- Grade 1-2 hypersensitivity-related and infusion related AEs until D29 visit.
- AE of special interest.
- Discontinuation of investigational product (for any reason).

#### 2. Study methodology

##### 2.1. Overall study design

This is a European prospective, multicentre, adaptive, randomized, double blinded clinical trial that randomly allocates participants (1:1 ratio) between 2 arms: Standard of Care (SoC) + placebo versus SoC + AZD7442.

##### 2.2. Treatments received

Patients will be randomly allocated between 2 arms SoC+ placebo versus SoC + AZD7442 (through IV infusion).

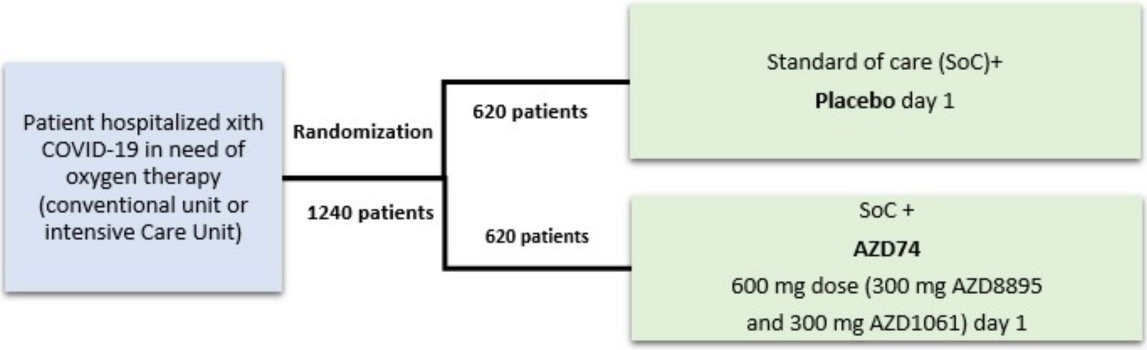

*Figure 1: DisCoVeRy trial arms, drugs and dosing schedule*

##### 2.3. Method of treatment assignment and randomization

The randomization procedure is managed and carried out by Vennlife Sciences. A detailed description of the randomization procedure is provided in the Randomization SOP prepared by VennLife Sciences[1]. The randomization will be stratified by :

- Antigenic status: positive or negative (obtained from the result of a rapid antigen test on nasopharyngeal swab performed at enrolment).
- Region (of each country included in the trial).
- Vaccination status: vaccination initiation yes or no (vaccination is initiated if at least one injection of any vaccine against SARS-CoV-2 was received prior to enrolment, whatever the delay).

#### 2.4. Study schedule

| Day +/- Window | Screening[1] | Baseline (Day 1)[2] | 2-14 | 15[3] ±2 | 29[4] ±3 | 90[4] ±7 | 180[5] ±14 | 365[5] ±14 | 456[5] ±14 |
|---|---|---|---|---|---|---|---|---|---|
| **ELIGIBILTY** | | | | | | | | | |
| Eligibility Criteria | X | | | | | | | | |
| Informed consent | X | | | | | | | | |
| Review SARS-CoV-2 results | X | | | | | | | | |
| Rapid Antigen test | X | | | | | | | | |
| **STUDY INTERVENTION** | | | | | | | | | |
| Randomization[12] | | X | | | | | | | |
| Standard of Care (SoC) + Placebo | | Administration of SoC according to guidelines[13] Single administration of placebo at day 1 | Administration of SoC according to guidelines | | | | | | |
| Or SoC + AZD7442 | | Administration of SoC according to guidelines[13] Single administration of AZD7442 at day 1 | Administration of SoC according to guidelines | | | | | | |
| **STUDY PROCEDURES** | | | | | | | | | |
| Demographics and Merdical History | X | | | | | X[6] | X[6] | X[6] | X[6] |
| Vital status | | | Daily while hospitalized | X | X | X | X | X | X |
| Medication review | X | | Daily while hospitalized[11] | X | X | X[14] | X[14] | X[14] | X[14] |
| NEWS-2 | | X | Days 3, 8 (all ± 1 day) if Hospitalized | X | X | | | | |
| Ventilation mode | | X | Daily while hospitalized[11] | X | X | | | | |
| Oxygenation | | X | Daily while hospitalized[11] | X | X | | | | |
| Ordinal scale | | X | | X | X | X | X | X | |
| Thoracic CT scan | | X | | | | | | | |
| Adverse event evaluation | X | X | Daily while hospitalized | X | X | X | X | X | X |
| **SAFETY LABORATORY** | | | | | | | | | |
| Safety biological and inflammatory tests[7,8] | X[9] | | Day 3, 8 (all ± 1 day) if hospitalized | X | X | X | X[10] | X[10] | |
| Urinary pregnancy test for females of childbearing Potential | X[9] | | | X | X | X | X[10] | X[10] | |

| | | | | | | | | |
|---|---|---|---|---|---|---|---|---|
| Qualitative (Ct value) PCR for SARS-CoV-2 in NP swab or lower respiratory tract sample | | X[15] | Day 8 (± 1 day) if Hospitalized[16] | | | | | |
| **RESEARCH LABORATORY** | | | | | | | | |
| Serum concentration of AZD7442 | | X (before and at end of infusion) | Days 1 (end of infusion) Days 3, 8 (all ± 1 day) if hospitalized | X | X | X | X 1 0 | X[10] |
| Serum sample for AZD7442 ADA Assessment | | X (before infusion) | | X | X | X | X 1 0 | X[10] |
| Serum for exploratory objectives | | X | Days 3, 8 (all ± 1 day) if hospitalized | X | X | X | X 1 0 | X[10] |
| Plasma for exploratory Objectives | | X | Days 3, 8 (all ± 1 day) if hospitalized | X | X | X | X 1 0 | X[10] |
| Whole blood for genetic analysis[18] | | X | | | | | | |
| Nasopharyngeal swab and/or lower respiratory tract samples (for patients in the Intensive Care Unit) for exploratory objectives | | X | Days 3, 8 (all ± 1 day) if hospitalized On the day of transfer into ICU, if applicable | X | X | | | |

*Notes:*

1. *Refer to Section 10.1 of the protocol for details of data to be collected at screening.*
2. *Baseline assessments will be performed prior to study drug administration (Day 1 pre-treatment)*
3. *Data will be collected only during hospitalization, except for the ordinal scale which will be collected by phone for discharged patients.*
4. *If discharged from the hospital, visits will be conducted in the outpatient setting.*
5. *By phone call or during a medical consultation*
6. *Information on re-hospitalization and SARS-CoV-2 reinfection*
7. *Creatinine; Liver enzymes and hepatic function (LDH, ALT, AST); cell counts (neutrophils and total lymphocyte counts, haemoglobin, platelets); coagulation and inflammation status (fibrinogen, D-Dimer, prothrombin time, C-reactive protein, ferritinemia, serum total immunoglobulins G. Serum total immunoglobulins G assessed only at screening)*
8. *Any laboratory tests performed as part of routine clinical care within the specified visit window can be used for safety laboratory testing.*
9. *Laboratory tests performed in the 36 hours prior to enrolment will be accepted.*
10. *During a medical consultation, in a subset of 25% of enrolled participants.*
11. *In hospitalized patients, medication review, oxygenation and ventilation mode will be collected up to day 29.*
12. *Randomization will be done in the 6 hours following rapid antigen testing.*
13. *SoC should be administered on day 1 in the 24 hours following randomization.*
14. *Concomitant medications will be captured based on potential relatedness to AEs post D29 visit.*
15. *Ct value and lineage of the virus performed during hospitalization and before infusion will be accepted.*
16. *Qualitative (Ct value) for SARS-CoV-2 in NP swab or lower respiratory tract sample if patient hospitalized at D8*
17. *If the participant agrees to the host genome analysis.*

#### 3. Sample size determination

The sample size calculations were conducted for a two arms trial with both arms assumed to continue until the end of the study.

The odds ratio represents the odds of improvement in the ordinal scale for treatment relative to control [Whitehead, 1993] shows that the sample size to detect a given odds ratio for 1:1 randomization using a 2-tailed test at level α is given by

$$\frac{12(z_{\alpha/2}+z_{\beta})^2}{\theta^2(1-\sum_{i=1}^{6} p_i^3)}$$

where θ is the log odds ratio, $p_i$ is the overall probability (combined over both arms) of being in the ith category of the ordinal outcome, and $z_{\alpha/2}$ and $z_{\beta}$ are the $1-\alpha/2$ and βth quantiles of the standard normal distribution.

The table below displays the outcomes under standard of care used for sample size determination. These were obtained from patients included in DisCoVery trial from July 1st to December 31, 2020, with results available in the database locked on January 6, 2021 for the DSMB. More precisely, D15 outcomes of 93 patients from the SOC arm, included in the moderate disease strata and with duration of symptoms of less of 9 days are reported and used.

| **Severity Outcome** | **Anticipated scenario**<br><br>outcome (%) |
|---|---|
| Death | 5 |
| Hospitalized, on mechanical ventilation or ECMO | 6 |
| Hospitalized, on non-invasive ventilation or high flow oxygen devices | 6 |
| Hospitalized, requiring supplemental oxygen | 17 |
| Hospitalized, not requiring supplemental oxygen | 7 |
| Not hospitalized, limitation on activities | 35 |
| Not hospitalized, no limitations on activities | 24 |

The targeted strength of evidence is 2-sided p=0.05, which corresponds to a (one-sided) false positive error rate of 0.025.

Since a large proportion of subjects are moderately ill patients, we have powered the study for an odds ratio of 1.5. For a power of 90%, we would need 413 patients per arm with Ag+ results.

Despite the changes in the inclusion/exclusion criteria, we decided not to change the number of subjects needed.

For the delay of symptoms before inclusion from 9 to 11 days, as the number of subjects was computed for the primary analysis on only positive antigen patients it will have no impact.

The inclusion new strata of vaccinated (fully or partly) patients could have an impact on the statistical power for an unchanged number of subjects, depending:

- On the distribution of the outcome at day 15 in vaccinated patients receiving SoC,
- On the expected treatment effect in vaccinated patients,
- On the proportion of patients vaccinated.

We decided to cap the proportion of vaccinated patients to 20% of antigen positive patients.

As can be seen in the table (reported in appendix) which presents power calculation, for a sample size of 413 per arm with Ag+ results, for various assumptions on treatment effect and outcome at day 15 in vaccinated versus non-vaccinated patients, the power is always greater than 85%, hence the decision not to change the number of subjects to be included.

The number of evaluable Ag- patients will be limited to at most 30% of the total number of patients, hence at most 177 Ag- patients per arm. At most 20% of vaccinated Ag- patients will be included.

- The number of evaluable patients per arm is thus 590. Therefore, we need to include 620 patients per arm (434 Ag+ and 186 Ag-) to account for 5% not evaluable patients (informed consent retracted, not valid,…).

### 4. General considerations

#### 4.1. Timing of analyses and database lock

Two analyses and database locks are planned:

- The primary (Prima) analysis will be performed after the first database lock once all patients have reached their D90 visit. This analysis will be conducted on the primary and key secondary endpoint, and other efficacy and safety endpoints available at day 90.

- The final analysis will be performed after the second and final database lock once all patients have completed the last study visit. It will cover all planned analysis including the other secondary and exploratory endpoints.

- Tables and Figures of each analysis will be reported first using the dummy randomization list in case some modifications are needed in the code (that will be tracked). Then the true randomization list will be used to report the results.

#### 4.2. Statistical software

Statistical analyses will be performed using SAS® version 9.4 (SAS Institute Inc., Cary, NC, USA). RStudio (2018, RStudio: Integrated Development for R) version 1.3.1335 (RStudio, Inc., Boston, MA, USA) powered with R software (version 4.0.3) will be used for graphical representation of selected analyses.

#### 4.3. Adaptive design

If good evidence emerges while the trial is continuing that some other treatment(s) should also be evaluated, then it will be centrally decided that one or more extra arms may be added while the trial is in progress.

#### 4.4. Type-I error rate

A two-sided Type-I error rate of 5% will be used.

#### 4.5. Interim analysis

There will be no pre-planned interim analysis for efficacy nor futility. If good evidence emerges while the study is continuing that some other treatment(s) should also be evaluated, then it will be decided that one or more extra arms may be added while the trial is in progress. All those decisions will be made globally by the Executive Group of the International Steering Committee of Solidarity from information of its DSMC.

A global independent data and safety monitoring board (DSMB) monitors interim data to make recommendations about early study closure or changes to conduct.

The AE / SAE data are reviewed every 2 weeks by the trial safety team. If there are a concerning number of unexpected AEs, the DSMB will be asked to review safety data. In addition, the DSMB will review safety data after 100 subjects are entered into the study, and then every 200 new patients are included, with a maximum of 1 DSMB meeting per week. Other ad hoc reviews will be undertaken if there are other specific safety concerns.
No pre-specified stopping guideline will be defined because there are various aspects of potential harm that could be studied. Given the double blinded nature of the trial, an independent statistician will prepare these closed reports for DSMB review and recommendations. Analyses will be presented with blinded codes for treatment arms to protect against the possibility that the DSMB report may fall into the wrong hands.

##### 4.6. Definition of analysis sets

For the two planned analyses described in section 4.1, the following analysis sets are defined:
- Positive intention-to-treat (Positive ITT): all randomized patients with positive SARS-CoV-2 antigenic status at randomization who will be analysed in their randomization group.
- Global intention-to-treat (Global ITT): all randomized patients with positive or negative SARS- CoV-2 antigenic status at randomization who will be analysed in their randomization group.
- Positive modified intention-to-treat (Positive mITT): patients of the Positive ITT in whom the allocated treatment was initiated.
- Global modified intention-to-treat (Global mITT): patients of the Global ITT in whom the allocated treatment was initiated.
- Positive Safety analysis set: all randomized patients with positive SARS-CoV-2 antigenic status at randomization and analysed in their actual treatment group.
- Global Safety analysis set: all randomized patients with positive or negative SARS-CoV-2 antigenic status at randomization and analysed in their actual treatment group.

##### 4.7. Baseline definition

Baseline data corresponds to demographic data, medical, surgical and treatment history. Regarding clinical and biological evaluation at baseline, it corresponds to the worst value for each component in the 24 hours (maximum) prior to the first intake of the study treatment

##### 4.8. Handling of missing data

The number, timing and pattern for missing value will be detailed for each outcome measure.

***a. Time to event outcomes***

- ***Time to sustained recovery***

Given that time to sustained recovery will be analysed using competing risk analysis with death as the competitive risk, only patients who are lost to follow up or those who withdraw from the study will be censored at their withdrawal or lost to follow up date.

- ***Incidence and duration of new oxygen use***

Patients who are not on oxygen, nor on mechanical ventilation, nor on ECMO and are: (i) lost to follow- up or (ii) withdrew from the trial and (iii) did not experience these events will be censored at their lost to follow-up or withdrawal date. Other patients who did not experience these events and still alive at D29 will be censored at day 29.

- ***Incidence and duration of invasive mechanical ventilation or ECMO use***

Patients who are not on mechanical ventilation nor on ECMO and are: (i) lost to follow-up or (ii) withdrew from the trial and (iii) did not experience these events will be censored at their lost to follow- up or withdrawal date. Other patients who did not experience these events and still alive at D29 will be censored at day 29.

- ***Time to discharge from hospital***

Given that time to discharge from hospital will be analysed using competing risk analysis with death as the competitive risk, only patients who are lost to follow up or those who withdraw from the study will be censored at their withdrawal or lost to follow up date.

***b. Other type of outcomes***

_Ordinal scale_

Missing data for the ordinal scale will be accounted for as follows:

- Death:
  - If the last scale value available before the missing data for a patient is “7. Death”, this value will be carried forward through day 365
  - If the last scale value available before the missing data for a patient is not “7. Death” but a death date is available, the value “7. Death” will be carried forward from the death date through day 365
- Non-hospitalized patients (without or before death date):
  - If the last scale value available before the missing data is “1. Not hospitalized, no limitations on activities” or “2. Not hospitalized, limitation on activities” and without new re-hospitalization between the date of the last available scale value and the date of the missing scale value, this last available scale value will be carried forward (until the next non-missing scale value, if values are available afterwards).
  - If the last scale value available before the missing data for a patient is not one of the two mentioned above (ie not a scale 1 or 2) but a discharge date is available and no new re-hospitalization is reported between discharge and the study visit for which the scale value is missing:
    - the last scale value available will be used on the discharge date (if the scale is missing at this time).
    - the value “2. Not hospitalized, limitation on activities” will be used from the discharge date + one day (until the next non-missing scale value, if values are available afterwards).
  - If the patient is re-hospitalized and no ordinal scale is reported for the re- hospitalization period, the worst scale value recorded for the patient during the index hospitalization will be used until the end of the re-hospitalization period.
- Hospitalized patients (without or before death or discharge date):

  - If the patient is reported as being on "room air" on the time of the missing scale value, then the value "3. Hospitalized, not requiring supplemental oxygen" will be used (until the next non-missing scale value, if values are available afterwards; or until the discharge date if a discharge date is available).
  - If the patient is reported as receiving Standard oxygenotherapy on the time of the missing scale value, then the value "4. Hospitalized, requiring supplemental oxygen" will be used (until the next non-missing scale value, if values are available afterwards; or until the discharge date if a discharge date is available).
  - If the patient is reported as being on non-invasive ventilation or high-flow oxygen device receiving oxygen on the time of the missing scale value, then the value "5. Hospitalized, on non-invasive ventilation or high flow oxygen devices" will be used (until the next non-missing scale value, if values are available afterwards; or until the discharge date if a discharge date is available).
  - If the patient is reported as being on invasive mechanical ventilation or ECMO/ECLS on the time of the missing scale value, then the value "6. Hospitalized, on invasive mechanical ventilation or ECMO" will be used (until the next non-missing scale value, if values are available afterwards; or until the discharge date if a discharge date is available).
  - Otherwise, for other days, the last observation will be carried forward (until the next non-missing scale value, if values are available afterwards; or until the discharge date if a discharge date is available).

*NEWS-2*

Missing data for computation of the NEWS-2 score will be accounted for as follows:

- Missing at baseline: the missing value of items which are used for the NEWS-2 derivation at baseline will be imputed by the mean.
- Missing values after death date of a patient: a NEWS value of 20 will be used (worst value)
- Other missing values: the last observation will be carried forward (until the next non- missing scale value, if values are available afterwards).

*Oxygenation or Worse*

For the creation of the variable for the secondary endpoint "duration of supplemental oxygen", missing data on the *Room air or Ventilation mode for oxygen support* will be accounted for as follows:

- If an ordinal scale value is available on the same day of the missing data:
  - For patients with an ordinal scale value of "1. Not hospitalized, no limitations on activities" or "2. Not hospitalized, limitation on activities" or "3. Hospitalized, not requiring supplemental oxygen", oxygen use will be imputed with "No"
  - For patients with an ordinal scale value of "4. Hospitalized, requiring supplemental oxygen", or "5. Hospitalized, on non-invasive ventilation or high flow oxygen devices" or "6. Hospitalized, on invasive mechanical ventilation or ECMO", the oxygen use will be imputed with "Yes"
- If the ordinal scale value is not available:
  - The last available data on the Room air or Ventilation mode for oxygen support will be carried forward (until the next non-missing scale value, if values are available afterwards); or until the discharge date if discharged before day 29, or until death in case of death before day 29, or until the day 29 in the other cases.

*Mechanical ventilation or Worse*

For the creation of the variable for the secondary endpoint "duration of mechanical ventilation", missing data on the mechanical ventilation use will be accounted for based on the ordinal scale value:

- If an ordinal scale value is available on the same day of the missing data:
  - For patients with an ordinal scale value of "6. Hospitalized, on invasive mechanical ventilation or ECMO", the mechanical ventilation use will be imputed with "Yes"; for other scale value (except "7. Death") it will be imputed with "No"
- If the ordinal scale value is not available:
  - If the last available mechanical ventilation or ECMO/ECLS information is "Yes", then the patient will be considered to be on mechanical ventilation through day 29, or until death, or until the discharge date if a discharge date is available (after discharge date, the patient will be considered as not being on mechanical ventilation).
  - If the last available mechanical ventilation or ECMO/ECLS information is "No", then the patient will be considered not to be on mechanical ventilation through day 29.

*Hospitalization and re-hospitalization*

If there is a missing data on the date of discharge but a value of the ordinal scale value of "1. Not hospitalized, no limitations on activities" or "2. Not hospitalized, limitation on activities", the date of discharge will be imputed with the day before the first date associated with "1. Not hospitalized, no limitations on activities" or "2. Not hospitalized, limitation on activities".

If a patient is discharged and no further hospitalization data are available, then the patient will be considered as not being re-hospitalised afterwards.

*Mortality*

If there is a missing data on the date of death but a value of the ordinal scale value of "7. Death", the date of death will be imputed with the first date associated with "7. Death".

#### 4.9. Derived and computed variables

The following variables will be derived:

- The time to sustained recovery, defined as:
  - For patients discharged before day 90 without new hospitalization and alive at least 14 days after discharge
    - [(Date of discharge from index hospitalization – Randomization date+1)+14]

  - For patients with several discharges before D90 and re-hospitalisation prior to achieving sustained recovery, the date of the first sustained recovery will be considered.
  - Patients discharged but for whom the information on the status 14 days after discharge is not available will be censored at date of last news on vital status post discharge.
  - Patients with no sustained recovery nor death before D90 will be censored either at day 90 or at the lost to follow-up date or at the withdrawal date.
  - Time to sustained recovery will not be calculated for patients who died before experiencing the outcome, because death is considered as a competitive risk.
  - For patients who died before achieving sustained recovery, time till death will be calculated as:
    - [Date of death – Randomization date+1]

    Of note**,** the shortest possible time to sustained recovery is 14 days (this would require the patient to be discharged from the hospital on the day of randomization), and a patient would have to be discharged from the index hospitalization no later than Day 76 to achieve *sustained recovery* by Day 90.

- The changes from baseline on ordinal scale on Days 29, 90, 180 and 365 calculated as the difference in ordinal scale between baseline and D29, D90, D180 and D365 respectively.
- NEWS-2 value (defined based on the NEWS scoring system reported in Appendix)
- The changes from baseline in NEWS-2 on Days 3, 8, 15 and 29 calculated as the difference in NEWS-2 value between randomization and D3, 8, 15 and 29 respectively.
- Oxygenation or worse: For patients with value of Ventilation mode in one of 3 modalities : “1. Standard oxygenotherapy”, “2. High flow oxygen device”, “3. Non Invasive Ventilation /CPAP” the value of oxygen, non-invasive ventilation and high flow oxygen will be accounted as “Yes”, for patients in Room air or other modalities of ventilation mode it will be accounted as “No”.
- Mechanical ventilation or worse: for patients with value of Ventilation mode in one of 2 modalities: “4. Invasive ventilation”, “5. ECMO” , the value of Mechanical ventilation will be given as “Yes”, for the patients in Room air or in the other modalities it will be accounted as “No”.
- The number of oxygenation-free days in the first 28 days (to Day 29) will be calculated as: [29- number of days on standard oxygenotherapy, high flow oxygen device or non-invasive ventilation]
- Time to new oxygen use, non-invasive ventilation or high flow oxygen devices during the first 28 days (to Day 29), defined as:
  - [Date of initiation of new oxygen use, non-invasive ventilation or high flow oxygen - Randomization date +1]
  - [Date of death - Randomization date +1] if patient died before experiencing the event.
- Durations of new oxygen use, non-invasive ventilation or high flow oxygen devices during the first 28 days (to Day 29), defined as:
  - [End date of new oxygen, non-invasive ventilation or high flow oxygen devices use – Initiation date for the first use during the study+1]
- The number of mechanical ventilation-free days during the first 28 days (to Day 29): [29- number of days on invasive mechanical ventilation or ECMO]
- Time to new invasive mechanical ventilation use during the first 28 days (to Day 29), defined as:
  - [Date of initiation new invasive mechanical ventilation - Randomization date +1]
  - [Date of death - Randomization date +1] if patient died before experiencing the event.

- Durations of invasive mechanical ventilation use during the first 28 days (to Day 29), defined as:
    - [End date of invasive mechanical ventilation use – Initiation date for the first use during the study+1]
- Mechanical ventilation or death between baseline and Day 15 categorized as Yes/No.
- Duration of hospitalization, defined as:
    - [Discharge date – Admission date+1]
- The time to discharge before D90, defined as:
    - [First Discharge date – Randomization date+1]
    - Patient with no discharge date will be censored either at day 90 or at the lost to follow- up date or at the withdrawal date.
    - Time to discharge will not be calculated for patients who die before discharge and before D90, instead their death status will be used as a variable in the competing risk analysis.
- Occurrence of new hospitalization between discharge from index hospitalization and Days 90, 180, 365, categorized as Yes/No
- Confirmed re-infection with SARS-CoV-2 between discharge from index hospitalization and Days 90, 180, 365, categorized as Yes/No.
- At least one occurrence of every AE for a given participant
- Duration of symptoms prior to enrolment, defined as:
    - [Inclusion date – Date of onset of COVID-19 symptoms + 1]

### 5. Description of study patients

#### 5.1. Patient disposition

The number of patients and the flowchart of the study will be presented as in the CONSORT flowchart (Figure 1). Two flowcharts will be presented: one for the Global ITT and mITT populations, with details on the antigenic and vaccination status (vaccination initiation yes, or no) at each step, and one for the positive ITT and mITT populations with details on the vaccination status (vaccination initiation yes, or no).

*Figure 1 CONSORT flowchart*

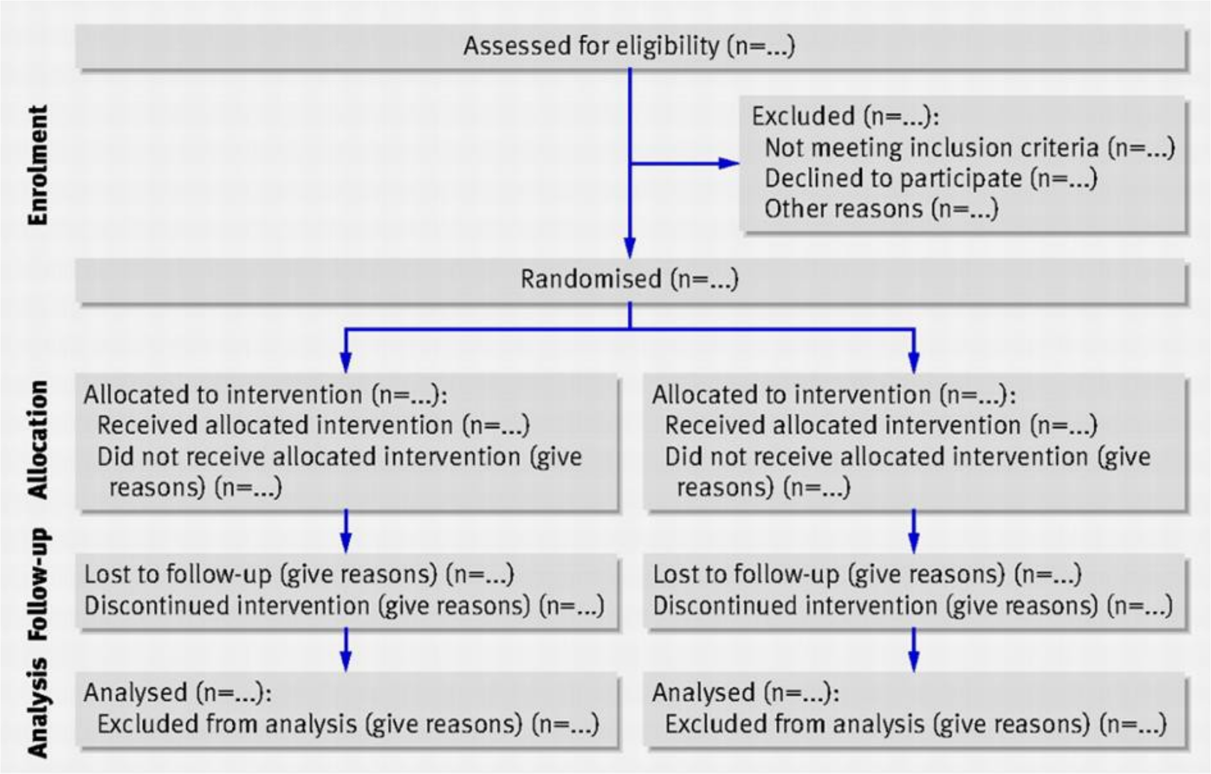

For the primary analysis, a summary table with the description of the number of included subjects, the number of randomized subjects, the number of subjects in whom the treatment was initiated, the number of subjects in whom the treatment was fully administered, the number of subject in whom the treatment was partially administered, the number of subjects who completed the follow up until D90, and the number of subjects who discontinued the study before D90, classified by main reason of withdrawal will be prepared by treatment group and overall stratified by antigenic status (for the Global ITT population) and vaccination status (vaccination initiation yes, or no).

For the final analysis, in addition to the afore mentioned summaries, the number of subjects who completed the study, and the number of subjects who discontinued the study, classified by main reason of withdrawal will be added and similarly the analysis will be presented by treatment group and overall stratified by antigenic status (for the Global ITT population) and vaccination status (vaccination initiation yes, or no).

A summary table with the description of the number and percentage of subjects in each analysis population will be prepared by treatment group and overall. A specific listing of subjects excluded from analysis sets will be provided with the reason(s) for exclusion.

For the primary and final analyses, a summary table with the number and percentage of subjects at each visit will be prepared by treatment group and overall stratified by antigenic and vaccination status (vaccination initiation yes, or no) on the subjects in Global ITT population.

Listings with end of study status and study visit dates will also be generated.

##### 5.2. Protocol violations

The following violations are defined for the purpose of analysis:

- Did not fulfil the eligibility criteria
- Incomplete study treatment administration.
- Received a treatment from another study.
- Withdrew consent or had consent withdrawn by a legally authorised representative.
- Lost to follow-up.

A detailed list of protocol deviations is presented in section 11 of the Study's monitoring plan V4.0 of 07 May 2021.

A summary table by treatment group and overall with the number and percentage of subjects presenting violations relating to inclusion/exclusion criteria will be prepared for the subjects in the Positive ITT population. A summary table by treatment group and overall with the number and percentage of subjects presenting other protocol violations (all violations judged relevant during the data review meeting) will also be prepared. The corresponding listings will be provided.

#### 6. Demographics and other baseline characteristics

The analyses of the demographic and baseline characteristics will be performed on the Positive mITT and Global mITT populations. These analyses will be stratified by randomization arm as well as randomization stratum (whenever possible).

##### 6.1. Descriptive statistics

Demographics and baseline characteristics will be summarized as follows:

- For continuous variables: sample size, mean, standard deviation, median, interquartile range, minimum and maximum.
- For categorical variables: sample size, proportion and number of patients in different categories.

##### 6.2. Demographic data

The following demographic characteristics will be summarized by treatment group and overall:

- Age
- Sex
- Place of life before inclusion
- European region

- Ethnicity
- Geographical origin

##### 6.3. Medical and surgical history

The following baseline medical and surgical history data will be summarized by treatment group, strata and overall:

- Smoking status
- Chronic cardiac disease
- Chronic pulmonary disease (not asthma)
- Asthma
- Chronic kidney disease (stage 1 to 4)
- Liver disease
- Chronic neurological disorder (including dementia)
- Active cancer (including haematological malignancy)
- Hematopoietic stem cell or solid organ transplantation
- Auto-inflammatory disease
- HIV infection
- Obesity (BMI $\geq$ 30)
- Diabetes

##### 6.4. Clinical evaluation

The following baseline clinical data will be summarized by treatment group, strata and overall:

- Delay between first laboratory-confirmed SARS-CoV-2 infection and admission date at facility
- Delay between first laboratory-confirmed SARS-CoV-2 infection and randomization
- Delay between randomization and admission date at facility
- Delay between onset of symptoms and admission date at facility
- Delay between onset of symptoms and randomization
- Department:
    - Conventional unit
    - Intensive Care Unit
- Systolic Blood pressure < 90 mmHg
- Body temperature
- Body weight
- Clinical status
- Ventilation mode
- 7-point ordinal scale at baselines
- NEWS2 Score

##### 6.5. Biological data

The following baseline biological data will be summarized by treatment group, strata and overall:

- Haemoglobin
- Absolute lymphocyte count

- Absolute neutrophil count
- Platelet Count
- Creatinine
- eGFR
- AST / SGOT
- ALT / SGPT
- LDH
- Prothrombin time
- Fibrinogen
- C-reactive protein (CRP)
- D-Dimers
- Ferritin
- Serum total immunoglobulins
- Plasma pregnancy test (woman only)
- Urine pregnancy test (woman only)

##### 6.6. Chest CT data

The following baseline chest CT data will be summarized by treatment group, strata and overall.

- Presence of major artifacts precluding analysis of lung parenchyma
- Presence of pulmonary embolism
- Extension of lung parenchyma lesions
- Presence of pleural effusion
- Presence of Mediastinal lymph node

##### 6.7. Previous and concomitant treatment

Previous and concomitant medications will be listed separately.
A previous treatment will be defined as a treatment prior to the date of first intake of the allocated treatment.
A concomitant treatment will be defined as a treatment that is taken any time during the treatment period (on or after the date of first intake of the allocated treatment).

In case of missing start and end date, or a recorded start date prior to dosing but missing end date, the treatment will be considered concomitant.
A table with the number and percentage of patients having taken at least one previous treatment and a table with the number and percentage of patients having taken at least one concomitant treatment will be generated by treatment group and overall (by medication class and overall).
The following treatments and treatment groups will be described:

- Antibiotics
  - Cefuroxime
  - Cefotaxime
  - Ceftriaxone
  - Amoxicilline
  - Amoxicillin-clavulanic acid
  - Céfotaxime
  - Azithromycine
  - Erythromycine

- Amikacine
- Gentamicin
- Piperacillin-Tazobactam
- Cefepime
- Ceftazidime
- Meropenem
- Flucloxacillin
- Fluroquinolones
- Anti-fungal treatments
- Other

- Anti-inflammatory
    - Corticosteroids
    - Tociluzimab
    - Sarilumab
    - Siltuximab
    - Anakinra
    - Baracitinib
    - Tofacitinib
    - Other anti-inflammatory or immunomodulatory drug
- Anti-virals
    - Remdesivir
    - Ritonavir-Nirmatrelvir (Paxlovid)
    - Molnupiravir
    - Oseltamivir
    - Aciclovir/valaciclovir
    - Ganciclovir/valganciclovir
    - Foscarnet
    - Other antivirals
- The following other medication groups
    - Oral Antidiabetics
    - Insulin therapy
    - NSAID
    - ACE inhibitors
    - Angiotensin II inhibitors
    - Diuretics
    - Antipyretics / Analgesics
    - Anticoagulant
    - Anti-thrombotic (aspirin or other)
    - Other anti-hypertensor treatments

##### 6.8. Study treatment

Study treatment are defined as:

- Placebo single IV infusion
- AZD7442 single IV infusion

A compliant patient will be defined as a patient who has received the complete infusion.

A treatment switch will be defined as a patient who has received the treatment he/she was not allocated to.

#### 7. Analysis

All the efficacy analyses of the primary, key secondary and clinical secondary endpoints will be conducted on the Positive mITT population.

All tests for the analysis of the primary endpoint, key secondary endpoint and clinical secondary endpoints will be stratified by vaccination status (vaccination initiation: yes, or no).

All analyses on safety endpoints will be performed on the Global Safety analysis set.

All tests for the analysis of safety endpoints will be stratified by vaccination status (vaccination initiation: yes, or no) and antigenic status.

##### 7.1. Analysis of the primary endpoint

***Endpoint***

The primary endpoint is defined as the value of the ordinal scale on day 15.

***Descriptive statistics and graphical representation***

The ordinal scale on day 15 will be summarized with the sample size, number and proportion of patients in each category overall with 95% CI and by treatment group.
Stacked barplots overall and by treatment group will be used to display the ordinal scale value from baseline to day 15.

***Statistical methodology***

The ordinal scale will be used to estimate a proportional odds model.[2,3] The primary hypothesis test will be based on a test of whether the common odds ratio for treatment is equal to one. Odds ratios are then interpreted as the odds of being “lower” or “higher” on the ordinal scale across the entire range of the scale. The hypothesis test is, for large sample sizes, nearly the same as the Wilcoxon rank sum test. Therefore, the procedure produces a valid p-value regardless of whether the proportional odds model is correct. Nonetheless, estimation and confidence intervals do require the model to be correct. The validity of the proportionality assumption will be graphically evaluated and tested using a score test, a non- significant test results being interpreted as evidence that the odds ratios are constant across all possible cut points of the outcome.

#### 7.2. Analysis of key secondary endpoint

***Endpoint***

The key secondary endpoint is the time from randomization to sustained recovery, defined as being discharged from the index hospitalization, followed by being alive and at home for 14 consecutive days prior to Day 90.

***Descriptive statistics and graphical representation***

The proportion of patients achieving sustained recovery will be summarized as percentages. Median time to sustained recovery will be reported along with the corresponding IQR.

The time to sustained recovery through Day 90 will be summarized using the cumulative incidence functions to compare the treatment groups, taking into account the "competing risk" of death in analysing sustained recovery.

***Statistical methodology***

The time to sustained recovery through Day 90, will be considered statistically significant if a p<0.05 is observed, and the primary endpoint was first found to be significant (hierarchical testing). In the event the primary endpoint was not found to be significant, the p-value for this analysis will be considered nominal as if no hypothesis testing has been done.

The test will compare the investigational agent versus the control group. Gray's test compares the cumulative incidence functions for sustained recovery between the treatment groups, taking into account the "competing risk" of death in analysing sustained recovery. Gray's test with rho=0 is the analogue of the log-rank test in the presence of competing risks. Cumulative incidence functions for sustained recovery will be estimated by treatment group using the Aalen-Johansen estimator[4], and the recovery rate ratio (RRR) (investigational agent versus control) for sustained recovery will be estimated using the Fine-Gray method[5,6], the RRR will be estimated as a point estimate with a 95% CI. The Aalen- Johansen estimator for cumulative incidence functions is the analogue of the Kaplan-Meier estimator in the presence of competing risks. The Fine-Gray method is the competing risks equivalent of Cox proportional hazards models; the RRR compares the cumulative incidence rates of sustained recovery between the study arms, and is a sub-distribution hazards ratio.

#### 7.3. Analysis of the secondary endpoints

##### 7.3.1. Ordinal scale

***Endpoints***

The secondary endpoint of the ordinal scale is:

- the value of the ordinal scale on Days 29, 90, 180 and 365.

***Descriptive statistics and graphical representations***

The ordinal scale on days 29, 90, 180 and 365 will be summarized according to proportions with confidence intervals on the difference or odds ratios for a binary or multiple category scale, respectively.

***Statistical methodology***

The ordinal scale on day 29, 90, 180 and 365 will be analysed using a proportional odds model similarly to the primary endpoint.

##### 7.3.2. National Early Warning Score 2 (NEWS-2)

***Endpoints***

The endpoints associated with the NEWS-2 are:

- Change from baseline to Days 3, 8, 15 (while hospitalized) and 29 in NEWS-2

***Descriptive statistics and graphical representations***

Change from baseline in ordinal scale on Days 3, 8, 15 and 29 will be summarized using mean, standard deviation, median, interquartile range, minimum and maximum.

***Statistical methodology***

Change from baseline to days 3, 8, 15 and 29 will be compared using a Student t-test or an ANCOVA model with the treatment group as effect.

##### 7.3.3. Oxygenation

***Endpoints***

The endpoints regarding oxygenation are:

- Number of oxygenation-free days in the first 28 days.
- Incidence of new oxygen, non-invasive ventilation or high flow oxygen devices use during the first 28 days (to Day 29).

***Descriptive statistics***

The number of oxygenation-free days in the first 28 days will be summarized according to median days with quartiles.

Incidence of new oxygen use, non-invasive ventilation or high flow oxygen devices will be summarized as percentages with confidence intervals.

Differences in median time to new oxygen use, non-invasive ventilation or high flow oxygen devices by treatment will be summarized with Kaplan-Meier curves and 95% confidence bounds. Kaplan-Meier survival probabilities and their 95% confidence intervals will be reported.

***Statistical methodology***

The number of oxygenation-free days in the first 28 days will be compared using a Student t-test or ANCOVA model with the treatment group as effect.

The time to new oxygen use, non-invasive ventilation or high flow oxygen devices will be compared between treatment arm and control using hazard ratios and confidence interval calculated by Cox proportional hazards model (model fitted by maximizing the partial likelihood). The Efron method will be used to handle ties.

###### 7.3.4. Mechanical ventilation

***Endpoints***

The endpoints regarding mechanical ventilation are:

- Number of mechanical ventilation-free days in the first 28 days.
- Incidence of invasive mechanical ventilation use during the first 28 days (to Day 29)
- Need for mechanical ventilation or death by day 15.

***Descriptive statistics***

The number of mechanical ventilation-free days in the first 28 days will be summarized according to median days with quartiles.
Incidence of new mechanical ventilation use will be summarized as percentages with 95% confidence intervals. Differences in median times to new mechanical ventilation by treatment will be summarized with Kaplan-Meier curves and 95% confidence bounds. Kaplan-Meier survival probabilities and their 95% confidence intervals will be reported.
Need for mechanical ventilation or death by Day 15 use will be summarized as percentages with 95% confidence intervals.

***Statistical methodology***

The number of mechanical ventilation (or ECMO)-free days in the first 28 days will be compared using a Student t-test or an ANCOVA model with the treatment group as effect.

The time to new mechanical ventilation use (or ECMO or death whichever occurs first) will be compared between treatment groups and control using hazard ratios and confidence interval calculated by Cox proportional hazards model (model fitted by maximizing the partial likelihood). The Efron method will be used to handle ties.

Need for mechanical ventilation or death by Day 15 will be compared between treatment arm and control using a Chi Squared test or stratified Cochran-Mantel-Haenszel test.

###### 7.3.5. Hospitalization

The time to discharge will be analysed using the same approach described for the analysis of the key secondary outcome.

***Endpoints***

The endpoint regarding hospitalization is time to discharge.

***Descriptive statistics and graphical representation***

Median number of hospitalization days will be reported along with the corresponding IQR.
Time to discharge will be summarized using the cumulative incidence functions to compare the treatment groups, taking into account the “competing risk” of death in analysing the outcome.

***Statistical methodology***

The cumulative incidence functions within each treatment group will be estimated using the Aalen- Johansen method, and treatment groups will be compared using Gray’s test with rho=0. The Aalen- Johansen estimates, and Gray’s test are analogues of the Kaplan-Meier estimates, and the log-rank test, respectively, taking into account the competing risk of death.

##### 7.3.6. Mortality

***Endpoints***

The endpoints regarding mortality are:

- in-hospital mortality rate
- mortality rate at 29 days
- mortality rate at 90 days
- mortality rate at 180 days
- mortality rate at 365 days
- mortality rate at 456 days

***Descriptive statistics***

In-hospital mortality rate and mortality rates at 29, at 90, at 180, 365 and 456 days will be summarized with the number and proportion of patients.

***Statistical methodology***

The Kaplan-Meier estimate will be used to estimate the mortality rates. We will calculate the Cochran- Mantel-Haenszel treatment difference and the 95% associated confidence interval in the percentage of mortality.

##### 7.3.7. Long term health status

***Endpoints***

The endpoints regarding long term health status are:

- Occurrence of new hospitalization between discharge and days 90, 180 and 365.
- Occurrence of a confirmed re-infection with SARS-CoV-2 between discharge and days 90, 180 and 365.

***Descriptive statistics***

Occurrence of new hospitalization will be summarized with the number and proportion of patients experiencing re-hospitalization between discharge and days 90, 180 and 365.
Occurrence of a confirmed re-infection with SARS-CoV-2 days will be summarized with the number and proportion of patients experiencing a SARS-CoV-2 re-infection between discharge and days 90, 180 and 365.

***Statistical methodology***
Proportion of patients experiencing re-hospitalization between discharge and days 90, 180 and 365 will be compared between the 2 study groups using a Chi Squared test or a stratified Cochran- Mantel-Haenszel test
Proportion of patients experiencing a SARS-CoV-2 re-infection between discharge and D90, 180 and 365 will be compared between the 2 study groups using a stratified Cochran- Mantel-Haenszel test.

###### 7.3.8. Safety

***Endpoints***
The safety endpoints are:

- Cumulative incidence of SAE evaluated through 456 days of follow up.
- Cumulative incidence of Grade 3 and 4 events (AEs) evaluated through 456 days of follow up.
- Cumulative incidence of Grade 1-2 hypersensitivity-related and infusion related AEs until D29 visit.
- Cumulative incidence of AEs of special interest evaluated through 456 days of follow up.
- Discontinuation of investigational therapeutics (for any reason).

***Descriptive statistics***
Each SAE, grade 3 and 4 events, Grade 1-2 hypersensitivity-related and infusion related AEs, and AEs of special interest will be summarized, by System Organ Class and Preferred Term, with the number and percentage of patients with at least one event and the total number of events for each endpoint.
Discontinuation of investigational therapeutics (for any reason) will be summarized with the number and percentage of patients by treatment group and overall.

***Statistical methodology***
Patients with at least one SAE, patients with at least one grade 3 or 4 AE, patients with at least 1-2 hypersensitivity-related and infusion related AEs and patients with at least one AE of special interest will be compared using a stratified Cochran-Mantel-Haenszel test.

#### 8. Sensitivity analysis

Sensitivity analysis for the primary, key secondary, and other clinical efficacy end-points will be conducted on the Global mITT population stratified by vaccination status (vaccination initiation yes, or no) and antigenic status at randomization.

Sensitivity analysis for safety endpoints will be performed on the Positive Safety analysis set.

#### 9. Subgroup analyses

Subgroup analyses will be performed for the primary efficacy endpoint and the key secondary efficacy endpoint, first for the Ag+ mITT population, and second for the full (Ag+/Ag-) mITT population whatever the vaccination status (vaccination initiation yes, or no), to evaluate the treatment effect across the following subgroups:

- Rapid antigen test result at inclusion (Ag+ vs Ag-, for the analysis on the Global ITT population).
- Vaccination status (yes vs no).
- Clinical status at inclusion (7-point ordinal scale of 3 OR 4 vs 5).
- Duration of symptoms prior to enrolment < 5 vs ≥ 5 days).
- Age (< 65 years vs ≥ 65 years).
- SARS-CoV-2 serology (seronegative vs seropositive) at inclusion.
- SARS-CoV-2 lineage at inclusion (categories to be defined from blind analysis).
- Sex.

For each population of analysis, a forest plot will display confidence intervals across subgroups.
Interaction tests will be conducted to determine whether the effect of treatment varies between subgroups.

### 10. Appendix:

#### 10.1. Power computation

These computations have been made for 413 subjects per arm and 20% vaccinated patients, for various assumptions on treatment effect and outcome at day 15 in vaccinated vs non-vaccinated patients

| **Outcome distribution in SoC arms in Vaccinated *vs* Non-vaccinated patients** | **OR of treatment effect in vaccinated patients** | | |
|---|---|---|---|
| | **OR = 1.3** (**lower** treatment effect than in non-vaccinated) | **OR = 1.5** (**same** treatment effect than in non-vaccinated) | **OR = 1.7** (**better** treatment effect than in non-vaccinated) |
| OR = 0.8 (**worse** outcome at day 15 in vaccinated than in non-vaccinated) | 86% | 90% | 93% |
| OR = 1 (**same** outcome at day 15 in vaccinated than in non-vaccinated) | 86% | 90% | 93% |
| OR = 1.2 (**better** outcome at day 15 in vaccinated than in non-vaccinated) | 86% | 90% | 93% |

#### 10.2. NEWS-2 scoring system

| **Physiological parameter** | **Score** 3 | 2 | 1 | 0 | 1 | 2 | 3 |
|---|---|---|---|---|---|---|---|
| Respiration rate (per minute) | ≤8 | | 9–11 | 12–20 | | 21–24 | ≥25 |
| $SpO_2$ Scale 1 (%) | ≤91 | 92–93 | 94–95 | ≥96 | | | |
| $SpO_2$ Scale 2 (%) | ≤83 | 84–85 | 86–87 | 88–92 ≥93 on air | 93–94 on oxygen | 95–96 on oxygen | ≥97 on oxygen |
| Air or oxygen? | | Oxygen | | Air | | | |
| Systolic blood pressure (mmHg) | ≤90 | 91–100 | 101–110 | 111–219 | | | ≥220 |
| Pulse (per minute) | ≤40 | | 41–50 | 51–90 | 91–110 | 111–130 | ≥131 |
| Consciousness | | | | Alert | | | CVPU |
| Temperature (°C) | ≤35.0 | | 35.1–36.0 | 36.1–38.0 | 38.1–39.0 | ≥39.1 | |

*

SpO2 scale: For patients confirmed to have hypercapnic respiratory failure on blood gas analysis on either a prior or their current hospital admission, and requiring supplemental oxygen, we recommend (i) a prescribed oxygen saturation target range of 88–92%, and (ii) that the dedicated SpO2 scoring scale (Scale 2) on the chart should be used to record and score the oxygen saturation for the NEWS. The decision to use SpO2 scale 2 should be made by a competent clinical decision maker and should be recorded in the patient's clinical notes. In all other circumstances, the regular NEWS SpO2 scale 1 should be used.

CVPU: Confusion or arousable only to voice (V) or pain (P), or unresponsive (U).

#### References

1. Lebel C. RTSM Randomisation, version 1. DisCoVeRy. Multi-center, adaptive, randomized trial of the safety and efficacy of treatments of COVID-19 in hospitalized adults. Venn Life Sciences; 2021
2. Agresti A. *An Introduction to Categorical Data Analysis*. John Wiley & Sons; 2018.
3. Hosmer DW, Lemeshow S. *Applied Logistic Regression*. Wiley New York; 2000.3
4. Aalen OO, Johansen S. An Empirical Transition Matrix for Non-Homogeneous Markov Chains Based on Censored Observations. Scand. J. Stat. 1978; 5(3):141–150.
5. Fine JP and Gray RJ. A proportional hazards model for the subdistribution of a competing risk. J Am Stat Assoc. 1999; 94:496-509.
6. Zhou B, Latouche A, Rocha V, Fine J. (2011). Competing Risks Regression for Stratified Data. Biometrics. 67(2):661-70.

### Multi-centre, adaptive, randomized trial of the safety and efficacy of treatments of COVID-19 in hospitalized adults

#### Statistical analysis plan for exploratory endpoints – Virologic efficacy in respiratory and blood samples.

CONFIDENTIAL

#### Signatures

| RESPONSIBILITY | NAME | SIGNATURE | DATE |
|---|---|---|---|
| **Statistician** | Lien HAN | | 2023-01-03 |
| **Methodologist** | Clément MASSONNAUD | | 2023-01-03 |
| **Chief methodologist** | France MENTRE | | 2023-01-03 |
| **Virologist** | Maude BOUSCAMBERT | | 2023-01-04 |
| **DisCoVeRy Trial steering committee** | Dominique COSTAGLIOLA | | 2023-01-18 |

#### Revision history

| VERSION NO. | EFFECTIVE DATE | DESCRIPTION |
|---|---|---|
| 1.0 | 18-01-2022 | First version |
| 1.1 | 03-01-2023 | Simplification since early stop of inclusions |

#### List of abbreviations

| ABBREVIATION | DESCRIPTION |
|---|---|
| ITT | Intention-to-treat |
| LOD | Limit of detection |
| LRT | Lower respiratory tract |
| mITT | Modified intention-to-treat |
| NormLOD | Normalized limit of detection |
| NP | Nasopharyngeal |
| SAP | Statistical analysis plan |

### 1. Overview of the study

#### 1.1. Introduction

This document summarizes the statistical methods that will be used to analyse the exploratory endpoints related to the treatment's (AZD 7442) virologic efficacy in respiratory samples (nasopharyngeal, NP swabs or lower respiratory tract, LRT) and in blood samples.
Analyses of the data generated by whole genome sequencing of the isolated virus will be described in an annex to this SAP, after all viral sequences have been generated.

A primary Statistical Analysis Plan (SAP) is available detailing the statistical methods of the study's primary, key secondary and secondary efficacy and safety endpoints.

The statistical analysis presented in this document will be performed after the first database lock once all patients have reached their D90 visit. The analyses described below are based upon the clinical study protocol V15.0 dated 20th September 2021.This new version of the Statistical Analysis Plan has been edited following the decision by the Discovery Steering Committee on June 23, 2022 to stop the inclusions on July 1st, 2022 due to a low rate of inclusion. It is therefore adapted to account for a smaller sample size than initially planned.

#### 1.2. Study objective relative to virologic efficacy

One of the exploratory objectives of the study is to evaluate the virologic efficacy of AZD 7442 compared to the control arm.

### 2. General considerations

#### 2.1. Definition of analysis sets

The analysis sets are the same as those defined in the primary SAP; used analysis sets for the analyses described in this document are:

- Positive intention-to-treat (Positive ITT): all randomized patients with positive SARS-CoV-2 antigenic status at randomization who will be analysed in their randomization group.
- Global intention-to-treat (Global ITT): all randomized patients with positive or negative SARS-CoV-2 antigenic status at randomization who will be analysed in their randomization group.
- Positive modified intention-to-treat (Positive mITT): patients of the Positive ITT in whom the allocated treatment was initiated.
- Global modified intention-to-treat (Global mITT): patients of the Global ITT in whom the allocated treatment was initiated.

#### 2.2. Type-I error rate and adaptive design

A two-sided Type-I error rate of 5% will be used.

##### 2.3. Normalized viral load and limit of detection

Normalized SARS-CoV-2 viral loads concern respiratory samples analysis. They are directly reported in the eCRF and will be analysed. The normalization uses cell quantification in the sample (either NP or LRT), and allows the expression of SARS-CoV-2 viral loads in $log_{10}$ copies/10 000 cells.
If the number of cells in the sample is below 500 cells / 5µL, the sample is considered to have a poor quality and is reported in the eCRF as "non-analysable", despite being available.
The limit of detection (LOD) for the viral load is 4 copies / 5µL and the normalized LOD (NormLOD) is: 10 copies/10 000 cells (=1 $log_{10}$ copies/10 000 cells).
Viral loads below the LOD are recorded as undetectable in the eCRF.

In blood, a quantitative SARS-CoV-2 viral load is measured and expressed in $log_{10}$ copies/mL. The limit of detection (LOD) for the viral load is 4 copies / 5µL of RNA and the LOD for the viral load in sample is 1 $log_{10}$ copies/mL.

##### 2.4. Handling of missing data

Missing data will not be replaced.

The number, timing and pattern for missing value will be detailed for each outcome measure.
A value is considered as missing if the sample was collected but not analysable or if the sample was not collected.

Given that time to time to first undetectable viral load will be analyzed using competing risk analysis with death as the competitive risk, only patients who are lost to follow up or those who withdraw from the study will be censored at their withdrawal or lost to follow up date.

#### 3. Analysis of the endpoints

The analysis of the virologic efficacy will be performed on the Positive mITT population. All tests will be stratified by vaccination status (vaccination initiation yes or no). Vaccination is considered as initiated if at least one injection of any vaccine against SARS-CoV-2 was received prior to enrolment, whatever the delay.

A sensitivity analysis will be performed on Global mITT population with stratification by vaccination status (vaccination initiation yes or no) and by antigenic status (positive or negative, obtained from the result of a rapid antigen test on nasopharyngeal swab performed at enrolment).

***Endpoints***

The virologic endpoints are:

Efficacy

1. Percent of subjects with detectable SARS-CoV-2 in NP swabs at baseline (Day 1 pre-treatment) and at Days 3, 8, 15 (while hospitalized) and 29
2. Percent of subjects with detectable SARS-CoV-2 in LRT samples at baseline (Day 1 pre-treatment) and at Days 3, 8, 15 (while hospitalized) and 29
3. Normalized quantitative SARS-CoV-2 viral load in NP swabs at baseline (Day 1 pre-treatment) and at Days 3, 8, 15 (while hospitalized) and 29
4. Change from baseline (Day 1 pre-treatment) of normalized quantitative SARS-CoV-2 viral load in

NP swabs at Days 3, 8, 15 (while hospitalized) and 29
5. Time to first undetectable normalized quantitative SARS-CoV-2 viral load in NP swabs through day 29
6. Normalized quantitative SARS-CoV-2 viral load in LRT samples at baseline (Day 1 pre-treatment) and at Days 3, 8, 15 (while hospitalized) and 29
7. Change from baseline (Day 1 pre-treatment) of normalized quantitative SARS-CoV-2 viral load in LRT samples at Days 3, 8, 15 (while hospitalized) and 29
8. Percent of subjects with detectable SARS-CoV-2 in blood at baseline (Day 1 pre-treatment) and at Days 3 and 8
9. Quantitative SARS-CoV-2 viral load in blood at baseline (Day 1 pre-treatment) and at Days 3 and 8
10. Change from baseline (Day 1 pre-treatment) of quantitative SARS-CoV-2 viral load in blood at Days 3 and 8

Of note, there is a discrepancy between the Statistical Analysis Plan and the protocol v15.00 of September 20, 2021, where secondary objectives 4, 5, 7 and 10 were not presented.

***Descriptive statistics and graphical representations***

The percent of subjects with detectable SARS-CoV-2 in NP swabs will be summarized with the number and proportion of patients with a positive PCR at each assessed day.

The percent of subjects with detectable SARS-CoV-2 in LRT samples will be summarized with the number and proportion of patients with a positive PCR at each assessed day.

The analysable $\log_{10}$ of normalized quantitative SARS-CoV-2 viral load in NP swabs will be summarized by the mean, standard deviation, median, interquartile range, minimum and maximum.

The analysable change from baseline (Day 1 pre-treatment) of $\log_{10}$ of normalized quantitative SARS-CoV-2 viral load in NP swabs will be summarized by the mean, standard deviation, median, interquartile range, minimum and maximum.

Median time to first undetectable SARS-CoV-2 viral load in NP swabs will be reported along with the corresponding IQR. The time to undetectable SARS-CoV-2 viral load through Day 29 will be summarized using the cumulative incidence functions to compare the treatment groups, taking into account the "competing risk" of death in analysing first undetectable SARS-CoV-2 viral load.

The analysable $\log_{10}$ of normalized quantitative SARS-CoV-2 viral load in LRT samples will be summarized by the mean, standard deviation, median, interquartile range, minimum and maximum.

The analysable change from baseline (Day 1 pre-treatment) of $\log_{10}$ of normalized quantitative SARS-CoV-2 viral load in LRT samples will be summarized by the mean, standard deviation, median, interquartile range, minimum and maximum.

The percent of subjects with detectable SARS-CoV-2 in blood will be summarized with the number and proportion of patients with a positive PCR at each assessed day.

The analysable $\log_{10}$ of quantitative SARS-CoV-2 viral load in blood samples will be summarized by the mean, standard deviation, median, interquartile range, minimum and maximum.

The analysable change from baseline (Day 1 pre-treatment) of $\log_{10}$ of quantitative SARS-CoV-2 viral load in blood samples will be summarized by the mean, standard deviation, median, interquartile range, minimum and maximum.

***Statistical methodology***
The number of subjects with SARS-CoV-2 detectable in NP swabs on Days 1, 3, 8, 15, and 29 will be compared between groups at each day using a stratified Cochran-Mantel-Haenszel test. For each day and each group, only patients with analysable viral loads will be considered.

The number of subjects with SARS-CoV-2 detectable in LRT samples on Days 1, 3, 8, 15, and 29 will be compared between groups at each day using a Chi Squared test or a stratified Cochran-Mantel-Haenszel test. For each day and each group, only patients with analysable viral loads will be considered.

The $log_{10}$ normalized quantitative SARS-CoV-2 viral load in NP swabs will be compared between groups using linear mixed models with repeated measures for each patient. The treatment arm will be the main exposure variable and the time of measurement (at baseline and on days 3, 8, 15, and 29), time by treatment interaction and stratification variables as covariates. Patient effect will be accounted for as random intercept and slope. Treatment effect will be tested and reported on the slope of time by treatment interaction term.
Results will be reported using slope expressed in $log_{10}$/day and slope difference, along with their estimated standard errors and 95% confidence interval.
In addition, the difference in the viral loads, and the difference in the change from baseline of the viral loads between treatment groups along with its se and 95%CI will be estimated at days 3, 8, 15 and 29 using the linear mixed effects model.
For this analysis, undetectable viral load values (i.e. values < 1 $log_{10}$ copies/10 000 cells) will be imputed to half the LOD, hence 0.7 $log_{10}$ copies/10 000 cells. In case of several consecutive undetectable values, only the first one will be replaced and the subsequent values will be discarded (until the next detectable value, if values are available afterwards).

Gray's test will be used to compare the cumulative incidence functions for first undetectable viral load between the treatment groups, taking into account the "competing risk" of death in analysing the time to first undetectable viral load. Gray's test with rho=0 is the analogue of the log-rank test in the presence of competing risks. Cumulative incidence functions for first undetectable viral load will be estimated by treatment group using the Aalen-Johansen estimator, and the recovery rate ratio (RRR) (investigational agent versus control) for first undetectable viral load will be estimated using the Fine-Gray method, the RRR will be estimated as a point estimate with a 95% CI. The Aalen- Johansen estimator for cumulative incidence functions is the analogue of the Kaplan-Meier estimator in the presence of competing risks. The Fine-Gray method is the competing risks equivalent of Cox proportional hazards models; the RRR compares the cumulative incidence rates of first undetectable viral load between the study arms, and is a sub-distribution hazards ratio.

The $log_{10}$ normalized quantitative SARS-CoV-2 viral load in LRT samples will be compared between groups using linear mixed models with repeated measures for each patient. The treatment arm will be the main exposure variable and the time of measurement (at baseline and on days 3, 8, 15, and 29), time by treatment interaction and stratification variables as covariates. Patient effect will be accounted for as random intercept and slope. Treatment effect will be tested and reported on the slope of time by treatment interaction term.
Results will be reported using slope expressed in $log_{10}$/day and slope difference, along with their estimated standard errors and 95% confidence interval.
In addition, the difference in the viral loads, and the difference in the change from baseline of the viral loads between treatment groups along with its se and 95%CI will be estimated at days 3, 8, 15 and 29

using the linear mixed effects model.
For this analysis, undetectable viral load values (i.e. values < 1 $\log_{10}$ copies/10 000 cells) will be imputed to half the LOD, hence 0.7 $\log_{10}$ copies/10 000 cells. In case of several consecutive undetectable values, only the first one will be replaced and the subsequent values will be discarded (until the next detectable value, if values are available afterwards).

The number of subjects with SARS-CoV-2 detectable in blood on Days 1, 3, and 8 will be compared between groups at each day using a Chi Squared test or a stratified Cochran-Mantel-Haenszel test. For each day and each group, only patients with analysable viral loads will be considered.

The $\log_{10}$ normalized quantitative SARS-CoV-2 viral load in blood samples will be compared between groups using linear mixed models with repeated measures for each patient. The treatment arm will be the main exposure variable and the time of measurement (at baseline and on days 3, and 8), time by treatment interaction and stratification variables as covariates. Patient effect will be accounted for as random intercept. Treatment effect will be tested and reported on the slope of time by treatment interaction term.
Results will be reported using slope expressed in $\log_{10}$/day and slope difference, along with their estimated standard errors and 95% confidence interval.
In addition, the difference in the viral loads between treatment groups along with its SE and 95%CI will be estimated at days 3, 8, 15 and 29 using the linear mixed effects model.
For this analysis, undetectable viral load values (i.e. values < 1 $\log_{10}$ copies/mL) will be imputed to half the LOD, hence 0.7 $\log_{10}$ copies/mL. In case of several consecutive undetectable values, only the first one will be replaced and the subsequent values will be discarded (until the next detectable value, if values are available afterwards).

#### 4. Subgroup analyses

Subgroup analyses will be performed for the the slopes of the change of normalized quantitative SARS-CoV-2 viral load in NP, on both Positive mITT and Global mITT population, to evaluate the treatment effect across the following subgroups:

- Rapid antigen test result at inclusion (positive or negative, only for the analysis on the Global mITT population)
- Vaccination status (vaccination initiation yes or no)
- Clinical status at inclusion (7-point ordinal scale of 3 OR 4 vs 5)
- Duration of symptoms prior to enrolment (< 5 vs ≥ 5 days)
- Age (< 65 years vs ≥ 65 years)
- SARS-CoV-2 serology (seronegative vs seropositive) at inclusion
- SARS-CoV-2 lineage at inclusion (categories to be defined from blind analysis)
- Sex

A forest plot will display confidence intervals across subgroups.
Interaction tests will be conducted to determine whether the effect of treatment varies between subgroups.

Multi-centre, adaptive, randomized trial of the safety and efficacy of treatments of COVID-19 in hospitalized adults

### Statistical analysis plan for exploratory endpoints – Serum neutralization of SARS-CoV-2 in blood samples.

CONFIDENTIAL

#### Signatures

| RESPONSIBILITY | NAME | SIGNATURE | DATE |
|---|---|---|---|
| **Statistician** | Lien HAN | | 15/05/2023 |
| **Methodologist** | Clément MASSONNAUD | | 12/05/2023 |
| **Chief methodologist** | France MENTRE | | 12/05/2023 |
| **Virologist** | Alexandre GAYMARD | | 12/05/2023 |
| **Immunologist** | Guislaine CARCELAIN | | 12/05/2023 |
| **Seroneutralisation** | Pascal POIGNARD | | 15/05/2023 |
| **DisCoVeRy Trial steering committee** | Dominique COSTAGLIOLA | | 16/05/2023 |

#### Revision history

| VERSION NO. | EFFECTIVE DATE | DESCRIPTION |
|---|---|---|
| 1.0 | 16-05-2023 | First version |

#### List of abbreviations

| ABBREVIATION | DESCRIPTION |
|---|---|
| ITT | Intention-to-treat |
| LOD | Limit of detection |
| mITT | Modified intention-to-treat |
| Anti-N IgG | SARS-CoV-2 anti-N immunoglobulins |
| Anti-S IgG | SARS-CoV-2 anti-spike protein immunoglobulins |
| SAP | Statistical analysis plan |

### 1. Overview of the study

#### 1.1. Introduction

This document summarizes the statistical methods that will be used to analyse the exploratory endpoints related to the neutralizing antibodies in serum samples.

A primary Statistical Analysis Plan (SAP) is available detailing the statistical methods of the study's primary, key secondary and secondary efficacy and safety endpoints.

The statistical analysis presented in this document will be performed after the first database lock once all patients have reached their D90 visit. The analyses described below are based upon the clinical study protocol V15.0 dated 20th September 2021.This Statistical Analysis Plan has been edited following the decision by the Discovery Steering Committee on June 23, 2022 to stop the inclusions on July 1st, 2022 due to a low rate of inclusion. It is therefore adapted to account for a smaller sample size than initially planned.

#### 1.2. Study objective

The objective is to evaluate the effect of AZD7442 on the titer of neutralizing antibodies against the three variants Delta, Omicron BA.1 and Omicron BA.2, separately, in patients' sera at Day 3 and Day 8 post-randomization.

### 2. General considerations

#### 2.1. Definition of analysis sets

The analysis sets are the same as those defined in the primary SAP; used analysis sets for the analyses described in this document are:

- Positive intention-to-treat (Positive ITT): all randomized patients with positive SARS-CoV-2 antigenic status at randomization who will be analysed in their randomization group.
- Global intention-to-treat (Global ITT): all randomized patients with positive or negative SARS-CoV-2 antigenic status at randomization who will be analysed in their randomization group.
- Positive modified intention-to-treat (Positive mITT): patients of the Positive ITT in whom the allocated treatment was initiated.
- Global modified intention-to-treat (Global mITT): patients of the Global ITT in whom the allocated treatment was initiated.

##### 2.2. Definition of variant of infection

The variant of infection of the patient will be recoded as a categorical variable of 3 modalities, based on the sequencing at baseline:

- Variants "wildtype", "alpha", "beta", "gamma", "delta" will be grouped as "pre-omicron"
- Variant "Omicron BA.1" will stay "Omicron BA.1"
- Variants "Omicron BA.2" and "Omicron BA.5" will be grouped as "Omicron BA.2/5"

##### 2.3. Type-I error rate and adaptive design

A two-sided Type-I error rate of 5% will be used.

##### 2.4. Normalized measurements and limit of detection

Values for neutralizing antibodies titers are reported as the 50% inhibitory concentration (IC50).

For this analysis, the limit of detection (LOD) is defined at a value of 30. Values below LOD (or "NN" – no neutralization) will be imputed to 15.

##### 2.5. Handling of missing data

Missing data for neutralizing antibodies titers will not be replaced.

The number, timing and pattern for missing value will be detailed for each outcome measure.
A value is considered as missing if the sample was collected but not analysable or if the sample was not collected.

Missing values for the variant of infection at baseline will be handled as follows:

- If the value at baseline is missing, it will be imputed using the value at day 3 or day 8, if available
- If no value is available at any time point, it will be imputed by comparing the date of symptoms onset to the epidemiological data on variants circulation in France at that time (*Santé Publique France* "Flash" surveys).
    - Before 2021/12/09: imputed to "pre-omicron" (> 95% of Delta in France up until then)
    - From 2022/01/06 to 2022/01/24: imputed to "omicron BA.1" (> 95% of BA.1 in France)
    - After 2022/03/28: imputed to "omicron BA.2/5" (> 95% BA.2 from this point in France)
    - Otherwise, recorded as "unknown"

#### 3. Analysis of the endpoints

The analysis of seroneutralization will be performed on the patients from the Global mITT population with known variant of infection at baseline. For the primary analysis, all tests will be stratified by vaccination status (vaccination initiation yes or no) and by variant of infection (pre-omicron ; omicron BA.1 ; omicron BA.2/5). Vaccination is considered as initiated if at least one injection of any vaccine against SARS-CoV-2 was received prior to enrolment, whatever the delay.

For all analyses, we will consider the IC50 titers of neutralizing antibodies targeted at the variants corresponding to the variant of infection of the patient:

- Neutralization against variant "Delta" for patients infected with a "pre-omicron" variant

- Neutralization against variant “BA.1” for patients infected with a “Omicron BA.1” variant
- Neutralization against variant “BA.2” for patients infected with a “Omicron BA.2/5” variant

***Endpoints***

- Neutralizing antibodies titers (IC50) at day 3 and day 8
- Change from baseline (day 1 pre-treatment) of neutralizing antibodies titers (IC50) at days 3 and 8
- Proportion of titers below LOD at days 3 and 8

***Descriptive statistics and graphical representations***

A flow-chart of the sera sampled and analysed at baseline (Day 1 pre-treatment), Day 3 and Day 8, according to each treatment group will be presented.

The $log_{10}$ neutralizing antibodies titers (IC50), at baseline (Day 1 pre-treatment), Day 3 and Day 8, for both treatment groups, will be summarized by the mean, standard deviation, median, interquartile range, minimum and maximum.

The distribution of the neutralizing antibodies titers (IC50) against the three variants will be presented on $log_{10}$ scale at baseline (Day 1 pre-treatment), Day 3 and Day 8, for both treatment groups.

The change from baseline (Day 1 pre-treatment) to Day 3 and to Day 8 will be computed as change of $log_{10}$ neutralizing antibodies titers. It will be summarized by the mean, standard deviation, median, interquartile range, minimum and maximum. Geometric mean ratio of neutralizing antibodies and their 95% confidence intervals will also be reported.

The number of subjects with undetectable neutralizing antibodies will be presented, at baseline (Day 1 pre-treatment), Day 3 and Day 8, for both treatment groups as numbers and proportions.

***Statistical methodology***

The $log_{10}$ neutralizing antibodies titers at Day 3 and Day 8 will be compared between the two treatment groups using an ANCOVA model with the treatment group as effect.

The change from baseline (Day 1 pre-treatment) to Day 3 and Day 8 of $log_{10}$ neutralizing antibodies titers will be compared between the two treatment groups using an ANCOVA model with the treatment group as effect.

The number of subjects with undetectable neutralizing antibodies titers at Day 3 and 8 will be compared between groups using stratified Cochran-Mantel-Haenszel test.

**Subgoup analyses**

The same analyses will be performed on the following subgroups:

- Vaccination status (yes/no): analyses will be stratified on the variant of infection
- Variant of infection (pre-omicron ; BA.1 ; BA.2/5): analyses will be stratified on the vaccination status
- Serology at baseline (seropositive/seronegative): analyses will be stratified on the variant of infection.
